# Omicron COVID-19 Immune Correlates Analysis of a Third Dose of mRNA-1273 in the COVE Trial

**DOI:** 10.1101/2023.10.15.23295628

**Authors:** Bo Zhang, Youyi Fong, Jonathan Fintzi, Eric Chu, Holly E. Janes, Lindsay N. Carpp, Avi Kenny, Marco Carone, David Benkeser, Lars W. P. van der Laan, Weiping Deng, Honghong Zhou, Xiaowei Wang, Yiwen Lu, Chenchen Yu, Bhavesh Borate, Christopher R. Houchens, Karen Martins, Lakshmi Jayashankar, Chuong Huynh, Carl J. Fichtenbaum, Spyros Kalams, Cynthia L. Gay, Michele P. Andrasik, James G. Kublin, Lawrence Corey, Kathleen M. Neuzil, Frances Priddy, Rituparna Das, Bethany Girard, Hana M. El Sahly, Lindsey R. Baden, Ruben O. Donis, Richard A. Koup, Peter B. Gilbert, Dean Follmann, United States Government (USG) COVID-19 Immune Assays Team, Moderna, Inc. Team, Coronavirus Vaccine Prevention Network (CoVPN)/Coronavirus Efficacy (COVE) Team, USG/CoVPN Biostatistics Team

**Affiliations:** Vaccine and Infectious Disease Division, Fred Hutchinson Cancer Center; Seattle, WA, USA; Public Health Sciences Division, Fred Hutchinson Cancer Center; Seattle, WA, USA; Biostatistics Research Branch, National Institute of Allergy and Infectious Diseases, National Institutes of Health; Bethesda, MD, USA; Clinical Monitoring Research Program Directorate, Frederick National Laboratory for Cancer Research, Frederick, MD, USA; Department of Biostatistics, University of Washington; Seattle, WA, USA; Department of Biostatistics and Bioinformatics, Rollins School of Public Health, Emory University; Atlanta, GA, USA; Department of Statistics, University of Washington; Seattle, Washington, USA; Moderna, Inc.; Cambridge, MA, USA; Biomedical Advanced Research and Development Authority; Washington, DC, USA; Division of Infectious Diseases, Department of Medicine, University of Cincinnati; Cincinnati, Ohio, USA; Department of Pathology, Microbiology and Immunology, Vanderbilt University Medical Center; Nashville, TN, USA; Department of Medicine, Division of Infectious Diseases, UNC HIV Cure Center, University of North Carolina at Chapel Hill School of Medicine; Chapel Hill, NC, USA; Department of Laboratory Medicine and Pathology, University of Washington; Seattle, WA, USA; Center for Vaccine Development and Global Health, University of Maryland School of Medicine; Baltimore, MD, USA; Department of Molecular Virology and Microbiology, Baylor College of Medicine; Houston, TX, USA; Brigham and Women’s Hospital; Boston, MA, USA; Vaccine Research Center, National Institute of Allergy and Infectious Diseases, National Institutes of Health; Bethesda, MD, USA

## Abstract

In the coronavirus efficacy (COVE) phase 3 efficacy trial of the mRNA-1273 vaccine, IgG binding antibody (bAb) concentration against Spike (BA.1 strain) and neutralizing antibody (nAb) titer against Spike (BA.1 strain) pseudovirus were assessed as correlates of risk of Omicron COVID-19 and as correlates of relative boost efficacy in per-protocol recipients of a third (booster) dose. Markers were measured on the day of the boost (BD1) and 28 days later (BD29). For SARS-CoV-2 naive individuals, BD29 Spike IgG-BA.1 strain bAbs and BD29 BA.1-strain nAbs inversely correlated with Omicron COVID-19: hazard ratio (HR) per 10-fold marker increase [95% confidence interval (CI)] = 0.16 (0.03, 0.79); P=0.024 and 0.31 (0.10, 0.96); P = 0.042, respectively. These markers also inversely correlated with Omicron COVID-19 in non-naive individuals: HR = 0.15 (0.04, 0.63); P = 0.009 and 0.28 (0.07, 1.08); P = 0.06, trend. Fold-rise in markers from BD1 to BD29 had similarly strong inverse correlations. For SARS-CoV-2 naive individuals, overall booster relative (three-dose vs two-dose) efficacy was 46% (95% CI: 20%, 64%) and correlated with BA.1 strain nAb titer at exposure. At 56, 251, and 891 arbitrary units (AU)/ml (10^th^, 50^th^, and 90^th^ percentile), the booster relative efficacies were −8% (95% CI: −126%, 48%), 50% (25%, 67%), and 74% (49%, 87%), respectively. Similar relationships were observed for Spike IgG-BA.1 strain bAbs and for the markers measured at BD29. The performance of bAb and nAb markers as correlates of protection against Omicron COVID-19 supports their continued use as surrogate endpoints for mRNA vaccination against Omicron COVID-19.

## Main Text

The COVE trial (*1, 2*) (NCT04470427) was a randomized, placebo-controlled phase 3 trial of the mRNA-1273 vaccine, which encodes prefusion-stabilized full-length Ancestral SARS-CoV-2 Spike (index strain, MN908947.3). Estimated vaccine efficacy (VE) against virologically confirmed COVID-19 (hereafter, “COVID-19”) in baseline SARS-CoV-2 negative participants was 93.2% [95% confidence interval (CI): 91.0%, 94.8%] over ~5 months of follow-up in those who received injections on Days 1 and 29 (D1, D29) in a per-protocol analysis (*2*). We previously applied multiple statistical frameworks to assess several immune markers including serum IgG binding antibodies (bAbs) against Spike and neutralizing antibodies (nAbs), measured on D57, as correlates of risk (CoRs) and correlates of protection (CoPs) against COVID-19 through ~4 months post dose 2 (*3, 4*). Using antibody decay models we also demonstrated that the nAb titer at the time of exposure correlated with COVID-19 (*5*). In all these analyses, the markers were measured against Ancestral Spike: bAbs against index (vaccine-insert) Spike and nAbs against index Spike with the D614G mutation (for simplicity, we refer hereafter to both of these as “Ancestral”). Each marker was shown to be a CoR and CoP of COVID-19, with strongest evidence for the nAb markers. During the follow-up time over which the markers were assessed, predominantly Ancestral SARS-CoV-2 lineages and minor genetic drift lineages circulated at the trial sites (*6*). Based on these and other analyses, nAb titer has been used as a surrogate endpoint for regulatory authorization or approval of booster doses and variant vaccines, accelerating decisions compared to requiring large phase 3 clinical trials (*7*).

In late 2021, the Omicron variant spread rapidly in southern Africa (*8*), becoming dominant in the United States by December 2021 (*9*) and with successive waves of Omicron subvariants dominating globally by mid-2022 (*10, 11*). All Omicron subvariants have demonstrated some level of immune escape, especially from neutralization by antibodies elicited by natural SARS-CoV-2 infection and/or COVID-19 vaccination (*12–19*). This raises the question of whether the above antibody markers, which were assessed against Ancestral strains, are also CoRs and CoPs against Omicron COVID-19. To address this knowledge gap, we measured nAb markers against Omicron (BA.1 strain) Spike pseudovirus and bAb markers against Omicron (BA.1 strain) Spike. Using follow-up data from the COVE trial extending through April 5, 2022 (past the unblinding stage and post-dose three, which was a booster dose), here we studied four measurements of these markers as CoRs and CoPs against Omicron COVID-19: (i) measured on the day of the booster dose (BD1), (ii) measured 28 days later (BD29), (iii) fold-rise from BD1 to BD29, and (iv) predicted at the time of exposure (COVID-19 illness visit). The overall objective of this work was to see if bAb and nAb markers continue to be supported as CoPs.

In assessing the antibody markers (i)-(iv) as correlates, four objectives were assessed. Objective 1 was to compare the four BA.1 strain correlates noted above to corresponding Ancestral strain correlates, toward understanding the significance of variant-matching. Objective 2 was to compare the BA.1 antibody/Omicron COVID-19 outcome relationship with the Ancestral antibody/Ancestral COVID-19 outcome relationship and see if similar antibody levels are associated with similar reductions in COVID-19 risk. This question was previously investigated by Cromer et al. (*20*) using a population-based approach, and we investigate the question here through a complementary approach consisting of individual-breakthrough analysis of the COVE trial.

All our previous COVE correlates analyses (*3, 4, 21, 22*) were conducted in baseline SARS-CoV-2 negative participants. Given that CoPs may be modified by prior infection, as well as the need to understand CoPs in previously infected persons given the vast majority of persons are seropositive (*23, 24*), another gap is knowledge of whether and how CoPs differ in non-naive compared to SARS-CoV-2 naive individuals. Objective 3 addressed this gap by studying immune correlates in COVE in both groups and testing modification of the correlates by prior infection. There is some evidence that baseline immune status influences COVID-19 vaccine protection, for example, Hertz et al. assessed IgG, IgA binding and neutralization titers at baseline in healthy individuals who received a fourth dose of BNT162b2 versus those who did not and showed that low baseline antibody responses correlated with higher risk of COVID-19 in both groups (*25*). In a large study, longitudinally measured anti-Spike IgG antibody strongly correlated with both Omicron BA.4/5 infection and disease with stronger and more durable protection seen in those vaccinated with prior infection compared to vaccinated without infection (*26*). Marking et al. showed that baseline Spike IgG, IgA, and neutralization titers in triple-vaccinated healthcare workers correlated inversely with omicron infection measured by serial PCR tests (*27*). BD1 antibody markers were assessed as correlates as noted above. Objective 4 considered all of the data together, assessing through multivariable statistical learning how to best predict Omicron COVID-19 based on the different types of variables among baseline factors, time point (BD1, BD29 absolute level, fold-rise), immunoassay (binding, neutralization), and immunologic assay strain (BA.1, Ancestral).

### Trial schema and participant demographics

**Figure S1** shows a schematic timeline of the mRNA-1273 vaccine doses, where the time interval between the second dose and the third (booster) dose differed between the original-vaccine arm (median time: 12.9 months) and crossover-vaccine arm (median time: 8.2 months), and the sampling days. **Figure S2** shows participant flow from enrollment through to the sampling population for the correlates analysis, which was a subset of COVE participants in the primary series per-protocol cohort (SARS-CoV-2 negative prior to receipt of two doses of mRNA-1273 and with no major protocol deviations) who received a third dose of mRNA-1273 vaccine prior to December 31, 2021 (n=15,713). Participants with SARS-CoV-2 infection diagnosis in the placebo arm prior to mRNA-1273 vaccination were excluded. **Figure S2** continues by showing participant flow through to the per-protocol boosted cohort for this study (n=14,251) and, via stratified case-control sampling (see Methods), the per-protocol three-dose correlates cohort (n=218; BD1 and BD29 antibody marker data were measured). Participants who tested SARS-CoV-2 RT-PCR+ at their BD1 visit were excluded. Definitions of the COVID-19 endpoint for the correlates analyses and of SARS-CoV-2 naive vs. non-naive status are given in the Supplementary Material. **Table S1** shows the numbers of participants in the per-protocol three-dose correlates cohort across the strata.

Demographic and clinical information for the per-protocol boosted cohort and the three-dose correlates cohort subset are provided in **Table S2.** Of all participants in the per-protocol boosted cohort, 31% were ≥65 years old, 24% were considered at-risk for severe COVID-19 (defined as having one or more comorbidities associated with elevated risk of severe COVID-19) and 48% had been assigned female sex at birth. Compared to the per-protocol correlates analysis cohort for the blinded-phase COVE correlates analyses (*3, 4*), the per-protocol boosted cohort was similar in age and sex, but lower in baseline risk (24% vs. 40%).

To assess the relative efficacy of the booster dose (three-dose vs. two-dose) and to evaluate antibody markers as correlates of booster protection, a dynamic unboosted and nonrandomized control group was identified consisting of 2753 participants who were in the baseline-negative per-protocol cohort according to the definition in Gilbert et al. (*4*), remained in the study through Dec 1, 2022 and had not received a booster dose by January 31, 2022 (**Figure S3**). For the relative booster efficacy analysis and the booster CoP analyses, this dynamic unboosted control group was compared to the per-protocol boosted cohort.

### Antibody marker response rates and levels

BD1 responses were positive/quantifiable in 100% or nearly 100% (depending on the marker) of per-protocol boosted recipients, across SARS-CoV-2 naives and non-naives, as well as across Omicron cases and non-cases (assay limits in **Table S3**; positive/quantifiable response defined in **Table 1** and **Figure 1**) for the two Ancestral strain markers. Response rates were generally slightly numerically lower for the two BA.1 markers (**Table 1**) compared to the Ancestral markers (**Table S4**), particularly for BA.1 strain nAbs in SARS-CoV-2 naive Omicron cases (84.1%; 95% CI: 69.1%, 92.6%) (**Table 1**). The geometric means (GMs) at BD1 were numerically higher for non-naives vs. SARS-CoV-2 naives, e.g. for Spike IgG-BA.1 strain bAbs [in arbitrary units (AU)/ml] 4406 (2907, 6678) vs. 3353 (2646, 4248) and for BA.1 strain nAbs (in AU/ml) 19.1 (11.7, 31.1) vs. 14.6 (12.1, 17.6) in non-cases (**Table 1**). Similar results were seen for the BD1 Ancestral strain markers (**Table S4**). **Figure 1** provides a visualization of participant-level (SARS-CoV-2 naives and non-naives) BD1 and BD29 BA.1 strain marker levels in non-cases and Omicron cases. Among non-naives, BD1 BA.1 strain marker levels were numerically higher in cases vs. non-cases, with GM ratios higher than 1 but with wide confidence intervals; GM ratios were closer to 1 in SARS-CoV-2 naives [e.g. for BA.1 strain nAbs, GM ratio (95% CI) was 1.48 (0.73, 3.00) in non-naives and 0.78 (0.58, 1.05) in SARS-CoV-2 naives] (**Table 1**). Similar results were seen for the Ancestral strain markers (**Table S4, Figure S4**).

**Fig. 1.**
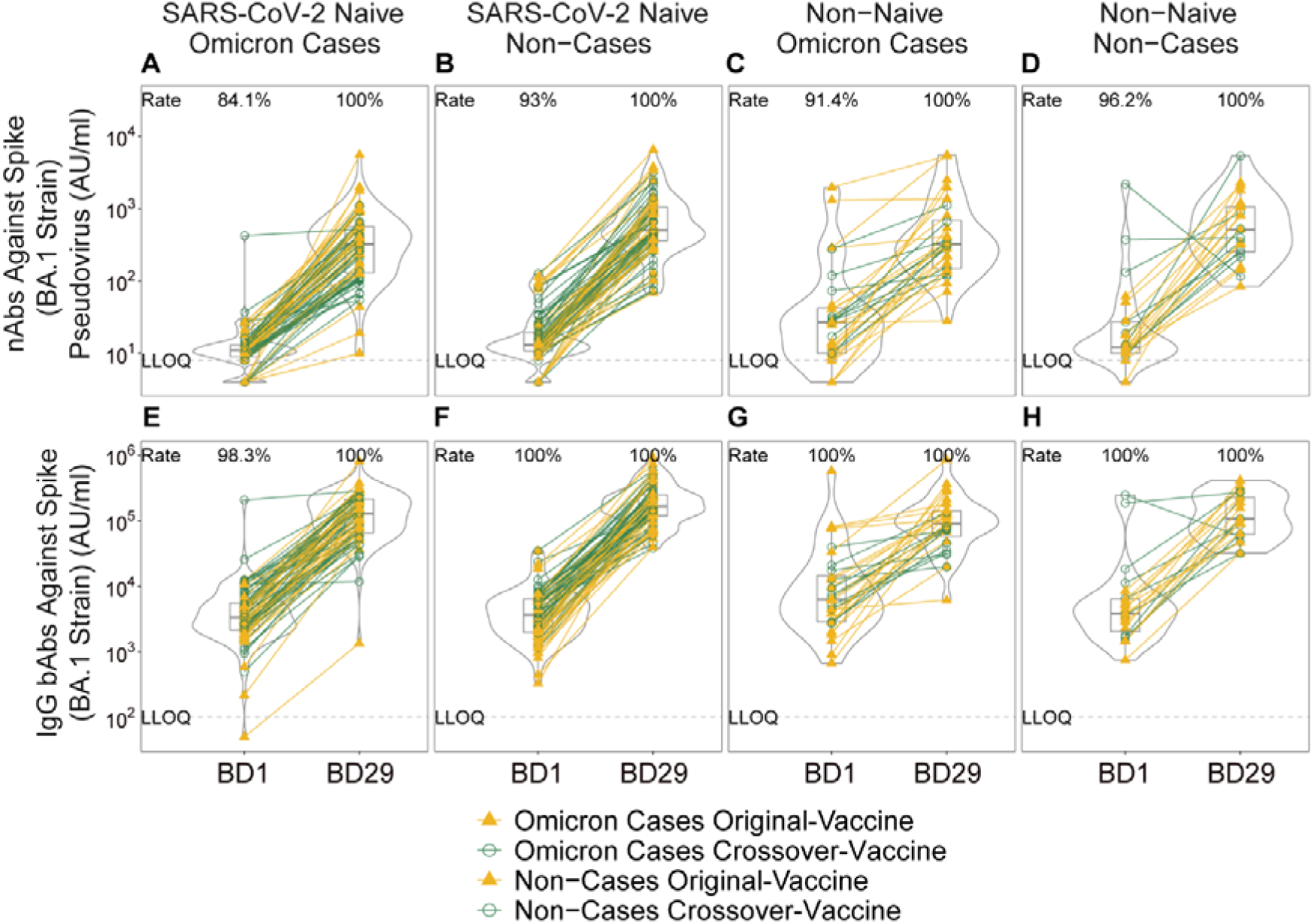
Distributions of BD1 and BD29 (A-D) neutralizing antibody (nAb) titer against Spike (BA.1 strain) pseudovirus and (E-H) IgG binding antibody (bAb) concentration against Spike (BA.1 strain), stratified by Omicron COVID-19 case vs. non-case status and by SARS-CoV-2 naive vs. non-naive status. Data points are from per-protocol boosted participants in the original-vaccine (filled triangles) or crossover-vaccine (open circles) arm, with lines (yellow: original-vaccine arm; green: crossover-vaccine arm) connecting the BD1 and BD29 data points for an individual participant. The violin plots contain interior box plots with upper and lower horizontal edges representing the 25^th^ and 75^th^ percentiles of antibody level and middle line representing the 50th percentile. The vertical bars represent the distance from the 25^th^ (or 75^th^) percentile of antibody level and the minimum (or maximum) antibody level within the 25^th^ (or 75^th^) percentile of antibody level minus (or plus) 1.5 times the interquartile range. Each side shows a rotated probability density (estimated by a kernel density estimator with a default Gaussian kernel) of the data. Positive response rates were computed with inverse probability of sampling weighting. LLOQ, lower limit of quantification. AU/ml, arbitrary units/ml. LLOQ = 8 AU/ml for nAb BA.1 and 102 AU/ml for Spike IgG BA.1. Positive (quantifiable) response for BA.1 strain nAb at a given timepoint was defined by value ≥ LLOQ at that timepoint. Positive response for Spike IgG-BA.1 strain bAb at a given timepoint was defined by value ≥ LLOQ at that timepoint. Omicron Case = COVID-19 endpoint in the interval [≥ 7 days post BD29 AND ≥ December 1, 2021 to April 5, 2022 (data cutoff date)]. Non-case = Did not acquire COVID-19 (of any strain) in the interval [BD1 to April 5, 2022]. SARS-CoV-2 naive (N) = No evidence of SARS-CoV-2 infection from enrollment through to BD1; Non-naive (NN) = Any evidence of SARS-CoV-2 infection in the interval [≥ 14 days after the first two doses of mRNA-1273, BD1].

**Table 1.** BD1, BD29, and Fold-Rise BA.1 strain neutralizing antibody (nAb) and Spike IgG-BA.1 strain binding antibody (bAb) response rates and geometric means by Omicron COVID-19 case vs. non-case status and by SARS-CoV-2 naive vs. non-naive status in the per-protocol boosted cohort, pooled across the original-vaccine and crossover-vaccine arms.

| Omicron Cases<sup>1</sup> |  |  |  |  |  | Non-Cases<sup>2</sup> |  |  |
| --- | --- | --- | --- | --- | --- | --- | --- | --- |
| Status<sup>3</sup> | Marker | Measurement | N<sup>4</sup> | Response Rate<sup>5</sup><br>(95% CI) | GMC or GMT<br>(AU/ml) (95% CI) | N<sup>4</sup> | Response Rate<br>(95% CI)<sup>5</sup> | GMC or GMT<br>(AU/ml) (95% CI) |
| SARS-CoV-2 Naive | BA.1 Strain nAbs | BD1 | 79 | 84.1%<br>(69.1%, 92.6%) | 11.4<br>(9.1, 14.3) | 84 | 93.0%<br>(84.6%, 96.9%) | 14.6<br>(12.1, 17.6) |
| SARS-CoV-2 Naive | Spike IgG-BA.1 Strain bAbs | BD1 | 79 | 98.3%<br>(88.3%, 99.8%) | 3621<br>(2621, 5004) | 84 | 100%<br>(100%, 100%) | 3353<br>(2646, 4248) |
| SARS-CoV-2 Naive | BA.1 Strain nAbs | BD29 | 79 | 100% (100%, 100%) | 259<br>(194, 347) | 84 | 100% (100%, 100%) | 491<br>(341, 706) |
| SARS-CoV-2 Naive | Spike IgG-BA.1 Strain bAbs | BD29 | 79 | 100% (100%, 100%) | 113,143 (87,402, 146,464) | 84 | 100% (100%, 0.0%) | 170,731 (137,772, 211,574) |
| SARS-CoV-2 Naive | BA.1 Strain nAbs | Fold-Rise (BD29/BD1) | 79 | - | 23<br>(17.5, 29.4) | 84 | - | 34<br>(24.6, 46.1) |
| SARS-CoV-2 Naive | Spike IgG-BA.1 Strain bAbs | Fold-Rise (BD29/BD1) | 79 | - | 31.2<br>(24.8, 39.3) | 84 | - | 50.9<br>(42.0, 61.6) |
| Non-Naive | BA.1 Strain nAbs | BD1 | 32 | 91.4%<br>(74.9%, 97.4%) | 28.3<br>(17.0, 47.1) | 23 | 96.2%<br>(74.0%, 99.6%) | 19.1<br>(11.7, 31.1) |
| Non-Naive | Spike IgG-BA.1 Strain bAbs | BD1 | 32 | 100%<br>(100%, 100%) | 7513<br>(4658, 12117) | 23 | 100%<br>(100%, 100%) | 4406<br>(2907, 6678) |
| Non-Naive | BA.1 Strain nAbs | BD29 | 32 | 100% (100%, 100%) | 346<br>(231, 517) | 23 | 100% (100%, 100%) | 572<br>(345, 949) |
| Non-Naive | Spike IgG-BA.1 Strain bAbs | BD29 | 32 | 100% (100%, 100%) | 90,534<br>(63,315, 129,453) | 23 | 100% (100%, 100%) | 148,330<br>(96,969, 226,897) |
| Non-Naive | BA.1 Strain nAbs | Fold-Rise (BD29/BD1) | 32 | - | 12.2<br>(7.6, 19.7) | 23 | - | 30.0<br>(18.3, 49.1) |
| Non-Naive | Spike IgG-BA.1 Strain bAbs | Fold-Rise (BD29/BD1) | 32 | - | 12.0<br>(7.2, 20.0) | 23 | - | 33.7<br>(22.6, 50.2) |

<sup>1</sup>Omicron case = COVID-19 Omicron BA.1 endpoint that occurred in the interval $\geq 7$ days post BD29 AND ≥ December 1, 2021 to April 5, 2022 data cutoff].
<sup>2</sup>Non-case = No acquirement of COVID-19 (of any strain) in the interval [BD1, April 5, 2022 data cutoff].
<sup>3</sup>SARS-CoV-2 Naive = No evidence of SARS-CoV-2 infection from enrollment through to BD1; Non-naive = Any evidence of SARS-CoV-2 infection in the interval $\geq 14$ days after the original two-dose series, BD1]
<sup>4</sup>N is the number of cases sampled into the subcohort within baseline covariate strata.
<sup>5</sup>Definition of “responder” for a marker measured on a given time-point: positive (quantifiable) response defined as BA.1 strain nAb titer on that time-point $\geq 8$ AU/ml; positive response defined as Spike IgG-BA.1 strain bAb concentration on that time-point $\geq 102$ AU/ml.
AU/ml, arbitrary units/ml; CI: confidence interval; GMC: geometric mean concentration; GMT: geometric mean titer

At BD29, 100% of SARS-CoV-2 naive and non-naive participants (Omicron cases and non-cases) had a positive/quantifiable response for each of the four markers (**Table 1**). For all four BD29 markers, GMs were lower in Omicron cases vs. non-cases: e.g. in SARS-CoV-2 naives, the GM ratios were 0.66 (0.47, 0.93) for Spike IgG-BA.1 strain bAbs and 0.53 (0.33, 0.84) for BA.1 strain nAbs (**Table 1**), with similar results for the two Ancestral strain markers (**Table S5**). Similar results were generally obtained in non-naives (**Tables 1, S5**), with the potential exception that Ancestral strain nAb levels were similar between Omicron cases vs. non-cases [GM ratio: 0.96 (0.53, 1.73)] (**Table S5**).

Fold-rise (BD29/BD1) values are given in **Table 1**. The lowest GM fold-rise (95% CI) [12.0 (7.2, 20.0)] was observed for Spike IgG-BA.1 strain bAbs in non-naive Omicron cases, with a very similar fold-rise [12.2 (7.6, 19.7)] in BA.1 strain nAbs in non-naive Omicron cases. The greatest GM fold-rise [50.9 (42.0, 61.6)] was observed for Spike IgG-BA.1 strain bAbs in SARS-CoV-2 naive non-cases (**Table 1**). The same pattern was seen in the fold-rise Ancestral strain markers (**Table S5**). Antibody response rates and geometric means by randomization arm are given in **Table S6**.

### Correlations among antibody markers

In SARS-CoV-2 naives, Ancestral strain bAbs and BA.1 strain bAbs were highly correlated at BD1 (weighted Spearman rank r = 0.90) and at BD29 (r = 0.96); Ancestral strain nAbs and BA.1 strain nAbs were moderately correlated at BD1 (r = 0.63) and highly correlated at BD29 (r = 0.89) (**Figures S7, S8**). Also, in SARS-CoV-2 naives, Ancestral strain bAbs and Ancestral strain nAbs were highly correlated at BD1 (r = 0.94) and BD29 (r = 0.87), while BA.1 strain bAbs and BA.1 strain nAbs were only moderately correlated at BD1 (r = 0.70) and BD29 (r = 0.78) (**Figures S7, S8**). Similar results were seen in non-naives (**Figures S9, S10**).

For all 4 markers, BD1 level and BD29 level were weakly correlated among SARS-CoV-2 naives (r = 0.26 for Spike IgG-Ancestral strain bAbs; r = 0.23 for Spike IgG-BA.1 strain bAbs; r = 0.34 for Ancestral strain nAbs; r = 0.38 for BA.1 strain nAbs) and non-naives (r = 0.23 for Spike IgG-Ancestral strain bAbs; r = 0.28 for Spike IgG-BA.1 strain bAbs; r = 0.32 for Ancestral strain nAbs; r = 0.35 for BA.1 strain nAbs) (**Figures S11, S12**).

### Boost, peak, and fold-rise marker correlates of risk analyses

**Figure S13** shows the estimated Omicron COVID-19 risk (all reported estimates were covariate-adjusted) across a range of levels of BD1 BA.1 strain nAbs and BD1 Spike IgG-BA.1 strain bAbs among SARS-CoV-2 naives and non-naives. Among SARS-CoV-2 naives, BD1 BA.1 strain nAbs had evidence as an inverse correlate of Omicron COVID-19 risk, with estimated risk decreasing with increasing BD1 titer (**Figure S13A**) and a HR per 10-fold increase of 0.31 (95% CI: 0.07, 1.37; P=0.12) (**Figure S13E**). In contrast, there was no evidence of association with Omicron COVID-19 for BD1 Spike IgG-BA.1 strain bAbs among SARS-CoV-2 naives, nor for either BD1 BA.1 marker among non-naives, with hazard ratios (HRs) generally close to 1 (**Figure S13**). Similarly, no evidence of association with Omicron COVID-19 was seen for the Ancestral strain markers at BD1, in both SARS-CoV-2 naives and in non-naives (**Figure S14**).

**Figure 2** shows the results of the same analysis for the BD29 BA.1 strain markers. Estimated risk decreased in both SARS-CoV-2 naives and non-naives with increasing BD29 marker level: For SARS-CoV-2 naives, Omicron COVID-19 risk through 121 days post-dose 3 was 15.6% (95% CI: 11.8%, 55.5%) at BD29 BA.1 strain nAb titer 100 AU/ml compared to 5.1% (1.2%, 8.3%) at BD29 BA.1 strain nAb titer 1000 AU/ml (**Figure 2A**). For non-naives, Omicron COVID-19 risk was 30.6% (4.9%, 95.5%) at BD29 BA.1 strain nAb titer of 100 AU/ml compared to 10.5% (0.5%, 69.8%) at 1000 AU/ml (**Figure 2B**); the risk estimates for non-naïves had much wider confidence intervals compared to those for SARS-CoV-2 naïves because the sample size of the former was much smaller (n = 204 vs. 14,047). Results were similar in both populations for BD29 Spike IgG-BA.1 strain bAbs (**Figure 2C, 2D**). The same analyses repeated with BD29 Spike IgG-Ancestral strain bAbs and Ancestral strain nAbs also yielded similar results (**Figure S15**), as expected due to the high correlation between the BD29 bAb and nAb readouts.

**Fig. 2.**
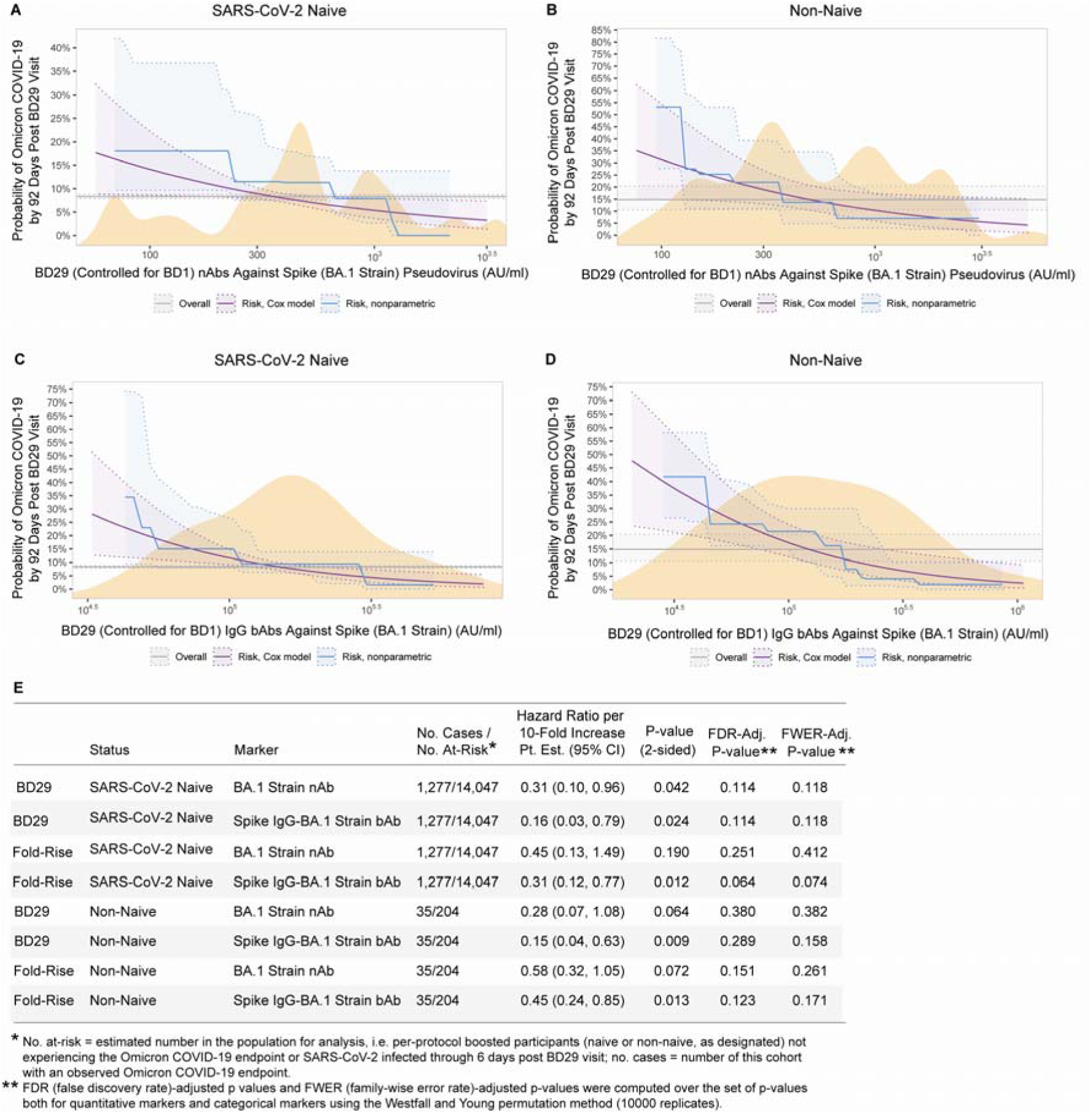
Analyses of BD29 and of Fold-Rise (BD29/BD1) BA.1 strain neutralizing antibody (nAb) titer and Spike IgG-BA.1 strain binding antibody (bAb) concentration as a correlate of risk of Omicron COVID-19. Curves show cumulative incidence of Omicron COVID-19, estimated using a Cox model (purple) or a nonparametric method (blue), in per-protocol boosted (A, C) SARS-CoV-2 naives and (B, D) non-naives by 92 days post BD29 by BD29 antibody marker level. The dotted lines indicate bootstrap pointwise 95% CIs. The horizontal gray line is the overall cumulative incidence of Omicron COVID-19 from 7 to 92 days post BD29 in the per-protocol boosted SARS-CoV-2 naive or non-naive population, as designated. The distribution of the marker in the respective analysis population, calculated by kernel density estimation, is plotted in orange. E) Hazard ratios of Omicron COVID-19 per 10-fold increase in each BD29 and fold-rise (BD29/BD1) BA.1 marker in per-protocol boosted SARS-CoV-2 naives or non-naives. Baseline covariates adjusted for: baseline risk score, at risk status, community of color status, BD1 marker level (paired to the BD29 marker studied).

Among SARS-CoV-2 naives, BD29 BA.1 strain bAbs and BD29 BA.1 strain nAbs were both inversely correlated with Omicron COVID-19 [HR per 10-fold increase (95% CI) = 0.16 (0.03, 0.79); P=0.024 and 0.31 (0.10, 0.96); P = 0.042, respectively] (**Figure 2E**). As expected due to the high correlation between BD29 bAb and nAb readouts, BD29 Spike IgG-Ancestral strain bAbs and BD29 Ancestral strain nAbs exhibited similar HR point estimates and trends toward correlating with Omicron COVID-19, though P-values were not statistically significant at the P<0.05 level [0.23 (0.04, 1.25); P = 0.089 and 0.33 (0.10, 1.13); P = 0.076, respectively)] (**Figure S15E**).

Among non-naives, BD29 Spike IgG-BA.1 strain bAbs inversely correlated with Omicron COVID-19 [HR = 0.15 (0.04, 0.63); P = 0.009] (**Figure 2E**). BD29 BA.1 strain nAbs trended toward correlating with Omicron COVID-19 [0.28 (0.07, 1.08); P = 0.06] (**Figure 2E**). BD29 Spike IgG-Ancestral strain bAbs also inversely correlated with Omicron COVID-19 [HR = 0.10 (0.01, 0.68); P = 0.019] whereas the result for BD29 Ancestral strain nAbs had a wide confidence interval without evidence of a correlation [HR = 0.45 (0.07, 2.95); P = 0.41] (**Figure S15E**).

Inverse correlations with Omicron COVID-19 were also seen for the fold-rise markers (**Figure 2E**; **Figures S16** and **S17**). Among SARS-CoV-2 naives, the HR per 10-fold increase in fold-rise from BD1 to BD29 in Spike IgG-BA.1 strain bAbs was 0.31 (0.12, 0.77); P=0.012, and in BA.1 strain nAbs it was 0.45 (0.13, 1.49); P=0.19 (**Figure 2E**). In non-naives, results were similar [HR = 0.45 (0.24, 0.85); P=0.013 for Spike IgG-BA.1 strain bAb fold-rise and 0.58 (0.32, 1.05); P=0.072 for BA.1 strain nAb fold-rise] (**Figure 2E**). Similar results were seen in both populations for the Ancestral fold-rise markers (**Figure S17**).

Next, a monotone nonparametric threshold regression method was used to assess cumulative incidence of Omicron COVID-19 across subgroups of per-protocol boosted participants defined by marker level exceeding a given threshold. In SARS-CoV-2 naives, the estimated cumulative incidence sharply decreased as the BD29 BA.1 strain nAb titer threshold increased, with estimated cumulative incidence = 9.39% (6.19%, 12.6%) above threshold 10 AU/ml; 5.65% (2.17%, 9.12%) above threshold 500 AU/ml; and 2.97% (0%, 6.06%) above threshold 1000 AU/ml (**Figure S18A**). In non-naives, the decrease in cumulative incidence was less steep with increasing BD29 BA.1 strain nAb titer threshold and the confidence intervals were much wider, due to the smaller sample size (**Figure S18B**). For BD29 Spike IgG-BA.1 strain bAbs, cumulative incidence also decreased with increasing threshold in both SARS-CoV-2 naives and non-naives, again with wider confidence intervals in non-naives (**Figure S18C, D**). Similar threshold-response relationships were also observed for the BD29 Ancestral markers (**Figure S19**) and for the BA.1 strain and Ancestral strain fold-rise markers (**Figures S20** and **S21**, respectively).

**Figure 3** shows Cox-model-based marginalized COVID-19 cumulative incidence curves among SARS-CoV-2 naives and non-naives for subgroups of boosted participants defined by tertile of BD29 Spike IgG-BA.1 strain bAbs and BA.1 strain nAbs. These curves also supported each marker as an inverse correlate of Omicron COVID-19 risk. Results for the equivalent analysis of the BD29 Ancestral markers and for the fold-rise markers are shown in **Figures S22-S24**, with HRs across tertiles reported for the BD1, BD29, and fold-rise markers in **Tables S7-S9**. HRs (High vs. Low tertile) for the BA.1 strain markers indicating inverse correlations included BD29 BA.1 strain nAbs in SARS-CoV-2 naives [0.27 (0.09, 0.78); P = 0.016] (**Table S8**), fold-rise Spike IgG-BA.1 Strain bAbs in SARS-CoV-2 naives [0.35 (0.13, 0.90); P = 0.030] (**Table S9**), and fold-rise Spike IgG-BA.1 Strain bAbs in non-naives [0.23 (0.07, 0.75); P = 0.015] (**Table S9**).

**Fig. 3.**
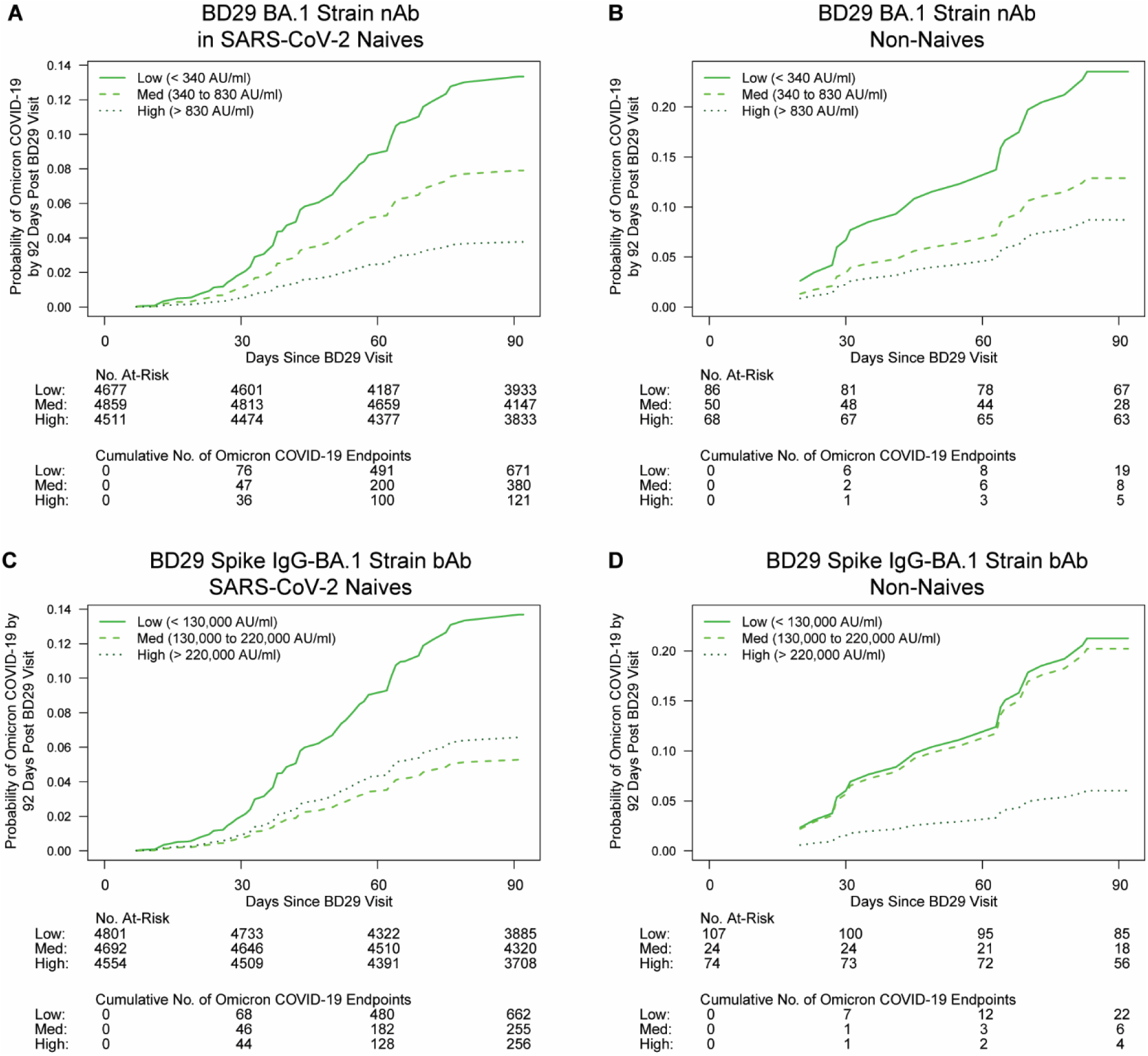
Cox-model-based marginalized Omicron COVID-19 cumulative incidence curves for subgroups of per-protocol boosted (A, C) SARS-CoV-2 naive or (B, D) non-naive participants defined by BD29 BA.1 strain antibody tertile. A, B: BD29 BA.1 strain neutralizing antibody (nAb); C, D: BD29 Spike IgG-BA.1 strain binding antibody (bAb). No. at risk = estimated number in the population for analysis, i.e. per-protocol (A, C) SARS-CoV-2 naive or (B, D) non-naive boosted participants not experiencing the Omicron COVID-19 endpoint or SARS-CoV-2 infected through 6 days post BD29 visit. Analyses were adjusted for baseline risk score, at-risk status, and community of color status.

Based on analysis of BD1 and BD29 antibody markers in the same model, for SARS-CoV-2 naive participants **Figure S25** shows the estimated Omicron COVID-19 risk across the range of BD29 Spike IgG-BA.1 strain bAb levels when fixing the BD1 Spike IgG-BA.1 strain bAb level to the 15^th^, 50^th^, and 85^th^ percentile. At each BD1 Spike IgG-BA.1 strain bAb percentile, COVID-19 risk decreased as BD29 Spike IgG-BA.1 strain bAb level increased, and there was no evidence of a different BD29 Spike IgG-BA.1 strain bAb association with Omicron COVID-19 by BD1 Spike IgG-BA.1 strain bAb level (interaction p > 0.20). Analyses repeated with the BA.1 strain nAb marker and among non-naives exhibited similar patterns (**Figures S26-S28**).

### Predicted-at-exposure antibody correlates of booster relative efficacy among SARS-CoV-2 naives

In SARS-CoV-2 naives, receipt of a third dose was estimated to provide a 46% (95% CI: 20%, 64%) relative reduction in Omicron COVID-19 throughout follow-up compared to an unboosted (two-dose recipient) control group. Using these unboosted participants as a dynamic control group, we analyzed time-varying predicted antibody levels where the instantaneous risk of Omicron COVID-19 on any given day depends on the predicted antibody level on that day using a Cox model with calendar time index [see Methods and (*5*)]. Model-predicted values on the day of disease onset correlated well with the actual antibody readouts on the day of disease onset (**Figures S29, S30**). Within the boosted, Omicron COVID-19 risk correlated similarly with predicted-at-exposure BA.1 strain nAbs and with BD29 BA.1 strain nAbs, with associated HRs (95% CIs) per 10-fold increase of 0.31 (0.12, 0.79) and 0.26 (0.09, 0.72), respectively. Results were similar for the bAb readout, with analogous HRs (95% CIs) of 0.24 (0.06, 0.88) for predicted-at-exposure Spike IgG-BA.1 strain bAbs and 0.21 (0.06, 0.74) for BD29 Spike IgG-BA.1 strain bAbs.

**Figure 4** provides correlates of booster relative efficacy curves for predicted-at-exposure nAb by contrasting the antibody association in the boosted participants with the overall hazard of the unboosted as a reference group. For boosted recipients with BA.1 strain nAb titers of 56 AU/ml (10^th^ quantile), 251 AU/ml (median) and 891 AU/ml (90^th^ quantile) at exposure, the proportion reduction in COVID-19 risk for booster relative (three-dose vs. two-dose) efficacy is −8% (95% CI −126%, 48%), 50% (95% CI 25%, 67%) and 74% (95% CI 49%, 876%) (**Figure 4A**).

**Fig. 4.**
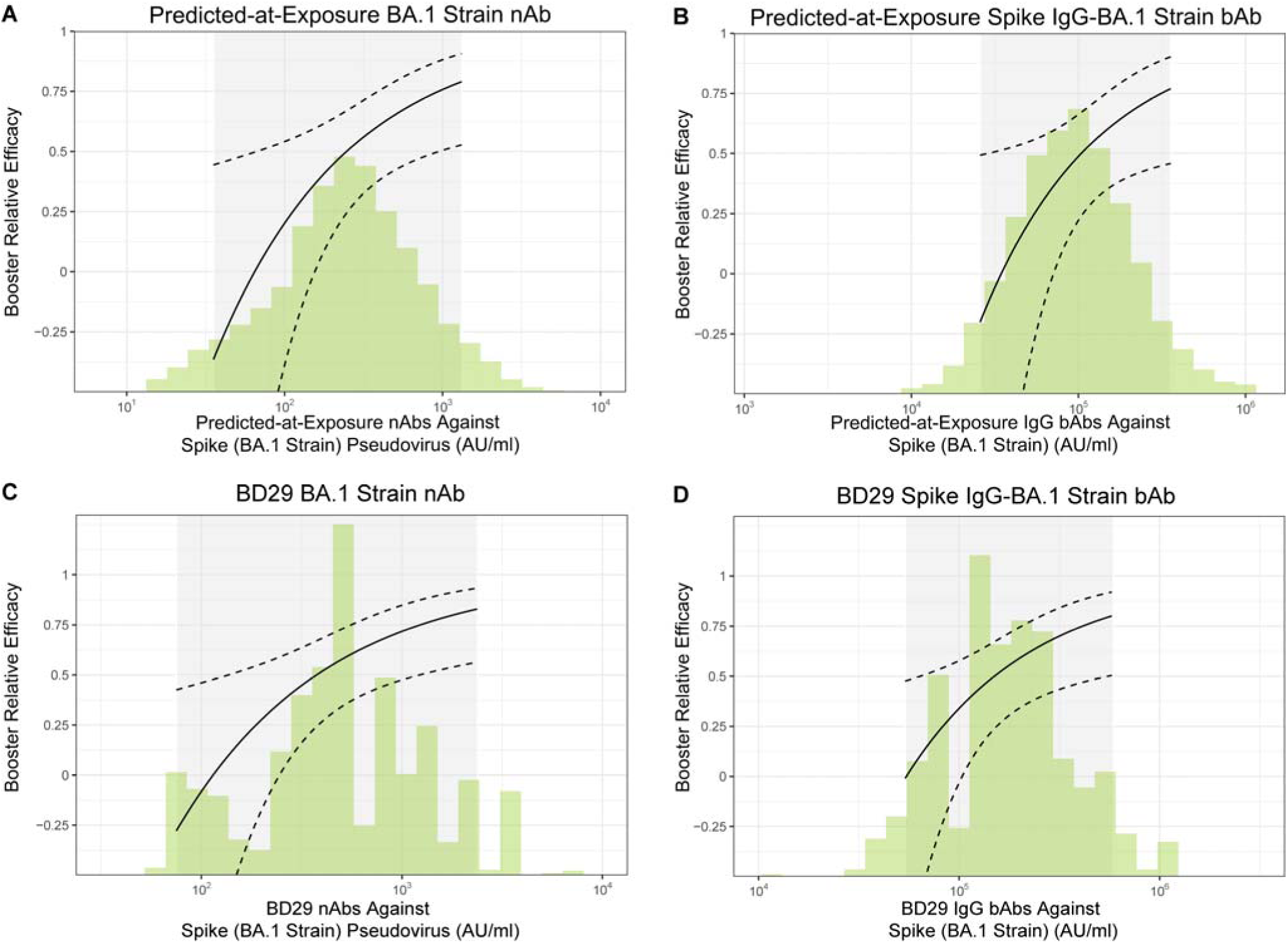
Correlate of booster relative efficacy curves against Omicron COVID-19 among SARS-CoV-2 naives as a function of predicted antibody level at exposure and measured antibody level at BD29. A) Predicted-at-exposure neutralizing antibody (nAb) titer against Spike (BA.1 strain) pseudovirus, B) Predicted-at-exposure IgG binding antibody (bAb) concentration against Spike (BA.1 strain), C) BD29 BA.1 strain nAb, D) Spike IgG-BA.1 strain bAb. The curves show the relative efficacy of three-dose mRNA-1273 vs. two-dose mRNA-1273. The dashed black lines are 95% confidence intervals. The green histograms are an estimate of the density of (A, B) predicted-at-exposure and (C, D) BD29 antibody marker level in per-protocol boosted SARS-CoV-2 naives.

Booster relative vaccine efficacy results for BD29 BA.1 strain nAb titer were similar to the above predicted-at-exposure results (**Figure 4C**) [booster relative efficacy = −7% (95% CI −113%, 46%) for 102 AU/ml (10^th^ quantile); 56% (95% CI 33%, 72%) for 479 AU/ml (median); 80% (95% CI 54%, 91%) for 1,738 AU/ml (90^th^ quantile)]. Analyses repeated with Spike IgG-BA.1 strain bAbs, Ancestral strain nAbs, and Spike IgG-Ancestral strain bAbs yielded similar results (**Figure 4B, 4D**; **Figure S31**). Analogous analyses with an unboosted control for the non-naives were not possible due to extreme confounding, specifically, different events tended to define a participant as non-naive in the boosted vs in the unboosted group. Among the boosted, participants generally became non-naive due to asymptomatic infections in Spring 2021, whereas among the unboosted, participants generally became non-naive due to COVID-19 in Fall 2021 (**Figure S32**).

### Comparison to Ancestral strain correlates study

It is of interest to ascertain whether a different amount of variant-matched antibody is needed for high-level three-dose vs. two-dose protection than for high-level two-dose vs. placebo protection. To answer this question, the Ancestral antibody/Ancestral COVID-19 VE curve estimated previously in baseline-negatives (*4*) can be compared with the BA.1 strain antibody/Omicron COVID-19 booster vaccine efficacy curve in SARS-CoV-2 naive participants. We scaled the BD29 Ancestral strain nAb marker to predict the BD29 BA.1 strain nAb marker such that the units can be absolutely quantitatively interpreted vs. the previous Ancestral antibody units (see Methods), hence enabling a comparison of the two vaccine efficacy curves in **Figure 5**. The results show that post dose 2 Ancestral strain nAb titer of 100 IU50/ml and 1000 IU50/ml is associated with 92% and 96% vaccine efficacy against Ancestral COVID-19, respectively, compared to post dose 3 scaled-Ancestral-strain-nAb-to-predict-BA.1-strain-nAb titer of 100 IU50/ml and 1000 IU50/ml associated with 21% and 77% booster relative efficacy against Omicron COVID-19, respectively. Forthcoming results from a BA.1 strain concordance study between the two neutralization assays in the previous and current correlates studies will enable greater accuracy for comparing the two protection curves in absolute assay units.

**Fig. 5.**
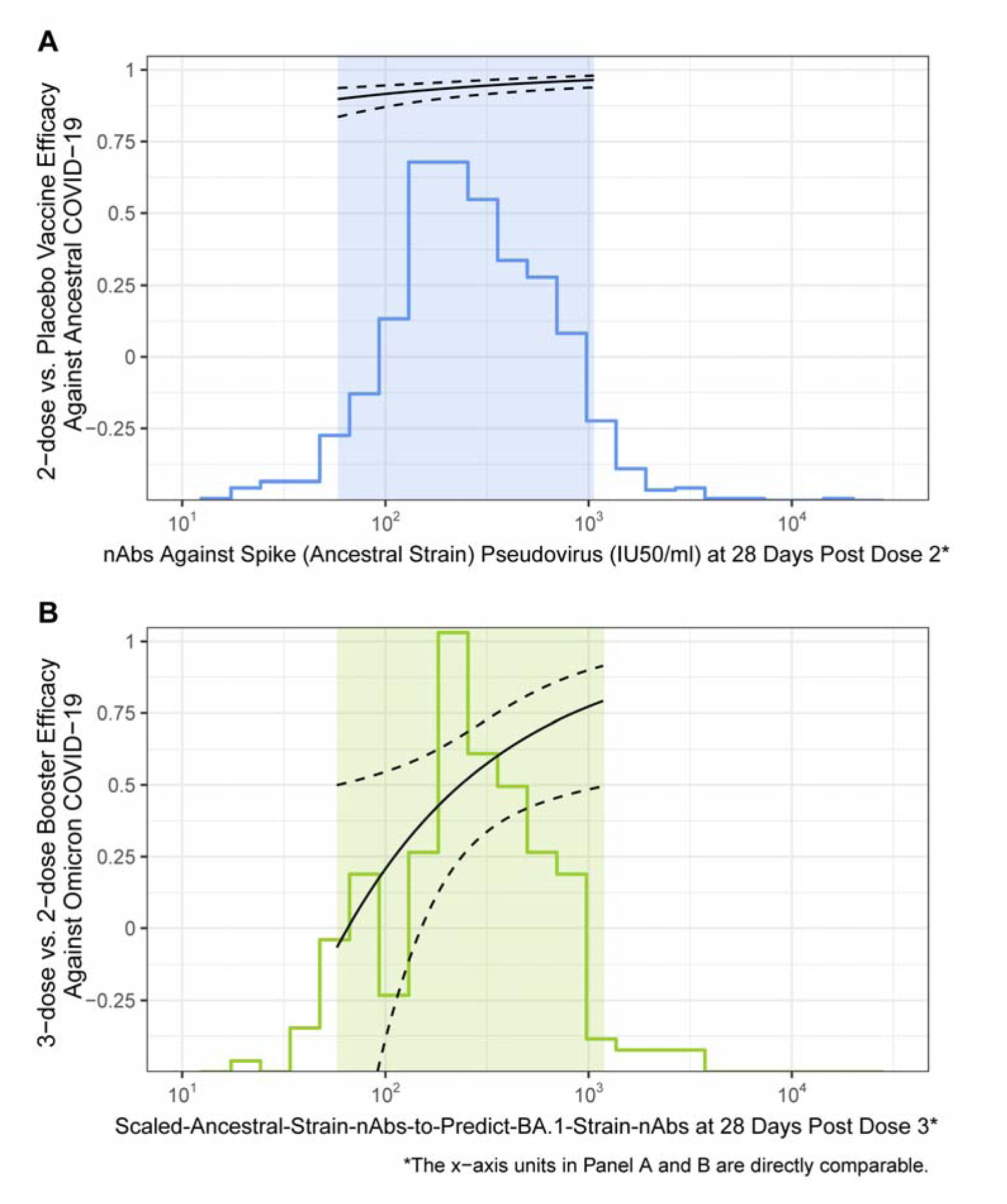
Matched neutralizing antibody, COVID-19 vaccine efficacy curves for Ancestral and Omicron eras among SARS-CoV-2 naive participants. (**A**) The solid curve graphs two-dose vs. placebo vaccine efficacy against Ancestral COVID-19 versus Day 57 (28 days post dose 2) Ancestral strain neutralizing antibody (nAb) titer in International Units (IU50/ml). (**B**) The solid curve graphs three-dose vs. two-dose booster relative efficacy against Omicron COVID-19 versus BD29 (28 days post dose 3) Scaled-Ancestral-strain-nAb-to-predict-BA.1-strain-nAb titer, with absolute scale comparable to the vaccine efficacy curve shown in panel (A) (see Methods). Solid lines are point estimates, dashed lines 95% confidence intervals. The blue histogram shows the distribution of post dose 2 Ancestral strain nAb titer and the green histogram the distribution of post dose 3 Scaled-Ancestral-strain-nAb-to-predict-BA.1-strain-nAb titer.

### Using observational cohort data to attempt to infer an unvaccinated group for comparison to the boosted (three-dose) cohort

A major difference between the two curves in **Figure 5** is their control groups: the Ancestral curve uses an unvaccinated group, whereas the Omicron curve uses a vaccinated (two-dose) but unboosted group. These curves all have the advantage of using data from the randomized COVE phase 3 efficacy trial. Due to the fact that the vast majority of participants originally assigned placebo were offered vaccine after participant unblinding, thus creating the crossover-vaccine group, no unvaccinated control group is available in the COVE data set for comparison to the boosted (three-dose) group. To venture to infer an unvaccinated group for the Omicron curve we reasoned as follows. The two-dose unboosted control group had a median of ~13 months follow-up between dose 2 and December 2021. Observational data have shown that by 13 months post dose 2, VE (compared to an unvaccinated control) against infection and hospitalization waned to 34% and 62%, respectively [eTable 2 in (*28*)]. At 1000 IU50/ml, the 77% booster relative efficacy translates to 1 − (1−0.77)×(1−VE)×100% = 85% to 91% efficacy relative to an unvaccinated group, after substituting VE with 0.34 and 0.62. [The formula VE(3-dose vs placebo) = 1 − (1 − VE(3-dose, 2-dose))×(1 − VE(2-dose, placebo)) was used here.] The analogous calculation at 100 IU50/ml translates a 21% booster relative efficacy to between 48% to 70% efficacy against an unvaccinated group. However, these results come with the caveats that accompany observational cohort study data as compared to randomized efficacy trial data.

### Predicting Omicron COVID-19 risk

We next applied ensemble machine learning to investigate the predictive power of individual or combinations of antibody markers for predicting the occurrence of Omicron COVID-19 in SARS-CoV-2 naive and non-naive per-protocol boosted participants, with objectives to identify best predictive models and to evaluate which component variables (among baseline factors, time point, immunoassay, strain) may be most important for prediction. Classification accuracy of each pre-specified variable set was quantified by the cross-validated area under the receiver operating characteristic (ROC) curve (CV-AUC) and its 95% CI for the discrete Super Learner model. All models included baseline risk factors (risk score, at risk status, community of color status). Results are shown in **Tables S10** and **S11**. Among SARS-CoV-2 naives, baseline risk factors alone had negligible predictive power [CV-AUC = 0.517 (0.404, 0.628)]. Models that additionally included subsets of antibody markers (nAb and bAb against the Ancestral and BA.1 strains at BD1 and BD29, and fold-rise from BD1 to BD29) were benchmarked against this baseline risk factors reference model in order to quantify their contribution towards improving prediction accuracy. Among models that included only antibody markers at BD1, the model that included both nAb and Spike IgG bAb against the BA.1 strain performed the best with CV-AUC of 0.641 (95% CI 0.552, 0.721).

Models that included BD29 antibody markers generally outperformed the BD1-only marker models, where the single-marker model with BD29 BA.1 strain nAb performed best [CV-AUC = 0.677 (0.591, 0.753)]. The corresponding model with BD29 Ancestral strain nAbs had a lower CV-AUC of 0.659 (0.572, 0.737). Among multivariable marker models, a model with BD29 IgG Spike bAb and nAb markers against both the Ancestral and BA.1 strains achieved a CV-AUC of 0.685 (0.599, 0.760). Replacing the BD29 markers in this model with the fold-rise markers resulted in lower prediction accuracy [CV-AUC = 0.601 (0.512, 0.685)]. Including the BD1 IgG Spike bAb and nAb markers against both the Ancestral and BA.1 strains on top of the BD29 markers marginally improved performance [CV-AUC = 0.686 (0.600, 0.761)].

Among non-naives, the reference model including baseline risk factors alone had higher CV-AUC point estimate [CV-AUC =0.561 (0.389, 0.720)] compared to the equivalent model in SARS-CoV-2 naives. Among all BD1-only marker models, the best performing model included anti-receptor binding domain (RBD) IgG Ancestral strain bAbs, Ancestral strain nAbs, and BA.1 strain nAbs and showed improved performance over the reference model [CV-AUC = 0.621 (0.465, 0.756)]. Models that included only one BD29 antibody marker generally performed worse compared to their counterparts among SARS-CoV-2 naives, where the best-performing model included BD29 RBD IgG-Ancestral strain bAbs [CV-AUC = 0.577 (0.411, 0.728)]. Models that included one variant-matching BD29 marker had negligible predictive power given their CV-AUC was lower than the model with baseline risk factors alone [CV-AUC = 0.559 (0.389, 0.719) for BA.1 strain nAbs; CV-AUC = 0.560 (0.397, 0.712) for Spike IgG-BA.1 strain bAbs]. Replacing the BD29 bAb marker with its corresponding fold-rise marker improved the prediction performance, e.g., CV-AUC = 0.651 (0.496, 0.780) for fold-rise of RBD IgG-Ancestral strain bAbs vs. 0.577 (0.411, 0.728) for absolute level. On the other hand, replacing the BD29 nAb marker with the fold-rise marker appeared to make a smaller difference [CV-AUC = 0.575 (0.408, 0.727) vs. 0.559 (0.389, 0.719) for BA.1 strain nAbs; CV-AUC = 0.586 (0.428, 0.728) vs 0.558 (0.377, 0.727) for Ancestral strain nAbs].

For non-naives, multivariable marker models generally performed better than single marker models. Models with a combination of BD29 bAb and nAb markers against the Ancestral strain were among the best-performing models; for instance, a model including RBD IgG-Ancestral strain bAbs and Ancestral strain nAbs achieved the highest predictive power with a CV-AUC = 0.712 (0.558, 0.829) and a model including Spike IgG-Ancestral strain bAbs and Ancestral strain nAbs achieved a CV-AUC of 0.656 (0.500, 0.784). Replacing BD29 markers in the multivariable models with corresponding fold-rise markers or both the BD1 and fold-rise markers generally resulted in slightly inferior performances. Further including the variant-matching markers against the BA.1 strain did not increase, and in many circumstances, decreased the prediction performance.

In summary, among SARS-CoV-2 naives, the multivariable learning supported prediction advantages of variant-matching over Ancestral strain antibody and BD29 absolute level over fold-rise, an advantage of nAb assay over bAb assay in parsimonious models with no improvement by including both assays, and no augmented predictive value of including BD1 markers in addition to BD29 markers. Among non-naives, the multivariable learning supported prediction advantages using a combination of BD29 bAb and nAb levels against the Ancestral strain over using any individual BD29 bAb or nAb levels and no added predictive value of further including variant-matching markers.

## Conclusions

In this paper we provide an extensive analysis of the correlation between antibody response to a third dose of mRNA-1273 and Omicron COVID-19 risk during the initial Omicron wave in the COVE trial. Strengths include use of a randomized clinical trial cohort with rigorous follow-up, careful virologic and symptom sampling, use of validated assays, and standardized analyses that facilitate comparison with the Gilbert et al. correlates analysis (*4*) following second dose during the Ancestral strain era of the same cohort. We find that antibody measured 28 days post third dose consistently correlates with Omicron COVID-19 risk, supporting its continued use as a surrogate endpoint for regulatory decision making. For the BA.1 strain nAb assay, BD29 titer was estimated to have a hazard ratio of 0.31 and 0.28 for SARS-CoV-2 naives and non-naives, which corresponds to a 69% and 72% lower Omicron COVID-19 risk in participants with a 10-fold higher BD29 titer. This association is similar to what was estimated in Gilbert et al. (*4*) for SARS-CoV-2 naives, with a hazard ratio of 0.42 (95% CI: 0.27, 0.65) for an Ancestral strain nAb assay during the Ancestral era. Consistent associations were seen with the bAb assays and with alternate statistical methods of analysis.

In addition to ‘peak’ BD29 antibody, we evaluated BD1 and BD29/BD1 fold rise as correlates of Omicron COVID-19. BD1 antibody, measured at a median of 11 months post 2^nd^ dose, was poorly correlated with BD29 antibody and generally did not correlate with Omicron COVID-19 except BA.1 strain nAbs inversely correlated with COVID-19 and multivariable models of non-naives suggested inverse correlations. In comparison, Hertz et al. measured baseline IgG a median of 6 months from 3^rd^ to 4^th^ Pfizer mRNA vaccine dose and showed individuals with low baseline IgG to index-strain receptor binding domain or index-strain S2 had significantly higher risk of COVID-19 during the Omicron wave of early/mid 2022 (*25*). BD1 antibodies might have been more predictive had boosting occurred closer to the 2^nd^ dose. For Spike IgG-BA.1 strain bAbs and BA.1 strain nAbs, the HR p-values for BD29 and BD29/BD1 fold-rise were mostly similar in our analysis (in SARS-CoV-2 naives: P = 0.024 and 0.012, respectively, for Spike IgG-BA.1 strain bAbs and P = 0.042 and 0.19, respectively for BA.1 strain nAbs; in non-naives P=0.009 and 0.013, respectively, for Spike IgG-BA.1 strain bAbs and P=0.064 and 0.072 for BA.1 strain nAbs), indicating similar levels of evidence.

In our analysis we focused on the BA.1 strain assays as the BA.1 strain Spike antigen and BA.1 strain Spike pseudovirus used in the bAb and nAb assays, respectively, were well-matched to the BA.1 Spike from the SARS-CoV-2 BA.1 strain that caused the Omicron COVID-19 cases accrued during follow-up. In contrast, the Ancestral strain Spike antigen and Ancestral strain Spike pseudovirus used in the Ancestral strain bAb and nAb assays were highly similar to most SARS-CoV-2 variants (i.e., only minor genetic drift from Ancestral) (*6*) that were circulating at the COVE trial sites during the blinded follow-up period. Unsurprisingly, the BD29 Spike IgG-Ancestral strain bAb and BD29 Ancestral strain nAb levels were much higher than the BD29 BA.1 levels for both assays. In SARS-CoV-2 naive Omicron COVID-19 cases, for example, the geometric mean BD29 Ancestral strain nAb titer was 12-fold higher than the geometric mean BD29 BA.1 strain nAb titer (3234 AU/ml vs. 259 AU/ml, numbers from Table S5 and **Table 1**, respectively). In SARS-CoV-2 naive non-cases, the geometric mean BD29 Ancestral strain nAb titer was 11-fold higher than the geometric mean BD29 BA.1 strain nAb titer (5492 AU/ml vs. 491 AU/ml, numbers from **Table S5** and **Table 1**, respectively). However, regardless of this overall difference in the Ancestral strain vs. BA.1 strain assay levels, in SARS-CoV-2 naives the BA.1 strain and Ancestral strain assay readouts are highly correlated and the hazard ratios of Omicron COVID-19 per 10-fold increase for BD29 Spike IgG-BA.1 strain bAbs and BD29 BA.1 strain nAbs were similar to those for BD29 Spike IgG-Ancestral strain bAbs and BD29 Ancestral strain nAbs (0.16 and 0.31; 0.23 and 0.33, respectively; **Figures 2E** and **S15E**). Moreover, the correlate of booster relative efficacy curves were quite similar for BA.1 strain and Ancestral strain nAbs, though shifted to the right. Thus, for SARS-CoV-2 naives, variant-antibody matching to variant COVID-19 may not give a meaningfully better prediction of booster relative efficacy. For non-naives, correlation point estimates suggested variant-antibody matching improved the correlate, albeit with insufficient precision to draw a conclusion.

Our study allowed a comparison of bAbs and nAbs as correlates. nAb assays appealingly measure in vitro function while bAb assays have the advantage of less technical measurement error. Measurements from both assays consistently correlated with Ancestral COVID-19 in SARS-CoV-2 naive participants after primary immunization in multiple studies (*7*). We demonstrated that BA.1 strain bAbs significantly correlated with Omicron COVID-19 with very similar hazard ratios and p-values for SARS-CoV-2 naives and non-naives. For BA.1 nAbs, the relationships for naives and non-naives were again very similar with slightly larger p-values than for bAbs (**Figure 2E**, **Figure S15E**). Our results do not demonstrate that one assay readout is a better Omicron COVID-19 correlate than the other.

Multivariable statistical learning analyses that analyzed the baseline risk factors and antibody markers together as joint predictors of COVID-19 yielded apparently distinct results for SARS-CoV-2 naives and non-naives. For SARS-CoV-2 naives, variant-matching the strain of the assay to the COVID-19 variant improved the correlate, and the absolute level of antibodies at BD29 yielded a better correlate than fold-rise of antibodies to BD29. In contrast, for non-naives there was no evidence that variant-matching improved the correlate, with the best predictive models based on Ancestral strain antibodies and including both RBD IgG bAbs and nAbs. Additionally, for non-naives the data supported better prediction with fold-rise than absolute level BD29 antibodies. One interpretation of these results is that antibody levels perform better as a mechanistic CoP for SARS-CoV-2 naives and more as a non-mechanistic CoP for non-naives. A potential explanation is that SARS-CoV-2 naive mRNA-1273 vaccine recipients have an immune response only to Spike antigen whereas non-naives have an immune response to many viral antigens that spans also additional arms of the immune system, hence lowering the predictive value of markers of vaccine response.

Our study has scope limitations, including: i) our analysis is for BA.1 Omicron COVID-19 and not current variants; ii) while we preferentially sampled those with severe COVID-19, it was too rare to study as a separate endpoint; iii) vaccination was with the Ancestral strain and non-naives predominantly acquired Ancestral strain infection; iv) non-naives were largely defined by asymptomatic infection a median of ~8 months prior to boost rather than symptomatic COVID; v) the sample size for non-naives was relatively small such that the multivariable learning analyses in particular had limited precision such that these results are interpreted as hypothesis generation in need of additional analysis; and vi) T-cell and B-cell responses were not studied, such that the contribution of other immune markers as potential correlates of protection could not be assessed.

We identified about a 50% reduction in Omicron COVID-19 for the boosted group compared to a non-randomized dynamic unboosted control which provides an opportunity to evaluate the association of antibody with boost relative efficacy. For the predicted-at-exposure model in SARS-CoV-2 naives we found that the relative efficacy ranged from approximately −8% to 74% for BA.1 nAbs of 56 AU/ml to 891 AU/ml (the 10^th^ and 90^th^ percentiles of the antibody distribution throughout follow-up), though with substantial uncertainty. At BD1, the mean BA.1 strain nAb titer was about 15 AU/ml, somewhat close to 56 AU/ml, thus roughly approximating the idea that as antibody wanes to approach that of unboosted individuals, relative efficacy wanes.

An important question is the transportability of CoP relationships across new variants. The HR of Omicron COVID-19 in SARS-CoV-2 naive boosted participants per 10-fold increase in “peak” post-boost (BD29) BA.1 strain nAb titer (**Figure 2E** of this work) was relatively similar to the HR of Ancestral COVID-19 in SARS-CoV-2 negative two-dose recipients per 10-fold increase in “peak” post-dose 2 (D57) Ancestral strain nAb titer [**Figure 3A** in (*4*)]: 0.31 (95% CI: 0.10, 0.96; P=0.042) and 0.42 (0.27, 0.65; P<0.001), respectively. A question is whether a given titer from a nAb assay matched to the circulating variant gives the same level of protection across variants — a variant-invariant absolute CoP. In **Figure 5** we showed that Ancestral strain nAb titer of 1000 IU50/ml was associated with a 96% reduction (two-dose vs. placebo) in Ancestral COVID-19, while a BA.1-strain-predicted-nAb titer of 1000 IU50/ml (BA.1) was associated with a 77% reduction (three-dose vs. two-dose) in Omicron COVID-19. However, these reductions are not directly comparable due to the different control groups. Using observational data, we crudely inferred that the booster relative efficacy (3 dose vs 2 dose) of 77% translated to a vaccine efficacy (3 dose vs unvaccinated) of between 85% to 91%, lower than the 96% vaccine efficacy of the Ancestral era. These estimates suggest that higher antibody levels are required to attain high boost protection against Omicron COVID-19 compared to levels required for original two-dose protection against Ancestral COVID-19. These data also suggest levels of neutralization that can be used in early phase studies to guide / be a potential target for high-level efficacy for vaccine strain selection. As noted in **Figure 5**, few persons achieved this level of neutralizing activity to BA.1 after the third dose of mRNA-1273. However, a limitation of this analysis is that the estimates are extrapolated and have statistical uncertainty, given that the vaccine effectiveness estimates used were from an observational study with potential bias in risk behavior, occurrence/timing of a third dose, and antibody response subject to residual confounding from inadequate statistical adjustment. Thus, this work could not definitively answer this important question.

## Supporting information

COVE Post-Dose 3 Omicron COVID-19 Correlates Statistical Analysis Plan

## Data Availability

Access to participant-level data and supporting clinical documents with qualified external researchers may be available upon request and is subject to review once the trial is complete.

## Author contributions

B.Z., Y.F., P.B.G., and D.F. conceptualized the study. F.P., R.D., X.W. also contributed to study concept and design. B.Z., Y.F., J.F., A.K., M.C., P.B.G, and D.F. developed methodology used in the study. B.Z., Y.F., H.E.J., J.F., A.K., M.C., D.B., W.D., H.Z., X.W., Y.L., C.Y., P.B.G., and D.F. curated the data. B.G., F.P., R.D., and X.W. were involved in data collection. B.Z., Y.F., H.E.J., J.F., E.C. A.K., M.C., W.D., H.Z., X.W., Y.L., C.Y., B.B., R.D., P.B.G., and D.F. conducted the analyses. B.Z., Y.F., H.E.J., J.F., A.K., M.C., D.B., W.D., H.Z., X.W., Y.L., C.Y., K.M., L.J., J.M., C.J.F., S.K., C.L.G., M.P.A., J.G.K., L.C., K.M.N., R.P., H.M.E.S., L.R.B., R.O.D., R.A.K., and P.B.G. contributed resources to the project. B.Z., Y.F., J.F., A.K., M.C., D.B., P.B.G, and D.F. developed software used in the analyses. C.R.H., C.H., R.O.D., R.A.K., and P.B.G. performed project administration. B.Z., Y.F., P.B.G., and D.F. validated the analysis results. B.Z., L.N.C., P.B.G., and D.F. wrote the original draft. All coauthors reviewed and edited the draft. ICMJE guidelines for authorship have been adhered to.

## Ethics

The mRNA-1273-P301 study was conducted in accordance with the International Council for Harmonisation of Technical Requirements for Pharmaceuticals for Human Use, Good Clinical Practice guidelines, and applicable government regulations. The Central Institutional Review Board approved the mRNA-1273-P301 protocol and the consent forms. All participants provided written informed consent before enrollment. Central IRB services for the mRNA-1273-P301 study were provided by Advarra, Inc., 6100 Merriweather Dr., Suite 600, Columbia, MD 21044.

All necessary patient/participant consent has been obtained and the appropriate institutional forms have been archived.

## Supplementary Materials

Materials and Methods

Figs. S1 to S34

Tables S1 to S11

Supplementary Text

