## Supplementary material for "Omicron COVID-19 Immune Correlates Analysis of a Third Dose of mRNA-1273 in the COVE Trial": COVE Post-Dose 3 Omicron COVID-19 Correlates Statistical Analysis Plan

### Statistical Analysis Plan for Study of Post Dose 3 and Exposure-Proximal Omicron Antibody as Immune Correlates for Omicron COVID-19 in the P301 COVE Study

USG COVID-19 Response Team / Coronavirus Prevention Network (CoVPN) Biostatistics Team

October 15, 2023

### Contents

**List of Tables** 3

**List of Figures** 4

**1 Outline** 5

**2 Stage 1 correlates sampling design** 5

**3 Objectives of this post booster dose Omicron correlates study** 5

**4 Stage 2 sampling design for addressing the objectives** 6

**5 Statistical analysis plan by objectives** 7

#### List of Tables

1 Stratified sampling design for measuring Omicron antibody at (BD1, BD29) for non-cases and at (BD1, BD29, DD1) for cases* . . . . . . . . . . . . . . . . . . . . . . . . . 8

#### List of Figures

1 Flow-chart of the stage 2 correlates study that evaluates Omicron antibody as a correlates of risk and of protection of Omicron COVID-19. . . . . . . . . . . . . . . . . 7

### 1 Outline

First, this document recapitulates the sampling design that was used for assessment of Stage 1 correlates (Gilbert et al., 2022b). Second, it states the study objectives to assess post dose 3 Omicron BA.1 antibody titer, and exposure-proximal antibody titer, as immune correlates for Omicron COVID-19. Third, it describes the sampling plan for enabling the immune correlates statistical analyses. Fourth, it specifies the statistical analysis plan that details how to assess each objective.

Three assays VAC62, VAC122, VAC123 have been selected for this study:

VAC62– PsVNA against ancestral D614G strain (PPD Vaccines)

VAC122– PsVNA against BA.1 (B.1.529) VAC122 (PPD Vaccines)

VAC123– MSD multiplex: S, RBD, N +S (D614, Gamma, Alpha, Beta, Delta AY4, Omicron BA.1) (PPD Vaccines)

### 2 Stage 1 correlates sampling design

A two-phase stratified case-cohort sampling design was applied for measuring D1, 29, 57 antibody levels after the two-dose primary series in per-protocol participants sampled into the immunogenicity subcohort and for all baseline negative per-protocol vaccine recipient COVID-19 endpoint cases occurring at least 7 days post D29 visit or at least 7 days post D57 visit. The implemented sampling design is described in the Supplementary Material of Gilbert et al. (2022b). The sampling design sought balanced numbers of baseline negative per-protocol participants in each of the six demographic strata defined by (Minority, Non-Minority) $\times$ (Age $\geq$ 65, Age $<$ 65 and 'at risk', Age $<$ 65 and Not 'at risk'), within each of the naïve and non-naïve populations. For sampling of non-cases for Stage 2 correlates, balance in these factors will also be pursued.

### 3 Objectives of this post booster dose Omicron correlates study

The following objectives are assessed separately in SAR-CoV-2 naïve and SAR-CoV-2 non-naïve individuals, as defined below. The study endpoint for all objectives is adjudicated "Omicron COVID-19" counted starting 7 days after the post-booster Day 29 (BD29) visit and starting December 1, 2021 or later. For Objectives 7 and 9, "instantaneous Omicron COVID-19" refers to the instantaneous hazard rate of Omicron COVID-19, i.e., the rate of Omicron COVID-19 over the next day of follow-up. The objectives are assessed primarily for two BA.1 markers: bAb to Spike BA.1 in the MSD multiplex (VAC123), and pseudovirus nAb-ID50 titer to BA.1 (VAC122), based on assays at PPD. In addition, some of the objectives will be repeated for the same markers measured against D614 (binding assay) or D614G (pseudovirus neutralization) instead of against BA.1.

**Objectives**

1. To assess BD29 Omicron Ab as a correlate of risk (CoR) against Omicron COVID-19
2. To assess fold-rise in Omicron Ab from BD1/pre-booster to BD29 as a CoR against Omicron COVID-19

3. To assess whether the CoR in 1. or 2. is modified by SARS-CoV-2 naïve/non-naïve status
4. To assess whether the CoR in 1. is modified by the BD1 antibody value
5. To assess BD29 Omicron Ab as a correlate of protection (CoP) against Omicron COVID-19
6. To assess fold-rise in Omicron Ab from BD1 to BD29 as a CoP against Omicron COVID-19
7. To assess Omicron Ab as an exposure-proximal CoR of instantaneous Omicron COVID-19
8. To assess whether the exposure-proximal CoR in 7. is modified by the BD1 antibody value
9. To assess Omicron Ab as an exposure-proximal CoP against instantaneous Omicron COVID-19
10. To assess mediation of the effect of the interval between dose 2 and dose 3 on Omicron COVID-19 through BD29 Omicron Ab value

#### 4 Stage 2 sampling design for addressing the objectives

Figure 1 shows the blood sampling schedule that enables the correlates studies. This correlates study is a stratified case-control study of post-dose 3 Omicron Ab in 3-dose vaccine recipients. The sampling approach samples Omicron COVID-19 endpoint cases starting 7 days after BD29 from each of the original vaccine and cross-over vaccine arms. Sampling stratified by randomization arm creates useful variability in the time between the two-dose vaccination series and the booster dose. The primary study endpoint is Omicron COVID-19 occurring at least 7 days post dose 3 Ab measurement at BD29 through to the data base lock in May, 2022. The sampling is done separately in the "naïve" cohort with no evidence of SARS-CoV-2 infection from enrollment through to BD1 and in the "non-naïve" cohort with any evidence of SARS-CoV-2 infection from ≥14 days after the second dose of mRNA-1273 vaccine through to BD1. Here, prior infection is defined inclusively based on any results of previous RT-PCR+, N-seroconversion, or a symptomatic COVID-19 endpoint with positive confirmatory testing. Stratifying the sampling by naïve/non-naïve status enables study of immune correlates in each of the naïve and non-naïve cohorts given the importance of understanding immune correlates in non-naïve populations as well as in naïve populations, and addressing whether and how prior infection modifies immune correlates (Objective 3). Figure 1 shows the schema of blood sample storage for potential antibody measurement in the COVE study.

The sample size of the correlates study in terms of participants with new antibody measurements is as follows:

1. Stratified random sample of N=256 three-dose vaccine recipients
2. 640 total samples/assays (Omicron BA.1 Ab measured at BD1, BD29 for all participants, and also at disease-day-one (DD1) that is the date of COVID-19 endpoint diagnosis for all cases)

Each of the correlates studies (in naïve and non-naïve individuals) is based on 64 vaccine cases with antibody data; in comparison ≈peak Ab correlates were defined based on 36 vaccine cases in the Stage 1 correlates analyses (Gilbert et al., 2022b) and exposure-proximal correlates were defined

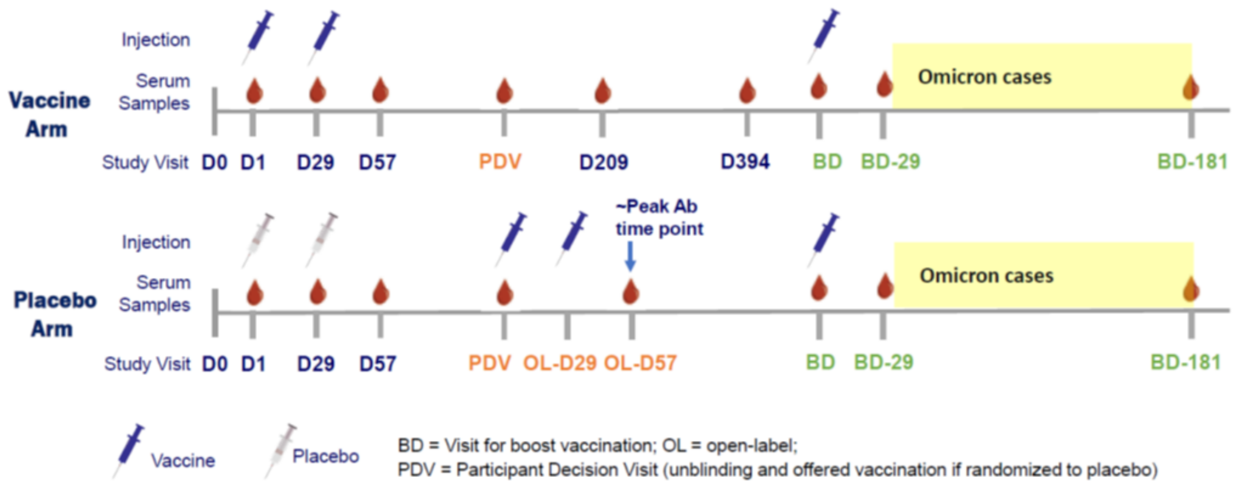

Figure 1: Flow-chart of the stage 2 correlates study that evaluates Omicron antibody as a correlates of risk and of protection of Omicron COVID-19.

based on 39 vaccine cases for an observational study of Pfizer's mRNA vaccine (Bergwerk et al., 2021).

Table 1 presents the sampling strata, where for eligibility participants must qualify per-protocol during the original follow-up period, received the first booster dose, have blood samples at BD1 and BD29, and cases are also required to have DD1 sample availability. Appendix A provides complete details of the Stage 2 sampling design that includes a prioritization of sampling of eligible participants and a dependency of sampling on demographic factors, which allows computation of inverse probability of sampling weights for all participants included in the correlates study.

For peak time correlates analyses of BD29 markers, in addition to requiring cases to have failure time starting 7 days after BD29 for inclusion, it is also required that the time interval between BD1 and BD29 falls in 19 to 45 days; otherwise the case is excluded from analysis.

### 5 Statistical analysis plan by objectives

#### 5.1 Descriptive statistics

Tables of immunogenicity will be reported separately by assay, which amounts to the following variables:

1. $\log_{10}$ nAb titer to D614G
2. $\log_{10}$ nAb titer to BA.1
3. $\log_{10}$ anti-Spike IgG to D614

Table 1: Stratified sampling design for measuring Omicron antibody at (BD1, BD29) for non-cases and at (BD1, BD29, DD1) for cases*

| | Boosted Sep23-Oct15 2021 | | Boosted Oct16-Oct31 2021 | | Boosted Nov 2021 | | Boosted Dec 2021 | | Total (Samples) |
|---|---|---|---|---|---|---|---|---|---|
| Original Vx Omicron case | 8N | 8NN | 8N | 8NN | 8N | 8NN | 8N | 8NN | 64 (192) |
| Original Vx non-case | 8N | 8NN | 8N | 8NN | 8N | 8NN | 8N | 8NN | 64 (192) |
| Crossover Vx Omicron case | 8N | 8NN | 8N | 8NN | 8N | 8NN | 8N | 8NN | 64 (192) |
| Crossover Vx non-case | 8N | 8NN | 8N | 8NN | 8N | 8NN | 8N | 8NN | 64 (192) |

*Case = COVID-19 endpoint in the interval [$\geq$ 7 days post BD29 AND $\geq$ Dec 1 2021, May 2022 data base lock date]. As described in the appendix the COVID-19 endpoint is documented to be Omicron BA.1 if possible whereas for some non-naïve COVID-19 endpoints there was not lineage data available to document the case to be Omicron BA.1.
Non-case = Did not acquire COVID-19 (of any strain) in the interval [BD1, data base lock date].
naïve = No evidence of SARS-CoV-2 infection from enrollment through to BD1;
Non-naïve = Any evidence of SARS-CoV-2 infection in the interval [$\geq$ 14 days after the second dose of mRNA-1273, BD1], operationalized as a COVID-19 endpoint or seroconversion from a blood sample up to the BD1 visit (evidence of infection by RNA PCR from a BD1 sample was not included as a qualifier for the Non-naïve group).

4. $\log_{10}$ anti-Spike IgG to Gamma
5. $\log_{10}$ anti-Spike to Alpha
6. $\log_{10}$ anti-Spike to Beta
7. $\log_{10}$ anti-Spike to Delta AY4
8. $\log_{10}$ anti-Spike to BA.1
9. $\log_{10}$ anti-RBD IgG to D614

Note that while descriptives are provided for all of the assay variables, the correlates analyses focus on four assay variables: $\log_{10}$ nAb ID50 titer to D614G, $\log_{10}$ nAb ID50 titer to BA.1, anti-Spike IgG to D614, and anti-Spike IgG to BA.1.

Inverse-probability weighting will be used in summarizing immunogenicity in order that estimates and inferences are for the population from which the whole study cohort was drawn. This whole population and the sample weights are defined in Section 6.1.

**Assay readouts accounting for assay limits (before multiplying the readouts by constants)**

The antibody markers have readouts in units defined by PPD reports, and the readouts account for the LOD, LLOQ, ULOQ assay limits derived by PPD for each assay.

The readout for the analysis of the two nAb ID50 titer markers is serum antibody concentration Ab[C], with labeling for plots "ID50 (AU/ml)." For D614G, the LLOQ for Ab[C] is 10 AU/ml. For D614G the ULOQ for Ab[C] is 281,600 AU/ml. Values $>$ ULOQ are assigned ULOQ. These LLOQs and ULOQs were derived for the D614G antigen in the PPD assay report "PPD Project

ID: RPJX. Assessment of Equivalency of Neutralization Antibodies Between the PPD VSDVAC 62 Microneutralization Assay and Historical ID50 Results Provided by Moderna That Were Generated Using the Duke Microneutralization Assay and the D614G Microneutralization Assay Version 1.0." PPD amended the ULOQ of 281,600 AU/ml based on FDA feedback that precision, relative accuracy and dilutional linearity of the SARS CoV-2 MN assay as well as ULOQ for the assay be based on the highest measurable sample that shows acceptable precision and accuracy.

For the Omicron BA.1 antigen, the PPD assay report "PPD Project Code: RVUJ2. Validation of A Microneutralization Assay for the Detection of SARS CoV-2 Neutralizing Antibodies (SARS CoV-2-NAb) for the Omicron Spike BA.1 Variant in Human Serum (SARS CoV-2 MN O) Version 1.0" yielded the LLOQ for Ab[C] of 8 AU/ml and a ULOQ of 24,503 AU/ml. Values > ULOQ are assigned ULOQ.

The LLOQ for PsV nAb ID50 is 10 for D614G and 8 for BA.1, respectively. PsV nAb ID50 D614G values below LLOQ = 10 are assigned the value of LLOQ/2 = 5 and PsV nAb ID50 BA.1 values below LLOQ = 8 are assigned the value of LLOQ/2 = 4.

**Multiplying nAb ID50 titer assay readouts by constants to place readouts on a comparable scale to units used in the previous immune correlates publications for Moderna COVE**

The PPD assay report RPJX cited above showed that Ab[C] is on the same AU/ml scale as the Duke ID50 titer readout with no need for multiplying Ab[C] by a constant, that is, analyses would be acceptable if they treat Ab[C] to have the same unitage as the Duke ID50 biomarker. That report estimated a scaling factor of 1.04 between the PPD Ab[C] readout and the Duke ID50 readout, and therefore we do apply this scaling factor, even though it has little impact.

In addition, we multiply PPD Ab[C] readouts by 0.242, which was the conversion factor used by Duke to convert their ID50 readouts to the IU50/ml scale. It might seem better to divide the PPD Ab[C] readouts by 1.275, as this was the conversion factor estimated by PPD in its calibration report "PPD Project Code: RQHQ. Calibration of the V62RS-X132-CVMN Reference Standard Used in the SARS CoV-2 Neutralizing Antibodies (SARS CoV-2-Nab) in Human Serum (SARS CoV-2 MN) Method to the WHO International Standard for anti-SARS-CoV-2 Immunoglobulin Lot 20/136 Version 1.0." However, to meet our greater objective to be able to compare readouts to those previously used in the blinded-phase Moderna COVE correlates studies (Gilbert et al., 2022b; Benkeser et al., 2023), we apply the Duke conversion factor. This means we can interpret anti-D614G ID50 titer readouts at BD1 and BD29 in the current correlates study on an apples vs. apples scale compared to the readouts used in the previous correlates studies. A section in the Supplemental Material of the booster correlates manuscript will explain the reasoning of this choice in greater detail. In sum, the original PPD Ab[C] ID50 readouts received from PPD are multiplied by 0.242 and then they are divided by 1.04, to constitute the reported ID50 (AU/ml) readouts. Then, the PPD anti-BA.1 Ab[C] ID50 readouts received from PPD are also multiplied by 0.242 and then they are divided by 1.04, for placing the readouts on a comparable scale to readouts against the D614G strain. Statistical reports are generated both using the un-scaled PPD assay units for each of D614G ID50 and BA.1 ID50, as well as using the scaled units for each of D614G ID50 and BA.1 ID50 (multiplying un-scaled readouts by 0.242/1.04).

In addition, it is of interest to consider D614G ID50 readouts scaled to be predicted ID50 values against BA.1; the advantage of doing this is the anchoring to the Duke/PPD D614G assay concordance study, as the Duke/PPD BA.1 concordance study is still ongoing. Based on data from the Duke assay on 26 3-dose mRNA-1273 participants, the geometric mean ratio of ID50 readouts to BA.1 vs. to D614G was 0.225. Therefore, it is of interest to scale D614G readouts by multiplying them by 0.225, which gives the readouts interpretations in terms of predicted ID50 against BA.1. Multiplying original PPD D614G ID50 units by (0.242/1.04)*0.225 = 0.052 creates the Predicted BA.1 units that can be quantitatively interpreted in comparison to the anti-D614G IU50/ml units, where for example a result of Predicted BA.1 ID50 is 2-fold lower than D614G IU50/ml can be properly interpreted as 2-fold lower titer against BA.1 than against D614G. The following data analysis will be included:

The blinded phase correlates study (Gilbert et al. 2022) estimated how two-dose vs. placebo vaccine efficacy varied by D614G nAb ID50 titer at 4 weeks post dose 2, with ID50 titer calibrated to the WHO 20/136 International Standard and reported in IU50/ml units. It is of interest to compare this ancestral antibody, ancestral COVID-19 vaccine efficacy curve with the BA.1 antibody, Omicron BA.1 COVID-19 booster vaccine efficacy curve, to ascertain whether a different amount of variant-matched antibody is needed for high-level booster protection than for high-level two dose vs. placebo protection. To do this, we defined a Predicted BA.1 ID50 biomarker at BD29 scaled such that it can be absolutely quantitatively interpreted vs. D614G IU50/ml units. This scaling was accomplished in two steps. First, the Duke/PPD D614G assay concordance study (BARDA, 2021) and the Duke assay International Standard calibration study (Huang et al., 2021) showed that multiplying D614G PPD nAb ID50 readouts by (0.242/1.04) transforms units to the IU50/ml scale previously used (Gilbert et al., 2022c). Second, based on data from 26 three-dose mRNA-1273 participants with Duke assay ID50 measured 4 weeks post dose 3 against both D614G and BA.1, the geometric mean ratio of ID50 against BA.1 vs. against D614G was 0.225 (Lyke et al., 2022; Atmar et al., 2022). Therefore, we multiplied the PPD D614G nAb IU50/ml values by 0.225, attaining the Predicted BA.1 ID50 values (thus original PPD BA.1 ID50 units are multiplied by (0.242/1.04)*0.225 = 0.052 to generate Predicted BA.1 ID50 units). The BD29 booster vaccine efficacy curve analysis was repeated for this biomarker, and results overlaid with the original Day 57 vaccine efficacy curve analysis, providing a means for absolute comparison of variant-matched titer levels associated with efficacy.

Note that for plotting labeling, IU50/ml and BAU/ml labeling is never used, because antibody responses to BA.1 are of primary interest, and international units do not exist for these readouts. For the responses against D614G or D614 the readouts indeed are in international units IU50/ml and BAU/ml; however, for consistency with BA.1 readouts plotting labels AU/ml are used, and footnotes of captions note that these units equate to international units.

The assay limits for the PPD VAC123 MSD multiplex assay are listed below. In particular, the LLOQs are taken as the LLOQs for the lowest dilution 1:500, and are as follows by antigen:

- Spike D614: 69
- B.1.1.529/BA.1: 102
- B.1.617.2/Delta: 150

- P.1/Gamma: 143
- B.1.1.7/Alpha: 52
- B.1.351/Beta: 111
- RBD D614: 79

In addition, the ULOQs are taken as the ULOQs for the highest dilution 1:500,000, and are as follows by antigen:

- Spike D614: 14,400,000
- B.1.1.529/BA.1: 1,180,000
- B.1.617.2/Delta: 8,000,000
- P.1/Gamma: 5,800,000
- B.1.1.7/Alpha: 8,800,000
- B.1.351/Beta: 5,000,000
- RBD D614: 5,800,000

The LLOQ for bAb Spike is 69 AU/ml for D614 and 102 AU/ml for BA.1. bAb spike D614 readouts below LLOQ = 69 AU/ml are assigned the value LLOQ/2 = 34.5 AU/ml, and bAb spike BA.1 readouts below LLOQ = 102 AU/ml are assigned the value LLOQ/2 = 51 AU/ml.

Reporting units for MSD binding antibody readouts in tables and figures are AU/ml. PPD did not develop a conversion factor from AU/mL to International Units (BAU/ml) for any of the MSD assays. There is no equivalency study of the PPD VAC123 MSD assay compared to the VRC MSD assay that was used in the first correlates study Gilbert et al. (2022b).

**Definition of participants with a positive response**

- Participants with a positive (quantifiable) pseudovirus neutralization response at each pre-defined timepoint are defined as participants who had ID50 value at the time point greater than or equal to the antigen-specific LLOQ; otherwise the response is not detectable. This definition is the same for both nAb D614G and nAb BA.1.
- Participants with a positive antigen-specific binding antibody response at each pre-defined timepoint are defined as participants who had a antigen-specific bAb measurement at the time point greater than or equal to the antigen-specific LLOQ (specified above); otherwise the response is negative.

**Tabular output**

- Average duration of follow-up post BD29 for cases and non-cases, stratified by naïve/non-naïve status
- Number (%) positive responses (including denominator that is the estimated number of participants in the population in the cell) with 95% CI at each time point (columns) by original randomization arm x case-control status x naïve/non-naïve status (rows). 95% CI calculated

based on Clopper-Pearson method. Table pools participants over the four boosting intervals listed in Table 1. The time points are BD1, BD29, and disease-day 1 (DD1) (only cases are included for DD1).

- Number (%) positive responses with 95% CI at BD1 by boosting interval (columns) and original randomization arm x case-control status x naïve/non-naïve status (rows). 95% CI calculated based on Clopper-Pearson method.
- Number (%) positive responses with 95% CI at BD29 by boosting interval (columns) and original randomization arm x case-control status x naïve/non-naïve status (rows). 95% CI calculated based on Clopper-Pearson method.
- Number (%) positive responses with 95% CI at DD1 by boosting interval (columns) and original randomization arm x case-control status x naïve/non-naïve status (rows). 95% CI calculated based on Clopper-Pearson method.
- Geometric mean (95% CI) of quantitative marker at each time point (columns) by original randomization arm x case-control status x naïve/non-naïve status (rows). 95% CIs using the t-distribution approximation of $\log_{10}$-transformed marker (base 10 of the logarithm is always used). Table pools participants over boosting interval. The time points are BD1, BD29, and DD1 (only cases are included for DD1).
- Geometric mean (95% CI) of quantitative marker at BD1 by boosting interval (columns) and original randomization arm x case-control status x naïve/non-naïve status (rows). 95% CIs using the t-distribution approximation of log-transformed marker
- Geometric mean (95% CI) of quantitative marker at BD29 by boosting interval (columns) and original randomization arm x case-control status x naïve/non-naïve status (rows). 95% CIs using the t-distribution approximation of log-transformed marker
- Geometric mean (95% CI) of quantitative marker at DD1 by boosting interval (columns) and original randomization arm x case-control status x naïve/non-naïve status (rows). 95% CIs using the t-distribution approximation of log-transformed marker.
- Geometric mean ratio (95% CI) of quantitative marker at BD29 and DD1 time points relative to BD1 time point (columns) by original randomization arm x case-control status x naïve/non-naïve status (rows). (i.e., geometric mean of fold-rise values from BD1 to BD29 and from BD1 to DD1.) 95% CIs using the t-distribution approximation of log-transformed marker at each time point. Table pools participants over boosting interval.
- Geometric mean ratio (95% CI) of quantitative marker at BD29 relative to BD1 time point by boosting interval (columns) and original randomization arm x case-control status x naïve/non-naïve status (rows). 95% CIs using the t-distribution approximation of log-transformed marker.
- Geometric mean ratio (95% CI) of quantitative marker at DD1 relative to BD1 time point by boosting interval (columns) and original randomization arm x case-control status x naïve/non-naïve status (rows). 95% CIs using the t-distribution approximation of log-transformed marker.

- Differences in positive response rates (95% CI) between cases and controls at each time point (columns) by original randomization arm x naïve/non-naïve status (rows). 95% CI the Wilson-Score method without continuity correction (Newcombe, 1998). Table pools participants over boosting interval. The time points are BD1 and BD29.
- Differences in positive response rates (95% CI) between cases and controls at BD1 by boosting interval (columns) and original randomization arm x naïve/non-naïve status (rows). 95% CI the Wilson-Score method without continuity correction (Newcombe, 1998).
- Differences in positive response rates (95% CI) between cases and controls at BD29 by boosting interval (columns) and original randomization arm x naïve/non-naïve status (rows). 95% CI the Wilson-Score method without continuity correction (Newcombe, 1998).
- Geometric mean ratio (95% CI) of quantitative marker between cases and controls at each time point (column) by original randomization arm x naïve/non-naïve status (rows). Table pools participants over boosting interval. The time points are BD1 and BD29.
- Geometric mean ratio (95% CI) of quantitative marker between cases and controls at BD1 by boosting interval (columns) and original randomization arm x naïve/non-naïve status (rows).
- Geometric mean ratio (95% CI) of quantitative marker between cases and controls at BD29 by boosting interval (columns) and original randomization arm x naïve/non-naïve status (rows).

**Graphical Output**

*Set 1 plots: BD1 and BD29 Ab distributions by case/non-case and naive-non-naive status*

1. BD1 antibody for the 2 $\log_{10}$ nAb ID50 titer markers (to D614G and to BA.1), 8 panels of violin/boxplots defined by 4 rows (cross-classification of original randomization arm with naïve/non-naïve) and 2 columns defined by D614G and BA.1 strain, where within each panel there are side-by-side violin/boxplots for cases and non-cases. These plots pool over the four boosting intervals.
2. Repeat 1. for BD29 antibody
3. Repeat 1. for BD29 - BD1 antibody
4. Repeat 1.–3. for the 2 $\log_{10}$ IgG anti-Spike markers (to D614 and to BA.1)
5. Repeat 1.–3. for the 2 $\log_{10}$ IgG anti-RBD markers (to D614)
6. BD1 antibody for the 6 $\log_{10}$ IgG anti-Spike markers (to D614, Gamma, Alpha, Beta, Delta AY4, BA.1), 24 panels of violin/boxplots defined by 4 rows (cross-classification of original randomization arm with naïve/non-naïve) and 6 columns defined by strain, where within each panel there are side-by-side violin/boxplots for cases and non-cases. These plots pool over boosting intervals.
7. Repeat 7. for BD29 antibody
8. Repeat 7. for BD29 - BD1 antibody

*Set 2 plots: Longitudinal plots BD1 to BD29 (and to DD1)*

1. For $\log_{10}$ nAb ID50 titer to D614G, for each of 4 rows (cross-classification of original randomization arm with naïve/non-naïve), plot 5 side-by-side violin/box plots, the the first 2 for BD1 non-cases, BD29 non-cases, with lines connecting individual's data points, and the last 3 for BD1 cases, BD29 cases, DD1 cases, with lines connecting individual's data points. To the right of this plot, place the parallel results for $\log_{10}$ nAb ID50 titer to BA.1. These plots pool over the four boosting intervals.
2. Repeat 1. for $\log_{10}$ anti-Spike IgG (for D614 and BA.1)
3. Repeat 1. for $\log_{10}$ anti-RBD IgG (for D614)

*Set 3 plots: Correlation plots across markers at a given time point*

1. For all 15 markers at BD1, a pairs plot similar to those in Gilbert et al. (2022b), pooling over boosting intervals, original randomized arm, case/non-case status, and naïve/non-naïve status. Spearman rank correlation coefficients are included (including IPS weights).
2. Repeat 1. for the 15 markers at BD29
3. Repeat 1. for the 15 difference markers BD29 - BD1 (i.e., $\log_{10}$ fold-rise markers)
4. Repeat 1.–3. restricting to the 6 markers of focus as defined in Section 5.1.

*Set 4 plots: Correlation plots for a given marker across time points*

1. For each of the 15 markers, a figure with 8 panels, with 4 rows (cross-classification of original randomization arm with naïve/non-naïve) of pairs plots, pooling over boosting intervals, for the marker measured over the time points BD1 and BD29 for non-cases (column 1) and over BD1, BD29, and DD1 for cases (column 2). Spearman rank correlation coefficients are included.

##### 5.2 Details on planned figures and tables for the first manuscript

**Proposed Figure 1 of the manuscript:** Include the nAb ID50 BA.1 marker and the IgG Spike BA.1 marker. 8 panels, 2 rows, 4 columns. Each panel shows the violin plots for BD1 and BD29 marker distributions, with lines connecting the BD1 and BD29 data points so the paired data/fold-rises are visible. The 2 rows are for (1) nAb marker and for (2) IgG Spike (the logic here is the y-axis range can always be the same). The 4 columns are for (1) Naive Omicron Cases; (2) Naive Non-Cases; (3) Non-naive Omicron Cases; (4) Non-naive Non-Cases Plotting symbols distinguish Original-Vaccine and Crossover-Vaccine.

Then a supp figure would do the same thing for nAb ID50 D614G and IgG Spike D614. And 2 other supp figures would do the same thing except replace BD29 marker with Fold-rise marker.

**Proposed Table 1 of the manuscript:** Like Table 1 in the 2022 Science paper, for the same 2 BA.1 markers of Figure 1 (nAb ID50, IgG Spike), focusing only on the BD29 time point, showing BD29 absolute level markers and fold-rise markers as separate rows, and separately for Naive and Non-Naive. So the rows would be (1) Naive, ID50 BA.1, BD29; (2) Naive, IgG Spike BA.1, BD29; (3) Naive, ID50 BA.1, Fold-rise; (4) Naive, IgG Spike BA.1, Fold-rise; (5) Non-Naive, ID50 BA.1,

BD29; (6) Non-Naive, IgG Spike BA.1, BD29; (7) Non-Naive, ID50 BA.1, Fold-rise; (8) Non-Naive, IgG Spike BA.1, Fold-rise.

Then a supp table that is the same except it is for nAb ID50 against D614G and IgG Spike against D614.

**Proposed Figure 2 of the manuscript:** Of the 'identical' structure/layout of Figure 2 in the 2022 Science paper, with Panel A for Naive, ID50 BA.1, BD29 and Panel B for Non-Naive, ID50 BA.1, BD29. Panel C would include results for 8 markers, the same 8 listed above for Table 1.

##### 5.3 Assessing Objectives 1–4 (≈peak Ab and pre-booster Correlates of Risk)

For the CoR Objectives 1.–4., the planned analysis is similar to the originally published Stage 1 CoR analysis, implementing baseline-covariate marginalized Cox regression and nonparametric monotone-constrained analysis in the stratified random sample of three-dose vaccine recipients, who were per-protocol during the original follow-up period, received the first booster dose, and have blood samples at BD1, BD29, and also at DD1 for cases. The Cox regression modeling is done using study time to be consistent with what was done originally for COVE; this approach could have reduced precision compared to using calendar time if calendar time predicts COVID-19. If calendar time does strongly predict COVID-19, the analyses may be repeated using the calendar time scale. Cox modeling for CoP objectives uses the calendar time scale (see Section 5.5).

For analyses of markers defined at BD29, the Cox model uses BD29 as the time origin, whereas for analyses of markers defined at BD1, the Cox model uses BD1 as the time origin. Specifically, output for the six analyzed markers listed in Section 5.1 is as follows, where the analyses are done separately for the naïve and non-naïve cohorts, as well as for pooling over the naïve and non-naïve cohorts.

1. (Obj. 1,2) Univariable Cox model results for each quantitative marker (hazard ratio, 95% CI, 2-sided p-value)
2. (Obj. 1,2) Univariable Cox model results for each tertilized marker (hazard ratios, 95% CIs, 2-sided p-values, Generalized Wald p-values)
3. (Obj. 1,2) Univariable Cox model marginalized marker-conditional mean cumulative incidence curves over time through to the last time point $t_0$, for Low, Medium, High tertile marker subgroups.
4. (Obj. 1,2) Univariable Cox model marginalized marker-conditional mean cumulative incidence curve over time through to the last time point $t_0$, with marker subgroups defined by the continuous value of the marker.
5. (Obj. 1,2) Univariable nonparametric monotonic-regression model (Kenny PhD dissertation) marginalized marker-conditional mean cumulative incidence curve over time through to the last time point $t_0$, with marker subgroups defined by the continuous value of the marker.
6. (Obj. 1,2) Multivariable Cox model for the two quantitative markers (anti-Spike IgG to BA.1, nAb ID50 titer to BA.1) (hazard ratios, 95% CIs, 2-sided p-values, generalized Wald test p-value)

7. (Obj. 1,2) Multivariable Cox model for the two tertilized markers (anti-Spike IgG to BA.1, nAb ID50 titer to BA.1) (hazard ratios, 95% CIs, 2-sided p-values, generalized Wald test p-values)

8. (Obj. 3,4) Multivariable Cox model for each of the two quantitative markers including an interaction term for naïve/non-naïve status (Obj. 3, 6) or for the BD1 antibody marker (Obj.4): A Wald p-value for interaction/effect modification is calculated

9. (Obj. 1, 2) Nonparametric threshold TMLE analysis the same as done in Gilbert et al. (2022b) with the method of Van der Laan and Gilbert (2022).

###### 5.3.1 Covariates adjusted for in CoR and CoP analyses

The following covariates are adjusted for in all CoR and CoP analyses: baseline behavioral risk score, heightened at-risk indicator, and indicator of White Non-Hispanic (same three variables as adjusted for in Gilbert et al. (2022b)). Analyses pooling over naive and non-naive include adjustment for naive status. Moreover, for the pooled analysis the multivariable superlearning CoR analyses also adjust for the interaction of heightened at-risk indicator with naive status.

In addition to the baseline covariates **X**, controlled risk CoP analysis that imagines "intervening" on a post-randomization event like BD29 antibody titer will also adjust for covariates measured after baseline but prior to BD29 and are associated with both the BD29 antibody titer and the endpoint; see Section 5.4 for details. Such covariates will include tertiles of BD1 antibody titer. For the analyses that pool over naïve and non-naïve, the analyses also adjust for naïve/non-naïve status. This is done because naïve/non-naïve status is strongly predictive of COVID-19, and likely will also be quite predictive of the BD29 antibody markers, such that it is likely a confounder of the effects of BD29 antibody markers on COVID-19.

The last time point $t_0$ for analysis is defined taking into account the smallest of the two latest COVID-19 endpoint failure times for naë and non-naë individuals, which is similar to as in Gilbert et al. (2022b) except only naïve individuals were studied previously. For the overall analysis of booster vaccine efficacy against Omicron, $t_0$ was selected as PENDING/TBD days, as the latest time point with reasonable precision in estimation.

###### 5.3.2 Machine learning analysis to estimate best models for predicting COVID-19

This analysis will only be pursued if the lower-dimensional CoR analyses of Objectives 1–4 generate substantial signal and motivate a machine-learning multivariable CoR analysis. The analysis will be conducted in the same way as done in Benkeser et al. for the multivariable Moderna correlates analysis of two-dose vaccine recipients (Benkeser et al., 2023), except 1) the markers involved and the baseline covariates involved are different and data from all participants are included, 2) to identify interactions between markers, SL.step.interaction will be added to the learner library, These variables are listed below, in the different sets for which a superlearner model is built. Cross-validated area under the ROC curve (CV-AUC) and variable importance analysis will also be conducted in the same way as done in (Benkeser et al., 2023).

1. Baseline demographics (= variables described in Section 5.3.1) and for analyses pooling over

naive and non-naive also include naive status and the interaction of heightened at-risk indicator with naive status.

2. Possible antibody marker variable sets accounting for assay type (bAb, nAb) where bAb refers to anti-Spike (D614, BA.1), anti-RBD (D614 only), and time point (BD1, BD29, BD29-fold-rise, which includes 2FR and 4FR variables – indicators of two-fold and four-fold rise)
    - BD1 bAb all BA.1
    - BD29 bAb all BA.1
    - BD29-fold-rise bAb all BA.1
    - BD1 nAb all BA.1
    - BD29 nAb all BA.1
    - BD29-fold-rise nAb all BA.1
    - BD1 bAb, BD29 bAb all BA.1
    - BD1 bAb, BD29-fold-rise bAb all BA.1
    - BD29 bAb, BD29-fold-rise bAb all BA.1
    - BD1 nAb, BD29 nAb all BA.1
    - BD1 nAb, BD29-fold-rise nAb all BA.1
    - BD29 nAb, BD29-fold-rise nAb all BA.1
    - BD1 (bAb, nAb) all BA.1
    - BD29 (bAb, nAb) all BA.1
    - BD29-fold-rise (bAb, nAb) all BA.1
    - BD1 (bAb, nAb,) BD29 (bAb, nAb) all BA.1
    - BD1 (bAb, nAb), BD29-fold-rise (bAb, nAb) all BA.1
    - BD29 (bAb, nAb), BD29-fold-rise (bAb, nAb) all BA.1
    - Repeat the above 18 sets for all D614 / D614G
    - Repeat the above 18 sets for all BA.1 and D614 / D614G

As for other analyses, the analysis is done separately for naïves, non-naïves, and naïves + non-naïves pooled.

#### 5.4 Assessing Objectives 5 and 6 (≈peak Ab as controlled risk Correlates of Protection)

##### 5.4.1 Primary controlled risk CoP analysis

Each of the BD29 and BD29 fold-rise markers is assessed as a controlled risk CoP as defined in Gilbert et al. (2022a), which is based on boosted participants only without a contrast of controlled risk in not-yet-boosted participants. This is analogous to CoR analysis of the vaccine arm only in the original blinded vaccine vs. placebo stage of the trial. This analysis reports E-values for each marker analyzed in tertiles and reports ignorance intervals and 95% estimated uncertainty intervals around the controlled risk curve estimate as a function of continuous immune marker value via the Cox modeling approach, the same as was done in the first Moderna CoP analysis (Gilbert et al., 2022b). As described in Gilbert et al. (2022a, Section 2.1), the objective of a controlled risk CoP analysis is to estimate the controlled risk parameter that assesses the causal effect of the antibody marker on COVID-19 risk. A controlled risk CoP analysis is different from a controlled vaccine efficacy CoP analysis (see Section 5.5), whose goal is to contrast participants in the vaccination arm and that in the not-yet-boosted arm.

Our primary interest is to assess the BD29 biomarker as a controlled risk CoP for the population who received the 3rd dose of mRNA-1273 vaccine in the COVE cohort. We will pursue this goal in the naïve and non-naïve populations, separately.

To be more specific, we will study the following causal estimand. Let $T(Ab1)$ denote the time to Omicron BA.1 COVID-19 after receiving the booster under assignment of all participants to $BD29 = Ab1$. For a fixed time $t_0$ after receiving the booster shot, define

$$r_M(Ab1) := \mathbb{E}_{\mathcal{P}_{\mathbf{L}}}[P(T(BD29 = Ab1) \leq t_0)],$$

where $\mathbf{L}$ denotes a vector of pre-treatment covariates ('pre-treatment' with respect to BD29) and $\mathcal{P}_{\mathbf{L}}$ is the distribution of $\mathbf{L}$ in the "per-protocol" naïve or non-naïve populations who received a booster.

Identification of $\mathbb{E}_{\mathcal{P}_{\mathbf{L}}}\{P(T(BD29 = Ab1) \leq t_0)\}$ from observed data depends on the ignorability assumption. One version of the ignorability assumption states that the BD29 titer level is independent of potential outcomes $T(Ab1)$ conditional on baseline covariates $\mathbf{X}$, including the minority indicator, high risk indicator and risk score, and covariates collected at BD1, including the matching biomarker level (or its tertiles) at BD1.

Under this ignorability assumption, the quantity $\mathbb{E}_{\mathcal{P}_{\mathbf{L}}}\{P(T(BD29 = Ab1) \leq t_0)\}$ is identified from observed data via the following g-computation formula (Gilbert et al., 2022a):

$$r_M(Ab1) := \mathbb{E}_{\mathcal{P}_{\mathbf{L}}}\{P(T(BD29 = Ab1) \leq t_0)\} = \mathbb{E}_{\mathcal{P}_{\mathbf{L}}}\{P(T \leq t_0 \mid BD29 = Ab1, \mathbf{L})\}, \quad (1)$$

where $\mathbf{L}$ is specified above. To faciliate interpretation, for a fixed BD1 Ab tertile, the controlled risk curve $r_M(Ab1)$ will be plotted against $Ab1$ and used to assess the BD29 biomarker of interest as a controlled risk CoP.

Analyses outlined above will be done with BD29 titer replaced by fold-increase from BD1 to BD29 (Objective 6).

##### 5.4.2 Exploratory controlled CoP analysis

We may pursue the following exploratory analyses. First, in addition to assessing the controlled risk CoP in the boosted population, we could also assess BD29 as a controlled risk CoP in the COVE trial population. Let $\mathcal{P}_{\mathbf{X}}$ denote the distribution of baseline covariates $\mathbf{X}$ in COVE. The parameter of interest would be $\mathbb{E}_{\mathcal{P}_{\mathbf{X}}}\{P(T(BD29 = Ab1) \leq t_0)\}$. Identification of $\mathbb{E}_{\mathcal{P}_{\mathbf{X}}}[P(T(BD29 = Ab1) \leq t_0 \mid \mathbf{X})]$ from observed data depends on the sequential ignorability assumption; see, e.g., Joffe and Greene (2009, Section 2.3) and Gilbert et al. (2022a, Supplementary Material B). One version of the sequential ignorability assumption states that the BD29 titer level is independent of potential outcomes $T(Ab1)$ conditional on baseline covariates $\mathbf{X}$, tertiles of BD1 marker level, and a person's naïve/non-naïve status as discussed in the primary controlled risk CoP analysis.

Under this version of sequential ignorability assumption, the quantity $\mathbb{E}_{\mathcal{P}_{\mathbf{X}}}\{P(T(BD29 = Ab1) \leq t_0 \mid \mathbf{X})\}$ is identified as follows (Gilbert et al., 2022a, Supplementary Material B):

$$\begin{aligned} & P(T(BD29 = Ab1) \leq t_0 \mid \mathbf{X}) \\ = & \sum_{a \in l,m,h; b \in 0,1} P(T \leq t_0 \mid BD29 = Ab1, BD1 = a, \text{Naïve} = b, \mathbf{X}) \times P(BD1 = a, \text{Naïve} = b \mid \mathbf{X}). \end{aligned} \tag{2}$$

where $a \in \{l, m, h\}$ denotes the the low, medium and high tertiles of the matching BD1 biomarker. In practice, the conditional probability $P(BD1 = a, \text{Naïve} = b \mid \mathbf{X})$ can be estimated via a multinomial regression. Finally, we standardize $P(T(BD29 = Ab1) \leq t_0 \mid \mathbf{X})$ to the COVE trial $\mathcal{P}_{\mathbf{X}}$ and obtain a controlled risk curve.

As a second exploratory analysis, we will study the controlled risk CoP in each randomization arm. Let $A = 1 =$ Vaccine if a participant was assigned to the vaccine arm and $A = 0 =$ Crossover if assigned to the placebo arm (and later crossed over to the vaccine arm) in the original COVE study; see Figure 1 for an illustration. Let $T(a, Ab1)$ denote the time to Omicron BA.1 COVID-19 after receiving the booster under assignment of all participants to $A = a$ and $BD29 = Ab1$. For a fixed time $t_0$ after receiving the booster shot, let $r_M(a, Ab1) := \mathbb{E}_{\mathcal{P}_{\mathbf{X}}}[P(T(A = a, BD29 = Ab1) \leq t_0 \mid \mathbf{X})]$, where a = Vaccine or Crossover, $\mathbf{X}$ denotes a vector of baseline covariates, and $\mathcal{P}_{\mathbf{X}}$ is the distribution of $\mathbf{X}$ in the COVE trial population.

Identification of $\mathbb{E}_{\mathcal{P}_{\mathbf{X}}}[P(T(A = a, BD29 = Ab1) \leq t_0)]$ from observed data depends is analogous to the two-stage g-computation discussed previously. Separate estimates of the curves $r_M(a, Ab1)$ in $Ab1$ for each $a = 0, 1$ will be produced. The contrast $r_M(\text{Vaccine}, Ab1)/r_M(\text{Crossover}, Ab1')$ will also be reported. This contrast characterizes the "joint effect" of being assigned to the vaccine versus crossover (which had an implication for the interval time and could potentially have an effect on the clinical outcome via a causal pathway not mediated by the BD29 antibody titer) and different levels of BD29 antibody titer.

In a third exploratory analysis, the potential outcome of interest is:

$$T(\Delta, Ab1) := T(\text{receiving booster } \Delta \text{ days after the 2nd vaccine}, BD29 = Ab1).$$

The identification of the controlled risk based on the potential outcome $T(\Delta, Ab1)$ will be based on a two-stage generalization g-computation discussed previously if the target population is the entire

COVE population and a single-stage g-computation if the population of interest is the boosted population. For selected values of $\Delta$, a controlled risk curve could be plotted as a function of BD29 titer level. In addition, controlled vaccine efficacy can be estimated and plotted for two distinct values of $\Delta$, e.g., the 10th and 90th percentiles.

Analyses outlined above will be done with BD29 titer replaced by fold-increase from BD1 to BD29 (Objective 6).

##### 5.5 Controlled VE CoP analysis of Objectives 5 and 6 based on boosted vs. not-yet boosted

In addition to the controlled risk CoP analysis, for assessing the BD29 antibody marker as a CoP against Omicron COVID-19, another approach measures the booster VE, defined as the hazard rate of COVID-19 for boosted vs. not-yet boosted individuals, or alternatively by the cumulative probability of COVID-19 by a given fixed time point for boosted vs. not-yet boosted individuals. We will study how the booster VE varies as a function of BD29 antibody level Ab1, through the stepped-wedge methodology designed by Fintzi and Follmann (2021).

To be more specific, at any time $t$, the risk set would consist of not-infected-by-Omicron participants who are at least 7 days post BD29 and not-yet boosted participants. Each boosted participant is associated with a BD29 antibody level and the hazard rate conditional on the BD29 Ab level, $\lambda_{\text{boost}}(t, Ab1)$, will be estimated. On the other hand, the hazard among the not-yet boosted participants, $\lambda_{\text{not-yet-boost}}(t)$, will also be estimated. The contrast $1 - \lambda_{\text{boost}}(t, Ab1)/\lambda_{\text{not-yet-boost}}(t)$ or boost efficacy by Ab1, will be reported and plotted as a function of of the BD29 antibody marker Ab1.

An estimate of the overall booster VE against Omicron COVID-19 provides a way to scale the controlled risk curve (marginalized Omicron COVID-19 risk vs. BD29 antibody level Ab1) to be a booster-controlled VE curve; see Gilbert et al. (2022d, Section 2.1) for the distinction between a controlled risk CoP analysis discussed in Section 5.3 and the controlled VE CoP analysis outlined in this section.

The cohort for the CoP analysis will be comprised of everyone who is unboosted as of 1 December 2021, plus the stratified case-control cohort (SCCC). Separate datasets and analyses will be constructed for the non-naive and naive cohorts. An illustrative version of the hazard function for peak antibody CoP analysis is given by

$$h(t) = h_0(t)\exp\{Z_i(t)[\beta_0 + \beta_1 Ab1] + X_i\theta\}w_i(t)I(t \in R_i)$$

where $R_i$ is defined to remove person $i$ from the risk set after event, censoring, or the 34 days postboost interval, $t$ is days since 1 December 2021, $Z_i(t)$ is 0 before boost and 1 after boost and $X_i$ is a vector of covariates. The weight $w_i(t)$ is a little complicated. There are three categories.

1. For those in the SCCC BD cohorts for period 1-3, $w_i(t)$ is the IPS weight.
2. For those in the SCCC BD period 4 cohort (boosted 1 December 2021 to 31 December 2021), $w_i(t) = 1$ prior to boosting and is the IPS weight after boosting.

3. For those not in the SCCC and unboosted/prior to boosting on 1 December 2021, $w_i(t)$ is 1 prior to boosting/COVID-19 event and 0 after boosting or COVID-19 event.

As an alternative, we will avoid weighting by imputing Ab1 in all vaccinees by empirical sampling with replacement from the distribution of BD29 antibody or either cases or controls, as appropriate. This should result in a much reliable estimate of $\beta_0$.

The same analysis will be conducted with the BD29 titer replaced by fold-increase from BD1 to BD29 (Objective 6).

**Repeating correlates analyses in the early period of follow-up in acknowledgment of waning vaccine efficacy**

Several studies have shown that mRNA booster vaccines have waning protection over time, including analysis of the COVE trial itself. Correlates of protection may be strongest and most interpretable during periods of substantial vaccine protection. Therefore, the ≈peak time point CoR and CoP analyses may be repeated using as the final time point $t_0 = 91$ days post Day 1 visit. This cut-point of 91 days is chosen in part to harmonize with the COVAIL immune correlates study that also assesses immune correlates restricting to COVID-19 endpoints occurring 91 days post booster. In addition, the immune correlates analyses may also be repeated restricting to COVID-19 endpoints occurring starting 92 days post Day 1 visit through to the final time point $t_0$ that was selected for the main correlates analyses.

##### 5.6 Assessing Objectives 7-9 (Exposure-Proximal Correlates of Risk and Correlates of Protection)

For assessing antibody as an exposure-proximal CoR (Objective 7), we use the below Cox model

$$h(t) = h_0(t)\exp\{Z_i(t)[\beta_0 + \beta_1 Ab_i(t-b)] + X_i\theta\}w_i(t)I(t \in R_i),$$

where $t$ is days since 1 December 2021 and $Ab_i(t-b)$ is the predicted antibody level for person $i$ at time $(t-b)$ post boost with other terms as defined in section 5.4. A CoR analysis will draw estimated curves with confidence bands of $exp\{\beta_1 Ab\}$ as a function of Ab ranging over the middle 95% of the distribution of predicted Ab. Assessing the Objective 8 will include a term for BD1 dichtomized at the median of the BD1 distribution and an interaction of dichtomized BD1 with $Ab_i$. Objective 9 will use the same model and provide CoP curves analogous to the CoR curves using $1 - exp(\beta_0 + \beta_1 Ab)$. Below we describe how we will impute $Ab(t)$.

In select cases the BD29 and DD1 antibody readouts will be used to calculate individual slopes using the form (BD29-DD1)/d where BD29 and DD1 are the antibody readouts and d the difference in days between BD29 and DD1. Denote the median slope as $\hat{\theta}$ which will be used to calculate individual antibody decay curves for *all* cases and non-cases using the formula

$$\text{Ab}(d) = \text{BD29} + \hat{\theta} \times d,$$

where $d$ is the number of day post BD29. This imputation will be performed for all individuals in the risk set at all event times. This approach has been applied to the Stage 1 blinded-phase COVE data, though the slope of decay there was estimated using data from Doria-Rose et al. (2021). Note that even though we have DD1 antibody value for the cases we don't use it in this approach in order to treat cases and controls the same way. It's bad if a covariate is measured one way for cases and another way for controls and the above symmetric imputation avoids this problem. Another reason not to impute DD1 is that the interval between BD29 and DD1 is random, which makes imputation problematic.

If the total variance is large relative to the within person variance, the above regression calibration approach may result in bias and to reduce such bias, an expected partial likelihood estimator may be considered.

The above analyses will be run separately for the naive and non-naive cohorts.

##### 5.7 Addressing Objective 10 on mediation of the effect of dose 2 to 3 interval on COVID-19 mediated through BD29 antibody

This question will be analyzed by a new method described in a manuscript under preparation (Hejazi et al., 2023). The exposure variable of interest $A$ must be dichotomous, so it will be defined as above vs. below the median number of days between dose 2 and dose 3. The putative mediator to study is BD29 $\log_{10}$ PsV nAb ID50 titer against BA.1, and the outcome is COVID-19, both variables defined the same as for the other ≈peak antibody correlates objectives. Covariates to adjust for $W$ will also be the same as used for the other ≈peak antibody correlates analyses. The analysis will be done with and without $V$ defined as the BD1 $\log_{10}$ PsV nAb ID50 titer against BA.1. The data set up fits the method of Hejazi et al. (2023) where $V$ is a likely confounder of the exposure-mediator relationship given that $V$ predicts both $A$ and the putative mediator.

The data analysis will be repeated for $\log_{10}$ PsV nAb ID50 titer against D614G as well as for each of the other markers $\log_{10}$ anti-Spike BA.1 IgG, $\log_{10}$ anti-Spike D614, IgG $\log_{10}$, and $\log_{10}$ anti-RBD D614 IgG.

This data analysis is based on a novel statistical method that is still being developed, which will be submitted as part of a statistical methods manuscript. Consequently, the results of this method will likely come later than results from the other analyses, and hence will likely be included in sequel manuscripts rather than in the first correlates manuscript resulting from this SAP.

#### 6 Specifications for general issues faced for most analyses

##### 6.1 Computation of inverse probability of sampling weights

Define six demographic categories, which were used in the stratified sampling design: Age $\geq 65$ minority; Age 18-64 'at risk' minority; Age 18-64 'not at risk' minority; Age $\geq 65$ non-minority; Age 18-64 'at risk' non-minority; Age 18-64 'not at risk' non-minority, i.e., to enrich/over-sample those Age $\geq 65$]. For each sampled participant, the inverse probability sampling weight is computed as numerator / denominator, where the numerator is the total number of per-protocol participants in the participant's cell (among the 32 of Table 1) that also have membership in the participant's demographic category (among the 6 listed above). The denominator is the total number of participants included in the numerator that were sampled for stage 2 correlates.

##### 6.2 Imputation of demographics variables for stratification and merging of sparse strata for weights computation

Wstratum depends upon the demo variables (age, at risk, minority), CalendarBD1Interval, naive, and trt. Controls with missing Wstratum won't be sampled, hence not part of ph1. On the other hand, cases with missing Wstratum are part of ph1 because they may be sampled. If cases have missing demo variables, we want to impute them so that we can assign weights, otherwise it gets too complicated to assign weights to cases. Imputation is performed over all cases and controls without missing demo variables. The latter are included to improve imputation performance. Imputation is performed for demo variables only, but can be enlarged if there are additional variables that provide info on the three demo variables. Due to the limited missingness, a single hard imputation is performed.

When there are strata with empty ph2 sample set, collapsing strata is performed in three steps. First, do it across demo strata within each of 32 sampling buckets. Second, if there are still empty strata, do it across the 4 calendar periods. Specifically, merge a period with the next period if not the last, and merge with the last period with the previous if needed. Third, do it across demo strata within each of 32 sampling buckets one more time because it is possible that collapsing across time periods in step 2 introduced empty demo strata. Assuming that DD1 may not be available for all cases with BD1 and BD29 markers, we will compute a different set of weights for DD1, which may be used for, e.g. computing positive response rates at DD1. We will first attempt to compute weights for DD1 using the Wstratum derived for computing BD29 weights. If it turns out that there are empty cells, we will re-collapse sampling strata to compute weights for DD1.

#### 7 Additional data analysis issues

##### 7.1 Exclude participants reporting being HIV positive from the correlates analysis

Because the lentivirus-based pseudovirus neutralization assay uses an HIV backbone, the presence of anti-retroviral drugs in serum can give a false positive neutralization signal. For this reason, the original immune correlates analysis Gilbert et al. (2022b) excluded participants who self-reported being HIV positive, because they would likely be taking anti-retroviral drugs. Consistent with the previous correlates analysis, this SAP also excludes participants who self-reported being HIV positive.

##### 7.2 Missing lineages

Some endpoint cases will likely have missing lineage/spike sequence. If the COVID-19 endpoint diagnosis date is $\geq$ January 15, 2022, then the lineage will be hard-imputed to be Omicron BA.1. If the COVID-19 diagnosis date is less than January 15, 2022, the lineage will be recorded as NA. Note that attempts were made to measure the lineage/sequence for 100% of selected cases, enabling addressing this issue in the data analysis. Data analyses will restrict to Omicron BA.1 cases, although if the number of non-naïve cases has more than 10% of missing lineages, then missing data methods may be used that account for missing lineage. A separate SAP describes an approach to doing this using hotdeck multiple imputation, similar to as in Sun et al. (2020), which may be added to this SAP if needed.

### Appendix A: Stage 2 Sampling: Stratified Case-Control Samples in 3-dose vaccine recipients

#### Appendix A.1 Stratified case-control sampling

Participants in P301 started receiving booster dose in Sep-2021 (first subject first booster dose 23-Sep-2021 in P301 Part C), and a total of 19,609 participates received a booster dose in Part C (Part C Safety Set, data cutoff date: 05-Apr-2022). Part C Safety Set will be used as the source dataset to sample for the stage 2 sampling. In this sampling plan, Omicron case, is approximated by adjudicated COVID-19 case (positive RT-PCR for SARS-CoV-2 with eligible symptoms) $\geq$ 7 days post BD29 AND $\geq$ 01-Dec-2021 given the emergence of Omicron (BA.1) wave. Primary endpoint COVID-19 cases with known Omicron BA.1 lineage are prioritized for sampling. The sampling of cases and controls are further stratified by the following:

1. Originally randomized to mRNA-1273 (mRNA-1273, original vaccine arm) vs. placebo recipients in the blinded phase (Part A) who received mRNA-1273 primary series in Part B (Placebo-mRNA-1273, cross-over vaccine arm), these two groups/arms create useful variability in the time between the two-dose vaccination series and the booster dose.

2. Calendar period a participant received a booster (23-Sep to 15-Oct-2021, 16-Oct to 31-Oct-2021, Nov-2021, Dec-2021).

3. naïve vs. non-naïve cohorts, where naïve participants are those with no evidence of SARS-CoV-2 infection through the day of receiving booster (BD-Day 1, or pre-booster, or BD1); and non-naïve participants are those with evidence of infection in [date of 2nd dose of the primary series + 14 days, BD1]. Infection is defined by either a positive RT-PCR for SARS-CoV-2, or conversion from non-positive to positive by Roche Elecsys assay (NP).

An equal number of cases vs. non-cases, 8 within each of the stratum defined by the above cross-classification will be sampled, as presented in Table 1 below. Primary endpoint COVID-19 cases with known Omicron BA.1 lineage are prioritized for sampling If fewer than 8 such eligible cases are available for sampling, then the remainder of the cell is filled with eligible cases with unknown lineage. For cases, antibodies at 3 timepoints: pre-booster Day 1 (BD1), 1 month/28 days after booster (BD29) and illness Day 1 (DD1) will be measured; for non-cases, antibodies at 2 timepoints: BD 1 and BD29 will be measured.

#### Appendix A.2 Specifications for Sampling

Participants who are in Per-protocol Primary Series analysis set and received booster dose (ADSL.PPPSFL = 'Y' AND ADSL.TR03SDT > .) are used for sampling. Eligible participants to be sampled also requires Case/Non-case, naïve/non-naïve, received booster during [23-Sep-2021, 31-Dec-2021] as defined in section 2.1.

For cases, severe Omicron COVID-19 cases (onset $\geq$ 01-Dec-2021) will be sampled first. Within each stratum, a random number generator function with seed of 1273 is used. Sampled participants who did not have sufficient serum samples available at the planned timepoints (BD1 and BD29 for non-cases, BD1, BD29, and DD1 for cases) will be replaced. Based on preliminary review of data, the number of non-naïve Omicron COVID-19 cases is very limited. Thus, for non-naïve Omicron cases, eligible primary endpoint COVID-19 cases will be sampled first. The remaining non-naïve cases will be sampled from infection cases (positive RT-PCR not necessarily with eligible symptom(s)), first from those with known Omicron BA.1 lineage, then from those likely to be Omicron BA.1. For these non-naïve Omicron COVID-19 cases and infections, every effort will be made to sample up to 8 participants, even if serum samples are not available at all 3 preferred timepoints (BD1, BD29, and DD1). If there are still not sufficient non-naïve cases, to fill out the 8 cases additional adjudicated COVID-19 cases 7 days post BD29 with onset $\geq$ 01-Dec-2021. In summary, in the situation available non-naïve cases are <8 in a cell, effort will be made: to sample a total of 16 cases for each arm per boosting calendar period as described above. Effort will be made to maintain 1:1 ratio between case: non-case for naïve and non-naïve cohort. In the situation when there are only x (<8) non-naïve cases to be sampled, 16-x naïve cases will be sampled to reach a total of 16 cases. Correspondingly, 16-x naïve non-cases and x non-naïve non-cases will be sampled be maintain 1:1 between cases: non-cases, as illustrated as an example in the table below:

| | | |
|---|---|---|
| Omicron case | 16-x | x |
| | N | NN |
| non-case | 16-x | x |
| | N | NN |

When feasible, for each set of 8N (naïve) or 8NN (non-naïve) in a cell, sample 2:1:1:2:1:1 from baseline demographic strata: Age $\geq$ 65 minority; Age 18-64 'at risk' minority; Age 18-64 'not at risk' minority; Age $\geq$ 65 non-minority; Age 18-64 'at risk' non-minority; Age 18-64 'not at risk' non-minority, i.e. to enrich/over-sample those Age $\geq$ 65].

#### Appendix B: Notes for construction of a mock data set

1. Need the mock data set to include all of the variables used for sampling as described in Table 1, which means adding a variable coding the four calendar boosting intervals, and adding a new variable to indicate naïve vs. Non-naïve. This is needed for computing sampling weights as well as for other purposes such as covariate adjustment.

2. The markers that need to be simulated are BD1, BD29 values (and DD1 values for cases) of:

    - $\log_{10}$ nAb titer to D614G
    - $\log_{10}$ nAb titer to BA.1
    - $\log_{10}$ anti-Spike IgG to D614
    - $\log_{10}$ anti-Spike IgG to Gamma
    - $\log_{10}$ anti-Spike to Alpha
    - $\log_{10}$ anti-Spike to Beta
    - $\log_{10}$ anti-Spike to Delta AY4

- $\log_{10}$ anti-Spike to BA.1
- $\log_{10}$ anti-RBD IgG to D614

For the naïve cohort, the BD1, BD29, and DD1 values could all be taken to be like D29, D57, and D29 values against D614/D614G of baseline negatives from the original COVE study, respectively, by assay type MSD/binding and pseudovirus neutralization, using BAU/ml and IU50/ml. For readouts to strains other than D614/D614G, will re-sample from the D614/D614G strain data. For the Non-naïve cohort, the BD1 and BD29 values could be taken from the D29 and D57 values against D614/D614G of baseline positives from the original COVE study, respectively, by assay type MSD/binding and pseudovirus neutralization. For DD1 values, could be taken from the D29 values against D614/D614G of baseline positives from the original COVE study. After readouts are calculated, for the MSD binding antibody data, values below the constant/non-strain-specific LLOQ (on the BAU/ml scale) are set to LLOQ/2, and for the nAb ID50 titer data, values below the constant/non-strain-specific LOD (on the IU50/ml scale) are set to LOD/2.

#### Appendix C: Miscellaneous

tfinal.tpeak is the minimum of tfinal.tpeak for each of four quadrants (2 trt * 2 naive status) and no larger than 105 (Dean et al's analyses). Within each qudrant, it is defined as smaller of the two: 1) time of the last case, 2) last time to have 15 ph2 samples at risk.

#### Appendix D: Definition of Stage-2 Per-Protocol Population

Booster dose correlates studies will be restricted to the "stage-2 per-protocol" (BDPerprotocol) population where the flag BDPerprotocol == TRUE if the participant satisfies the following two criteria:

1. The participant was in the original blinded-phase per-protocol cohort as in Gilbert et al. (2022b);
2. The participant received the booster dose (third mRNA-1273 dose) before and including December 31, 2021;

and belongs to one of the following 4 sampling strata:

1. "Case/Naïve"
2. "Case/Non-Naïve"
3. "Non-Case/Naïve"
4. "Non-Case/Non-Naïve"

where "naïve," "non-naı̈ve," "case," and "non-case" are defined as follows:

**Naïve** No evidence of SARS-CoV-2 infection detected by elecsys or RT-PCR from enrollment through BD1;

**Non-Naïve** Any evidence of SARS-CoV-2 infection in the interval [14 days after the second dose of mRNA-1273 vaccine, BD1];

**Case** 1) If a participant is naïve, then case is Omicron COVID-19 event in the interval [max(7 days post BD29, 01 Dec2021), 16 May 2022 database lock date]; 2) If a participant is non-naïve, then case is SARS-CoV-2 infection detected by Elecsys or RT-PCR in the interval [max(7 days post BD29, 01 Dec2021), 16 May 2022] and the Elecsys test was not positive at BD-D1 pre-booster;

**Non-Case** No evidence of SARS-CoV-2 infection detected by Elecsys or RT-PCR in the interval (BD1, 16 May 2022 database lock date].

For all analyses of the peak immune correlates (i.e., BD29 response), the "per-protocol" status further requires that

1. The participant did not miss a BD29 visit;
2. The participant had a BD29 measurement (approximate peak measurement) that was between 19 and 45 days, both inclusive, of the BD1 visit;
3. The participant was not censored or had any evidence of infection before 7 days post BD29 visit.
4. The participant did not live with HIV.
5. The participant did not acquire a non-adjudicated Omicron endpoint.
6. The participant did not test SARS-CoV-2 RT-PCR positive at the BD1 visit.

In the correlates studies of the "per-protocol" population, a study participant would be considered "naïve" by BD1 if there was no evidence of SARS-CoV-2 infection (RT-PCR+, Roche Elecsys seropositive, or a symptomatic COVID-19 endpoint followed by positive confirmatory testing) from enrollment to BD1 (including the BD1 visit). A study participant is considered "non-naïve" if the participant showed any evidence of COVID-19 starting 14 days post the second immunization in the primary series and through the BD1 visit.

Because a participant who tested PCR+ at BD1 was excluded from the per-protocol cohort, the person would not be associated with a naive/non-naive status.
