## Supplementary Materials for "Omicron COVID-19 Immune Correlates Analysis of a Third Dose of mRNA-1273 in the COVE Trial"

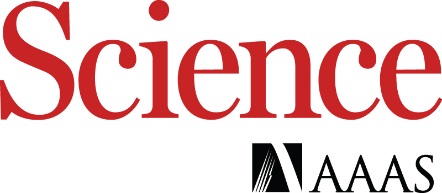

Supplementary Materials for

Omicron COVID-19 Immune Correlates Analysis of a Third Dose of mRNA-1273 in the COVE Trial

Bo Zhang, Youyi Fong, Jonathan Fintzi, Eric Chu, Holly E. Janes, Lindsay N. Carpp, Avi Kenny, Marco Carone, David Benkeser, Lars van der Laan, Weiping Deng, Honghong Zhou, Xiaowei Wang, Yiwen Lu, Chenchen Yu, Christopher R. Houchens, Karen Martins, Lakshmi Jayashankar, Chuong Huynh, Carl J. Fichtenbaum, Spyros Kalams, Cynthia L. Gay, Michele P. Andrasik, James G. Kublin, Lawrence Corey, Kathleen M. Neuzil, Frances Priddy, Rituparna Das, Bethany Girard, Hana M. El Sahly, Lindsey R. Baden, Ruben O. Donis, Richard A. Koup, Peter B. Gilbert, Dean Follmann*, on behalf of the United States Government (USG) COVID-19 Immune Assays Team; Moderna, Inc. Team; Coronavirus Vaccine Prevention Network (CoVPN)/Coronavirus Efficacy (COVE) Team; and USG/CoVPN Biostatistics Team

**This PDF file includes:**

Materials and Methods

Figs. S1 to S34

Tables S1 to S11

Supplementary Text

### United States Government (USG) COVID-19 Immune Assays Team

(PubMed listed, and ordered alphabetically by affiliation)

| Affiliation | Team Members |
| --- | --- |
| Biomedical Advanced Research and Development Authority (BARDA), Washington, DC | Oleg Borisov, Flora Castellino, Brett Chromy, Mark Delvecchio, Ruben O. Donis, Tremel Faison, Corey Hoffman, Christopher Houchens, Tom Hu, Chuong Huynh, Pennie Hylton, Lakshmi Jayashankar, Aparna Kolhekar, James Little, Karen Martins, Jeanne Novak, Carol Sabourin, Evan Sturtevant, Xiaomi Tong, John Treanor, Danielle Turley, Leah Watson |
| Boston Consulting Group, Boston, MA | Gian King, Andrew Li, Najaf Shah, Smruthi Suryaprakash, Jue Xiang Wang |
| Division of AIDS, NIAID, NIH, Bethesda, MD | Patricia D'Souza |
| Division of MID (Microbiology and Infectious Diseases), NIAID, NIH, Bethesda, MD | Janie Russell |
| Duke University, Durham, NC | David Beaumont, Kendall Bradley, Jiayu Chen, Xiaoju Daniell, Thomas Denny, Elizabeth Domin, Amanda Eaton, Kelsey Engel, Wenhong Feng, Juanfei Gao, Hongmei Gao, Kelli Greene, Sarah Hiles, Leihua Liu, Kristy Long, Kellen Lund, Charlene McDanal, David C. Montefiori, Marcella Sarzotti-Kelsoe, Francesca Suman, Haili Tang, Jin Tong, Olivia Widman |
| The Tauri Group, an LMI company - Contract Support for U.S. Department of Defense (DOD) Joint Program Executive Office for Chemical, Biological, Radiological and Nuclear Defense (JPEO-CBRND) Joint Project Manager for Chemical, Biological, Radiological, and Nuclear Medical (JPM CBRN Medical), Fort Detrick, MD, USA | Christopher S. Badorrek, Gregory E. Rutkowski |
| Vaccine Research Center, NIAID, NIH, Bethesda, MD | Akua Abrah, Obrimpong Amoa-Awua, Manjula Basappa, Robin Carroll, Erykah Coe, Jevone Fentress, Britta Flach, Suprabhath Gajjala, Nazaire Jean-Baptiste, Richard A. Koup, Bob C. Lin, Adrian McDermott, Christopher Moore, Mursal Naisan, Muhammed Naqvi, Sandeep Narpala, Sarah O’Connell, Abhinaya Srikanth, Clare Whittaker, Weiwei Wu |

### Moderna, Inc. Team

(PubMed listed, ordered alphabetically)

| Affiliation | Team Members |
| --- | --- |
| Moderna, Inc., Cambridge, MA | Weiping Deng, Shu Hahn, Jacqueline Miller, Rolando Pajon, Honghong Zhou |

### Coronavirus Vaccine Prevention Network (CoVPN)/Coronavirus Efficacy (COVE) Team

(PubMed listed, and ordered alphabetically by institution affiliation)

| **Affiliation/Funding*** | **Study Group** | **Location** |
| --- | --- | --- |
| AB Clinical Trials | Atoya Adams, MD, MBA, Eric Miller | Las Vegas, NV |
| Accel Research Sites | Bruce G. Rankin DO, John Hill MD, Steven Shinn MD, Marshall Nash MD | DeLand, FL |
| Advanced Clinical Research | Sinikka L. Green MD, Colleen Jacobsen, Jayasree Krishnankutty, Sikhongi Phungwayo | Cedar Park, TX |
| Alliance for Multispecialty Research | Richard M. Glover, II MD, Drs. Stacy Slechta, Troy Holdeman, Robyn Hartvickson, Amber Grant | Newton, KS |
| Alliance for Multispecialty Research | Terry L. Poling MD, Terry D. Klein MD, Thomas C. Klein MD, Tracy R. Klein MD | Wichita, KS |
| Alliance for Multispecialty Research | William B. Smith MD, Richard L. Gibson MD, Jennifer Winbigler MD, Elizabeth Parker PA | Knoxville,TN |
| Baptist Health Center for Clinical Research | Priyantha N. Wijewardane, MD, Eric Bravo MD, Jeffrey Thessing MD, Michelle Maxwell APRN, Amanda Horn APRN | Little Rock, AR |
| Baylor College of Medicine, NIAID 1UM1AI148575-01S2 | Hana El Sahly MD, Jennifer Whitaker MD, Catherine Mary Healy MD, Christine Akamine MD | Houston, TX |
| Benchmark Research | Laurence Chu, MD, R. Michelle Chouteau, MD | Austin, TX |
| Benchmark Research | Michael J. Cotugno MD, George H. Bauer, Jr. MD | Metairie, LA |
| Benchmark Research | Greg Hachigian MD, Masaru Oshita MD, Michael Cancilla NP, Deborah Murray NP, Kristen Kiersey NP | Sacramento, CA |
| Benchmark Research | William Seger MD, Mohammed Antwi, Allison Green, Anthony Kim | Fort Worth, TX |
| Brigham and Women’s Hospital, NIAID UM1AI069412, NCATS UL1RR025758 | Lindsey R Baden MD, Michael Desjardins MD, Jennifer A Johnson MD, Amy Sherman MD, Stephen R Walsh MD | Boston, MA |
| Carolina Institute for Clinical Research | Judith Borger DO, Ryan Starr DO, Scott Syndergaard DO, Nafisa Saleem MD | Fayetteville, NC |
| Centex Studies | Joel Solis MD, Martha Carmen Medina PA-C, Westly Keating PA-C, Edgar Garcia PA-C, Cynthia Bueno PA-C | McAllen, TX |
| Clinical Research Atlanta | Nathan Segall MD, Nathan Segall, Jon Finley, Mildred Stull | Stockbridge, GA |
| Clinical Trials of Texas | Douglas Scott Denham DO, Thomas Weiss MD, Ayoade Avworo DNP, Parke Hedges MD | San Antonio, TX |
| Coastal Carolina Research Center | Cynthia Becher Strout MD, Rica Santiago, Yvonne Davis, Patty Howenstine, Alison Bondell | Mount Pleasant, SC |
| Cornell Clinical Trials Unit - Weill Cornell Uptown & Weill Cornell Chelsea, NIAID UM1AI068619,  NCAT UL1TR002384 | Kristin Marks MS MD, Grant Ellsworth, MS, MD, Tina Wang, MD, Timothy Wilkin, MD, MPH, Mary Vogler, MD, Carrie Johnston, MD, MS | New York, NY |
| Covid19 Prevention Network (CoVPN, NIAID-NIH) | Michele P Andrasik, Jessica G Andriesen, Gail Broder, Lawrence Corey, Niles Eaton, Kathleen M Neuzil, Huub G Gelderblom, James G Kublin, Rachael McClennen, Nelson  Michael, Merlin Robb, Carrie Sopher | Seattle, WA |
| DM Clinical Research | Vicki E. Miller MD, MPH, Fredric Santiago MD, Blanca Gomez FNP-C, Insiya Valika PA-C, Amy Starr FNP-C | Tomball, TX |
| Emory University – Ponce de Leon Clinical Research Site, NIAID 3UM1AI068614-14S1 | Colleen Kelley MD MPH, Valeria D Cantos MD, Sheetal Kandiah MD MPH, Carlos del Rio MD | Atlanta, GA |
| Emory University – Hope Clinic, NIAID 1UM1AI148576-01 | Nadine Rouphael MD, Paulina Rebolledo, Srilatha Edupuganti, Daniel Sans Graciaa | Decatur, GA |
| Emory University School of Medicine, NIAID 1UM1AI148576-01 | Evan J Anderson MD, Andres Camacho-Gonzalez MD, Satoshi Kamidani MD, Christiana A Rostad MD, Meghan Teherani MD | Atlanta, GA |
| George Washington University, NIAID UM1AI068619 | David Joseph Diemert MD, Elissa Malkin, Marc Siegel, Afsoon Roberts, Gary Simon | Washington, DC |
| Hackensack University Medical  Center | Bindu Balani MD, Carolene Stephenson, Steven Sperber, Cristina Cicogna | Hackensack, NJ |
| Henry Ford Health System | Marcus J. Zervos MD, Paul Kilgore MD, MPH, Mayur Ramesh MD, Erica Herc MD, Kate Zenlea MPH | Detroit, MI |
| Hope Research Institute | Abram Burgher MD, Ann Marie Milliken | Phoenix, AZ |
| Hope Research Institute | Joseph D. Davis MD, Brendan Levy, Sandra Kelman | Chandler, AZ |
| Hope Research Institute | Matthew W. Doust MD, Denise Sample, Sandra Erickson | Phoenix, AZ |
| J. Lewis Research | Shane Glade Christensen MD, Christopher Matich, James Longe, John Witbeck | Salt Lake City, UT |
| J. Lewis Research | James Todd Peterson MD, Alexander Clark, Gerald Kelty, Issac Pena-Renteria | Salt Lake City, UT |

| **Affiliation/Funding*** | **Study Group** | **Location** |
| --- | --- | --- |
| Jacksonville Center for Clinical Research | Michael J. Koren MD, Darlene Bartilucci MD, Jeffery Jacqmein MD, Alpa Patel MD, Carolyn Tran MD | Jacksonville, FL |
| Javara | Christina Kennelly MD, Robert Brownlee, Jacob Coleman, Hala Webster | Charlotte, NC |
| Johnson County Clin-Trials | Carlos A. Fierro MD, Natalia Leistner, Amy Thompson, Celia Gonzalez | Lenexa, KS |
| Kaiser Permanente Washington Health Research Institute, NIAID 1UM1AI148373-01 | Lisa A Jackson MD MPH, Janice Suyehira MD | Seattle, WA |
| Laguna Clinical Research Associates | Milton Haber MD, Maria M. Regalado MD, Veronica Procasky RN JD, Alisha Lutat | Laredo, TX |
| Lynn Health Science Institute | Carl P. Griffin MD, Raymond Cornelison, William Schnitz, Shanda Gower | Oklahoma City, OK |
| Lynn Institute of the Rockies | Ripley R. Hollister MD, Jeremy Brown DO, Melody Ronk PA-C | Colorado Springs, CO |
| M3 Wake Research | Wayne Lee Harper MD, Lisa Cohen DO, Lynn Eckert PA-C, Matthew Hong MD | Raleigh, NC |
| MediSync Clinical Research Hattiesburg Clinic | Rambod Rouhbakhsh MD, MBA, Elizabeth Danford MD, John Johnson MD, Richard Calderone MD | Petal, MS |
| Meridian Clinical Research | Shishir Kumar Khetan MD, Oyebisi Olanrewaju AC-CRNP, Nan Zhai NP-C, Kimberly Nieves AC-CRNP, Allison O'Brien AC-CRNP | Rockville, MD |
| Meridian Clinical Research | Paul Simon Bradley MD, Amanda Lilienthal MSN NP-C, Jim Callis PA-C | Savannah, GA |
| Meridian Clinical Research | Adam Benson Brosz MD, Andrea Clement PA, Whitney West APRN, Luke Friesen PA, Paul Cramer APRN | Grand Island, NE |
| Meridian Clinical Research | Frank Steven Eder MD, Ryan Little FNP, Victoria Engler FNP, John Tarbox FNP, Heather Rattenbury-Shaw DO | Binghamton, NY |
| Meridian Clinical Research | David Jon Ensz MD, Tavane Harrison, Allie Oplinger | Dakota Dunes, SD |
| Meridian Clinical Research | Brandon James Essink MD, Jay Meyer MD, Frederick Raiser, III MD, Kimberly Mueller APRN, Roni Gray PA | Omaha, NE |
| Meridian Clinical Research | Keith William Vrbicky MD, Charles Harper MD, Chelsie Nutsch MD, Wendell Lewis III MD, Cathy Laflan MD | Norfolk, NE |
| Meridian Clinical Research | Jordan L. Whatley MD, Nicole Harrell MD, Amie Shannon MD, Crystal Rowell APRN, FNP- C, Christopher Dedon APRN, FNP-C | Baton Rouge, LA |
| NIH | Mamodikoe Makhene MD MPH | Bethesda, MD |
| New Horizons Clinical Research | Gregory Mark Gottschlich MD, Kate Harden PA-C, Melissa Gottschlich PA-C, Mary Smith MSN, FNP-C, Richard Powell MD | Cincinnati, OH |
| Optimal Research | Murray A. Kimmel DO, Simmy Pinto MD | Melbourne, FL |
| Optimal Research | Timothy P. Vachris MD, Mark Hutchens MD, Stephen Daniels DO, Margaret Wells MD | Austin, TX |
| Optimal Research | Mimi Van Der Leden MD, PhD, Peta Gay Jackson Booth MD | Rockville, MD |
| Palm Beach Research Center | Mira Baron MD, Pamela Kane DO, Shannen Seversen PA-C, Mara Kryvicky PA-C, Julia Lord PA-C | West Palm Beach, FL |
| Paradigm Clinical Research Center | Jamshid Saleh MD, Matthew Miles, Rafael Lupercio | Redding, CA |
| Quality of Life Medical & Research Centers | John W. McGettigan Jr. MD, Walter Patton MD, Riemke Brakema MD, Karin Choquette MSN, ABNP-C, Jonlyn McGettigan MSN, RN | Tucson, AZ |
| Rancho Paseo Medical Group | Judith L. Kirstein MD, Marcia Bernard NP | Banning, CA |
| Rapid Medical Research | Mary Beth Manning MD, Joan Rothenberg MD, Toby Briskin MD, Denise Roadman PAC, Sharita Tedder-Edwards FNP | Cleveland, OH |
| Research Centers of America | Howard I. Schwartz MD, Surisday Mederos, Barbara Corral, Jennifer Schwartz, Nelia Sanchez-Crespo | Hollywood, FL |
| Rutgers New Jersey Medical School, NIAID UM1AI068619 | Shobha Swaminathan MD, Amesika Nyaku MD MS, Tilly Varughese MD, Michelle DallaPiazza MD | Newark, NJ |
| Saint Louis University, NIAID 1UM1AI148685-01 | Sharon E Frey MD, Irene Graham MD, Getahun Abate MD PhD MSc, Daniel Hoft MD PhD | St. Louis, MO |
| St. Vincent's Health System | Leland N. Allen III MD, Leslie Anne Edwards MSN, CRNP, William Simpson Davis Jr., MS PA-C, Jessica Maria Mena, PA | Birmingham, AL |
| Suncoast Research Group | Mark E. Kutner MD, Jorge Caso MD, CPI, Maria Hernandez Moran APRN, Marianela Carvajal APRN, Janet Mendez APRN | Miami, FL |
| Sundance Clinical Research | Larkin T. Wadsworth III MD, Horacio Marafioti, Lyly Dang, Jennifer Berry, Lauren Clement | St. Louis, MO |
| Synexus Clinical Research | Michael Ryan Adams MD, Leslie Iverson PA | Murray, UT |
| Synexus Clinical Research | Joseph Lee Newberg MD, Laura Pearlman MS, MD, MBA | Chicago, IL |
| Synexus Clinical Research | Paul Joseph Nugent DO, Leonard Singer | Cincinnati, OH |

| **Affiliation/Funding*** | **Study Group** | **Location** |
| --- | --- | --- |
| Synexus Clinical Research | Michele Diane Reynolds MD, Jennifer Bashour MD, Robert Schmidt MD | Dallas, TX |
| Synexus Clinical Research | Neil Parmanand Sheth MD, Kenneth Steil DO | Glendale, AZ |
| Synexus Clinical Research | Ramy Joseph Toma MD, William Kirby MD, Pink Folmar MD, Samantha Williams NP | Birmingham, AL |
| Synexus Clinical Research | Judith White MD, Robert Meyer MD, Sejal Patel MD, Prity Patel APRN | Orlando, FL |
| Tekton Research | Paul Pickrell MD, Stefanie Mott FNP-C, Carol Ann Linebarger MD, Hussain Malbari MD, David Pampe MD | Austin, TX |
| Texas Center for Drug Development | Veronica G. Fragoso MD, Lisa Holloway MD, Cecilia McKeown-Bragas MD, Teresa Becker MD, Vicki Miller MD | Houston, TX |
| Trial Management Associates | Barton G. Williams MD, William H. Jones MD | Wilmington, NC |
| VA Greater Los Angeles Healthcare System | Michael Lewis MD, Elham Ghadishah, Joseph Yusin, Mai Pham | Los Angeles, CA |
| University of California Los Angeles, NIAID UM1AI068619 | Jesse L Clark MD, Steven Shoptaw PhD, Michele Vertucci PA, NP, Will Hernandez NP | Los Angeles, CA |
| University of California San Diego, NIAID UM1AI068636 | Stephen A. Spector MD, Amaran Moodley MD, Jill Blumenthal MD, Lisa Stangl NP, Karen Deutsch NP | La Jolla, CA |
| University of Chicago | Kathleen M. Mullane DO PharmD, David Pitrak MD, Cheryl Nuss FNP, Judy Pi PharmD | Chicago, IL |
| University of Cincinnati, NIAID UN1AI068619 | Carl Fichtenbaum MD, Margaret Powers-Fletcher PhD, Michelle Saemann RN, Sharon Kohrs RN | Cincinnati, OH |
| University of Colorado Denver,  Anschutz Medical Campus, NIAID UM1AI068636 | Thomas B. Campbell MD, Andrew Lauria, Jose Castillo Mancilla, Hillary Dunlevy | Aurora, CO |
| University of Illinois at Chicago –  Project WISH, NIAID UM1AI068619 | Richard M Novak MD, Andrea Wendrow, Scott Borgetti, Ben Ladner | Chicago, IL |
| University of Maryland School of Medicine, NIAID 1UM1AI148689-01 | Karen L Kotloff MD, Matthew Laurens, Milagritos Tapia, Lisa Chrisley, Cheryl Young | Baltimore, MD |
| University of Miami, NIAID 3UM1AI068614-14S1 | Susanne Doblecki-Lewis MD, Maria Luisa Alcaide, Jose Gonzales-Zamora, Stephen Morris | Miami, FL |
| University of North Carolina at Chapel Hill, NIAID UM1AI068619 | Cynthia Gay MD MPH, Erin Hoffman, Susan Pedersen, Maria Bullis, Mandy Tipton, Carolina Pastrana Medina, Catherine Kronk, Nicole Maponga, Julie Nelson, Becky Straub, Amy James Loftis, David Wohl MD, Joseph Eron, Jr. MD | Chapel Hill, NC |
| University of Pennsylvania, NIAID 3UM1AI068614-14S1 | Ian Frank MD, Debora Dunbar, David Metzger, Florence Momplaisir | Philadelphia, PA |
| University of Pittsburgh Medical Center, NIAID 1UM1AI148452-01 | Judith Martin MD, Alejandro Hoberman MD, Timothy Shope MD MPH, Gysella Muniz MD | Pittsburgh, PA |
| University of Texas Medical Branch, NIAID 1UM1AI148575-01 | Richard Rupp MD, Amber Stanford PA-C, Megan Berman MD, Laura Porterfield MD | Galveston, TX |
| VA Greater Los Angeles Healthcare System | Michael Lewis MD, Elham Ghadishah, Joseph Yusin, Mai Pham | Los Angeles, CA |
| Vanderbilt University Medical Center, NIAID 1UM1AI148452-01 | Clarence Buddy Creech II MD, Shannon Walker MD, Stephanie Rolsma MD PhD, Robert Samuels, Isaac Thomsen MD | Nashville, TN |
| Vanderbilt University Medical Center, NIAID 3UM1AI068614-14S1 | Spyros Andrews Kalams MD, Greg Wilson MD | Nashville, TN |
| Velocity Clinical Research | Gregg H. Lucksinger MD, Kevin Parks MD, Ryan Israelsen MD, Jaleh Ostovar FNP-C, Kary Kelly FNP-C | Medford, OR |
| Velocity Clinical Research, San Diego | Jeffrey Scott Overcash MD, Hanh Chu, Kia Lee, Karla Zepeda | La Mesa, CA |
| VitaLink Research | Luis I. De La Cruz MD, Steve Clemons, Elizabeth Everette, Suzanna Studdard | Greenville, SC |
| VitaLink Research | Gowdhami Mohan MD, Stefanie Tyson, Alyssa-Kay Peay, Danyel Johnson | Anderson, SC |
| VitaLink Research-Spartanburg | Gregory J. Feldman MD, May-Yin Suen, Jacqueline Muenzner, Joseph Boscia, Farhan Siddiqui | Spartanburg, SC |
| Wake Forest University Health Sciences | John Sanders MD,PhD, James Peacock MD, Julio Nasim MD | Winston Salem, NC |
| WR-Clinical Research Center of Nevada | Michael L. Levin MD, Julie Hussey MSN APRN FNP-C, Marcy Kulic MD | Las Vegas, NV |
| WR-ClinSearch | Mark Montgomery McKenzie MD, Teresa Deese, Erica Osmundsen, Christy Sweet | Chattanooga, TN |
| WR-Global Medical Research | Valentine Mbepson Ebuh MD MA MSc, Elwaleed Elnagar MD, Georgette Ebuh DNP APRN FNP-C, Genevieve Iwuala FNP | Dallas, TX |
| WR-Medical Center for Clinical Research | Laurie J. Han-Conrad MD, Todd Simmons MD, Denis Tarakjian MD | San Diego, CA |

*Funding of institutions by the National Institute of Allergy and Infectious Diseases (NIAID) and/or research support by the National Center for Advancing Translational Science (NCATS) as indicated. All other institutions were funded by Office of the Assistant Secretary for Preparedness and Response, Biomedical Advanced Research and Development Authority. The content of this publication is solely the responsibility of the authors and does not necessarily represent the official views of the funding sources.

### CoVPN/COVE Team (cont’d): COVE Trial Investigators and Study Teams

| **Principal Investigator** | **Study Team** | **Institution** | **Location** |
| --- | --- | --- | --- |
| Atoya Adams, MD, MBA | Miriah Campbell, Eric Miller, Daisy Langarica, Alia Bober, Diana Giraldo | AB Clinical Trials | Las Vegas, NV |
| Michael Ryan Adams, MD | Leslie Iverson, Andryelle Toledo, Melinda Bullington, Alicia Hanten, Carolyn Taylor, Shannon Wright, Chase Carnahan, Rachel Law, Natalie Smith, Julie Taylor, Jared- Robert Blake, Stefanie Vasconez, Courtney Jensen | Synexus Clinical Research | Murray, UT |
| Leland N. Allen III, MD | Leslie Anne Edwards, William Simpson Davis, Jr., Ronald Meza, Jordan Stauffer, John Farringer, Faith Holmes, Rhonda Buzbee, Cristina Velez, Huse Lisa, Lisa Huse, Camelia Speegle, Gregory Prestage, Mary Perez, Jessica Space, Matthew Todd, Jessica McDowell, Marha Bunnell-Pollak, Jackie Ziegler, Jasmine Ali, Dumitru Sirbu, Kellie Williams, Logan Sawyer, Richelle Chambliss, Samantha Blackmon, Stephanie Brennan, Tiffany Gibbs, Alexandria Anderson, Caitlin Roll, Candace Robinson,  Zachary McCoy, Jessica Bartlett, Kimberly Cornelison, Chris Bovell, Vincent Baglini, Christy Greenhalgh, Jessica Maria Mena, David House, Matt Honold, Esteban Zurita | St. Vincent's Health System | Birmingham, AL |
| Evan J. Anderson, MD | Kathleen Stephens, Francine Dyer, Maya Stagg, Aaliyah Carron, Austin Lu, Julia Barton, Sy Tran, Leisa Bower, Esther Park, Jianguo Xu, Rebecca Gonzalez, Vy Ngo, Mike Shepard, Lezly Roxxette Zepeda, Karen Sytsma, Sandra Rojas-Honan, Felicia Glover, Susan Rogers, Theda Gibson, Christina A. Rostad, Andres Camacho- Gonzalez, Teresa Ball, Satoshi Kamidani, Mehgan Farah Teherani, Vikash Patel, Etza Peters, Peggy Kettle, Lisa Macoy, Cindy Lubbers, Amber Samuel, Laila Hussaini,  Kathryn Zaks, Caroline Ciric, Meg Taylor, Oliver Smith, Amy Muchinsky, Sydney Biccum, Laura Clegg, Dean Kleinhenz, Angelle Ijeoma, Hannah Huston | Emory University School of Medicine | Atlanta, GA |
| Lindsey Baden, MD | Xhoi Mitre, Jon Gothing, Bruce Bausk, Jessica Cauley, Natalie Izaguirre, Lewis Novack, Michael Seaman, Katherine Yanosick, Henry Rutherford, Junghyun Kim, Dominique Betterbed, Kathleen Garvey, Lauren Clore, Alexander Mills, Deepesh Duwadi, Alessandra Setaro, Kyl Bowman, Kevin McManus, Sidali Beriane, Fadi Ghantous, Christy Lavine, Jasper Ophel, Joseph Sapiente, Jessica Dorning, Tessa Speidel, Lauren Garneau, Robert Dannemiller, Kirquenique Rolle, Mulika Chhorn, Bailey McCarthy, Hana Flaxman, Milenko Tanasijevic, Cameron Nutt, Javier Barria, Andre Avila-Paz, Buteau Malhaika, Tong Alexandra, Tenaizus Woods, Bethany Evans, Hannah Jin, LaKeisha Gandy, Stephanie St. Pierre, Carolyn Darcy, Michael Corrado, James Maguire, Adetoun Okenla, Tamara Roldon Sevilla, David Kubiak, Cassandre Titus, Movita Harrigan, Maria Alvarado, Rose Theodat, Amy Sherman, Laura Platt, Kirsten Goodman, Laura Nicholson, Wilfredo Matias, Emily Koleske, Ruth Rodriguez, Nicole Taikeff, Jun Bai Park Chang, Julia Klopfer, Phoebe Cunningham, Elizabeth Sampson, Karen Magsipoc, Maureen Macgowan, Lauren Donahue, Haley Schram, Noah Abasciano, Megan Powell, Janet Morgan, Yazed Alsowaida, Olivia Riccardi, Neha Limaye, Virginia Loudermilk, Austin Kim, Kevin Zinchuk, Caitlin Grant, Charles Kelly, David Mellace, Jamie Myers, Erika Gribb, Jose Licona, Monica Feeley, Stephen R Walsh, Jennifer A Johnson, Ann Woolley, Alexis Liakos, Jane Kleinjan,  Jon Gothing, Nicolas Issa, Michael Desjardins, Raphael Dolin, Alka Patel, Opeyemi Talabi, Christin Price, Paulette Chandler, Elizabeth W Karlson, Allison P Moriarty | Brigham and Women's Hospital | Boston, MA |
| Bindu Balani, MD | Smith Kerowyn, Sergio Garcia, Charo Valdez, Shelly Chin, Caitlin DiBello, Silvia Lara, Chika Ekweghariri, Abena Roberts, Abimbola Coker, Marie-Therese Estanbouli, Greg Eskinazi, Michael Tortoriello, Jay Elkareh, Meral Karakoc, Olga Spathis, Patrice Hassoun, Carolene Stephenson, Steven Sperber, Kaur Harveen, Cristina Cicogna, Ciaran Mannion | Hackensack University Medical Center | Hackensack, NJ |
| Mira Baron, MD | Pamela Kane, Maria Bermudez, Shannen Seversen, Mara Kryvicky, Julia Lord, Terri Barr, Daisy Acevedo, Elena Acosta, Delta Anderson, Alexandra Arango, Anne Bauer, Joshua Egbehor, Tim Flanary, Audrey Haber, Carol Henao, Patti Isaacson, Peter Jacob, Sakaiya Jackson, Karen Kodes, Ludovic La-Branche, Kimarie Lee- Russell, Carol Liso, Cristina Liso, Stephanie Morse, Michelle Navarrette, Christy Norcross, Nora Norcross, Annette Pitts, Mary Sergalis, David Scott, Tytiana  Spearman, Danielle Theodore, Brian Thomas, Jennifer Torres | Palm Beach Research Center | West Palm Beach, FL |
| Judith Borger, DO | Jennifer Angell, Nicole Austin, Deanna Benz, Lucian Cappoli, Nicole Davis, Lynn Eckert, Kathryn Hostetter, Stephanie Keating, Jeanette Mangual-Coughlin, Avia McClain-Stocker, Ifeanyi Momodu, Cheryl Norris, Brennan Opanasenko, Stacey Saldua, Nafisa Saleem, Amy Sheets, Ryan Starr, Scott Syndergaard, Jennifer  Thomas, Michelle Wallace, Jeffery Pemberton, Mitchell Arildsen, Dan Tomita | Carolina Institute for Clinical Research | Fayetteville, NC |
| Paul Simon Bradley, MD | Taja Adams, Stephanie Ailey, Kira Bell, Shanice Bennett, Vincent Bernades, Jim Callis, Bounphone Chanthavong. Taryn Collett, Anne Crouch, Shannon Davis, Morgan Deal, Mimi Duncan, Brandon Essink, Laura Falcone, Debra Gabrielson, Brooke Halpern, Anyfa Hanna, Cassie Heisey, Dawn Kalloniatis, Andrew Kimball, Jeanette Lee, Amanda Lilienthal, Ginny McNew, Crystal Neely, Kay Lynn Olmsted,  Nicole Osborn, Chevon Roberts, Pechoka Sanders, Cynthia Seedorf, Kathryn Stoddard, Jonathan Whelan, Stella Yoon | Meridian Clinical Research | Savannah, GA |

| **Principal Investigator** | **Study Team** | **Institution** | **Location** |
| --- | --- | --- | --- |
| Adam Benson Brosz, MD | Rhonda Richter, Debra Gabrielson, Kayla Flege, Ashley Bell, Karen Jo Johnson, Paul Cramer, Jessica Stanton, Andrea Clement, Whitney West, Laura Falcone, Amanda Friesz, Kathy Osborne, Summer Tophoj, Kimber Breeden, Susan Newman, Douglas  Herbek, Lindsey Mettenbrink, Luke Friesen, Alison Pierce | Meridian Clinical Research | Grand Island, NE |
| Abram Burgher, MD | Stephanie Catanzaro, Shauna Harrell, Magen Hess, Nate Alderson, Bettie D'Nise Corcoran, Norma Frederick, Adrian Alejo, Brian DeCraene, Karen Wakefield, Scarlett  Hammett, Susan DeCraene, Ann Marie Milliken, Neil Pearson, Donald Terral Harper | Hope Research Institute | Phoenix, AZ |
| Thomas B. Campbell, MD | Andrew Lauria, Jenelynn Kimble, Steven Johnson, Matin Krsak, Andrew Monte, Patrisha Adkins, Michelle Barron, Suzanne Fiorillo, Amy Harrison, Anderson Victoria, Nga Le, Sara Berech, Jose Castillo-Mancilla, Kristine Erlandson, Laurel Ware, Josie Marshall, Stephen Bartlett, Hillary Dunlevy | University of Colorado Denver, Anschutz Medical Campus | Aurora, CO |
| Shane Glade Christensen, MD | Christopher Mickelson, Jessica Shaw, Emily Raming, Amy Nelson, Gabrielle Lewis, Jenessa Folsom, Mikaela Jones, Dylan Owen, Rachel Pugmire, Jennifer Bradley, Annjanette Kemp, Krista Marti, Allyson Christensen, Madison Ellis, Holly Anderson, Emily Bloomquist, Ross Brunetti, Thomas Conner. Jr., Gina Cox, Diana Grazulis, Wesley Lewis, James Longe, Christopher Matich, Bryan Nelson, Sarah Scott, John  Witbeck, Stephen Wood | J. Lewis Research | Salt Lake City, UT |
| Laurence Chu, MD | Jennifer Bacchi, Maria Barrientes, Lamar Box, Christian Casas, R. Michelle Chouteau, Katherine Davis, Tambra Dora, Cindy Duran, Pamela Fidler, Ruth Fitch, Brooke Harris, Isaiah Knight, Jennifer Leyva, Michelle Listz, Jennifer Montes, Javier Perez, Jessica Ruff, Dean Skiles, Sean Turnbow, Francesca Vigil, Breana Wade, Kelly Weber | Benchmark Research | Austin, TX |
| Jesse L. Clark, MD | Sandy MacNicoll, Somaieh Talebi, Timothy Hall, Steven Shoptaw, Emery Chang, Michael Li, David Goodman, Paul Adamson, Oladunni Adeyiga, Inez Bentancourt, Susan Reed, Christopher Blades, Jasmin Tavares, Demetria Villanueva, Simone Riley, Jonathan Veloz, Schuyler Thomas, Will Hernandez, Jennifer Baughman,  Mitchell Stern, Michele Vertucci | University of California, Los Angeles | Los Angeles, CA |
| Michael J. Cotugno, MD | Kyra Lawson, Kim Harper, Edwin Adamson, George H. Bauer Jr., Julie Bilich, Brenda Lawson, Brandon Illickal, Lois Eaglin, Heather Salisbury, Jeff Segner | Benchmark Research | Metairie, LA |
| Clarence Buddy Creech II, MD | Shanda Phillips, Naomi Kown, Katherine Sokolow, Wendy Winn, Katherine Wright, Shannon Walker, Stephanie Rolsma, Anna Gallion, April Hanlotxomphou, Deborah Myers, Robert Adkisson, Natalia Jimenez, Cindy Trimmer, Roberta Winfrey, Matthew Donio, John Oleis, Donna Torr, Shelly McGehee, Robert Samuels, Sandra Yoder, Eric Brady, Isaac Thomsen, Madeleine Guy, Emma Alexander, Lana Howard, Krisha Alexander, Shane Moore, Tacora Wright, Tara Evans, Ursula Powell, Jenna Caserta, Valerie Mitchell, Meryk Moore, Melissa Lehman, Diane Anders, Constance Dotye,  Crystal Rice, Lamar Bowman, Sherri Hails, Monique Bennett, Nicki Soper, Leigh Howard | Vanderbilt University Medical Center | Nashville, TN |
| Joseph D. Davis, MD | Sandra Kelman, Sandra Braden, Sabrina Bolland, Mia Munoz, Jose Barocio, Brendan Levy, Dhwani Shah, Neil Pearson, Stephanie Catanzaro, Nathan Alderson, Susan  DeCraene, Maureen Godfrey, Skyla Clark | Hope Research Institute | Chandler, AZ |
| Luis I. De La Cruz, MD | Amy Ford, Taylor Wilson, Cindy Smith, Austin Lambert, Erin Zeiler, Kaelyn Rowland, Marlee Smith, Suzanna Studdard, Zandra Hamilton, Meredith Benfield, Sara Poff, David Godwin, Elizabeth Everette, Steven Clemons, Kayla Peay, Stephanie Gilreath | VitaLink Research | Greenville, SC |
| Douglas Scott Denham, DO | Thomas Weiss, Parke Hedges, Ayoade Avworo, Kay Scroggins, Leisel Koerber, Antonio Gutierrez, Nathan Cortez, Andrea Gomez, Darlington Akahara, Michelle Smith, Kristy Trevino, Beatriz Herrera, Shaiane Dickerson, Kerry de Jesus, Matthew Korte, Cynthia Ramos, Reanna Martinez, Erica Leal, Shakera Flores, Paul Esparza, Brian Hemming, Melinda Axton, D’Andre White, Terri Perez, Carolina Coronado, Rebecca Many, Clayton Stone, Kimberly Evans, Anshumaan Maharaj, Stephen Brick, Steffanie Barrera, Staci Poettgen, Dawn Killian, Gerardo Pena, Karol Perez, Victoria Hernandez, Kevin Martinez, Amy Griffith, Nolan Payton, Quincey Hogue, Jamie  Padilla, Emily Mendez, Lily Hays, Maristelle Co, Nicholas Trinidad, Ismael Rodriguez, Amy Lewis, Cindi Nellis, Lele Simmons, Marissa Johnson | Clinical Trials of Texas | San Antonio, TX |
| David Joseph Diemert, MD | Linda Witkin, Aimee Desrosiers, DeEnna Wedding, Bertran Walton, LaKeisha Queen, Ryan Mouton, Caroline Thoreson, Manya Magnus, Jennifer Wald, Erika Faust, Nicholas Heredia, Robbie Kattappuram, Hira Qadir, Chelsea Ware, Hannah Yellin, Kegan Dasher, Daniel Mullen, Jeanne Jordan, Taylor Ladson, Madison Lintner, Kaitlyn Macnair, Bitana Saintilma, Kelly Thomas, Samantha Walker, Neha Rampally, Madhu Balachandran, Elissa Malkin, David Parenti, Hana Akselrod, Marc Siegel, Gary Simon, Afsoon Roberts, Aileen Chang | George Washington University | Washington, DC |

| **Principal Investigator** | **Study Team** | **Institution** | **Location** |
| --- | --- | --- | --- |
| Susanne Doblecki- Lewis, MD | Maria Luisa Alcaide, Jose Gonzalez-Zamora, Stephen Morris,  Yimy Puerto, Annie Salvarrey, Claudia Balgas, Claudia Santos, Katherine King, Brahian Steven Erazo, Mayra Fernandez, Leopoldo Cordova-Garcia, Elisa Corzo- Sanchez, Edgar Fernandez, Loreta Padron, Stefani Ann Butts, Kenia Moreno, Juan Casuso, Maria de Pilar Valanzasca, Thomas Tanner, Marilyn Fernandez, Mary Aloise, Inza Patton, Vivian Pastrana, Sendy Puerto, Irma Barreto Ojeda, Junlin Long, Barbara Huang, Gilianne Narcisse, Vanessa Perez | University of Miami | Miami, FL |
| Matthew W. Doust, MD | Denise Sample, Sandra Erickson, Nate Alderson, Adrian Alejo, Stephanie Catanzaro, Susan DeCraene, Cassie Enricco, Sandra Erickson, Alex Guereque, Shauna Harrell, Shana Harshell, Stephanie Junker, Stephanie Laufenberg, Madison Mikulak, Makayla Morra, Nicole Olson, Neil Pearson, Jasmin Redden, Monique Romo, Denise Sample,  Dhwani Shah, Sahara Vega, Emma Kar | Hope Research Institute | Phoenix, AZ |
| Valentine Mbepson Ebuh, MD | Elwaleed Elnagar, Georgette Ebuh, Genevieve Iwuala, Catina Adams, Marissa Cervenka, Ezgar Del Real, Shraddha Dubal, Elwaleed Elnagar, Jenifer Fiatte, Kathy Harrell, Genevieve Iwuala, Vicki Martinez, Robert Miranda, Brennan Opanasenko,  Destiny Robinson, Liz Ruiz, Amy Sheets, Shoniece Wallace | WR-Global Medical Research | Dallas, TX |
| Frank Steven Eder, MD | Ryan Little, Victoria Engler, John Tarbox, Heather Rattenbury-Shaw, Deborah Hubish, Jessie Taylor, Debra Gabrielson, Jessica Fellows, Jennifer  Molstead, Kathe Olmstead, Ashley Conover, Tammy Kohn, Chelsea Briar, Corrine Young, Collen McVannan, Kelli Quick, Shaylynne Hubanks, Kimber Breeden, Ann Marie Sampson, Traci Hull, Tarin Gordon, Susan Owen, Kate Macarak, Tonya Rackett, Jacob Blattstein, Partidge Jane Aton, Nicole Croft, Carolyn Grausgruber, Rebecca Miller, Ryan Little, Victoria Engler, John Tarbox, Heather Rattenbury-Shaw, Nathan Kimball, Courtney Heisey, Ginny McNew, Abigail Wine, Cindi VanKuren,  Jared Frick, Tammy Dennis, Andrew Kimball | Meridian Clinical Research | Binghamton, NY |
| Hana M. El Sahly, MD | Jennifer A. Whitaker, C. Mary Healy, Christine Akamine, Wendy A Keitel, Robert L Atmar, Annette Nagel, Sandra Francisco, Thea Marie Cordero, Janet Brown, Jennifer Christensen, Caroline Doughty-Skierski, Connie Rangel, Carrie Kibler, Coni Cheesman, Lisreina Toro, Chanei Henry, Chianti Wade Bowers, Pedro Piedra, Kathy Bosworth, Kayla Burrell, Jesus Banay, Tykel Eddy, Trent Davis, Shetel Anassi, Yvette  Rugeley, Olga Rybina-Willis | Baylor College of Medicine | Houston, TX |
| David Jon Ensz, MD | Pamela Allen, Taylor Bergh, Kimber Breeden, Avery Dunn, Brandon Essink, Debra Gabrielson, Rylea Gulick, Tavane Harrison, Courtney Heisey, Andrew Kimball, Shelby Klaschen, Jessica Knight, Makayla Langston, Meagan Miller, Allie Oplinger, Heather Persinger, Alison Pierce, Kathryn Stoddard, Kayla Sturgeon, Jamie Thompson,  Melissa Wiseman | Meridian Clinical Research | Dakota Dunes, SD |
| Brandon James Essink, MD | Jay Meyer, Frederick Raiser, Kimberly Mueller, Roni Gray, Riley Brockman, Tabitha Campbell, Carrie Essink, Laura Falcone, Roni Gray, Linda Layton, Jay Meyer,  Kimberly Mueller, Tiffany Nemecek, Frederick "Fritz" Raiser, III, Jessica Satorie, Chelsea Steinmetz, Nicole Osborn, Cassie Heisey, Maria Nguyen | Meridian Clinical Research | Omaha, NE |
| Gregory J. Feldman, MD | May-Yin Suen, Brittany Cooksey, Madison Fowler, Sarah Chynoweth, Gary Clemons, Laura Jolly, Charlie Jordan, Heather Allison, Steve Clemons, Amber Brittany Belcher, Allison Kelly, Marsha Gossett, Wendy Taylor, Amy Witt, Kendal Nelson, Jeffrey Witt,  Jacqueline Muenzner, Elizabeth Everette, Supinder Channa, Allison Ayers, Joseph Boscia, Farhan Siddiqui | VitaLink Research- Spartanburg | Spartanburg, SC |
| Carl J. Fichtenbaum, MD | Maggie Powers-Fletcher, Michelle Saemann, Sharon Kohrs, Kimberly Mullins, Lindsay Davis, Moises Huaman, Angela Snyder, Kristin Weghorn, Brenda Miller, Elizabeth Costea, Lisa Schira, Romana Saeed, Helen Shelton, Kathleen Ballman, Laura Browning-Cho, Sherry Donaworth, Chris Goddard, Jeanine Goodin, Elizabeth Niederegger, Lisa Hachey, Tamara Maus, Pam Fletcher, Makayla Bishop, Victoria Straughn, Shaina Horner, Carrie Christofield, Dana Burns, Jason Mayes, Kelly Windholtz, Lisa Proffitt, Faizan Qureshi, Michelle O'Neil, Arustamyan Lisa, Sarah Trentman, Eva Whitehead, Jennifer Baer, Linda Hinds, Jaasiel Chapman, D’Vaughn House, Gary Frazier, Judy Houston, Lisa Altenau, Mary Burns, Dorice Smith, Justin Ragle, Eric Mueller, Cynthia Nypaver, Jaime Robertson, Anissa Moussa, Geronimo  Feria Garzon, Sierra Bennett, Marlena Petrie | University of Cincinnati | Cincinnati, OH |
| Carlos A. Fierro, MD | Mazen Zari, Celia Gonzalez, Natalia Leistner, Mary Easley, Mary Provost, Krista Estrada, Ann Geier, Amy Thompson, Heather Barker, Karol Moore, Kelly Moen, Monica Atwood, Amber Wolf, Brandi Dickerson, Manyvohn Rinehart, Dina Hammine, Angela Eichler, Casey Johnson, Nathan Arthur | Johnson County Clin-Trials | Lenexa, KS |
| Veronica G. Fragoso, MD | Lisa Holloway, Cecilia McKeown-Bragas, Teresa Becker, Vicki Miller, Leena Mir, Elton Oliveira, Moez Talpur, Enya Rentas-Sherman, Gabriela Maria Becerra, Dewayne Hicks, Robert Krbashyan, Shakira Barr, Ashraf Jafri, Herman Ortiz, Zohair Harianawala, Chandra Tobin, Norma Gonzalez, Saji Perinjelil, Khorshid Amirkhosravi, Tracy Kowalski, Biman Goswami, Waheeda Sureshbabu, Amy Anderson, Berenice  Ferrero, Simeen Khan, Chen-Ho Yang, Nazanin Zarinkamar, Scott Ward, Crystal Reese, Miyosha Lewis, Olga Konshina, Lorrian Yates.Joel Cano, Quiana Wilson, Kara | Texas Center for Drug Development, Inc. | Houston, TX |

| **Principal Investigator** | **Study Team** | **Institution** | **Location** |
| --- | --- | --- | --- |
|  | Sikes, Diana Chehab, Joanna Quezan, Maryam Rabbani, Sadaf Batla, Abbyssinia Moges. Diego Carrington, Matthew Joseph, Laura Grissanty, Dean Jang, Dustin McFadden, Misbah Baloch, Elisa Moralez, Abdeali Dalal, Frances Saubon, William  Fernandez, Jenny Toress, Blessing Felix, Zain Rizvi |  |  |
| Ian Frank, MD | Annet Davis, Eileen Donaghy, Nicole Sundo, Juan Ramirez, Laura Schankel, Dana Brown, Katharine Bar, Dana Brown, Christopher Chianese, Gillain Constantino, Dovie Watson, Kathleen Degnan, Helen Koenig, William Short, Petra Alexander, Eileen Mergliano, Jie Ho, Michele Wisniewski, Debora Dunbar, Liani Santini-Lopez, Rosemarie Kappes, Angela Cabassa, Tammy Chen, Berry SotoVega, Deborah Kim, Devon Cliett, Kate Kearns, Jillian Baron, Vivian Leung, Florence Momplaisir, Sarah  Wood, Tameka Matthews, David Metzger, Richard Tustin | University of Pennsylvania | Philadelphia, PA |
| Sharon E. Frey, MD | Irene Graham, Getahun Abate, Daniel Hoft, Heather Douds, Cassandra Zehenny, Joan Siegner, Helay Hassas, Kim Cooper, Shirley Dettlebach, Sabrina DiPiazza, Carol Duane, Linda Eggemeyer-Sharpe, Lauren Foreman, Jerry Hutter, Ryan Kerr, Kate Liefer, Tracy Montauk, Karla Mosby, Janice Tennant, Nicole Purcell, Kiana Wilder, Kathleen Chirco, Sharon Irby-Moore, Kathleen Koehler, Melissa Loyet, Thomas Pacatte, Susan Stewart, Azra Blazevic, Tamara Blevins,  Chase Colbert, Christopher Eickhoff, Lainey Mejia Jauregui, Keith Meyer, Krystal Meza, Amanda Nethington, Huan Ning, Brittany Williams, Mei Xia, Yinyi Yu, Stanley Doublin, Mary Pat Eastman, Eric Eggemeyer, Mikayla Frye, Michelle Harris, Aleshia  McCoy, Donna Duncan, Gwendolyn Tatum, Nicole Purcell, Kiana Wilder, Tammy Grant, Claudia Castillo Paredes, Rong Hou, Jin Wang, Qian Wang, Sarah George | Saint Louis University | St. Louis, MO |
| Cynthia Gay, MD | David Wohl, Joseph Eron, Jr., , Janette Goins, Ulrike Adam, Ekundayo Nylander- Thompson, Anna Furlong, XinHong Ao, Kathy Guerrero, Melinda Hart, Kathleen Loeven, Rachael Mossey, Esther Speight, Rachel White, Chloe Twomey, Kristen Gray, , Patti Vasquez, April Welch, Camille O’Reilly, Maureen Furlong, Noshima Darden-Tabb, Elizabeth DuBose, Marie Oriol, Dynesha Perry, Maria Stetson, Maria Bullis, Shelby Turner, Ebony Harrington, Alexander Bradley, Susan Pedersen, Becky Straub, Sandra Barnhart, Tevnan Keller, Mandy Tipton, Abigail Riddick, Kristi Kirkland, Maggie Harman, Tania Hossain, Centhla Washington, Erin Hoffman, Carolina Pastrana Medina, William Johnson, Samantha Earnhardt, Amy James Loftis, Catherine Kronk, Yaa Ofori-Marfoh, Julie A Nelson, Nicole Maponga, Lina Rosengren-Hovee, William Zhao, Jennifer Thompson, Sarah Law, Holly Milner, Jonathan Oakes, Rachel Cook, Erin Cardot, Oesa Vinesett, Victoria  Rucinski, Joy Wannamaker, Tanailly Giralt Smith, Eliza DuBose, Chidinma Okafor | University of North Carolina at Chapel Hill | Chapel Hill, NC |
| Richard M. Glover, II, MD | Stacy Slechta, Troy Holdeman, Robyn Hartvickson, Amber Grant, Jennifer Bennett, Lindsey Brewer, Janelle Brown, Kelsey Burden, Melissa Burton, Brianna Burton, Jordan Danby, Sheri Duncan, Amber Grant, Robyn Hartvickson, Lisa Hemmelgarn, Sherry Henning, Jeri King, Riley King, Colton King, April Kitterman, Shannen Lassiter, Cayla Lawless, Janna Martinez, Ragene Moore, Marissa Mueller, Aaron Nguyen, Justin Phillips, Jordan Reheis, Rebecca Ring, Katherine Saengerhausen, Shannon Thomas, Dylan Thomas, Cindy Thome, Denae Villines, Amber Wenzel, Eilleen Wilbert, Avi Woods, Caressa Presley, Brianna Newport, Olivia Allen, Miranda  Santiago, Cheryl Sauerwein, Jill Longstaff, Sadie Allen, Candace Heckart | Alliance for Multispecialty Research | Newton, KS |
| Gregory Mark Gottschlich, MD | Melissa Gottschlich, Steven Anderson, Gregory Mark Gottschlich II, Mary Woeste, Kate Harden, Cindy Young, Michael Pordy, Audrius Ruksenas, Lacy Baird, Kim Krogman, Lori Stanton, Melissa Fuson, Mason Urban, Christine Watson, Richard Powell, Mary Smith, Jacob Sekinger, Diamond Russell, Nicole Lim, Mylene Asmar- Rios, Yusef Museitif, Craig Mitchell, Tarik Whitham, Zachary Rutledge, Troy Porter, Andrea Newlands, Jami Ramsey, Mary Frances Curry, Nishay Holloman, Crystal  Barket, Michelle Spear, Shelley Mahan, Taeleigha Greene, Zachary Eardley, Gen Moussa, Mary Ann Gottschlich | New Horizons Clinical Research | Cincinnati, OH |
| Sinikka L. Green, MD | Julie Hamilton, Alex Fuller, Jeanette Dickhaus, Colleen Jacobson, Triny Cooper, Michelle Jackson, Taylor Evans, Tabitha Judd, Kathryn Alexander, Megan Rosallo, Sikhongi Phungwayo, Robin Dotson, Dana Finley, Michael Vasquez, Cyndi Foster, Gregg Lucksinger, Sarah Smiley, Jayasree Krishnankutty, Ray Coon, Grishma  Dhimmer, Melanie Wilkerson, Tatum Shawver, Marcedes Coffman, Devin Teal, Laura Crenshaw | Advanced Clinical Research | Cedar Park, TX |
| Carl P. Griffin, MD | William Schnitz, Andrea Romero, Kim Hamilton, Raymond Cornelison, Angela Genovese, Shelly Brunson, April Green, Lacey Dietz, Kim Calloway, Chris Hyatt, Destiny Heinzig-Cartwright, Chalimar Rojo, Sharee Wright, Kathi Shaw, Michael  Pojezny, Avery Keller, Krystal Hightower, Dalia Tovar, Shanda Gower | Lynn Health Science Institute | Oklahoma City, OK |
| Milton Haber, MD | Maria Candelario, Martha Bunnell-Pollak, Lauren Wade, Jackie Ziegler, Deena  Ramirez, Perla Avalos, Maria Drada, Jasmine Ali, Jessica McDowell, Kehinde Busari, Patricia Church, Ronald Meza, Marco Vela, Esteban Zurita, Chris Connolly, Ruben | Laguna Clinical  Research Associates | Laredo, TX |

| **Principal Investigator** | **Study Team** | **Institution** | **Location** |
| --- | --- | --- | --- |
|  | Del Bosque, Alisha Lutat, Chelsea Fleming, Brett Potthoff, Anita Suri, Cynthia Priester, Brenda Hernandez, Veronica Procasky, Eva Cerreta, Matt Honold, Melinda  Rodriguez, Maria Regalado, Jordan Stauffer |  |  |
| Greg Hachigian, MD | Michael Cancilla, Ricardo Castellanos, Angela Cuellar, Yaman Darmarathne, Shaila Faulker, Yana Gordeyeva, Michelle Hisey, Ashley Jungsten, Kristin Kiersey, Pawandeep Nagra, Nav Nagra-Kooner, Jazmin Nauta, Masaru Oshita, Kenneth Quick, Julie Raygoza, Amanny Sadek, Melisa Tinder, Jhoana Torres, Deborah  Murray, Kristen Kiersey | Benchmark Research | Sacramento, CA |
| Laurie J. Han- Conrad, MD | Brandon Baldwin, Lucian Cappoli, Tenisha Garcia, Ella Grach, Brenda Grande, Nicolle Mendez, Natalie Moy, Matthew Musikanth, Karen Mylerberg, Brennan Opanasenko, Mark Pulera, Patti Sanchez-Emery, Mireles Sarah, Todd Simmons, Denis Tarakjian | WR-Medical Center for Clinical Research | San Diego, CA |
| Wayne Lee Harper, MD | Toni Bland, Lori Bridges, Lucian Cappoli, Lisa Cohen, Leah Corts, Annie Craft, James Earnhardt, Lynn Eckert, Aubrey Farray, Laura Hoer, Matthew Hong, Chris Hoyle, Jenee Jiggetts, Brian Joseph, Bradley Killebrew, Kendra Lisec, Lucie Mangala, David Musante, Adnan Nasir, Amanda Olsen, Brennan Opanasenko, Marci Parks, Marion Peoples, Katherine Schuch, Judith Shand, Sabine Ucik, Douglas Wadeson, Barbara Wheeler | M3 Wake Research | Raleigh, NC |
| Ripley R. Hollister, MD | Jeremy Brown, Brandy Ball, Jeremy Brown, Valerie Dyster, Dalia Jeronimo, Shelby Pickle, Michael Pojezny, Melody Ronk, Kathi Shaw, Bobbi Shofner, Jami Wagner, Meghan York, Jill York | Lynn Institute of the Rockies | Colorado Springs, CO |
| Lisa A. Jackson, MD, MPH | Marilyn Nguyen, Maya Dunstan, Barbara Carste, Sarah Friend, Diana McFeters, Lynn Gross, Mohamed Ajenah, Jana Ffitch, Audra Mccoy, David Skatula, Susan Lasicka, Kimberly Brinker, Karen Sherwin, Melissa Scheer, Paula Lins, Roger Calvert, Roxanne Erolin, Stella Lee, Vi Tran, Stephanie Pimienta, Bruce Douglas, Lee Barr, Colin Fields, Erika Kiniry, Joe Choe, Janice Suyehira, Joyce Benoit, Michael Witte,  Rebecca Lau | Kaiser Permanente Washington Health Research Institute | Seattle, WA |
| Spyros Andrews Kalams, MD | Greg Wilson, Kyle Rybczyk, Katie Crumbo, Carly Griffin, Latoya Hannah, Amy Kerrigan, Valerie Mitchell, Jenna Caserta, Mary Downey, Nicole Swindle, Shonda Sumner, Amber Massey, Trudy Sullivan, Rita Smith, Cindy Nochowicz, Eric Olson, Christian Warren, Josh Simmons, Dana King, Gwendolyn Rees, Matt Donio, Jesse  Case, Keith Richardson, Jarissa Greenard | Vanderbilt University Medical Center | Nashville, TN |
| Colleen Kelley, MD, MPH | Valeria D. Cantos, Sheetal Kandiah, Carlos del Rio, Christina Bacher, Hannah Huston, Juliet Brown, Divya Bhamidipati, Nithin Gopalsamy, Brittany Lynn Speigel, Elizabeth (Betsy) Hall, Brandon Spratt, Kiran Dhillon, Caitlin Moran, Michael Chung, Felecia Wright, Marcia Peters, Rondell Jaggers, Vanessa Soliman, Ron Gaston, Christopher Foster, Sarah Wiatrek, Bezuayehu Mandefro, Pamela Weizel, Pamela Lankford-Turner, Anandi Sheth, John Gharbin, Catherine Abrams, Philip Powers, Paulina Rebolledo, Christin Root, Tiraje Lester, Sha Yi, Damien Swearing, Fred Ede, Isaac Perez, Kelly Likos, Meen Dhir, Aastha KC, Gabriela Georgial, Tucker Colvin,  Nabeel Yar Khan, Valarie Hunter, D’Jamel Young, Felecia Atkinson | Emory University Emory University – Ponce de Leon Clinical Research Site | Atlanta, GA |
| Christina Kennelly, MD | Jacob Coleman, Brittany Bundeff, Melissa C. Hennessey, Kenneth Owen, Caroline Wilds Wilds, Jennifer Womack, Susan Martello, Chiedza Hooker, Robert Brownlee, Melissa James, Deborah Wesley-Farrington, Lori Whiteheart, Hala Webster, David Framm, Cortney Fretz, Gwyn Gibson, Susan Donahue, Kelly Woodell, Linda McCarty,  Jim Vesely, Scott Chatterton, Andrew Ottesen, Enrico Belgrave, Krishna Shah, James Chester Alexander, Brittain Callahan | Javara | Charlotte, NC |
| Shishir Kumar Khetan, MD | Taja Adams, Tanya Alexander, Tanya Alexaner, Sydney Barmoy, Jake Bart, Kira Bell, Ira Berger, Jemario Blackwell, Priscilla Buahin, Bounphone Chanthavong, Juliana DeVito, Azure Erskine, Brandon Essink, Laura Falcone, Debra Gabrielson, Beau Garland, Barb Geiger, Tiana Oliver, Courtney Heisey, Sucharita Katikala, Andrew Kimball, Heather Lang, Jeanette Lee, Asefa Mekonnen, Devan Myers, Kimberly Nieves, Allison O'Brien, Oyebisi Olanrewaju, Nicole Osborn, Adunola Oshiyoye, Rahul Patel, Alan Pollack, April Poole, Collin Smith, Kathryn Stoddard, Chao Wang, Sean  Whelan, Jonathan Whelan, Graciela Zapata, Nan Zhai | Meridian Clinical Researc | Rockville, MD |
| Murray A. Kimmel, DO | Alexa Diec, Ann Riley, Bette Denmat, Bram Swarr, Christina Raidl, Dania Billman, Denise Dixon, Donald Dawson, Elaine Crudo, James Crowley, Katrina Carlson, Kaylie Worzick, Laura Worth, Lisbeth Gordon, Marion Oliver, Robert Holt, Simmy Pinto, Taylor Atkinson, Traci Mitchell, Lana Ghomrawi, Norma Rokoff | Optimal Research | Melbourne, FL |
| Judith L. Kirstein, MD | Jared Bradshaw, Krista Forster, Jeanette Dickhaus, Marcia Bernard, Erica Sanchez, Nikki Abels, Cynthia Kunakom, Vanessa Vandergoot, Jessica Fisher, Carol Remigio, Jourdan Manfred, Frederick Lloyd, Tiffany Williams, Clarisse Baudelaire, Lovette  Cherelle, Nolan Mackey, Alan Valenzuela, Theodore Wyman, Alyssa Taber, Karen Myers, Craig Koch | Rancho Paseo Medical Group | Banning, CA |
| Michael J. Koren, MD | Shannon Trull, Amanda Elwood, Mary Strickland, Ivy Gulliermo, Chistopher Ganzhorn, Sonia Gerardo, Taylor Johnson, Victoria Kaposchansky, Cassie Lawler, Laura Little, Amanda Pratt, Sheldon Warren, Andrea West, Emery Noles, Nathanial | Jacksonville Center for Clinical Research | Jacksonville, FL |

| **Principal Investigator** | **Study Team** | **Institution** | **Location** |
| --- | --- | --- | --- |
|  | Grant, Jillian Agnew, Lori Alexander, Brenda Anderson, Deirdre Arrington, Sara Benner, Lisa Carl, Allison Crain, Nafisa Ishaku, Robert Nix, Sharon Smith, Amber Devries, Sandy Salceiro, Opara Chukwudi, Mikaela Karney-Trull, Ramil Castillo, David Graham, Gail Lowe, Alexander Hill, Carolyn Tran, Jeffry Jacqmein, Darlene Bartilucci, Alpa Patel, Janet Garvey, Mitchell Rothstein, Kenneth Aung-Din, Margaret Gannaway, Arman Mughal, Sandra Fuit, Jolenne Wolfer, Erin Schelhorn, Jacob  Wolfer, Madison Martinez, Melissa Parks, Patricia Neal |  |  |
| Karen L. Kotloff, MD | Matthew Laurens, Milagritos Tapia, Lisa Chrisley, Cheryl Young, Barbara Albert, Robin Barnes, Shernel Barrett, Andrea Berry, Melissa Billington, Shannon Bittner, Colleen Boyce, Faith Pa'Ahana Brown, James Campbell, Regina Carpenter, Jamonie Carter, Ginny Cummings, Brenda Dorsey, Jorge Flores, DeAnna Friedman-Klabanoff, Shirley George, Nancy Greenberg, Hassan Haji, Elizabeth Hammershaimb, Susan Holian, Leslie Howe, Myounghee Lee, Alyson Kwon, Kirsten Lyke, Alma Valle Maldonado, Jennifer Marron, Kaitlin Mason, Monica McArthur, Rosa McBryde, Sherry McCammon, Sandra Molina, Kathleen Neuzil, Daniele Nitkowski , Justin Ortiz,  Rekha Rapaka, Mardi Reymann, Toni Robinson, Wanda Somrajit, Mark Travassos | University of Maryland, School of Medicine | Baltimore, MD |
| Mark E. Kutner, MD | Amanda Colina, Isett Caro, Frances Beltran, Jessie Downs, Jonathan Fernandez, Mariete Renden, Mirnaya Mujica-Alabaci, Susel Figueredo, Yanelis Dominguez, Jaime Blandon, Bryan Ruiz, Leidy Montoya, Edgardo Rodriguez, Jessie Downs, Jason Rothscheld, Janett Acle, Yaime Martinez, Soraya Ricardo, Maria Hernandez Moran, Eloisa Guerra, Heidie Perez, Claudia Rodriguez, Victoria Moreno, Vanessa Hechavarria, Saray Carvajal, Daniel Lopez, Carlos Iviricu, Neiner Enriquez, Paola Garcia, Chris Hoyle, Marianela Carvajal, Janet Mendez, Edisleidy Mesa, Marco Ramirez, Dalila Del Valle, Jennifer Ortega, Yeni Hernandez, Jhobana Vargas, Carmen Amador, Juan Delgado, Maury Santos, Meredith Arguelles, Leyanis Coello, Vanessa Ansorena, Jorge Caso, Stacy Machado, Raydel Valdes, Giann Lightbourn,  Dayami Dovales, Alain Chang | Suncoast Research Group | Miami, FL |
| Mimi Van Der Leden, MD, PhD | Chrishea Harvey, Tricia Oyeyemi, Aicha Moutanni, Stephanie Melton, Peta-Gay Jackson Booth, Jennifer Yoon, Gloria Kim, Atanas Filev, Francis Uwandi, Meyling Lopez, Janice Spreitzer, Courtney Gennes, Xiangfei Cheng, Matthew Van Sickle, Nick  Bart, Brianne Okunji, Frank Maloba | Optimal Research | Rockville, MD |
| Michael L. Levin, MD | Brennan Opanasenko, Yajaira Ramos, Shonda Lester, Rebecca Boucher, Shawn Harrell, Shon Boucher, Patti Sanchez, Nina Scharbach, Alex Sanchez, Shyane Raniello, Wendy Guerra, Krystal Tyner, Kimberly Temple, Ruby Ortiz, Daniel Terreault, Amy Kill, Jade Odynski, Adolfo DeLeon, Debbie Carter, Eduardo Rodriguez, Julia Gass, Sara Esparza, Sierra Dansbee, Tammy Harrison, Marcy Kulic,  Lucian Cappoli, Mora KIm, Matthew Fenner, Heather Jimenez, Shraddha Dubal, Julie Hussey | WR-Clinical Research Center of Nevada | Las Vegas, NV |
| Michael Lewis, MD | Nancy Mohler, Mai Pham, Ron Waldorf, Elham Ghadishah, Samantha Feril, Stella Lee, Dzuyen Nguyen, Ruoxiang Wang, Justine Velandria, Benjamin Dreskin, Joseph Yusin, Lauren Vigil, Sara Wong, Suchi Tiwari, Joseph Pisegna, Sunita Dergalust,  Wayman Lee, Krissa Caroff | VA Greater Los Angeles Healthcare System | Los Angeles, CA |
| Gregg H. Lucksinger, MD | Jaleh Ostovar, Craig Koch, Danuel Hamlin, Kelly Chase, Jeanette Dickhaus, Edward Kerwin, Frederick Forde, Allison Alvord, Dawn Stewart, Dan Hamlin, Kevin Parks, Ryan Israelsen, Kary Kelly, Tiffany Smith, Melissa Myers, Ryan Rackley, Audrey Kuehl, Savannah Peterson, Hannah Hall, Jay Weisbart, Alison Dodenhoff, Emily Kelly | Velocity Clinical Research | Medford, OR |
| Mary Beth Manning, MD | Carol Salango, Alec Ireland, Lisa Hoagland, Jeanette Dickhaus, Toby Briskin, Joan Rothenberg, Michael Gaston, Sharita Tedder-Edwards, Denise Roadman, Megan Sokolowski, Tina Shickluna, Katherine Bielanski, Samantha Hood, Talia Chandler, Brianna Arman, Melinda DeLong, Naqib Ahmad, Karly Tarase, Jade Svoboda, Lisle Merriman, Melisa Sebera, Emma Landskroner, Amy Maroun, Brooke Glivar, Jennifer Gaston, Sarah Dzigiel, Cassiandra Uminski, Karol Sabol, Devan Patel, Nick Zarbo,  Briana Jackson, Brian Sharpe, Nicole, Baitt, Kaitlyn Duffy, Gabrielle Jacobs, Ann Czuprun, Tracee Cash, Diamond Ivey, Kaitlyn RubelI | Rapid Medical Research | Cleveland, OH |
| Kristen Marks, MD | Grant Ellsworth, Tina Wang, Timothy Wilkin, Mary Vogler, Carrie Johnston, Marshall Glesby, Roy Gulick, Ole Vielemeyer, Rebecca Fry, Todd Stroberg, Caitlin Rhoades, Noah Goss, Shaun Barcavage, Valery Hughes, Jonathan Berardi, Caroline Greene, Sarah Galloway, Caique Mello, Ashley Machado, Mia Crowley, Monique Williams, Katherine Fee, Elizabeth DeJesus, Andrew Yu, Minkyung Lee, Susan Herder, Mary Ann Zweibel, Patrice Weller, Antonio Rivera-Lopez, Edward Kenny, Hetal May, Natella Fridman, Parul Shah, Ruby Lee, Venus Fernandez, Victoria Lesina, Celine Arar, Byron Bullough, Kinge-Ann Marcelin, Brian Mangano, Jessenia Fuentes, Jiamin Li, Genessi Rodriguez, Catherine Jerry, Nadi Islam, Liqun Cai, Wayne Burns, Akinbayo Caulcrick, Andrika Thomas, Barbara Batog, Guoan He, Sara Yoder, Tamara  Crowder, Gianna Resso, Sophia Alvarez, Tahera Begum, Elizabeth Connolly, Roxanne Rosario, Paul Kim, Steven Wang, Vasilika Koci | Cornell Clinical Trials Units - Weill Cornell Chelsea and Uptown | New York, NY |

| **Principal Investigator** | **Study Team** | **Institution** | **Location** |
| --- | --- | --- | --- |
| Judith Martin, MD | Alejandro Hoberman, Timothy Shope, Gysella Muniz, Sonika Bhatnagar, Kumaravel Rajakumar, Anne-Marie Rick, Peri Unligil, Jennifer Nagg, Melissa Andrasko, Mary Ann Sieber, Jennifer Opal, Lalicia Roman, Spenser Kinsey, Michelle Burke, Matthew Lee, Dominic Kramer, Linette Milkovich, Emily Dougherty, Emily Carney, Shannon  Mance, Nader Shaikh, Diana Kearney, Jamie Fries, Lisa Vavro, Shayla Goller | UPMC University Center | Pittsburgh, PA |
| John W. McGettigan, Jr., MD | Walter Patton, Jennifer Schnider, Riemeka Brakema, Heeten Desai, Mikell Brett Karsten, Patricia Jalomo, Cindy Finch Benoy, Karin Choquette, Jonlyn McGettigan, Yvonne De Los Reyes, Melissa Cozzens, Amanda Hermosillo, Cindy Montgomery, Susan Tarwid, Annette Elzy, Tianna Young, Saysamone Banks, Cristina Fernandez, Damaris Atondo, Zoe Sesma, Norma Barrientos, Maggie Tono, Kisha Adams, JoAnn  Wilkins, Arianna Bermudez, Carol Sayer, Julie McDowell, Angelina Navarro, Mercedes Sullivan, Crystal Mata, Sheldon Gingrich, Aaliyah Sestiaga, Gia Longo | Quality of Life Medical & Research Centers | Tucson, AZ |
| Mark Montgomery McKenzie, MD | Tiffany Jewell, Zackery Harmon, Michael Elizabeth, Christy Sweet, Teresa Deese, Catherine Schon, Misti Earwood, Lou Cappoli, Brennan Opanasenko, Lisa Guider, Michelle Forgey, Justian Jarrett, Rachel Scott, Elizabeth Michael, Erica Osmundsen, Andrew Wood, Shelly Brooks, Gisela Heintz, Lilian Nukuna | WR-ClinSearch | Chattanooga, TN |
| Vicki E. Miller, MD | Sajjad Naqvi, Soofia Masood, Fredric Santiago, Sonia Guerrero, Subhash Koneru, Nirja Shah, Andrea Torres, Ramani Gali, Talha Baig, Heather Leary, Afifah Ayub, Nayab Goher, Patti Tate, Reagen Reed, Muhammad Irfan, Amy Starr, Alefiyah Motiwala, Julia Kenny, Victoria Aguilar, Jessica Arguijo, Insiya Valika, Victoria Aguilar, Jagruti Patel, Anna Pena, Faryal Mahmood, Blanca Gomez, Nancy Torres, Kristyn Latil, Tarori Mark, Laura Djampou, Lindsey Kueng, Marianne Tadros, Mohammad Millwala, Monica Murray, Murtaza Marvi, Shivani Shah, Vanessa Gonzalez, Zohair Harianawala, Zainab Rizvi, Ambily Dileep, Jaquelyn Gonzales, Ragen Powell, Carolina Deandres, Syed Fahad Ali Kazmi, Sandra Natalia Perez, Shannon Amacker,  Shiela Varghese | DM Clinical Research | Tomball, TX |
| Gowdhami Mohan, MD | Rodolfo Barrera, Emma Partin, Kelly White, Ashley Rochester, Charles Thompson, Stefanie Tyson, Ashten Sheriff, Alyssa-Kay Peay, Kayla Corn, Barbara A. Richardson, Kristin Miller, Steven Clemons, Cameron King, Emma Partin, Gary Clemons, Brianna  Starr, Danyel Johnson, Taylor Davis, Niki Tyson | VitaLink Research | Anderson, SC |
| Kathleen M.  Mullane, DO, PharmD | David L. Pitrak, Cheryl Nuss, Karen Cornelius, Randee Estes, Amy Luckett, Michelle Moore, Judi Pi, Stephen Schrantz, Jill Stetkevych | University of Chicago | Chicago, IL |
| Joseph Lee Newberg, MD | Mary Reyes, Nicole Leahy, Victoria Andriulis, Herbert Whinna, Patricia James, Lana Ghomrawi, Carole Kempfer, Miriam Arroyo, Maria Castro, Anna Maddox, Reuben Martinez, Jacquilyn McCormick-Burks, Laura Pearlman, Rosalinda Vazquez, Shaheera Suleiman, Neha Atal, Rosalind Vazquez | Synexus Clinical Research | Chicago, IL |
| Richard M. Novak, MD | Regina Harden, Maria Schwarber, Michael Pacini, Rebeca Gansari, Margie Villarreal, Stephanie Martin, Michelle Lee, Richard Morrissy, , Taylor Ellis, Samuel Rene, Tara Cobbs, Claudia Preciado, Scott Borgetti, Maximo Brito, Olamide Jarrett, Mahesh Patel, Tracy Cable, Charity Ball, Maryann Holtcamp, Rodrigo Burgos, Sarah Michienzi, Emily Drwiega, Mikayla Johnson, Fischer Herald, Benjamin Ladner, Minseung Chu, Carolyn Dickens, Alfredo Mena Lora, Stockton Mayer, Andrea Wendrow, Habiba Sultana, Nanu Nunwar, David Chan, Marla Schwarber, Khandaker Anwar, Mahmood Ghassemi, Md Ruhul Amin, Doris Carroll, Rosa Valencia, Michelle Agnoli, Elena Llinas, Samuel Rene, Liam Morrissy, Adrian Raygoza, Addis Mekkonnen, Lisa Lindemann, Daniel Meslar, Karen Pacini, Corey Ringhisen, Amy Kennedy-Krage, Claudia Miller, Lorna Sanchez McCann, Gizelle Alvarez, Nia Moragne-Oneal, Nusirat Williams, Ian Feather, Nikki Griffith, Wardrick Nealon, Renyce Powell, Nila Safaeian, Monica Gingell, Diana Bahena, Gerald Beck, Brad Farrington, Rod Reyes, Monica Wilson, Juline Wondrasek, Kimberly Shapiro, Shannon Whitted, Victoria Roehl, Braulio Carrasco, Michael Chen, Olivia Murray,  Yasiel Lacalle, Tessa Eckley, Anna Schluckebier, Kevin Cao, Elise DeBruyn | University of Illinois at Chicago - Project WISH | Chicago, IL |
| Paul Joseph Nugent, DO | Leonard Singer, Jennifer Jones, April Smith, Georgettea Geuss, Lana Ghomrawi, Christine Bennett, Norma Blevins, Linda Brotherton, Michele Byrd, Krista Doss,  Victoria Holden, Christine Hull, Jean Montgomery, Nancy Cipollone, Savanah Torline, Brandon Brown, Meagan Thomas, Katie Ziska, Dana Sias, Hannah Wagner | Synexus Clinical Research | Cincinnati, OH |
| Jeffrey Scott Overcash, MD | Hanh Chu, Kia Lee, Karla Zepeda, John Rodriguez, Adam Prince, Yashveer Dubbula, Elizabeth Tomatsu Michael Voskanian, Crystle Rajania, Stephanie Ramirez, Claudia Camacho, Lauren Arnett, Kecia Darbeau, Ashley Smith, Kimberly Quillin, Cesar Ramirez, Daniel Robitaille, Erica Sanchez, Allie Davis, Michael Waters, Pat Kappen, Valerie Horne, Thao Vuong, Andrew Dennis, Nikki Abels, Dominique Panis, Richard McQuaid, Whitley Harbison, Erika Trujillo, Andrea Garcia, Jose Jacob Esparza, Carlos Vera, Raquel Taitingfong, Cathy Meza, He Pu, Jackielynn Smith, Shandel Odom, Zahira Nieves, Ashliegh Lindsay, Ariana Nasatka, Jose Cazarez, Nora  Martinez, Angela Hunt, Antonio Delgado, Linda Vega, Angela Anorve, Erica Martinelli, Melania Riordan, Sylvia Lindholm, Gina Ciezkowski, Grecia Perez, Jacob Pineda, | Velocity Clinical Research, San Diego | La Mesa, CA |

| **Principal Investigator** | **Study Team** | **Institution** | **Location** |
| --- | --- | --- | --- |
|  | Nathan Tyler, Ranya Salem, Amara Yilmaz, Jessica Gonzales, Zabrina Ruiz, Laura Castillo, Yajaira Contreras, Angelica Guzman, Makenna Orel, Jeffery Alvarez, Gordon  Bovee, Roxana Ramirez, Joan Esquivel |  |  |
| James Todd Peterson, MD | Christopher Mickelson, Madeline Maldonado, Alison Charlton, Ashley Bragg, Sean Hansen, Emily Wilcox, Colby Bostock, Megan Henry, Pam Iwasaki, Bradley Young, Katelyn Walker, Joy Nguyen, Lindsey Bevan, Megan Grimmett, Madeline Grote, Heather Littell, Natalie Bee, Alexander Clark, Shana Eborn, Susan Edwards, Dan Henry, Heather Jackson, Gerald Kelty, Issac Pena-Renteria, Jacqueline Rohrer, Jack  Taylor, Brooke Barrick, Ty Henry, Anna Dansie, Kenadie Hamblin | J. Lewis Research | Salt Lake City, UT |
| Paul Pickrell, MD | Susan Bonner, Blaire Graham, Staci Taggart, Hussain Malbari, Tiffany Lemuz, Ethan Shotton, Andrew Bell, Megan Malek, David Pampe, Carol Ann Linebarger, Michelle Peterson, Brandi Chalman, John Luna, Elizabeth Santellanes, Christina Martinez, Lisa Johnson, Lisa Savage, Melissa Winn, Wendi McKenzie, Eileen Euperio, Stefanie Mott, Paul Menefee, Katie Caballero, Darrell O'Brien, Morgan Schulle, Kate Jurek, Olivia Hapanowicz | Tekton Research, Inc. | Austin, TX |
| Terry L. Poling, MD | Meenakshi (Kavya) Natesan, Patricia Contreras, Denise Hole, Avi Woods, Jill Hiebert, Melissa Burton, Olivia Eagleson, Laura Holz, Terri Ford, Cindy Thome, Terry D Klein, Gregory Greer, Diandra Henriques, Tracy R Klein, Thomas C Klein, Christa Shue,  Gina Young, Brenna Sprout | Alliance for Multispecialty Research | Wichita, KS |
| Bruce G. Rankin, DO | Jennifer Dittman, Lora Parahovnik, Crystal Paccione, Melissa Hodges, Katina Marchione, Matt Maxwell, Any Dominy, Diana Toney, Andrea Marrafino, Laura Isbell, Leandro Fernandez, Claxton Copeland, Michelle Tutt, Adam VanDeusen, Kevin Feldman, Clark Mason, Tifany Huertas, Over Seijas, Jennifer Cline, Christian Beierschmitt, Ryan Hobbick, Jessica Gilliam, Jeanette de Leon, Iman Mencia, Daniel Layish, Vienna Bauer, Shatonia Fields, Albert Garcia, Carrie Rycort, Tasha Brocato, Marshall Nash, Samantha Watts, Amy Houck-Dominy, Angela Hammerle, Teresa Logsdon, Erika Wierzbicki, Taylor Martin, Ranie Hutchins, Fadhel Alyunis, Gail Lavine, Jeffery Hood, Robert Duran, Michelle Jones, Ginny McClanahan, Heather Jackson, Leandra Fernandez, Douglas Winter, Antonio Rivera, Amber Vasquez, Thais Truffa, Daniel Campbell, Grace Newcomb, Elizabeth Orlando, Steven Shinn, John  Hill, Christina Isbell, Dhaneshwar Oomrow, Alicia Cevera | Accel Research Sites | DeLand, FL |
| Michele Diane Reynolds, MD | Jennifer Bashour, Robert Schmidt, Cynthia Mayeux, Uvoka Huffman, Lisa Nicholson, Jacklyn Newton, Lynn Yauch, Cathy Monroe, Kathleen Carty, Angelica Banks, Taylor Werner, Pamela Echols, Pauline Jackson, Chana Hines, Lorine Cook, Cristina Puig,  Patrick Brooks, Jennifer Ruiz, Deanna Bowman, Ladina Garcia | Synexus Clinical Research | Dallas, TX |
| Rambod Rouhbakhsh, MD, MBA | John Johnston, Richard Calderone, Tasha Stevenson, Tameka Fortune, Brandi Pace, Adreanna Pou, Jerrica Sullivan, Yolanda Lewis, April Rouse, Tiffany Jefferson, Elizabeth Danford, Jeff Repper, Mason Boutwell, Alexycia Washington, Krista Hirth,  Meagan Grabel | MediSync Clinical Research Hattiesburg Clinic | Petal, MS |
| Nadine Rouphael, MD | Renata Dennis, Tigisty Girmay, Michelle Wiles, Sharon Curate-Ingram, Lauren Hewitt, Alexis Ahonen, Mari Hart, Sarah Bechnak, Erin Carter, Lauren Nolan, Daniel Sans Graciaa, Geoffrey Kamau, Easton Beshears, Sy Tran, Mary Atha, Mary Bower, Ghina Alaaedine, Brandy Johnson, Jacob Usher, Eileen Osinski, Erin Scherer, C. Tae Stallworth, Stephanie Ramer, Rose Pope, Esther Park, Francine Dyer, Laura Clegg, Rebecca Gonzalez, Stacey Wheeler, Susan Rogers, Vy Ngo, Vanessa Soliman, Kristen Unterberger, Bernadine Panganiban, Christopher Huerta, Juton Winston, Ali Alvarez, Jianguo Xu, Colleen Kelley, Paulina Rebolledo, Nicholas Scanlon, Jessica Traenkner, Matthew Collins, Hollie Macenczak, Cassie Grimsely-Ackerley, Tiffany Lee, Amy Anderson, Michele Paine McCullough, Hannah Huston, Daniella Carter, Lisa Harewood, Srilatha Edupuganti, Varun Phadke, Mindee Adamson, Jeanne Allen, Debbie Bartenfeld, Lily Berz, Amy Cromwell, Sergio Cruz, Fred Ede, Monica Godfrey, Evan Gutter, Angelle Ijeoma, Sara Jo Johnson, Vinit Karmali, Dean Kleinhenz, Jennifer Kleinhenz, Alexandra Koumanelis, Maranda Leary, Tiraje Lester, Juliet Alise Morales, Shashi Nagar, Julia Paine, Dilshad Rafi Ahmed, Brittany Robinson, Amanda Rosner, Renee Silver, Trevor William Simon, Talib Sirajud-Deen, Damien Swearing,  Maliya Tolbert, Pamela Turner, Chia Uzuegbunam, Claire Wan, Dongli Wang, Erika Wimberly, Jean Winter, Joy Winters, Yong Xu, Sha Yi | Emory University - Hope Clinic | Decatur, GA |
| Richard Rupp, MD | Amber Stanford, Megan Berman, Laura Porterfield, Gerianne Casey, Hala Ghoson, Doreen Jones, Michael Willig, Cori Burkett, Robert Cox, Amy McMahan, Diane Barrett, Kristin Pollock | University of Texas Medical Branch | Galveston, TX |
| Jamshid Saleh, MD | Matthew Miles, Rafael Lupercio, Vicky Martin, Marla Clark, Matthew Pohlmeyer, Ruba Zanaid, Veronica Blevins, Tara Ulberg, Carlyee Chambers, Marisol Corrales, Emily  Crews, Mohamed Yassin, Sarah Sandberg, Frank Chen, Mandy Swanson | Paradigm Clinical Research Center | Redding, CA |
| John W. Sanders, MD, MPH | Stacy Harpe-Hall, Jesse Hopkins, Ann Schweppe, Jaymous Fayssoux, Kathryn Bender, James Peacock, Katharine Pearsall, Brandy Snyder, Deidre Knox, Megan  Thorpe, Melissa Ellingson, Brittany Bundeff, Lisa Ashworth, Meredith Hiatt, Ritu Rathee, Stacy Woodliff, Brian Strittmatter, Amanda Wright, Daisy DeWeese-Gatt, | Wake Forest University Health Sciences | Winston Salem, NC |

| **Principal Investigator** | **Study Team** | **Institution** | **Location** |
| --- | --- | --- | --- |
|  | Caryn Morse, John Williamson, Samantha Wheeler, Lori Whiteheart, Susan Donahue, James Lovette, Kaitlyn Van Leuvan, Kelly Ledbetter, Scott Chatterton, Julio Nasim, Amie Sidberry, Ashley Davis, Carter Noecker, Chie Hooker, Johanna Breenan, Sam  Cable, Anna Bowman, Stephanie Boothe, Shea Overcash |  |  |
| Howard I. Schwartz, MD | Carlos Valladares, Jocelyn Morrera, Yulexis Amestoy, Tori Wallenburg, Thelma Beltran, Terry Piedra, Monica Garces, Alexandra Galvis, Wanda Delgado, Catherine Casas, Lesly Miguel Sosa, Vivian Rosales, Jose Fernando Henriquez, Mikael Yaniz, Beatriz Rivera, Peter Ventre, Gabriella Huyke, Maria Companioni, Jessie De Vega, Brianna Gamez, Stephanie Diaz, James Jean-Mary, Americo Padilla, Nikita Notise, Yorlina Luquetta, Monifa Wilson-Morris, Kenia Gutierrez, Roilan Garcia, Karla Pentzke, Leyda Valentin, Lazara Novas, Marilein Camacho, Jazmin Henfield, Laymis Alvarez, Myriam Rosado, Maxine Bryant, Maria Pinero, Laura Raucci, Francisco Ramirez, Angelic Gamez, Mailin Perez, Yasmin Baddour, Hary Leon Joseph, Yaquelin De la Cruz, Dunia Torres, Rosaidaliz Carreira, Chanella Garcia, Surisaday Mederos, Jose Muniz, Karenda Plotka, Sara Gomez, Maria Soto, Cathy Cruz, Nelia Sanchez-Crespo, Jennifer Schwartz, Barbara Corral, Matthew Muniz, Dayana Deltejo,  Ana Castro, Reem Hassan | Research Centers of America | Hollywood, FL |
| Nathan Segall, MD | Michelle Sowell, Nancy Levine, Erynn McKinley, Hannah Smith, Karen Hickson, Elizabet West, Patrizia Greene, Jon Finley, Mildred Stull, Susan Jones, Jennifer LeBrun, Pamela Talbott, Kwannda Whatley, Jeffrey Jones, Michelle Binns, Donna  Toepfer, Cynthia Steele, Grace Newville, Gillian Waite, Cynthia Pinckney, Karen Yangapatty, Kiara Tyner, Kimberly Cobb, Kourtney Richardson | Clinical Research Atlanta | Stockbridge, GA |
| William Seger, MD | Kimberly Pullen, Jean Seignon, Anthony Kim, Mohammed Antwi, Allison Green, Lizzy Seger, Elizabeth Boydston, Abdur Rafay Qadri, Deborah Devlin, Tasha Todd, Oluwatosin Akingbala, Alma Guel, Tisha Davis, Melody Dufrene, Samantha Loudermilk, Virginia Loudermilk, Crystal Starr, John Villegas, Ben Seger, Katherine Hollie | Benchmark Research | Fort Worth, TX |
| Neil Parmanand Sheth, MD | Kenneth Stell, David Beckett, Enitt Gonzalez, Donna McGunigal, Amanda Burns, Nancy Wood, Shelley Miceli, Christina Avila, Rebecca Baker, Laura Vigliotti, Sarah  Kading, Samer Salama | Synexus Clinical Research | Glendale, AZ |
| William B. Smith, MD | Richard L Gibson, Jennifer Winbigler, Elizabeth Parker, Madison Watts, Suzann Cloninger, Talya Thomas | Alliance for  Multispecialty Research | Knoxville,TN |
| Joel Solis, MD | Martha Carmen Medina, Xavier Morales, Hank Heller, Blake Torrence, Joanna Gurrola-Mahoney, Cynthia Bueno, Heather Holloway, Irving Salinas, Joel Perez, Paola Garcia, Erica Canales, Blanca Urbina, Brancisilio Gutierrez, Carolina Cantu, Chelsea Vargas, Cindy Vasquez, Cody McIntire, Gabriela Gutierrez, Hugo Sosa, Irvin Munoz, Jessica Estrada, Jonna Lopez, Kaegan Knox, Mirella Melendez, Natalia Valle, Natalie Echavarria, Nicole Litton, Amber Victor, Nancy Torrence, Madhu Shreya, Mathew Maran, Asfak Alam, Westly Keating, Tara Green, Devora Torrence, Gerardo Sedas, Shruti Konda, Prem Jangam, Mario Echavarria, Alejandro Silva, Anne McNulty, Daniel Contreras, Daniel Gomez, Edgar Garcia, Elizabeth Weber, Luis Lopez, Samuel Ramirez, Kayla Lopez, Pedro Penalo, Angel Salinas, Jaime Solis, Shannon Moyer, Aryana Ibarra, Guadalupe Gurrola, Jenna Anastasiades, Uchechi  Ehiemua, Sara Solorzano | Centex Studies, Inc. | McAllen, TX |
| Stephen A. Spector, MD | Amaran Moodley, Jill Blumenthal, Baharin Abdullah, Christina Addington, Juan Carlos Alcantar, Deyna Arellano, Bernadette Cale, Brendan Costello, Tammelita Cotlon- Pineda, Fanny Delebecque, Karen Deutsch, Aram Dimayuga, Son Do, Yasmeen Esshaki, Aileen Everhart, Cindy Ewing, Veronica Figueroa, Medardo Gaytan, Crystal Groom, Carolyn Hernandez, Heather Huitema, Benjamin Hull, Sylvia Isaac, Jaclyn Jaskowiak, Cindy Knott, Leander Lazaro, Thuan Le, Megan Loughran, Michelle Madey, Rosalva Martha-Patten, Colleen McLellan, Jeff Ledford-Mills, Asami Mimura, Patty Moraes, Jennifer Morales, Jessica Nasca, Phirum Nguyen, Marielys Padilla- Martinez, Dennis Perpetua, Mike Pizza, Shannon Ransom, Emily Rizo, Carlos Rojas, Thaine Ross, Marie Sagrado, Eugene Sato, Lisa Stangl, Ji Sun, Nancy Tang, Mina  Trivedi, Rodney Trout, Donna Voss, Lindsey Woronicz | University of California, San Diego | La Jolla, CA |
| Cynthia Becher Strout, MD | Rica Santiago, Yvonne Davis, Patty Howenstine, Alison Bondell, Jaime  Robertson, Anissa Moussa, Geronimo Feria Garzon, Sierra Bennett, Marlena Petrie | Coastal Carolina Research Center | Mount Pleasant, SC |
| Shobha Swaminathan, MD | Amesika Nyaku, Tilly Varughese, Rondalya Deshields, Michelle L DallaPiazza, Elise Lewis, Jennifer Punsal, Mario Portilla, Malithi Desilva, Christina Daliani, Susana Rivera, Aidan Ziobro, Andressa Rebellatto, Brian Murloy, Christina Ninan, Ernest Pianim, Eunice Wang, Merit Henen, Muhammad Usman, Rebecca Kim, Shiao Wang, Gener Eric Cruz, Bethany Birago, Joyell Arscott, Dina Meawad, Christie Lyn  Costanza, Francesca Escaleira, Zoraida Cruz-Barahona, Jared Khan, Valeria Cadorett, Jamir Tuten, Travis Love, Eric Asencio, Sukhwinder Singh | Rutgers New Jersey Medical School | Newark, NJ |

| **Principal Investigator** | **Study Team** | **Institution** | **Location** |
| --- | --- | --- | --- |
| Ramy Joseph Toma, MD | Olivia Graves, Josiah Robinson, Patricia Hammonds, Lana Ghomrawi, Kara Quinnelly, Shaun O'Conor, Michael Lambe, Rachell Stewart, William Kirby, Pink Folmar, Rachel Culbreth, Heidi Leblanc, Julie McDaniel, Rian Montgomery, Andrea Woodle, Samantha Williams, Hunter Russell, Shereen Lowe, Maureen Mayer, Hollis  Ryan, Elaine Reese | Synexus Clinical Research | Birmingham, AL |
| Timothy P. Vachris, MD | Mark Hutchens, Stephen Daniels, Margaret Wells, Sandra Clancy, Rebecca Martinez, Jessica Buot, Merissa Daugherty, Julie Hamilton, Kimberly Hernandez, Ashli Alejandro, Amy Collins, Monique Gawlik, Patricia Johnson, Maria Moreno, Ashley  Washington, Tina Rountree, Daniel Dore, Ravi Davuluri, Ashlee Brunaugh, Jorge Martinez, James Hermon, Vianai Carreno, Mia Rountree, Colleen Coelho | Optimal Research | Austin, TX |
| Keith William Vrbicky, MD | Charles Harper, Chelsie Nutsch, Wendell Lewis III, Cathy Laflan, Linden DeBoer, Kayla Andal, Misty Appeldorn, Jenniger Grebe, Russell Herstein, Catherine King, Samantha Wieseler, Alisha Kiepke, Christy Lee, Kelsey Kelley, Kelli James, Ashley Frisch, Courney Green, Taysha Hingst, Jeni Hoppe, Kimber Breeden, Debra Gabrielson, Ginny McNew | Meridian Clinical Research | Norfolk, NE |
| Larkin T. Wadsworth III, MD | Ashley Dale, Christy Schultz, Rebecca Munsch, Anya Penly, Liz Garner, Stephanie Tesson, George Cherniawski, Angie Kean, Dan Reed, Courtney Kubiak, Maureen Dempsey, Heather Cherniawski, Breanna Galibert, Kristin Branson, Laura Hartupee, Karen Knapp, Horacio Marafioti, Lyly Dang, Jennifer Berry, Lauren Clement, Megan  Dandurand | Sundance Clinical Research | St. Louis, MO |
| Jordan L. Whatley, MD | Patricia Whatley, Christopher Dedon, Anika Payne, Amie Shannon, Kristen Losavio, Nicole Harrell, Mary Margaret Dobson, Lindsey Hall, Chaney Bennett, Crystal Rowell, Mimi Dimmick, Amy Thomassie, Kimber Breeden, Cody LaFleur, Makaylea Truitt, Taryn Collett, Emily Best, Alexandra Caillouet, | Meridian Clinical Research | Baton Rouge, LA |
| Judith White, MD | Amy Edridge, Chelsea Montalvo, Eugenia Clark, Lisa Russell, Zahra Somji, Lesli Leimer, Robert Meyer, Christine Murphy, Prity Patel, Sejal Patel, Ruben Moliere, Samantha Merveillard, Yarnick Mirjah, Bryn Walls, Joey Cruz, Aaron Cooper, Jessica Bienaime, Ashley Gilcrist, Alisa Petit, Tyler Knightly, Kimberly Stokes, Christina Rosario, Talhia Matos, Ilona Boggs, Nicholas Weber, Felix Busot, Linda Colon, Heather Gillenwater, Cristina Kaplun, Melissa Caputi, Shayna Siplin, Daminee Shah, Samuel Martin, Alexis Waldorf, Vihar Upadhyay, Adolfo Henriquez, Saskia Singh, Maria Roberts, John Caporelli, Shirley Salvador, Quevina Scarver, Vanessa Garcia, Taylor Moore, Jayasen Singh, Curshinda Galvin-Burch, Mary Kesner, Jasmin Gil, Shay Gray, Steven Monsegur, Michele Steinmetz, Michael Lambe, Heather Powell,  Sandra Torres, Shaban Katbeh, Taylor Wilson | Synexus Clinical Research | Orlando, FL |
| Priyantha N. Wijewardane, MD | Natalie Johnson, Martha Evans, Sondra Wright, Richard Pellegrino, Lastida Burns, Natasha Williams, Haylee Rowe, Kayla Graham, Amanda Horn, Eric Bravo, Jeffrey Thessing, A. Michele Maxwell, Amy Cooper, Lauren Evans, Tonya Cato, Haylee Tucker, Lesa Gann, Hannah Jones, Amanda May, Tiffany Walker, A. LeiAn Diaz, Laura Khalil, Lydia Purcell, Timothy Campbell, Charlotte Garcia-Velez, Andrea Scarborough, Beatrice A. Miller, Keith Bracy, Aujania Thompson, Cassandra Johnson,  Krishana Day, Freddie Hicks, Jamie Pettus | Baptist Health Center for Clinical Research | Little Rock, AR |
| Barton G. Williams, MD | Flo Abbott, Nicole Burton, Alice Cipollini, Madison Croucher, Philip Dattilo, Erin Harrelson, Kelsey Heston, James Ingram, William H Jones, Karla Lane, Brandy Lowman, Evan Lucas, Megan Marles, Morgan Mathis, Angie Northcott, Clyda  Pasquantonio, Alyssa Valente, Ciara Winders, Stephanie Graham | Trial Management Associates | Wilmington, NC |
| Marcus J. Zervos, MD | Paul Kilgore, Mayur Ramesh, Jelena Verkler, Pardeep Pabla, Andrew Clark, Katrina Williams, Dee Dee Wang, Beverley Duthie, Samia Arshad, Alandra White, Anna Kern, Ashley Mattern, Bilqis Mosed, Dana Parke, Doreen Dankerlui, Dragana Spasevska, Hanah Woods, Helina Misikir, Howard Klausner, Janay Scott, Jessica Heinonen, John Zervos, Joseph Miller, Kate Zenlea, Kristin Eis, Marissa Vasquez, Maurice Slaughter, Meaghan Flynn, Michael Garcia, Michelle Sankah, Nina Paeilli, Philip Benson, Robert Devore, Stevanya Baho, Tony Eljallad, Tyler Prentiss, Yaman Ahmed, Sharon Mathys, Linda Kaljee, Jeffrey Van Laere, Claudia Hanni, Hassan Zafar, Mona Desai, Gina Maki, Mary Perri, Dora Vager, Shannon Thomas, Autumn Robinson, Isis Hamilton, Sonia Eliya, Jehan Jazrawi, Biljana Popovic, Sharon Zahul, Joshua Ruzzin, John Laguio, Ali Mathena, Bobby Cook Jr., Marlene Hesler, Rochelle Fleming, Terria Minniefield, John Simons, Sherese Henderson, Ashley Hopkins, Rebecca McFarlane, Raeshell Carson, Jonathan Williams, Katherine Reyes, Erica Herc, Indira Brar, Mayur Ramesh, John McKinnon, Lacquis Duncan, Tim Asmar, Margaret Beyer, Kaleem Chaudhry, Madison Lee, Jo-Ann Rammal, Karthik Sridasyam, Siddesh Veer,  Angelique Buluran, Kimberlyn Lott, Jeremiah Rooker, Alayna Wilder, Kathleen Wilson, Allison Weinmann, Hassan Mourtada | Henry Ford Health System | Detroit, MI |

### United States Government (USG)/Coronavirus Prevention Network (CoVPN) Biostatistics Team

(PubMed listed, and ordered alphabetically by institution affiliation)

| Affiliation | Team Members |
| --- | --- |
| Biomedical Advanced Research and Development Authority (BARDA), Washington, DC | Di Lu, James Zhou |
| Department of Biostatistics and Bioinformatics, Rollins School of Public Health, Emory University | David Benkeser |
| Vaccine and Infectious Disease Division, Fred Hutchinson Cancer Research Center, Seattle, WA | Jessica Andriesen, Bhavesh Borate, Lindsay N. Carpp, Andrew Fiore-Gartland, Youyi Fong*, Peter B. Gilbert*, Ying Huang*, Yunda Huang, Ellis Hughes, Ollivier Hyrien, Holly E. Janes*, Michal Juraska, Yiwen Lu, April K. Randhawa, Brian Simpkins, Brian D. Williamson, Lars W.P. van der Laan, Chenchen Yu |
| Biostatistics Research Branch, NIAID, NIH, Bethesda, MD | Michael P. Fay, Dean Follmann, Martha Nason |
| Division of Biostatistics, School of Public Health, University of California, Berkeley, CA | Nima S. Hejazi |
| Department of Biostatistics, University of Washington, Seattle, WA | Marco Carone, Kendrick Li, Wenbo Zhang |
| Department of Statistics, University of Washington, Seattle, WA | Alex Luedtke |
| Department of Population Health Sciences, Weill Cornell Medical College, New York, New York | Iván Díaz |

*YF, PBG, YH, and HEJ are also affiliated with the Department of Biostatistics, University of Washington, Seattle, WA. PBG is also affiliated with the Public Health Sciences Division, Fred Hutchinson Cancer Research Center, Seattle, WA.

### Materials and Methods

#### COVE Trial

The COVE trial (NCT04470427), conducted in the United States, enrolled adults aged 18 and over at appreciable risk of SARS-CoV-2 infection and/or high risk of severe COVID-19 disease (*1, 2*). In the primary two-dose series, each mRNA-1273 dose was 100 μg; the third (booster) dose was 50 μg. All study participants provided written informed consent before enrollment and the protocol and consent forms were approved by the central institutional review board.

#### Omicron COVID-19 Endpoints

Correlates analyses were conducted for the first occurrence of acute symptomatic COVID-19 with virologically-confirmed SARS-CoV-2 infection, referred to as “COVID-19”. This adjudicated COVID-19 endpoint was identical to that in the primary analysis (*1, 2*) and the primary series correlates analyses (*3, 4*) of the COVE trial. For BD29 marker correlates analyses, all endpoints starting 7 days after BD29 through to April 5, 2022 (the data cut-off date of the current analysis) were counted (**Figure S1**); for correlates analyses involving the unboosted control, any remaining unboosted participants were censored on January 31, 2022.

The rationale for excluding COVID-19 endpoints between 1 and 6 days post-BD29 is that participants with these endpoints might had SARS-CoV-2 infection before BD29 and may have generated anamnestic responses that affected the BD29 antibody level. The BD29 study visit was not always 29 days post the BD1 visit because of the allowable study visit windows (within 19 and 45 days, both inclusive, of the BD1 visit; see “per-protocol” exclusions in **Figure S2**).

A non-case (or control) was defined as a participant who showed no evidence of SARS-CoV-2 infection (neither Elecsys+ nor RT-PCR+) between BD1 and April 5, 2022.

#### Definition of SARS-CoV-2 Naive or Non-Naive Status

In the per-protocol boosted cohort, a study participant was considered SARS-CoV-2 naive by BD1 if there was no evidence of SARS-CoV-2 infection (RT-PCR+, Roche Elecsys seropositive, or a symptomatic COVID-19 endpoint followed by positive confirmatory testing) from enrollment to BD1 (including the BD1 visit). A study participant was considered non-naive if the participant showed any evidence of infection between 14 days post the second vaccination in the primary mRNA-1273 two dose series and the BD1 visit (including testing seropositive at BD1; testing RT-PCR+ at BD1 were excluded from the per-protocol boosted cohort as discussed above). Based on this definition, 204 participants were classified as “non-naive” in the “per-protocol” boosted cohort and the other 14,047 participants “SARS-CoV-2 naive.”

#### Case-Control Sampling Design

The correlates study adopted a case-control sampling design stratified by the COVE trial randomization arm, a participant’s SARS-CoV-2 naive and non-naive status at BD1, four calendar periods of BD1 visits, and a person’s baseline demographics; see the “Statistical Analysis Plan for Study of Post Dose 3 and Exposure-Proximal Omicron Antibody as Immune Correlates for Omicron COVID-19 in the P301 COVE Study” (SAP), available as a supplementary file, for details. A total of 218 participants (163 SARS-CoV-2 naive and 55 non-naive) had their BD1 and BD29 antibody markers measured and met the per-protocol criteria to be included in the final analysis (**Figure S2**). Of these samples, 111 were Omicron COVID-19 cases (79 BD1 SARS-CoV-2 naive and 32 BD1 non-naive) and 107 were non-cases (84 BD1 SARS-CoV-2 naive and 23 BD1 non-naive).

Appendix A of the SAP describes how an Omicron case is approximated by adjudicated COVID-19 case (positive RT-PCR for SARS-CoV-2 with eligible symptoms) ≥ 7 days post BD29 AND ≥ December 1, 2021 given the emergence of Omicron (BA.1) wave. Primary endpoint COVID-19 cases with known Omicron BA.1 lineage were prioritized for sampling.

#### Pseudovirus Neutralizing Antibody Assay

Serum nAb activity against SARS-CoV-2 was measured in validated assays utilizing lentiviral vector pseudotyped with full-length Spike of the Ancestral (D614G) strain (hereafter referred to as “Ancestral strain”) NC_045512.2 (PPD Vaccines VAC62) and with full-length Spike of the BA.1/B.1.529 strain (hereafter referred to as “BA.1 strain”) (PPD Vaccines VAC122). The readout of each assay is serum antibody concentration Ab[C], reported in units ID50 titer [arbitrary units (AU)/ml] with labeling ID50 (AU/ml). For the Ancestral and BA.1 strains, the lower limit of quantitation (LLOQ) is 10 and 8 AU/ml, respectively; values < LLOQ were set to LLOQ/2. For the Ancestral strain, the upper limit of quantitation (ULOQ) is 281,600 AU/ml; for the BA.1 strain, the ULOQ is 24,503 AU/ml. For each strain, values > ULOQ were assigned ULOQ.

#### Binding Antibody Assay

Serum IgG bAbs against Spike antigens of the Ancestral (D614) (hereafter referred to as “Ancestral strain”), Gamma, Alpha, Beta, Delta AY4, and Omicron BA.1 strains and against the RBD antigen (Ancestral strain) were measured using a validated solid-phase electrochemiluminescence S-binding IgG immunoassay (PPD Vaccines VAC123). The readout of each assay is serum binding antibody concentration, reported in AU/ml. The assay limits are listed in Table S3. For Ancestral strain Spike and and BA.1 strain Spike the LLOQ is 69 and 102 AU/ml, respectively. For Ancestral strain RBD the LLOQ is 79 AU/ml. For each strain values < LLOQ were set to LLOQ/2. A factor to enable conversion from AU/mL to International Units (BAU/ml) was not developed for this assay. Data analyses restricted to Spike IgG-BA.1 strain bAbs, Spike IgG-Ancestral strain bAbs, and RBD IgG-Ancestral strain bAbs, where the latter marker was only studied in the multivariable statistical learning analyses.

#### Statistical Methods

All data analyses were prespecified in the SAP. All correlates of risk analyses adjusted for at-risk status [defined in (*1*)], predicted baseline risk score, and community of color classification (all persons other than white non-Hispanic). All controlled correlates of protection analyses further adjusted the BD1 level of the matching antibody marker.

Assessment of BD1, BD29, and Fold-Rise Markers as Correlates of Risk (CoRs)

Univariate analyses assessed each of the BD1, BD29, and Fold-Rise markers as CoRs of Omicron COVID-19 in the per-protocol boosted cohort (original-vaccine and crossover-vaccine arms combined). Analyses were performed as described previously (*4, 5*). In brief, the survey R package (*6*) was used to obtain point and 95% confidence interval (CI) estimates of the covariate-adjusted hazard ratio of Omicron COVID-19 across marker tertiles and per 10-fold increase in marker level. The analyses used inverse probability sampling–weighted Cox regression. Wald-based P values for an association of each antibody marker with Omicron COVID-19 are also reported. The same Cox models were also used to estimate antibody marker conditional cumulative incidence of the COVID-19 primary endpoint, with bootstrap 95% CIs reported. Nonparametric dose-response regression (*7*) was also used to estimate antibody marker conditional cumulative incidence of the COVID-19 primary endpoint, with influence function–based 95% CIs reported.

Cox regression models were also fitted with an interaction term between BD1 and BD29 marker levels, to investigate whether the BD1 marker modifies the effect of the BD29 biomarker. These models adjusted for at risk status, predicted baseline risk score, and community of color classification, as well as for BD1 marker level.

Point estimates of antibody marker threshold conditional cumulative incidence of Omicron COVID-19 and 95% point-wise CIs were calculated using nonparametric targeted minimum loss–based threshold regression (*8*).

Machine learning analysis (*9*) was also performed to estimate the best models for predicting Omicron COVID-19. The multivariable analysis was conducted as before (*5*), but with a few differences detailed in the SAP.

#### Antibody Decay and Cox Modeling for Booster Efficacy and Exposure Proximal Correlates

For each participant with antibody measurements on BD1 and DD1, a slope was calculated and the median slope was used to predict antibody concentration or titer at each following day for each individual with BD29 antibody using the formula Ab(d) = Ab29 + B d, where Ab29 is an individual’s log10 antibody concentration or titer on BD29, d is the number of days post BD29 and B is the median slope described above. This imputed antibody level was then used as a time-varying covariate in a Cox model

$h\left( t \right)=h_{0}\left( t \right) exp\{X\alpha+ Z(t) [ \beta_{0}+\beta_{1}Ab\left( d(t) \right)$] }w(t) R(t)

where X includes minority status, high risk, and risk score , t is the number of days since 12/1/2021, and Ab(d(t)) is an individual’s predicted log10 antibody level on day t, which is d(t) days post BD29, and Z(t) is 1 after boosting and 0 before, w(t) a weight and R(t) identifies when an individual is in the risk set (i.e. excluded from BD1 to BD29 + 6). Booster efficacy estimates remove Ab(d(t)) from the above model. BD29 is also assessed as a correlate using the above equation with Ab(d(t)) replaced with BD29 antibody.

#### Comparison to Ancestral Strain Correlates Study

The blinded phase correlates study (*4*) estimated how two-dose vs. placebo vaccine efficacy varied by Ancestral strain nAb titer at 4 weeks post dose 2 (Day 57), with Ancestral strain nAb titer calibrated to the WHO 20/136 International Standard and reported in International Units (IU/ml; IU50/ml was used for reporting of 50% inhibitory dilution nAb titer). It is of interest to compare this Ancestral strain antibody marker, Ancestral COVID-19 vaccine vs. placebo efficacy curve with the BA.1 strain antibody marker, Omicron COVID-19 booster relative (three-dose vs. two-dose) efficacy curve, to ascertain whether a different amount of variant-matched antibody is needed for high-level booster three dose vs. two dose protection than for high-level two dose vs. placebo protection. To do this, we defined a Predicted BA.1 strain nAb biomarker at BD29 scaled such that it can be absolutely quantitatively interpreted vs. Ancestral nAb in IU/50 ml units. This scaling was accomplished in two steps. First, the Duke/PPD Ancestral strain assay equivalency study (*10*) and the Duke assay International Standard calibration study (*11*) showed that multiplying PPD Ancestral strain nAb (ID50) readouts by (0.242/1.04) transforms units to the IU50/ml scale previously used (*4*). Second, based on data from 26 three-dose mRNA-1273 participants with Duke assay ID50 measured 4 weeks post dose 3 against both the Ancestral strain and the BA.1 strain (*12, 13*) [see Table 1 and Supplementary Table 2 in Hejazi et al. (*14*)], the geometric mean ratio of BA.1 strain nAb (ID50) vs. against Ancestral strain nAb (ID50) was 0.225. Therefore, we multiplied the PPD Ancestral strain nAb IU50/ml values by 0.225, attaining the Predicted BA.1 strain nAb values (thus original PPD BA.1 strain nAb units are multiplied by (0.242/1.04)*0.225 = 0.052 to generate Predicted BA.1 strain nAb ID50 units). The BD29 booster relative efficacy curve analysis was repeated for this biomarker, and results overlaid with the original Day 57 vaccine efficacy curve analysis, providing a means for absolute comparison of variant-matched titer levels associated with efficacy.

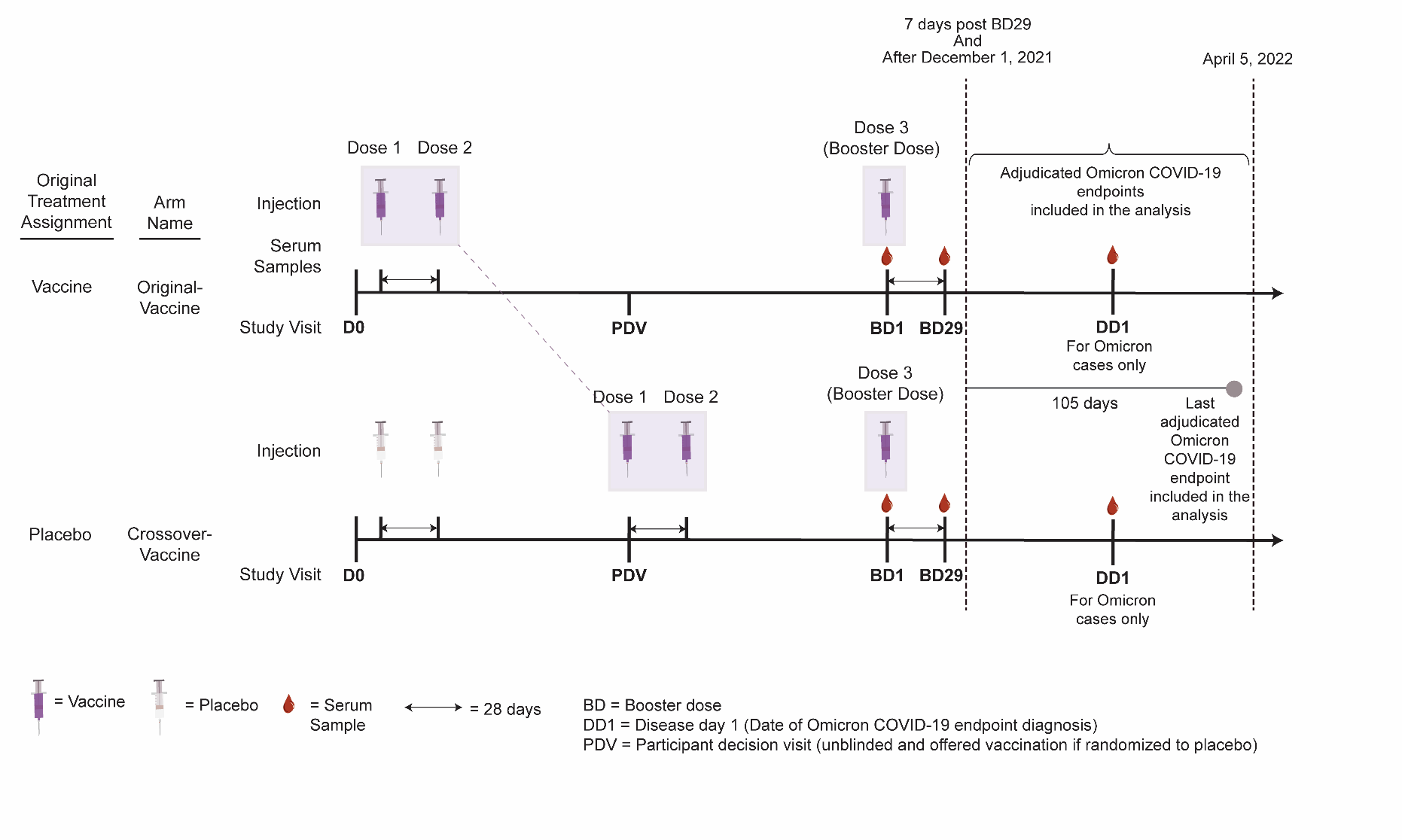

Figure S1. Timing of doses, study visits for serum sampling, and follow-up for Omicron COVID-19 endpoints included in the analysis. The median time interval between the second dose and the third (booster) dose was 12.9 months in the original-vaccine arm and 8.2 months in the crossover-vaccine arm.

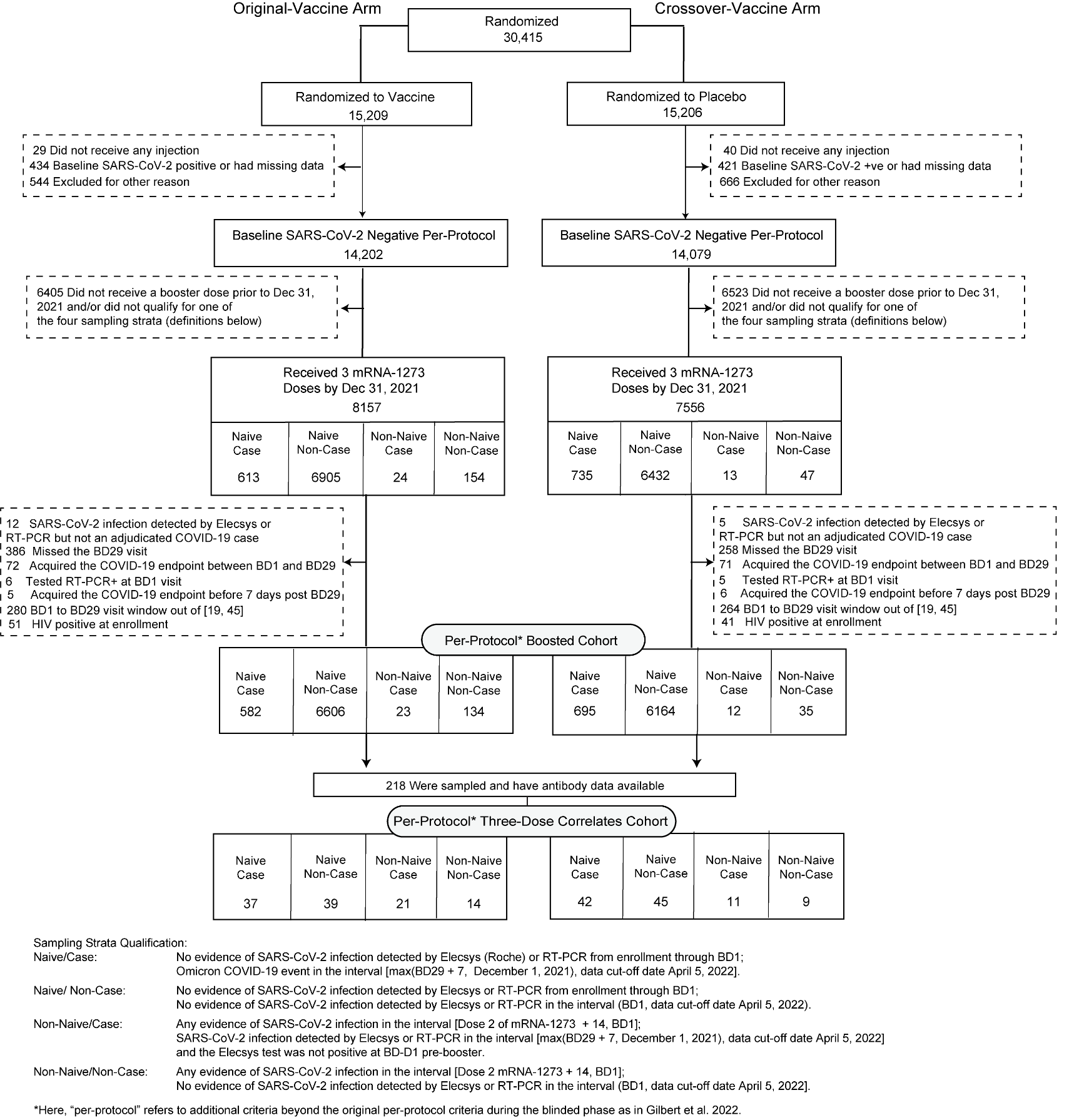

### Figure S2. Flowchart of study participants from enrollment in COVE through to the per-protocol three dose correlates cohort.

### Table S1. n = 218 sampled participants in the per-protocol three-dose correlates cohort (Fig. S2) by sampling stratum (N = SARS-CoV-2 naive and NN = Non-Naive) and time period of receipt of third (booster) dose.

|  | Time Period of Receipt of Third (Booster) Dose (BD1 Visit) | | | | | | | | Total |
| --- | --- | --- | --- | --- | --- | --- | --- | --- | --- |
|  | Sep 23 to Oct 15, 2021 | | Oct 16 to Oct 31,  2021 | | November, 2021 | | December, 2021 | |  |
| Original-Vaccine Arm $\times$ Omicron Case | 8N | 8NN | 11N | 5NN | 7N | 7NN | 11N | 1NN | 58 |
| Original-Vaccine Arm $\times$ Non-Case | 8N | 7NN | 10N | 2NN | 8N | 4NN | 13N | 1NN | 53 |
| Crossover-Vaccine Arm $\times$ Omicron Case | 9N | 5NN | 12N | 1NN | 10N | 5NN | 11N | 0NN | 53 |
| Crossover-Vaccine Arm $\times$ Non-Case | 9N | 4NN | 12N | 0NN | 10N | 4NN | 14N | 1NN | 54 |

Omicron Case= COVID-19 endpoint in the interval [≥ 7 days post BD29 AND ≥ December 1, 2021 to April 5, 2022 data cutoff date]. As described in the SAP (Appendix A) the COVID-19 endpoint is documented to be Omicron BA.1 if possible whereas for some non-naive COVID-19 endpoints there was not lineage data available to document the case to be Omicron BA.1.

Non-case = Did not acquire COVID-19 (of any strain) in the interval [BD1, data cutoff date].

SARS-CoV-2 naive (N) = No evidence of SARS-CoV-2 infection from enrollment through to BD1; Non-naive (NN) = Any evidence of SARS-CoV-2 infection in the interval [≥ 14 days after the first two doses of mRNA-1273, BD1]

### Table S2. Demographic and clinical information of the per-protocol boosted cohort and the per-protocol three-dose correlates cohort (original-vaccine and crossover-vaccine arms combined)

|  | **Per-Protocol Boosted Cohort**  **(Original Vaccine and Crossover Vaccine Combined)** | | | **Per-Protocol Three-Dose Correlates Cohort**  **(Original Vaccine and Crossover Vaccine Combined)** | | |
| --- | --- | --- | --- | --- | --- | --- |
| **Characteristics** | **SARS-CoV-2 Naive (N=14047)** | **Non-Naive (N=204)** | **Total (N=14251)** | **SARS-CoV-2 Naive (N=163)** | **Non-Naive (N=55)** | **Total (N=218)** |
| **Age** | | | |  |  |  |
| Age < 65 | 9683 (68.9%) | 155 (76.0%) | 9838 (69.0%) | 121 (74.2%) | 44 (80.0%) | 165 (75.7%) |
| Age ≥ 65 | 4364 (31.1%) | 49 (24.0%) | 4413 (31.0%) | 42 (25.8%) | 11 (20.0%) | 53 (24.3%) |
| Mean (Range) | 54.1 (18.0, 95.0) | 51.6 (19.0, 84.0) | 54.0 (18.0, 95.0) | 54.4 (20.0, 80.0) | 49.3 (21.0, 77.0) | 53.1 (20.0, 80.0) |
| **BMI** | | | |  |  |  |
| Mean ± SD | 29.3 ± 6.7 | 30.2 ± 7.8 | 29.3 ± 6.7 | 29.9 *±* 7.7 | 29.8 *±* 5.7 | 29.8 *±* 7.2 |
| **Risk for Severe COVID-19** | | | |  |  |  |
| At-risk | 3359 (23.9%) | 55 (27.0%) | 3414 (24.0%) | 43 (26.4%) | 14 (25.5%) | 57 (26.1%) |
| Not at-risk | 10688 (76.1%) | 149 (73.0%) | 10837 (76.0%) | 120 (73.6%) | 41 (74.5%) | 161 (73.9%) |
| **Age, Risk for Severe COVID-19** | | | |  |  |  |
| Age < 65 At-risk | 2086 (14.9%) | 37 (18.1%) | 2123 (14.9%) | 29 (17.8%) | 10 (18.2%) | 39 (17.9%) |
| Age < 65 Not at-risk | 7597 (54.1%) | 118 (57.8%) | 7715 (54.1%) | 92 (56.4%) | 34 (61.8%) | 126 (57.8%) |
| Age ≥ 65 | 4364 (31.1%) | 49 (24.0%) | 4413 (31.0%) | 42 (25.8%) | 11 (20.0%) | 53 (24.3%) |
| **Sex Assigned at Birth** | | | |  |  |  |
| Female | 6783 (48.3%) | 100 (49.0%) | 6883 (48.3%) | 94 (57.7%) | 23 (41.8%) | 117 (53.7%) |
| Male | 7264 (51.7%) | 104 (51.0%) | 7368 (51.7%) | 69 (42.3%) | 32 (58.2%) | 101 (46.3%) |
| **Hispanic or Latino Ethnicity** | | | |  |  |  |
| Hispanic or Latino | 2496 (17.8%) | 39 (19.1%) | 2535 (17.8%) | 32 (19.6%) | 12 (21.8%) | 44 (20.2%) |
| Not Hispanic or Latino | 11405 (81.2%) | 165 (80.9%) | 11570 (81.2%) | 131 (80.4%) | 43 (78.2%) | 174 (79.8%) |
| Not reported and unknown | 146 (1.0%) | 0 (0%) | 146 (1.0%) | 0 (0%) | 0 (0%) | 0 (0%) |
| **Race** | | | |  |  |  |
| White | 11189 (79.7%) | 156 (76.5%) | 11345 (79.6%) | 136 (83.4%) | 41 (74.5%) | 177 (81.2%) |
| Black or African American | 1383 (9.8%) | 28 (13.7%) | 1411 (9.9%) | 14 (8.6%) | 5 (9.1%) | 19 (8.7%) |
| Asian | 613 (4.4%) | 8 (3.9%) | 621 (4.4%) | 4 (2.5%) | 3 (5.5%) | 7 (3.2%) |
| American Indian or Alaska Native | 109 (0.8%) | 1 (0.5%) | 110 (0.8%) | 1 (0.6%) | 1 (1.8%) | 2 (0.9%) |
| Native Hawaiian or Other Pacific Islander | 29 (0.2%) | 1 (0.5%) | 30 (0.2%) | 2 (1.2%) | 0 (0%) | 2 (0.9%) |
| Multiracial | 310 (2.2%) | 6 (2.9%) | 316 (2.2%) | 2 (1.2%) | 3 (5.5%) | 5 (2.3%) |
| Other | 267 (1.9%) | 4 (2.0%) | 271 (1.9%) | 3 (1.8%) | 2 (3.6%) | 5 (2.3%) |
| Not reported and unknown | 134 (1.0%) | 0 (0%) | 134 (0.9%) | 1 (0.6%) | 0 (0%) | 1 (0.5%) |
| **Underrepresented Minority Status** |  |  |  |  |  |  |
| White Non-Hispanic | 10133 (72.1%) | 140 (68.6%) | 10273 (72.1%) | 115 (70.6%) | 38 (69.1%) | 153 (70.2%) |
| Communities of Color | 3914 (27.9%) | 64 (31.4%) | 3978 (27.9%) | 48 (29.4%) | 17 (30.9%) | 65 (29.8%) |

This table summarizes the per-protocol boosted cohort, which was randomly sampled within 12 strata deﬁned by enrollment characteristics: Assigned treatment arm × Baseline SARS-CoV-2 naive vs. non-naive status (deﬁned by serostatus and NAAT testing) × Randomization strata (Age < 65 and at-risk, Age < 65 and not at-risk, Age ≥ 65)× Minority status (Minority vs. Non-minority) deﬁned by White Non-Hispanic vs. all others [same as in (El Sahly et al. (*2*))]. “At Risk” refers to participants believed to be at increased risk of severe COVID-19 illness and comprised six self-reported health/comorbidities, as in El Sahly et al. (*2*) “Minority” includes Blacks or African Americans, Hispanics or Latinos, American Indians or Alaska Natives, Native Hawaiians, and other Pacific Islanders. Non-Minority includes all other races with observed race (Asian, Multiracial, White, Other) and observed ethnicity Not Hispanic or Latino. Numbers and percentages are based on inverse probability of sampling weighting.

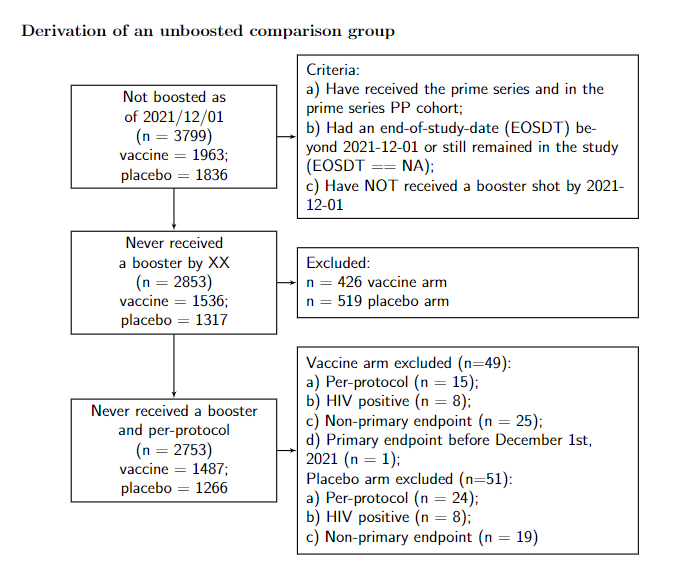

### Figure S3. Flow of baseline-negative per-protocol [according to the definition in Gilbert et al. (*4*)] participants who were still in the study and had not received a third (booster) dose as of December 1, 2022 through to inclusion in the exposure-proximal CoP analysis. These 2753 participants were used to enrich the analysis cohort for the exposure-proximal CoP analysis.

### Table S3. Assay limits for A) the PPD pseudovirus neutralizing antibody (nAb) assay and B) the PPD VAC123 MSD multiplex assay by antigen. Note that the Ancestral strain Spike used for pseudotyping in the nAb assay has the D614G mutation, whereas the Ancestral strain Spike used in the bAb assay does not (D614).

| **A** | **Neutralizing antibody assay** | |
| --- | --- | --- |
|  | Ancestral strain | Omicron BA.1 |
| LLOQ (AU/ml) | 10 | 8 |
| ULOQ (AU/ml) | 281,600 | 24,503 |
| **B** | **Binding antibody assay (Spike IgG)** | |
|  | Ancestral strain | Omicron BA.1/B.1.1.529 |
| LLOQ^1^ (AU/ml) | 69 | 102 |
| ULOQ^2^ (AU/ml) | 14,400,000 | 1,180,000 |

^1^LLOQs were taken as the LLOQs for the lowest dilution (1:500).

^2^ULOQs were taken as the ULOQs for the highest dilution (1:500,000).

For all assays, values < LLOQ were set to LLOQ/2 and for the neutralizing antibody assay, values > ULOQ were set to ULOQ. AU = arbitrary units.

### Table S4. BD1 Ancestral strain neutralizing antibody (nAb) and BD1 Spike IgG-Ancestral strain binding antibody (bAb) response rates and geometric means stratified by Omicron COVID-19 case vs. non-case and by SARS-CoV-2 naive vs. non-naive status in the per-protocol boosted cohort, pooled across the original-vaccine and crossover-vaccine arms

|  |  |  | **Omicron Cases^1^** | | | **Non-Cases^2^** | | | **Comparison** | |
| --- | --- | --- | --- | --- | --- | --- | --- | --- | --- | --- |
| **Status^3^** | **Marker** | **Measurement** | **N^4^** | **Response Rate^5^ (95% CI)** | **GMC or GMT (AU/ml) (95% CI)** | **N^4^** | **Response Rate^5^ (95% CI)** | **GMC or GMT (AU/ml) (95% CI)** | **Response Rate  Difference (Omicron Cases- Non-Cases)  (95% CI)** | **Ratio of GM  (Omicron Cases/Non-Cases)  (95% CI)** |
| SARS-CoV-2 naive | Ancestral strain nAbs | BD1 | 79 | 100%  (100%, 100%) | 124  (89.7, 170) | 84 | 99.7%  (98.1%, 100%) | 114  (75.9, 173) | 0.003  (0, 0.019) | 1.08  (0.64, 1.82) |
| SARS-CoV-2 naive | Spike IgG-Ancestral strain bAbs | BD1 | 79 | 100%  (100%, 100%) | 19100.77  (13777.90, 26480.05) | 84 | 100%  (100%, 100%) | 18213.80  (14060.12, 23594.56) | 0  (0, 0) | 1.05  (0.69, 1.59) |
| Non-Naive | Ancestral strain nAbs | BD1 | 32 | 100%  (100%, 100%) | 437  (247, 770) | 23 | 100%  (100%, 100%) | 148  (82.3, 264) | 0  (0, 0) | 2.96  (1.31, 6.68) |
| Non-Naive | Spike IgG-Ancestral strain bAbs | BD1 | 32 | 100%  (100%, 100%) | 51318  (30442, 86512) | 23 | 100%  (100%, 100%) | 24521  (16224, 37062) | 0  (0, 0) | 2.09  (1.08, 4.07) |

^1^Omicron case = COVID-19 Omicron BA.1 endpoint that occurred in the interval [≥ 7 days post BD29 AND ≥ December 1, 2021 to April 5, 2022 data cutoff].

^2^Non-case = No acquirement of COVID-19 (of any strain) in the interval [BD1, April 5, 2022 data cutoff].

^3^SARS-CoV-2 naive = No evidence of SARS-CoV-2 infection from enrollment through to BD1; Non-naive = Any evidence of SARS-CoV-2 infection in the interval [≥ 14 days after the original two-dose series, BD1]

^4^N is the number of cases sampled into the subcohort within baseline covariate strata.

^5^Definitions of “responder” for each BD1 marker: positive (quantifiable) response defined as BD1 Ancestral strain nAb ≥ 10 AU/ml; positive response defined as BD1 Spike IgG-Ancestral strain bAb ≥ 69 AU/ml.

AU/ml, arbitrary units/ml; CI: confidence interval; GMC: geometric mean concentration; GMT: geometric mean titer.

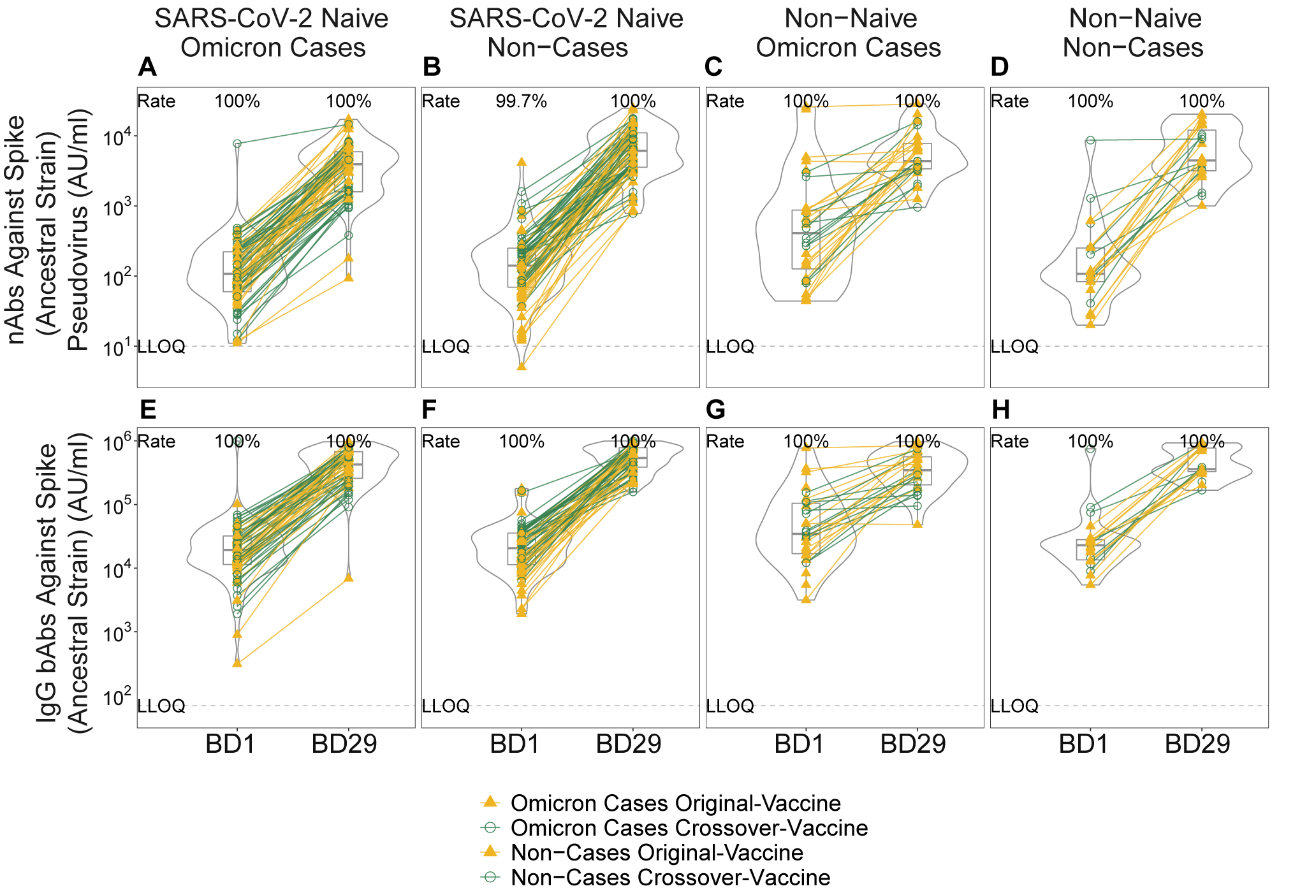

Figure S4. Distributions of BD1 and BD29 (A-D) Ancestral strain neutralizing antibody (nAb) titer and (E-H) Spike IgG-Ancestral strain binding antibody (bAb) concentration, stratified by Omicron COVID-19 case vs. non-case status and by SARS-CoV-2 naive vs. non-naive status. Data points are from per-protocol boosted participants in the original-vaccine (filled triangles) or crossover-vaccine (open circles) arm, with lines (yellow: original-vaccine arm; green: crossover-vaccine arm) connecting the BD1 and BD29 data points for an individual participant. The violin plots contain interior box plots with upper and lower horizontal edges representing the 25^th^ and 75^th^ percentiles of antibody level and middle line representing the 50^th^ percentile. The vertical bars represent the distance from the 25^th^ (or 75^th^) percentile of antibody level and the minimum (or maximum) antibody level within the 25^th^ (or 75^th^) percentile of antibody level minus (or plus) 1.5 times the interquartile range. Each side shows a rotated probability density (estimated by a kernel density estimator with a default Gaussian kernel) of the data. Positive response rates were computed with inverse probability of sampling weighting. LLOQ, lower limit of quantification. LLOQ = 10 AU/ml for Ancestral strain nAbs and 69 AU/ml for Spike IgG-Ancestral strain bAbs. Positive response for Ancestral strain nAbs at a given timepoint was defined by value ≥ LLOQ at that timepoint. Positive response for Spike IgG-Ancestral strain bAbs at a given timepoint was defined by value ≥ LLOQ at that timepoint. Omicron Case = COVID-19 endpoint in the interval [≥ 7 days post BD29 AND ≥ December 1, 2021 to April 5, 2022 data cutoff date]. Non-case = Did not acquire COVID-19 (of any strain) in the interval [BD1 to April 5, 2022]. SARS-CoV-2 naive = No evidence of SARS-CoV-2 infection from enrollment through to BD1; Non-naive = Any evidence of SARS-CoV-2 infection in the interval [≥ 14 days after the first two doses of mRNA-1273, BD1].

### Table S5. BD29 and Fold-Rise Ancestral strain neutralizing antibody (nAb) and Spike IgG-Ancestral strain binding antibody (bAb) response rates and geometric means by Omicron COVID-19 case vs. non-case status and by SARS-CoV-2 naive vs. non-naive status in the per-protocol boosted cohort, pooled across the original-vaccine and crossover-vaccine arms

|  |  |  | **Omicron Cases^1^** | | | **Non-Cases^2^** | | | **Comparison** | |
| --- | --- | --- | --- | --- | --- | --- | --- | --- | --- | --- |
| **Status^3^** | **Marker** | **Measurement** | **N^4^** | **Response Rate^5^ (95% CI)** | **GMC or GMT (AU/ml) (95% CI)** | **N^4^** | **Response Rate^5^ (95% CI)** | **GMC or GMT (AU/ml) (95% CI)** | **Response Rate  Difference (Omicron Cases- Non-Cases)  (95% CI)** | **Ratio of GM  (Omicron Cases/Non-Cases)  (95% CI)** |
| SARS-CoV-2 Naive | Ancestral Strain nAbs | BD29 | 79 | 100% (100%, 100%) | 3234  (2385, 4387) | 84 | 100% (100%, 100%) | 5492  (3866, 7802) | 0 (0, 0) | 0.59  (0.37, 0.94) |
| SARS-CoV-2 Naive | Spike IgG-Ancestral Strain bAbs | BD29 | 79 | 100% (100%, 100%) | 467178 (364292, 599121) | 84 | 100% (100%, 100%) | 652950 (516281, 825799) | 0 (0, 0) | 0.72 (0.51, 1.01) |
| SARS-CoV-2 Naive | Ancestral Strain nAbs | Fold-Rise | 79 | - | 26.2  (21.1, 32.4) | 84 | - | 48.0  (37.4, 61.6) | - | 0.54  (0.39, 0.76) |
| SARS-CoV-2 Naive | Spike IgG-Ancestral Strain bAbs | Fold-Rise | 79 | - | 24.5  (20.0, 30.0) | 84 | - | 35.9  (29.4, 43.7) | - | 0.68  (0.51, 0.91) |
| Non-Naive | Ancestral Strain nAbs | BD29 | 32 | 100% (100%, 100%) | 5536  (4012, 7637) | 23 | 100% (100%, 100%) | 5759  (3526, 9407) | 0 (0, 0) | 0.96  (0.53, 1.73) |
| Non-Naive | Spike IgG-Ancestral Strain bAbs | BD29 | 32 | 100% (100%, 100%) | 419795  (301040, 585399) | 23 | 100% (100%, 100%) | 619269  (441303, 869004) | 0 (0, 0) | 0.68  (0.42, 1.09) |
| Non-Naive | Ancestral Strain nAbs | Fold-Rise | 32 | - | 12.7  (7.5, 21.6) | 23 | - | 39.0  (23.9, 63.9) | - | 0.32  (0.16, 0.67) |
| Non-Naive | Spike IgG-Ancestral Strain bAbs | Fold-Rise | 32 | - | 8.2  (4.7, 14.1) | 23 | - | 25.3  (16.7, 38.2) | - | 0.32  (0.16, 0.64) |

^1^Omicron case = COVID-19 Omicron BA.1 endpoint that occurred in the interval [≥ 7 days post BD29 AND ≥ December 1, 2021 to April 5, 2022 data cutoff].

^2^Non-case = No acquirement of COVID-19 (of any strain) in the interval [BD1, April 5, 2022 data cutoff].

^3^SARS-CoV-2 naive = No evidence of SARS-CoV-2 infection from enrollment through to BD1; Non-naive = Any evidence of SARS-CoV-2 infection in the interval [≥ 14 days after the original two-dose series, BD1]

^4^N is the number of cases sampled into the subcohort within baseline covariate strata.

^5^Definitions of “responder” for the BD29 markers: positive (quantifiable) response defined as BD29 Ancestral strain nAbs ≥ 10 AU/ml; positive response defined as BD29 Spike IgG-Ancestral strain bAbs ≥ 69 AU/ml.

AU/ml, arbitrary units/ml; CI: confidence interval; GMC: geometric mean concentration; GMT: geometric mean titer.

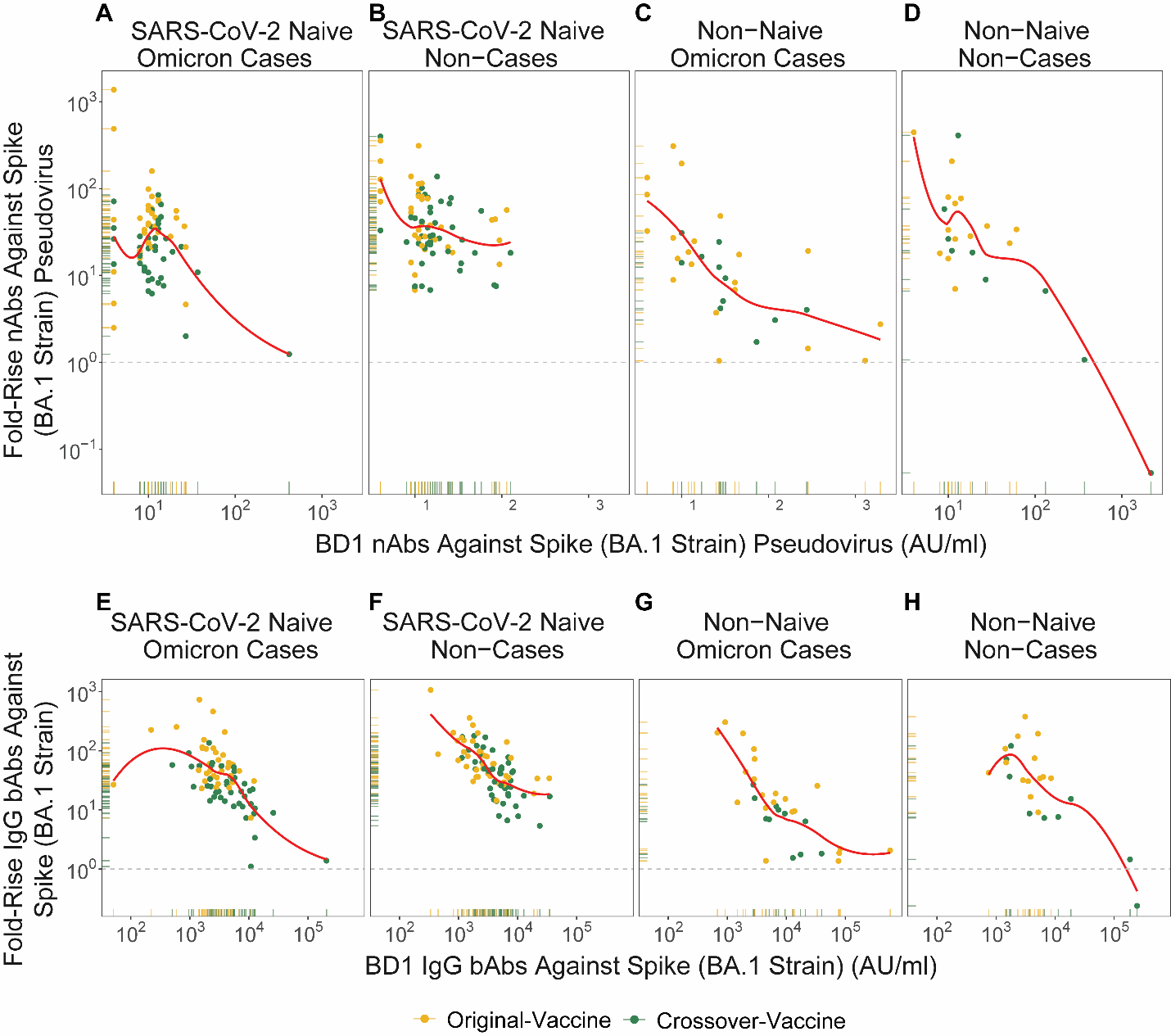

Figure S5. Scatterplots with rugs of BD1 and Fold-Rise (BD29/BD1) (A-D) BA.1 strain neutralizing antibody (nAb) and (E-F) Spike IgG-BA.1 strain binding antibody (bAb) level, stratified by Omicron COVID-19 case vs. non-case status and by SARS-CoV-2 naive vs. non-naive status. Data points are from per-protocol boosted participants in the original-vaccine (yellow) or crossover-vaccine (green) arm. Omicron Case = Omicron COVID-19 endpoint in the interval [≥ 7 days post BD29 AND ≥ December 1, 2021 to April 5, 2022 data cutoff date]. Non-case = Did not acquire COVID-19 (of any strain) in the interval [BD1 to April 5, 2022]. SARS-CoV-2 naive = No evidence of SARS-CoV-2 infection from enrollment through to BD1; Non-naive = Any evidence of SARS-CoV-2 infection in the interval [≥ 14 days after the first two doses of mRNA-1273, BD1].

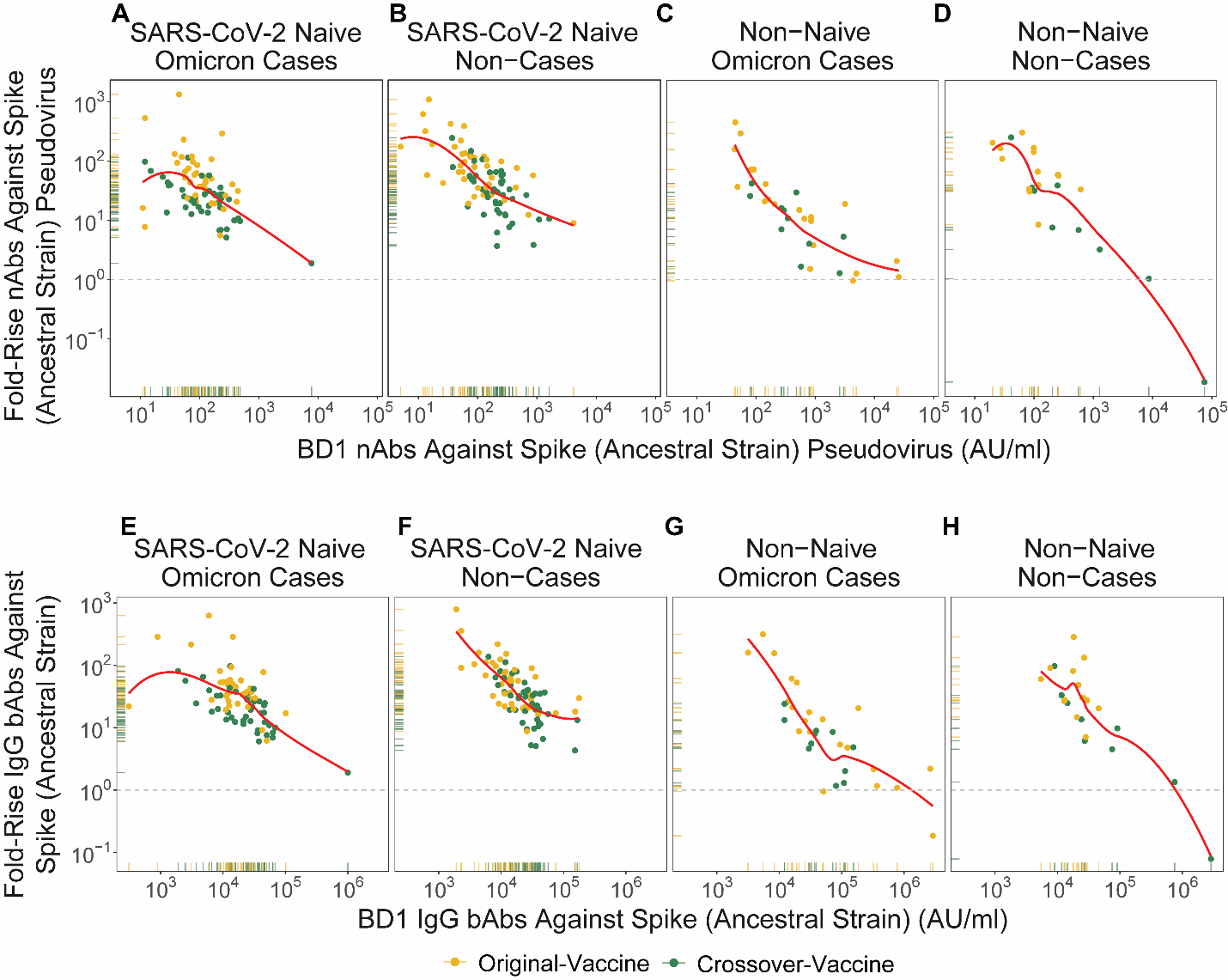

Figure S6. Scatterplots with rugs of BD1 and Fold-Rise (BD29/BD1) (A-D) Ancestral strain neutralizing antibody (nAb) and (E-F) Spike IgG-Ancestral strain binding antibody (bAb) level, stratified by Omicron COVID-19 case vs. non-case status and by SARS-CoV-2 naive vs. non-naive status. Data points are from per-protocol boosted participants in the original-vaccine (yellow) or crossover-vaccine (green) arm. Omicron Case = Omicron COVID-19 endpoint in the interval [≥ 7 days post BD29 AND ≥ December 1, 2021 to April 5, 2022 data cutoff date]. Non-case = Did not acquire COVID-19 (of any strain) in the interval [BD1 to April 5, 2022]. SARS-CoV-2 naive (N) = No evidence of SARS-CoV-2 infection from enrollment through to BD1; Non-naive (NN) = Any evidence of SARS-CoV-2 infection in the interval [≥ 14 days after the first two doses of mRNA-1273, BD1].

### Table S6. BD29 and BD29/BD1 Fold-Rise neutralizing antibody (nAb) and binding antibody (bAb) response rates and geometric means in non-cases in the per-protocol boosted cohort, shown separately by SARS-CoV-2 naive vs. non-naive status and by study arm

| **Arm** | **Status** | **Marker** | **Visit** | **N** | **Response Rate (95% CI)** | **GMC or GMT (AU/ml) (95% CI)** |
| --- | --- | --- | --- | --- | --- | --- |
|  |  | **BA.1 Strain** |  |  |  |  |
| Original-Vaccine | SARS-CoV-2 Naive | BA.1 Strain nAbs | BD29 | 6606 | 100 (100, 100) | 527  (276, 1006) |
| Original-Vaccine | SARS-CoV-2 Naive | Spike IgG-BA.1 Strain bAbs | BD29 | 6606 | 100 (100, 100) | 194468  (134566, 281036) |
| Original-Vaccine | SARS-CoV-2 Naive | BA.1 Strain nAbs | Fold-Rise | 6606 | - | 37.4  (21.6, 64.8) |
| Original-Vaccine | SARS-CoV-2 Naive | Spike IgG-BA.1 Strain bAbs | Fold-Rise | 6606 | - | 74.8  (58.1, 96.4) |
| Original-Vaccine | Non-Naive | BA.1 Strain nAbs | BD29 | 134 | 100 (100, 100) | 616  (336, 1131) |
| Original-Vaccine | Non-Naive | Spike IgG-BA.1 Strain bAbs | BD29 | 134 | 100 (100, 100) | 165136  (98711, 276261) |
| Original-Vaccine | Non-Naive | BA.1 Strain nAbs | Fold-Rise | 134 | - | 40.3  (26.5, 61.4) |
| Original-Vaccine | Non-Naive | Spike IgG-BA.1 Strain bAbs | Fold-Rise | 134 | - | 46.5  (32.2, 67.3) |
| Crossover-Vaccine | SARS-CoV-2 Naive | BA.1 Strain nAbs | BD29 | 6164 | 100 (100, 100) | 455  (329, 629) |
| Crossover-Vaccine | SARS-CoV-2 naive | Spike IgG-BA.1 Strain bAbs | BD29 | 6164 | 100 (100, 100) | 148498  (117406, 187823) |
| Crossover-Vaccine | SARS-CoV-2 Naive | BA.1 Strain nAbs | Fold-Rise | 6164 | - | 30.1  (21.8, 41.6) |
| Crossover-Vaccine | SARS-CoV-2 Naive | Spike IgG-BA.1 Strain bAbs | Fold-Rise | 6164 | - | 33.7  (24.7, 46.0) |
| Crossover-Vaccine | Non-Naive | BA.1 Strain nAbs | BD29 | 35 | 100 (100, 100) | 432  (212, 879) |
| Crossover-Vaccine | Non-Naive | Spike IgG-BA.1 Strain bAbs | BD29 | 35 | 100 (100, 100) | 98349  (58932, 164131) |
| Crossover-Vaccine | Non-Naive | BA.1 Strain nAbs | Fold-Rise | 35 | - | 9.7  (1.9, 49.4) |
| Crossover-Vaccine | Non-Naive | Spike IgG-BA.1 Strain bAbs | Fold-Rise | 35 | - | 9.8  (2.8, 33.9) |
|  |  | **Ancestral strain** |  |  |  |  |
| Original-Vaccine | SARS-CoV-2 Naive | Ancestral Strain nAbs | BD29 | 6606 | 100 (100, 100) | 5859  (3101, 11067) |
| Original-Vaccine | SARS-CoV-2 Naive | Spike IgG-Ancestral Strain bAbs | BD29 | 6606 | 100 (100, 100) | 715670  (471776, 1085650) |
| Original-Vaccine | SARS-CoV-2 Naive | Ancestral Strain nAbs | Fold-Rise | 6606 | - | 77.6  (52.4, 115) |
| Original-Vaccine | SARS-CoV-2 Naive | Spike IgG-Ancestral Strain bAbs | Fold-Rise | 6606 | - | 53.1  (38.8, 72.8) |
| Original-Vaccine | Non-Naive | Ancestral Strain nAbs | BD29 | 134 | 100 (100, 100) | 6297  (3433, 11550) |
| Original-Vaccine | Non-Naive | Spike IgG-Ancestral Strain bAbs | BD29 | 134 | 100 (100, 100) | 682910  (452891, 1029752) |
| Original-Vaccine | Non-Naive | Ancestral Strain nAbs | Fold-Rise | 134 | - | 61.4  (41.6, 90.8) |
| Original-Vaccine | Non-Naive | Spike IgG-Ancestral Strain bAbs | Fold-Rise | 134 | - | 35.6  (24.7, 51.2) |
| Crossover-Vaccine | SARS-CoV-2 Naive | Ancestral Strain nAbs | BD29 | 6164 | 100 (100, 100) | 5125  (3859, 6806) |
| Crossover-Vaccine | SARS-CoV-2 Naive | Spike IgG-Ancestral Strain bAbs | BD29 | 6164 | 100 (100, 100) | 591821  (474520, 738120) |
| Crossover-Vaccine | SARS-CoV-2 Naive | Ancestral Strain nAbs | Fold-Rise | 6164 | - | 28.7  (20.3, 40.6) |
| Crossover-Vaccine | SARS-CoV-2 Naive | Spike IgG-Ancestral Strain bAbs | Fold-Rise | 6164 | - | 23.5  (17.7, 31.3) |
| Crossover-Vaccine | Non-Naive | Ancestral Strain nAbs | BD29 | 35 | 100 (100, 100) | 4092  (2534, 6608) |
| Crossover-Vaccine | Non-Naive | Spike IgG-Ancestral Strain bAbs | BD29 | 35 | 100 (100, 100) | 425821  (283019, 640676) |
| Crossover-Vaccine | Non-Naive | Ancestral Strain nAbs | Fold-Rise | 35 | - | 6.9  (1.2, 40.3) |
| Crossover-Vaccine | Non-Naive | Spike IgG-Ancestral Strain bAbs | Fold-Rise | 35 | - | 6.8  (1.8, 25.9) |

Fold-Rise = BD29/BD1. N is the number of cases sampled into the subcohort within baseline covariate strata. Non-case = No acquirements of COVID-19 (of any strain) in the interval [BD1, data cutoff date]. SARS-CoV-2 naive = No evidence of SARS-CoV-2 infection from enrollment through to BD1. Non-naive = Any evidence of SARS-CoV-2 infection in the interval [≥ 14 days after the original 2-dose series, BD1]

GMC: geometric mean concentration; GMT: geometric mean titer.

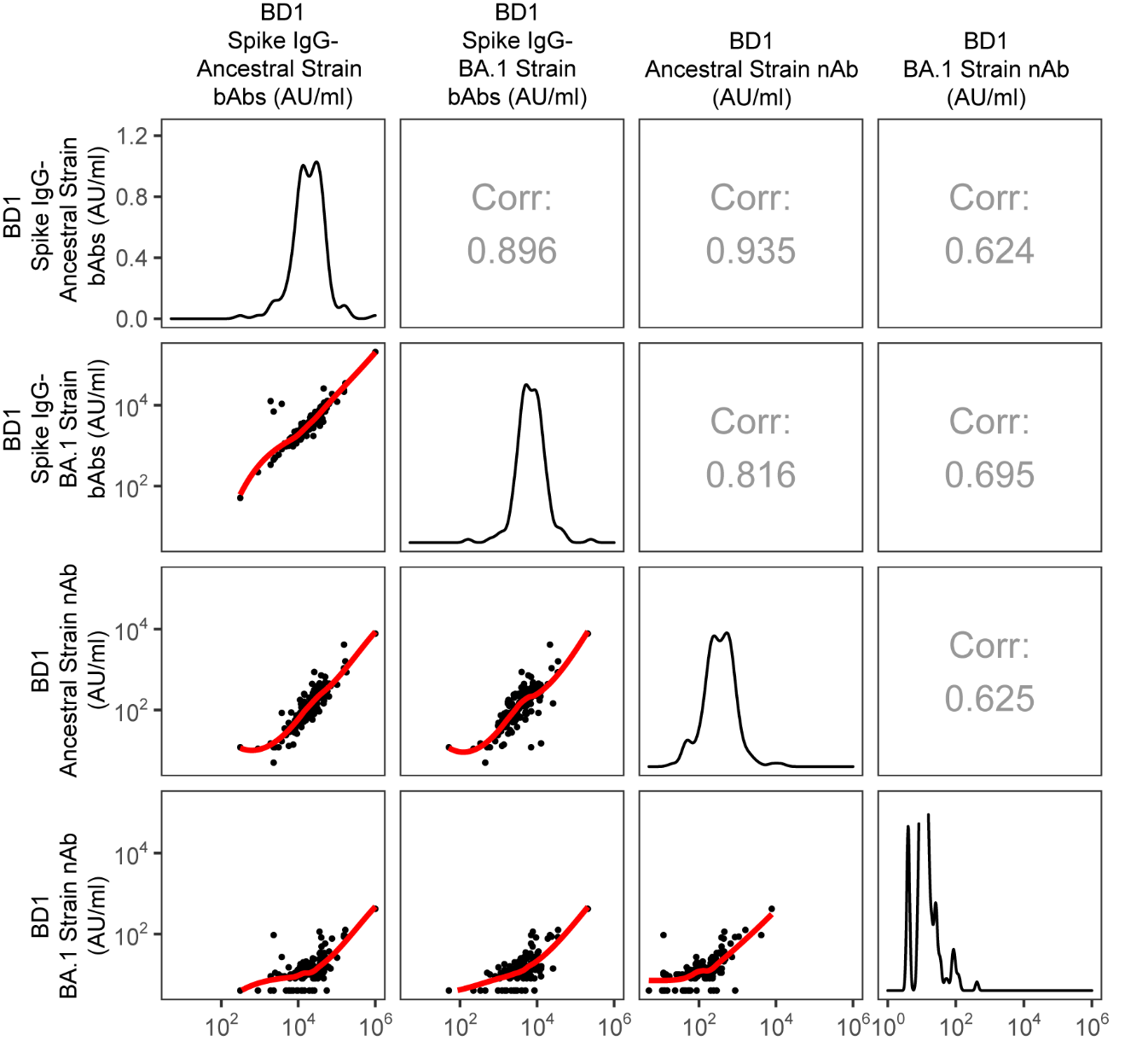

### Figure S7. Correlations of BD1 antibody markers among SARS-CoV-2 naives in the per-protocol boosted cohort. Corr = Inverse probability weight adjusted Spearman's rank correlation coefficient.

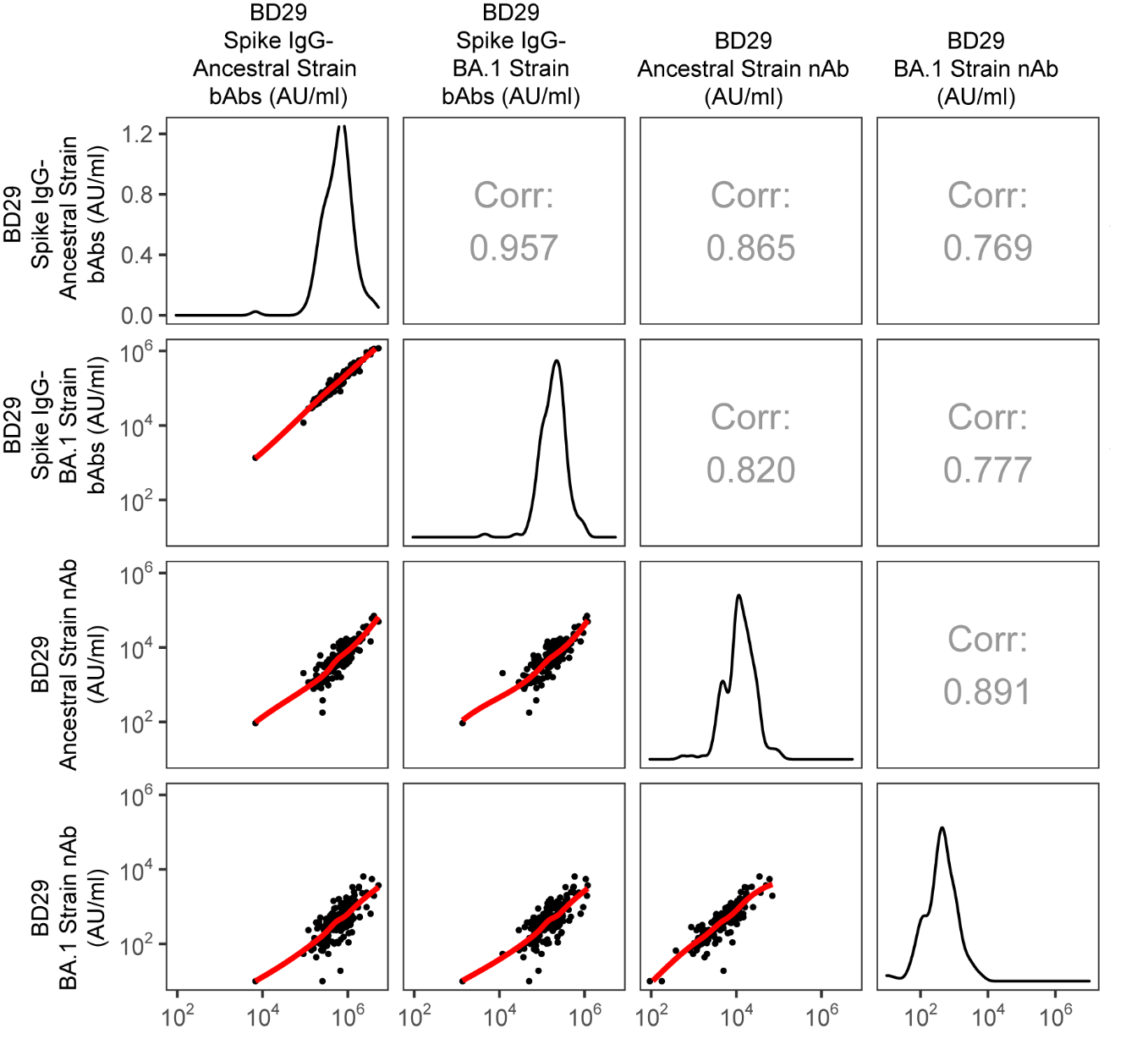

### Figure S8. Correlations of BD29 antibody markers among SARS-CoV-2 naives in the per-protocol boosted cohort. Corr = Inverse probability weight adjusted Spearman's rank correlation coefficient.

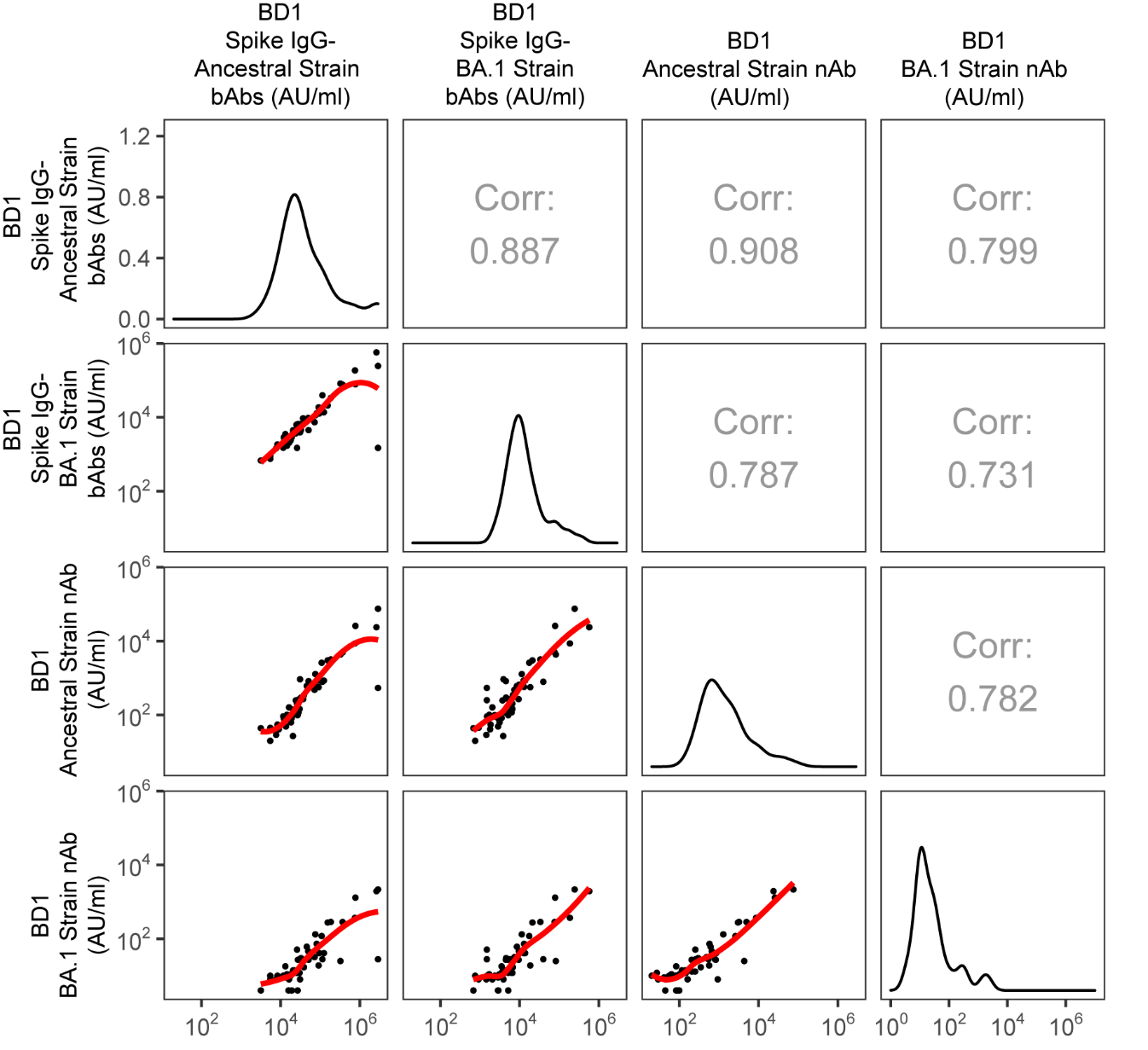

### Figure S9. Correlations of BD1 antibody markers among non-naives in the per-protocol boosted cohort. Corr = Inverse probability weight adjusted Spearman's rank correlation coefficient.

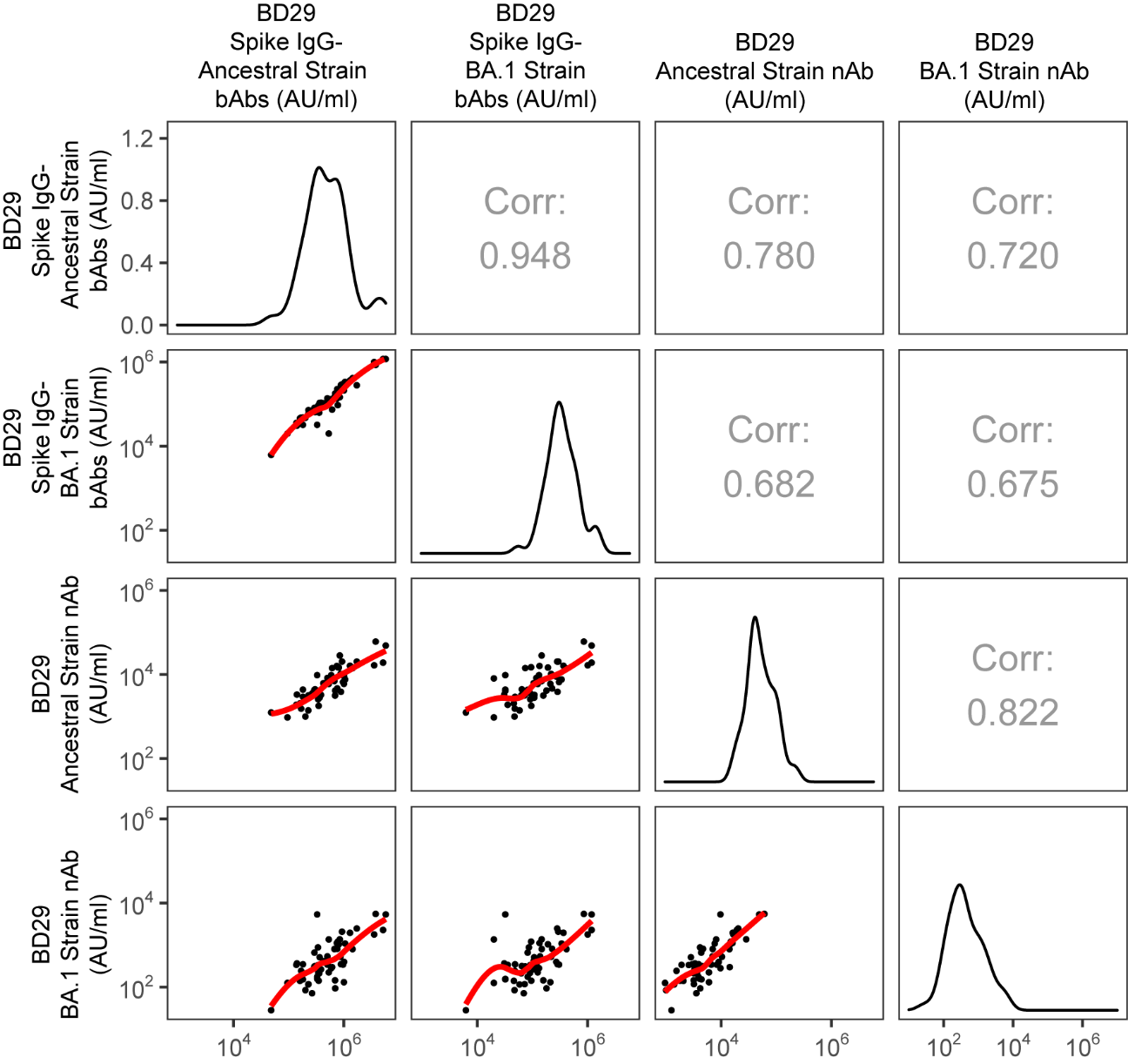

### Figure S10. Correlations of BD29 antibody markers among non-naives in the per-protocol boosted cohort. Corr = Inverse probability weight adjusted Spearman's rank correlation coefficient.

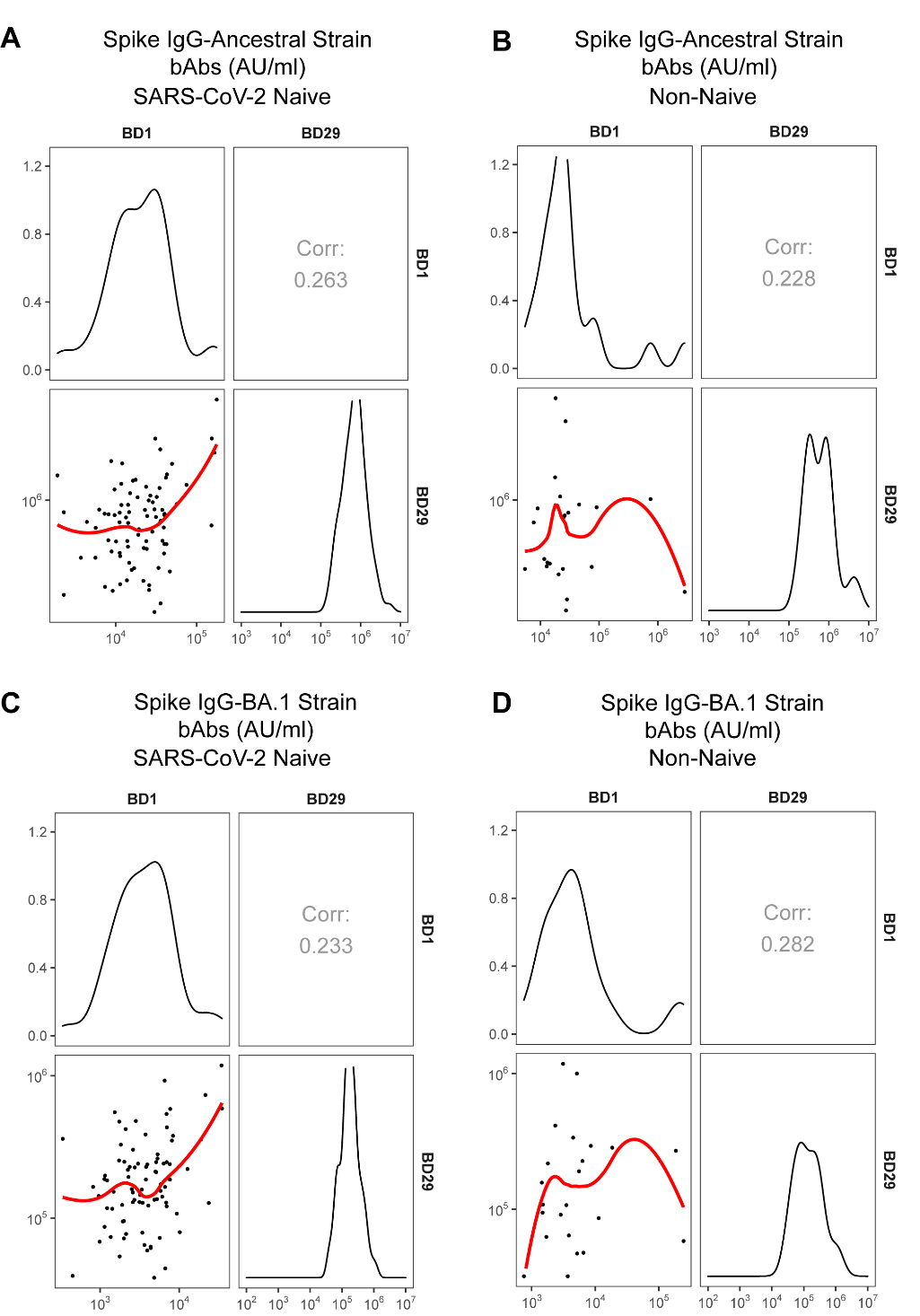

### Figure S11. Correlations between BD1 and BD29 (A, B) Spike IgG-Ancestral strain binding antibody (bAb) and (C, D) Spike IgG-BA.1 strain bAb concentrations among (A, C) SARS-CoV-2 naives and (B, D) non-naives in the per-protocol boosted cohort. Corr = Inverse probability weight adjusted Spearman's rank correlation coefficient.

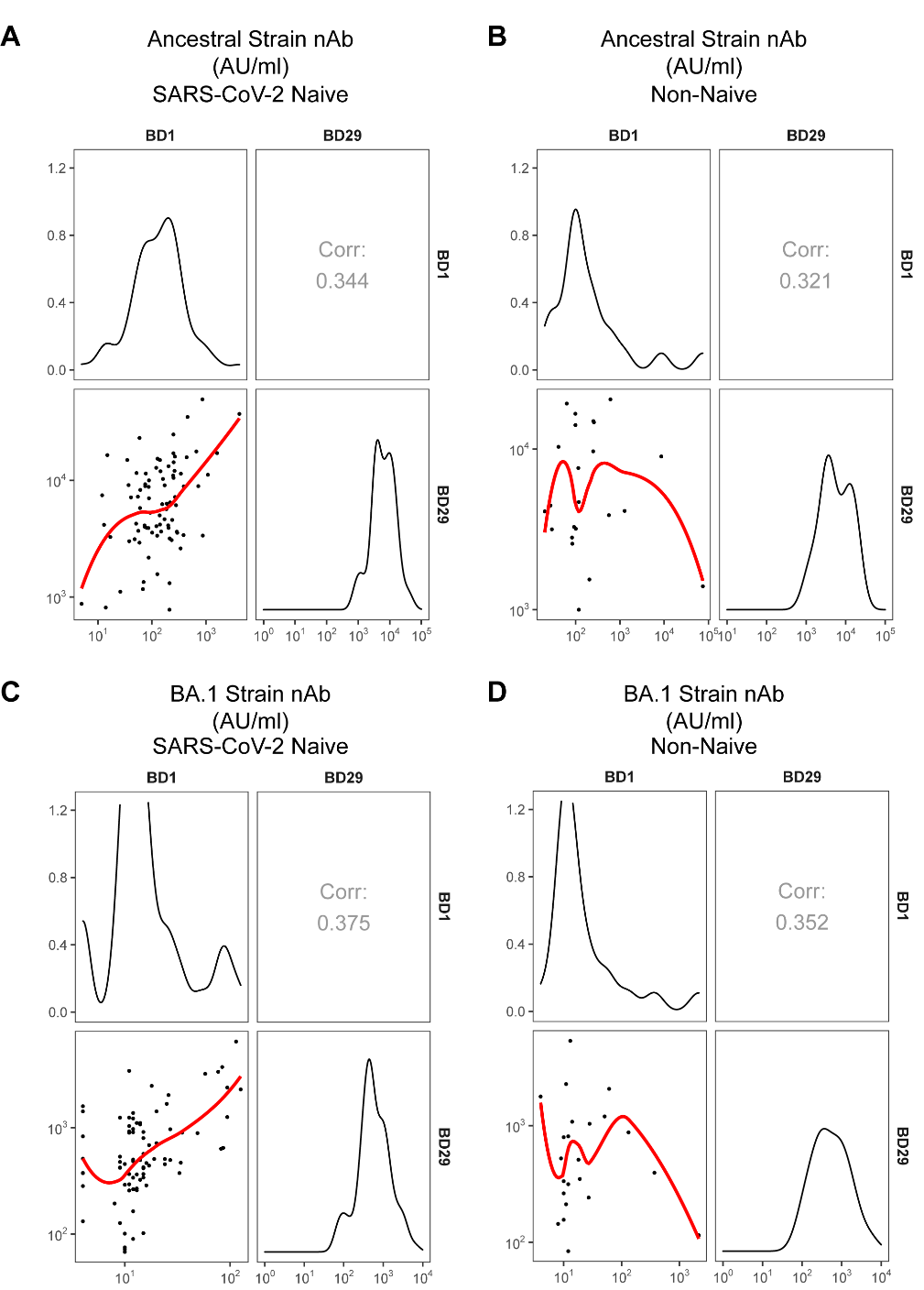

### Figure S12. Correlations between BD1 and BD29 (A, B) Ancestral strain neutralizing antibody (nAb) and (C, D) BA.1 strain nAb titers among (A, C) SARS-CoV-2 naives and (B, D) non-naives in the per-protocol boosted cohort. Corr = Inverse probability weight adjusted Spearman's rank correlation coefficient.

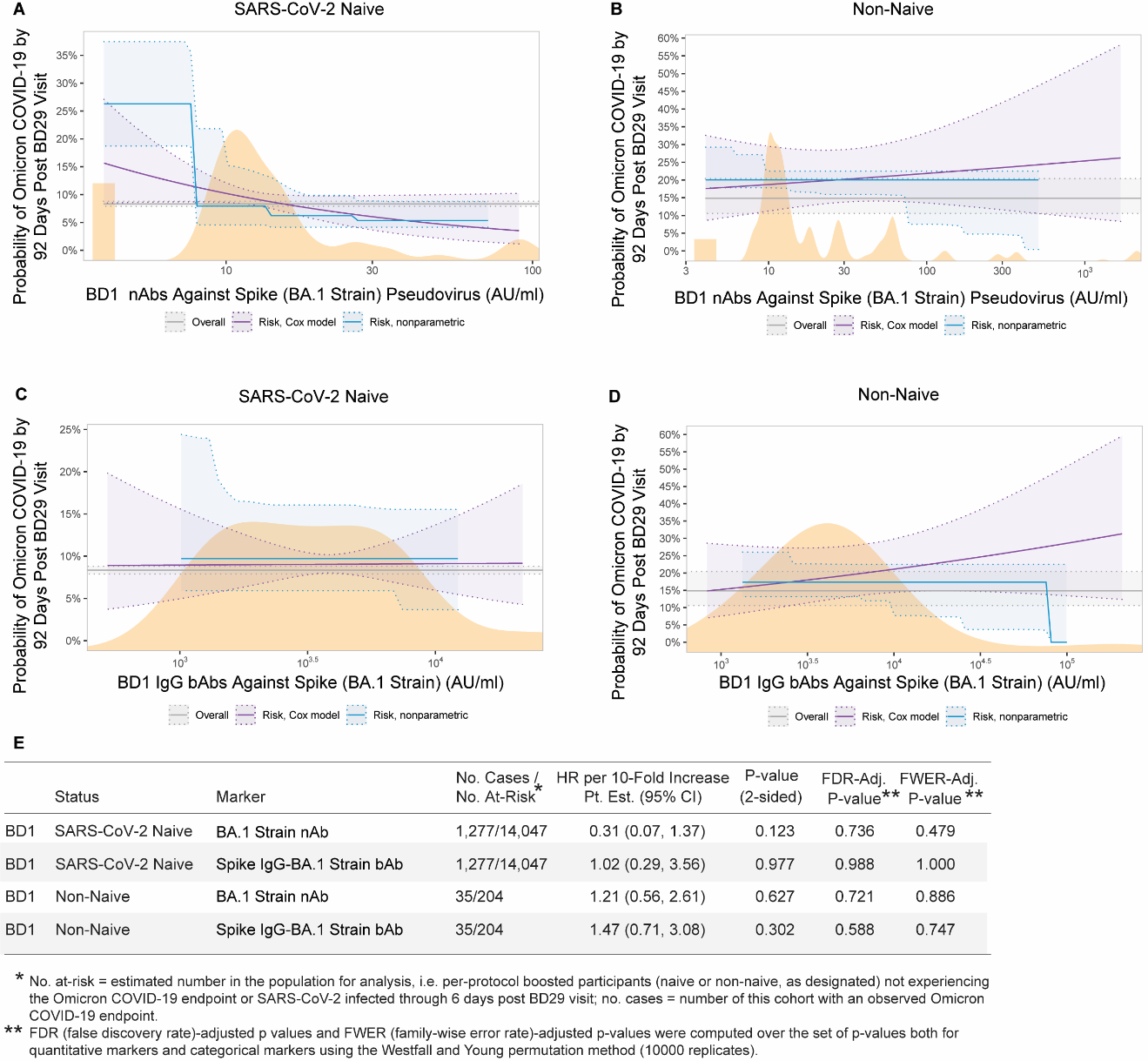

### Figure S13. Analyses of BD1 BA.1 strain neutralizing antibody (nAb) titer and Spike IgG-BA.1 strain binding antibody (bAb) concentration as a correlate of risk of Omicron COVID-19. Curves show cumulative incidence of Omicron COVID-19, estimated using a Cox model (purple) or a nonparametric method (blue), in per-protocol boosted (A, C) SARS-CoV-2 naives and (B, D) non-naives by 92 days post BD29 by BD1 antibody marker level. The dotted lines indicate bootstrap pointwise 95% CIs. The horizontal gray line is the overall cumulative incidence of Omicron COVID-19 from 7 to 92 days post BD29 in the per-protocol boosted SARS-CoV-2 naive or non-naive population, as designated. The distribution of the marker in the respective analysis population, calculated by kernel density estimation, is plotted in orange. E) Hazard ratios of Omicron COVID-19 per 10-fold increase in each BD1 BA.1 strain marker in per-protocol boosted SARS-CoV-2 naives or non-naives. Baseline covariates adjusted for: baseline risk score, at risk status, and community of color status.

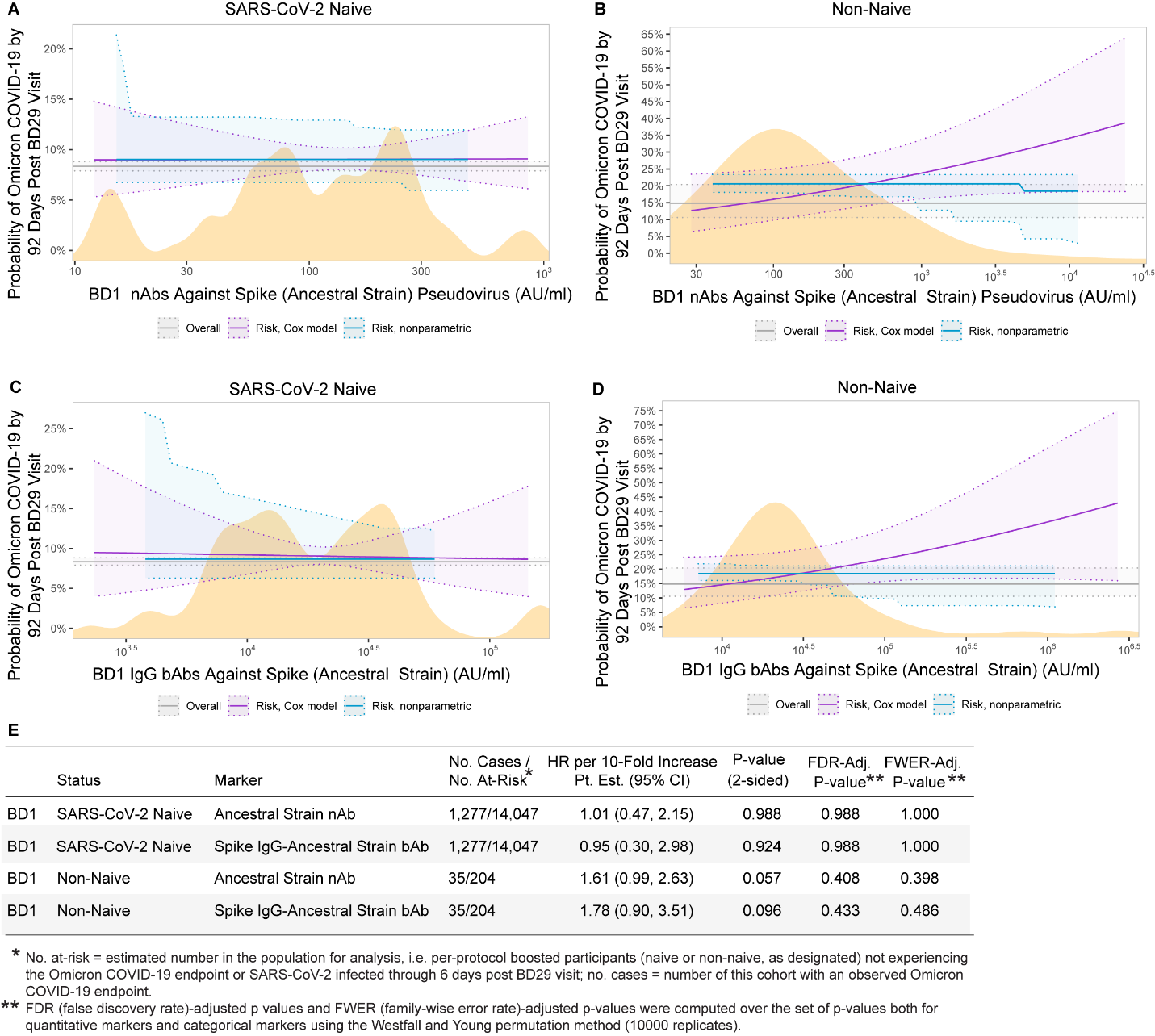

### Figure S14. Analyses of BD1 Ancestral strain neutralizing antibody (nAb) titer and Spike IgG-Ancestral strain binding antibody (bAb) concentration as a correlate of risk of Omicron COVID-19. Curves show cumulative incidence of Omicron COVID-19, estimated using a Cox model (purple) or a nonparametric method (blue), in per-protocol boosted (A, C) SARS-CoV-2 naives and (B, D) non-naives by 92 days post BD29 by BD1 antibody marker level. The dotted lines indicate bootstrap pointwise 95% CIs. The horizontal gray line is the overall cumulative incidence of Omicron COVID-19 from 7 to 92 days post BD29 in the per-protocol boosted SARS-CoV-2 naive or non-naive population, as designated. The distribution of the marker in the respective analysis population, calculated by kernel density estimation, is plotted in orange. E) Hazard ratios of Omicron COVID-19 per 10-fold increase in each BD1 Ancestral strain marker in per-protocol boosted SARS-CoV-2 naives or non-naives. Baseline covariates adjusted for: baseline risk score, at risk status, and community of color status.

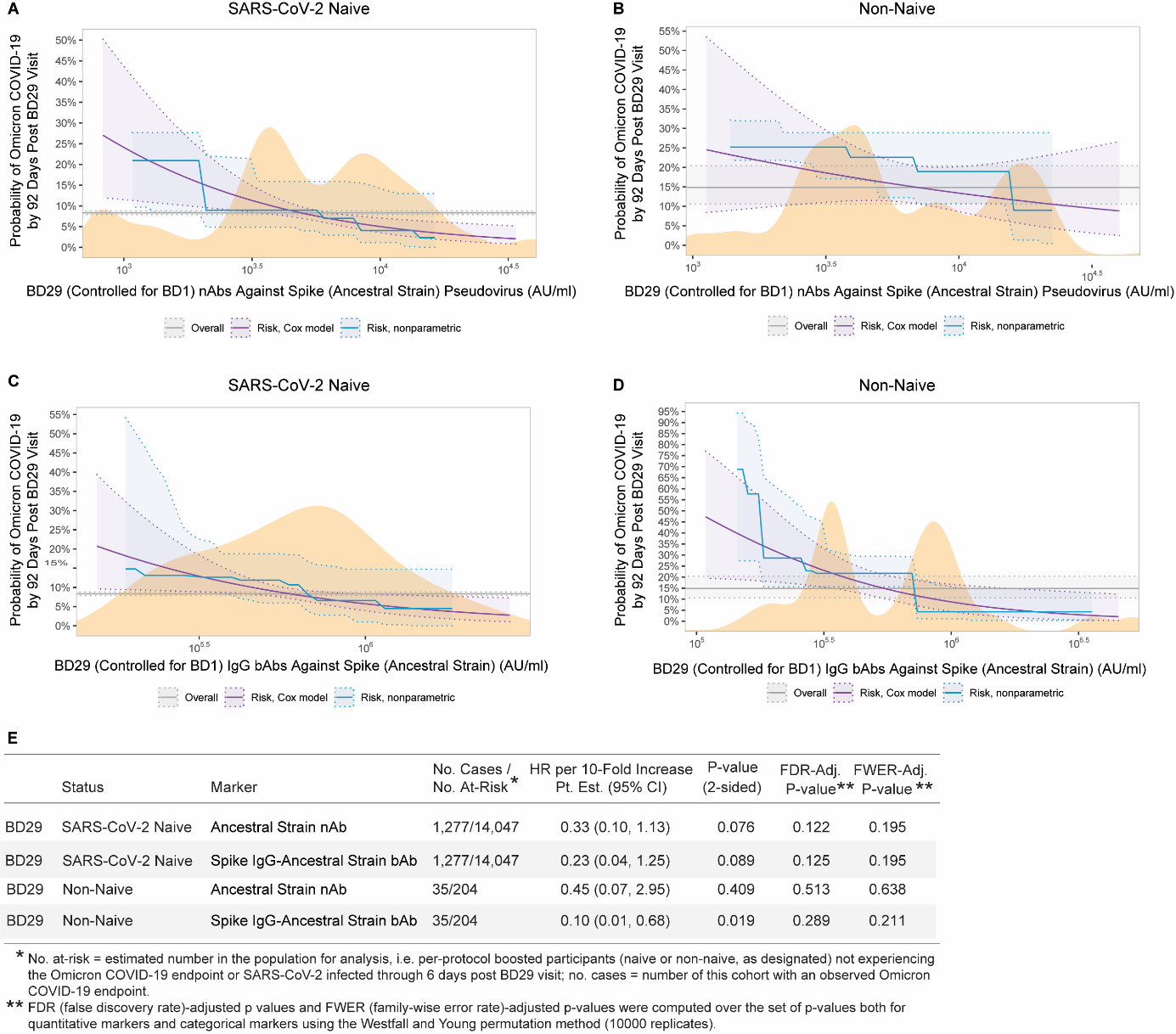

### Figure S15. Analyses of BD29 Ancestral strain neutralizing antibody (nAb) titer and Spike IgG-Ancestral strain binding antibody (bAb) concentration as a correlate of risk of Omicron COVID-19. Curves show cumulative incidence of Omicron COVID-19, estimated using a Cox model (purple) or a nonparametric method (blue), in per-protocol boosted (A, C) SARS-CoV-2 naives and (B, D) non-naives by 92 days post BD29 by BD29 antibody marker level. BD29 marker levels were controlled for BD1 marker levels. The dotted lines indicate bootstrap pointwise 95% CIs. The horizontal gray line is the overall cumulative incidence of Omicron COVID-19 from 7 to 92 days post BD29 in the per-protocol boosted SARS-CoV-2 naive or non-naive population, as designated. The distribution of the marker in the respective analysis population, calculated by kernel density estimation, is plotted in orange. E) Hazard ratios of Omicron COVID-19 per 10-fold increase in each BD29 Ancestral marker in per-protocol boosted SARS-CoV-2 naives or non-naives. Baseline covariates adjusted for: baseline risk score, at risk status, and community of color status.

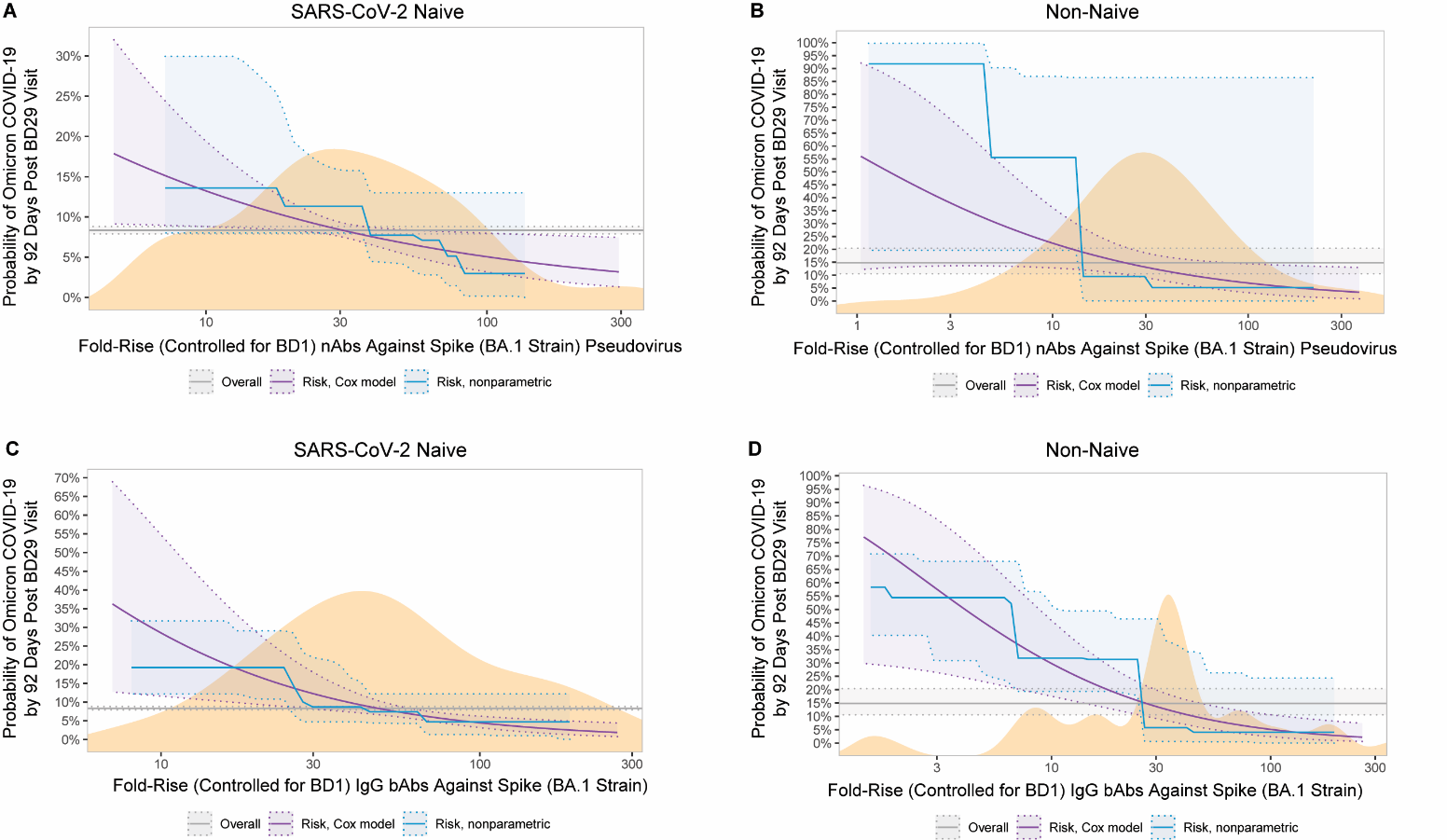

### Figure S16. Analyses of fold-rise (BD29/BD1) BA.1 strain neutralizing antibody (nAb) titer and Spike IgG-BA.1 strain binding antibody (bAb) concentration as a correlate of risk of Omicron COVID-19. Curves show cumulative incidence of Omicron COVID-19, estimated using a Cox model (purple) or a nonparametric method (blue), in per-protocol boosted (A, C) SARS-CoV-2 naives and (B, D) non-naives by 92 days post BD29 by BD29/BD29 antibody marker level. BD29 marker levels were controlled for BD1 marker levels. The dotted lines indicate bootstrap pointwise 95% CIs. The horizontal gray line is the overall cumulative incidence of Omicron COVID-19 from 7 to 92 days post BD29 in the per-protocol boosted SARS-CoV-2 naive or non-naive population, as designated. The distribution of the marker in the respective analysis population, calculated by kernel density estimation, is plotted in orange. Baseline covariates adjusted for: baseline risk score, at risk status, and community of color status.

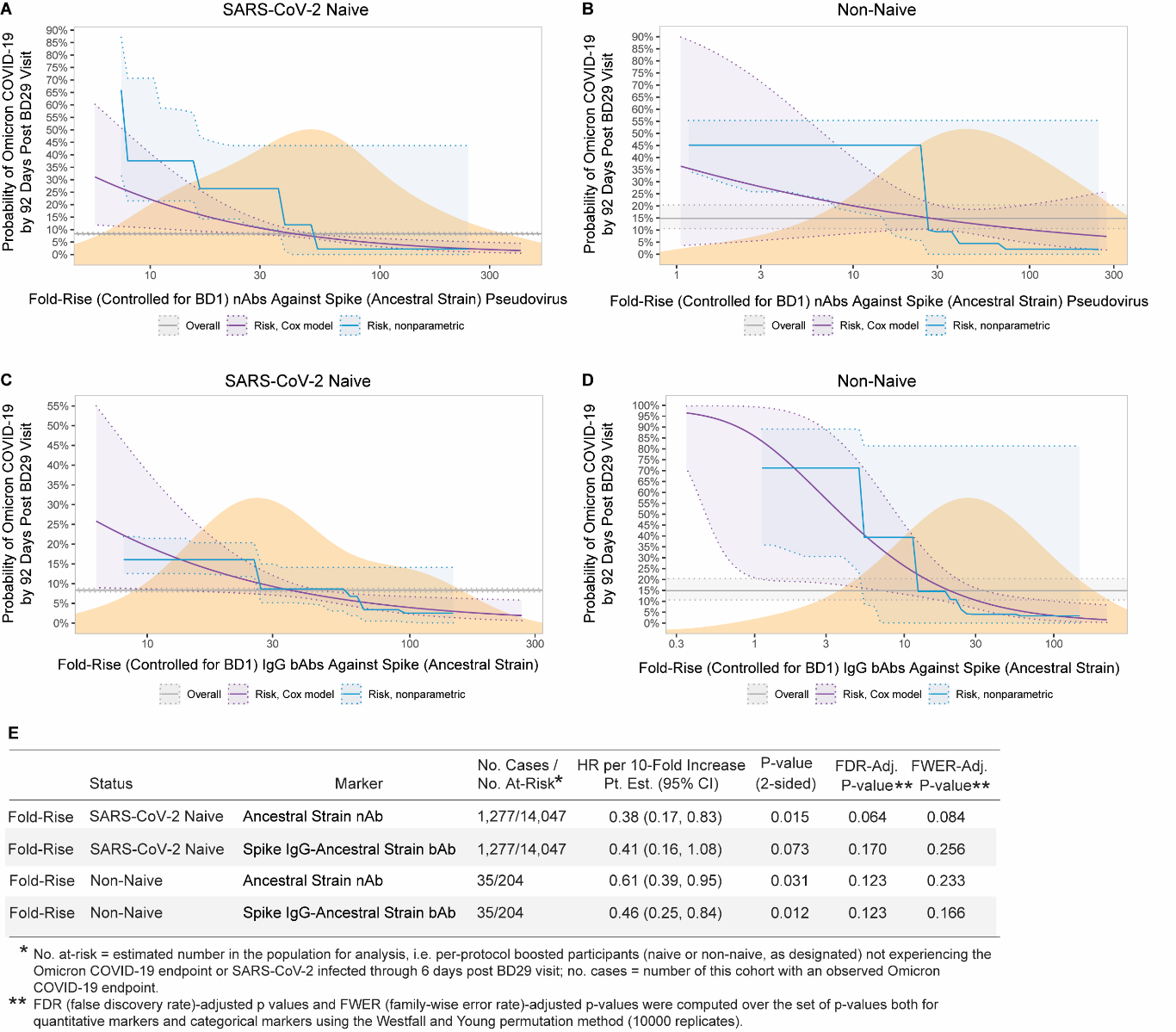

### Figure S17. Analyses of fold-rise (BD29/BD1) Ancestral strain neutralizing antibody (nAb) titer and Spike IgG-Ancestral strain binding antibody (bAb) concentration as a correlate of risk of Omicron COVID-19. Curves show cumulative incidence of Omicron COVID-19, estimated using a Cox model (purple) or a nonparametric method (blue), in per-protocol boosted (A, C) SARS-CoV-2 naives and (B, D) non-naives by 92 days post BD29 by BD29/BD1 fold-rise. BD29 marker levels were controlled for BD1 marker levels. The dotted lines indicate bootstrap pointwise 95% CIs. The horizontal gray line is the overall cumulative incidence of Omicron COVID-19 from 7 to 92 days post BD29 in the per-protocol boosted SARS-CoV-2 naive or non-naive population, as designated. The distribution of the marker in the respective analysis population, calculated by kernel density estimation, is plotted in orange. E) Hazard ratios of Omicron COVID-19 per 10-fold increase in each fold-rise (BD29/BD1) Ancestral marker in per-protocol boosted SARS-CoV-2 naives or non-naives. Baseline covariates adjusted for: baseline risk score, at risk status, and community of color status.

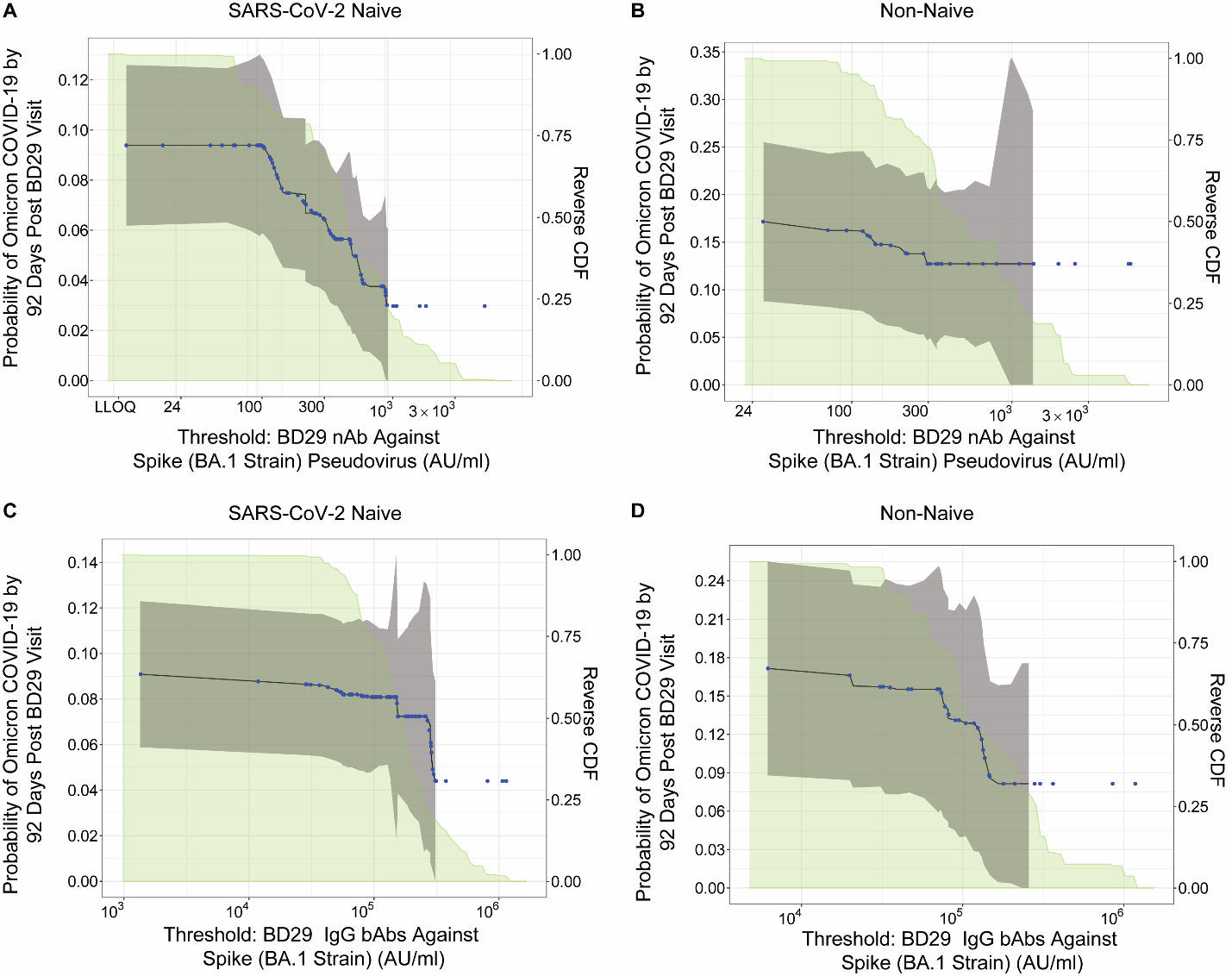

Figure S18. Cumulative incidence of Omicron COVID-19 by 92 days post BD29 by per-protocol boosted subgroups of (A, C) SARS-CoV-2 naives and (B, D) non-naives defined by (A, B) BD29 BA.1 strain neutralizing antibody (nAb) titer or (C, D) BD29 Spike IgG-BA.1 strain binding antibody (bAb) concentration above a threshold. The reverse cumulative distribution function (CDF) of each antibody marker is overlaid in green. Estimates and confidence intervals were adjusted using the assumption that the true threshold-response is nonincreasing. The blue dots correspond to marker values where an event is observed. The gray shaded area is pointwise 95% CIs.

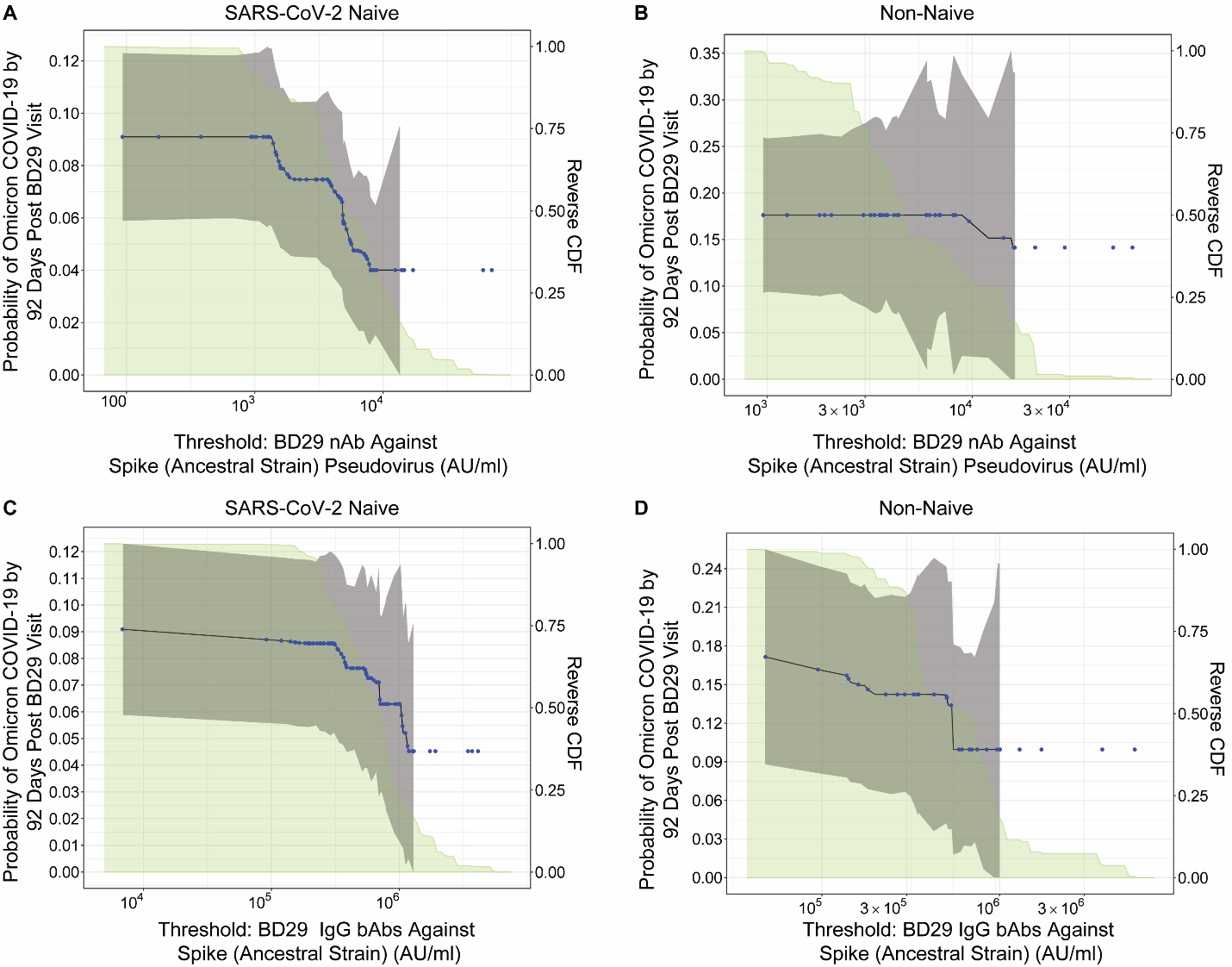
Figure S19. Cumulative incidence of Omicron COVID-19 by 92 days post BD29 by per-protocol boosted subgroups of (A, C) SARS-CoV-2 naives and (B, D) non-naives defined by (A, B) BD29 Ancestral strain neutralizing antibody (nAb) titer or (C, D) BD29 Spike IgG-Ancestral strain binding antibody (bAb) concentration above a threshold. The reverse cumulative distribution function (CDF) of each antibody marker is overlaid in green. Estimates and confidence intervals were adjusted using the assumption that the true threshold-response is nonincreasing. The blue dots correspond to marker values where an event is observed. The gray shaded area is pointwise 95% CIs.

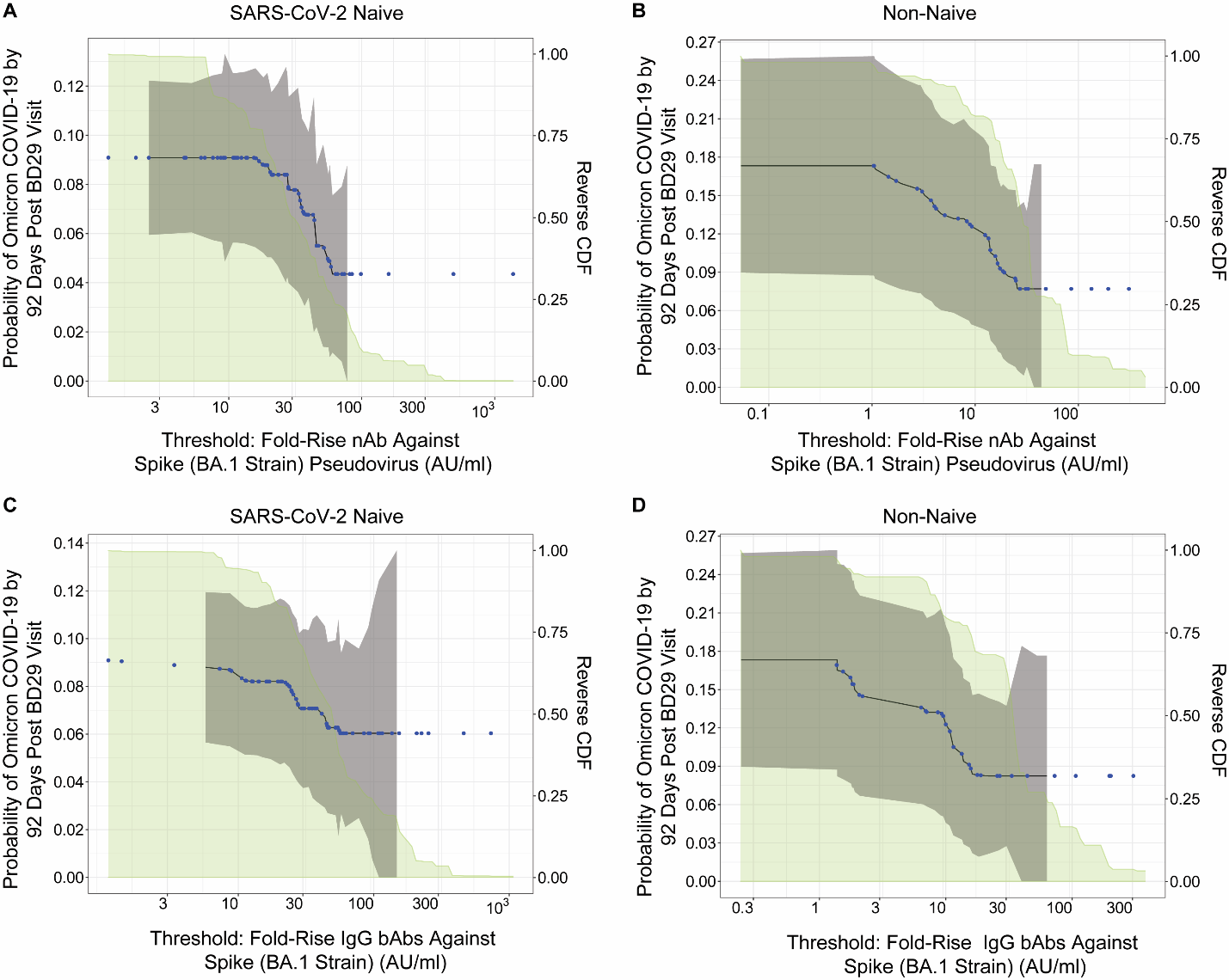
Figure S20. Cumulative incidence of Omicron COVID-19 by 92 days post BD29 by per-protocol boosted subgroups of (A, C) SARS-CoV-2 naives and (B, D) non-naives defined by (A, B) Fold-rise (BD29/BD1) BA.1 strain neutralizing antibody (nAb) titer or (C, D) Fold-rise (BD29/BD1) Spike IgG-BA.1 strain binding antibody (bAb) concentration above a threshold. The reverse cumulative distribution function (CDF) of each antibody marker is overlaid in green. Estimates and confidence intervals were adjusted using the assumption that the true threshold-response is nonincreasing. The blue dots correspond to marker values where an event is observed. The gray shaded area is pointwise 95% CIs.

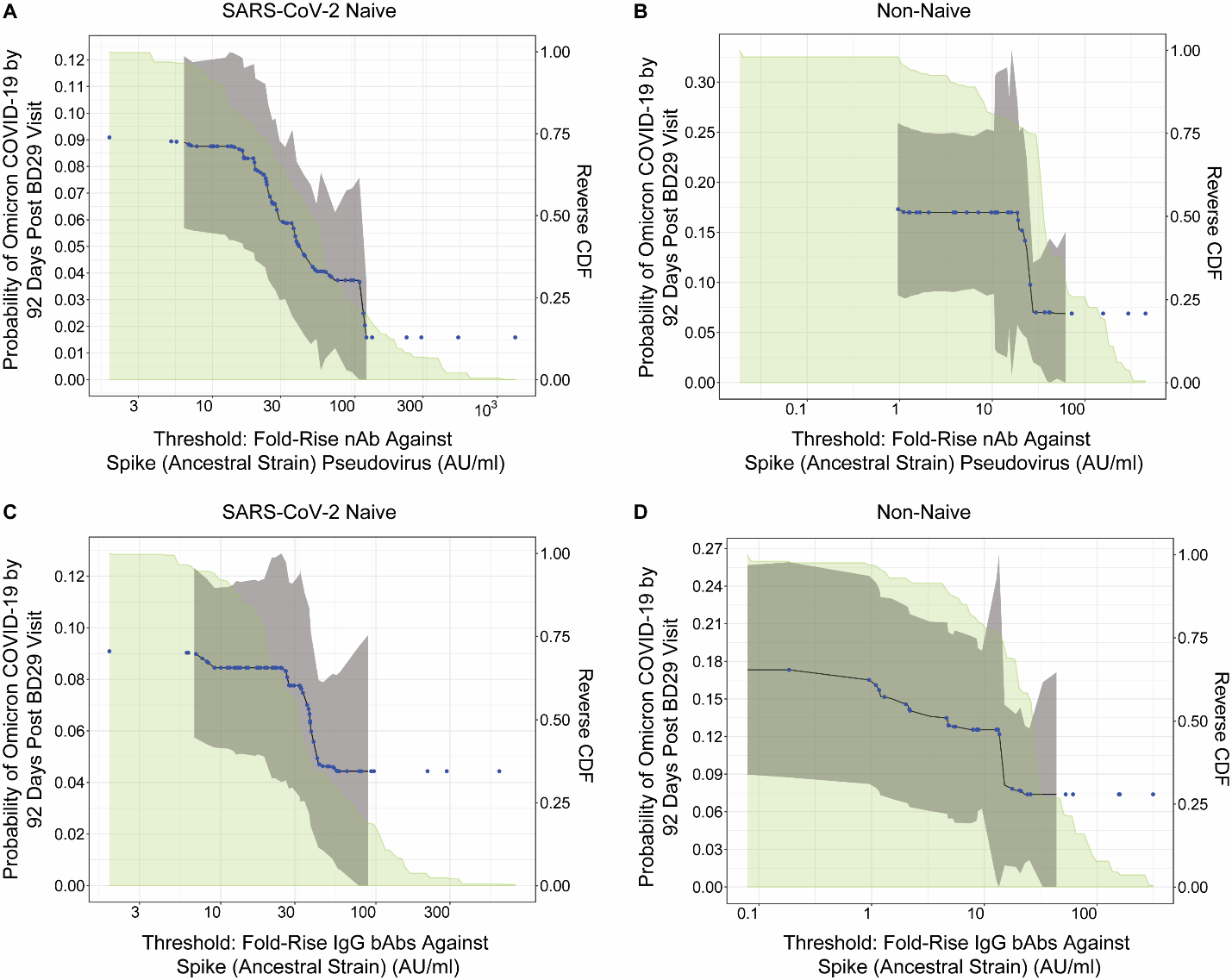
Figure S21. Cumulative incidence of Omicron COVID-19 by 92 days post BD29 by per-protocol boosted subgroups of (A, C) SARS-CoV-2 naives and (B, D) non-naives defined by (A, B) Fold-rise (BD29/BD1) Ancestral strain neutralizing antibody (nAb) titer or (C, D) Fold-rise (BD29/BD1) Spike IgG-Ancestral strain binding antibody (bAb) concentration above a threshold. The reverse cumulative distribution function (CDF) of each antibody marker is overlaid in green. Estimates and confidence intervals were adjusted using the assumption that the true threshold-response is nonincreasing. The blue dots correspond to marker values where an event is observed. The gray shaded area is pointwise 95% CIs.

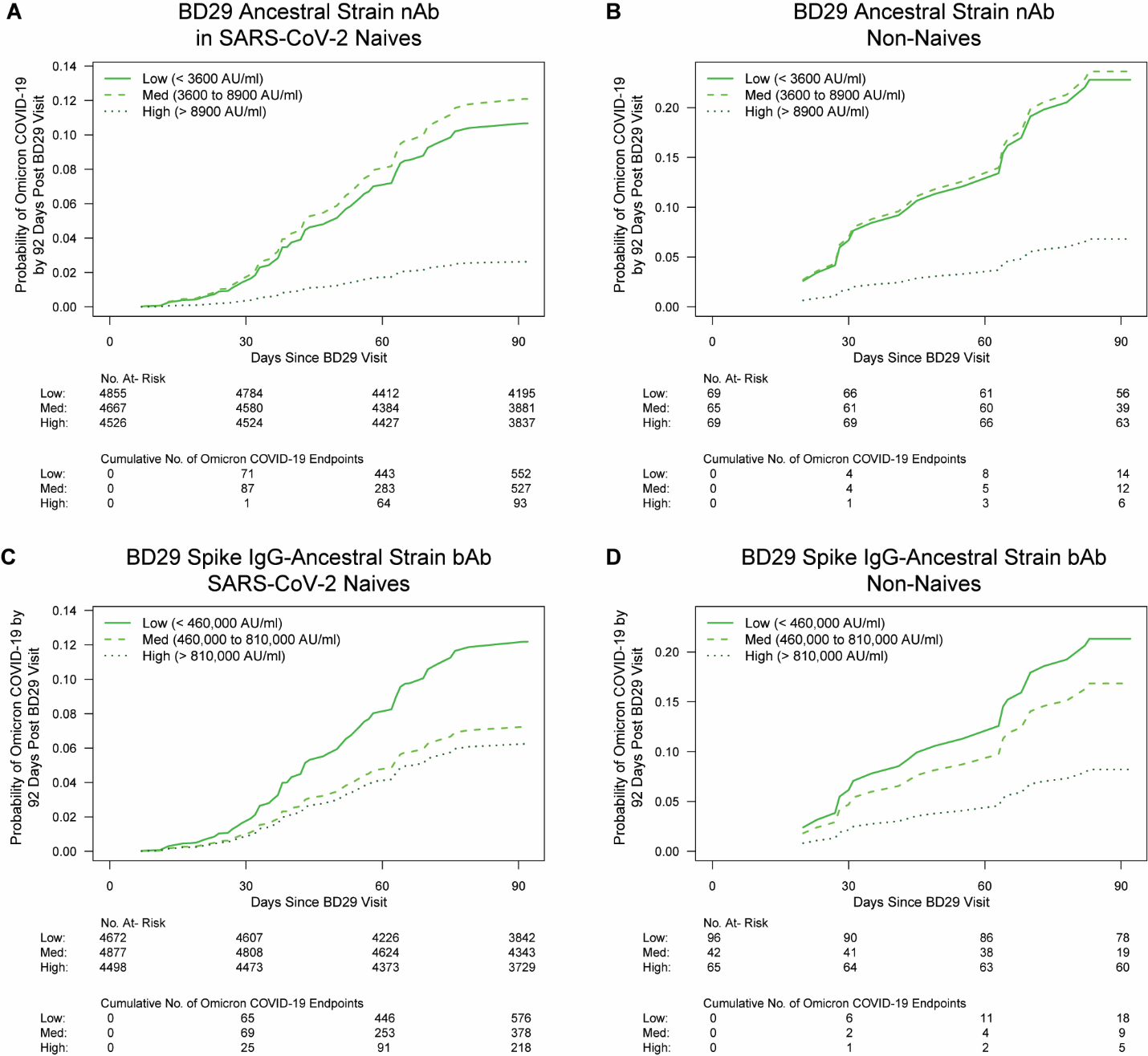

### Figure S22. Cox-model-based marginalized Omicron COVID-19 cumulative incidence curves for subgroups of per-protocol boosted (A, C) SARS-CoV-2 naive or (B, D) non-naive participants defined by BD29 Ancestral strain antibody tertile. A, B: BD29 Ancestral strain neutralizing antibody (nAb); C, D: BD29 Spike IgG-Ancestral strain binding antibody (bAb). No. at risk = estimated number in the population for analysis, i.e. per-protocol (A, C) SARS-CoV-2 naive or (B, D) non-naive boosted participants not experiencing the Omicron COVID-19 endpoint or SARS-CoV-2 infected through 6 days post BD29 visit. Analyses were adjusted for baseline risk score, at-risk status, and community of color status.

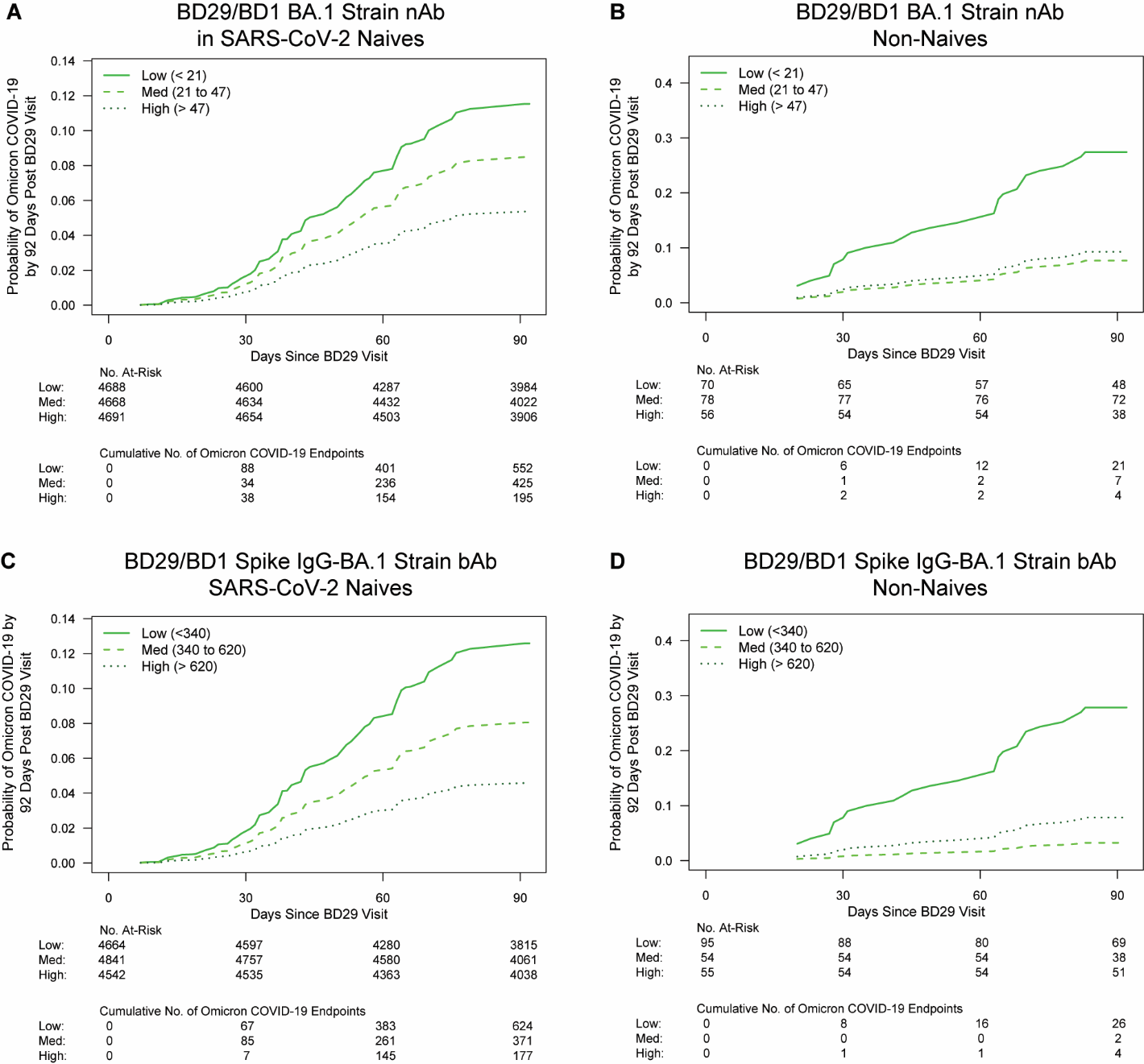

### Figure S23. Cox-model-based marginalized Omicron COVID-19 cumulative incidence curves for subgroups of per-protocol boosted (A, C) SARS-CoV-2 naive or (B, D) non-naive participants defined by fold-rise (BD29/BD1) BA.1 strain antibody tertile. A, B: Fold-rise BA.1 strain neutralizing antibody (nAb); C, D: Fold-rise Spike IgG-BA.1 strain binding antibody (bAb). No. at risk = estimated number in the population for analysis, i.e. per-protocol (A, C) SARS-CoV-2 naive or (B, D) non-naive boosted participants not experiencing the Omicron COVID-19 endpoint or SARS-CoV-2 infected through 6 days post BD29 visit. Analyses were adjusted for baseline risk score, at-risk status, and community of color status.

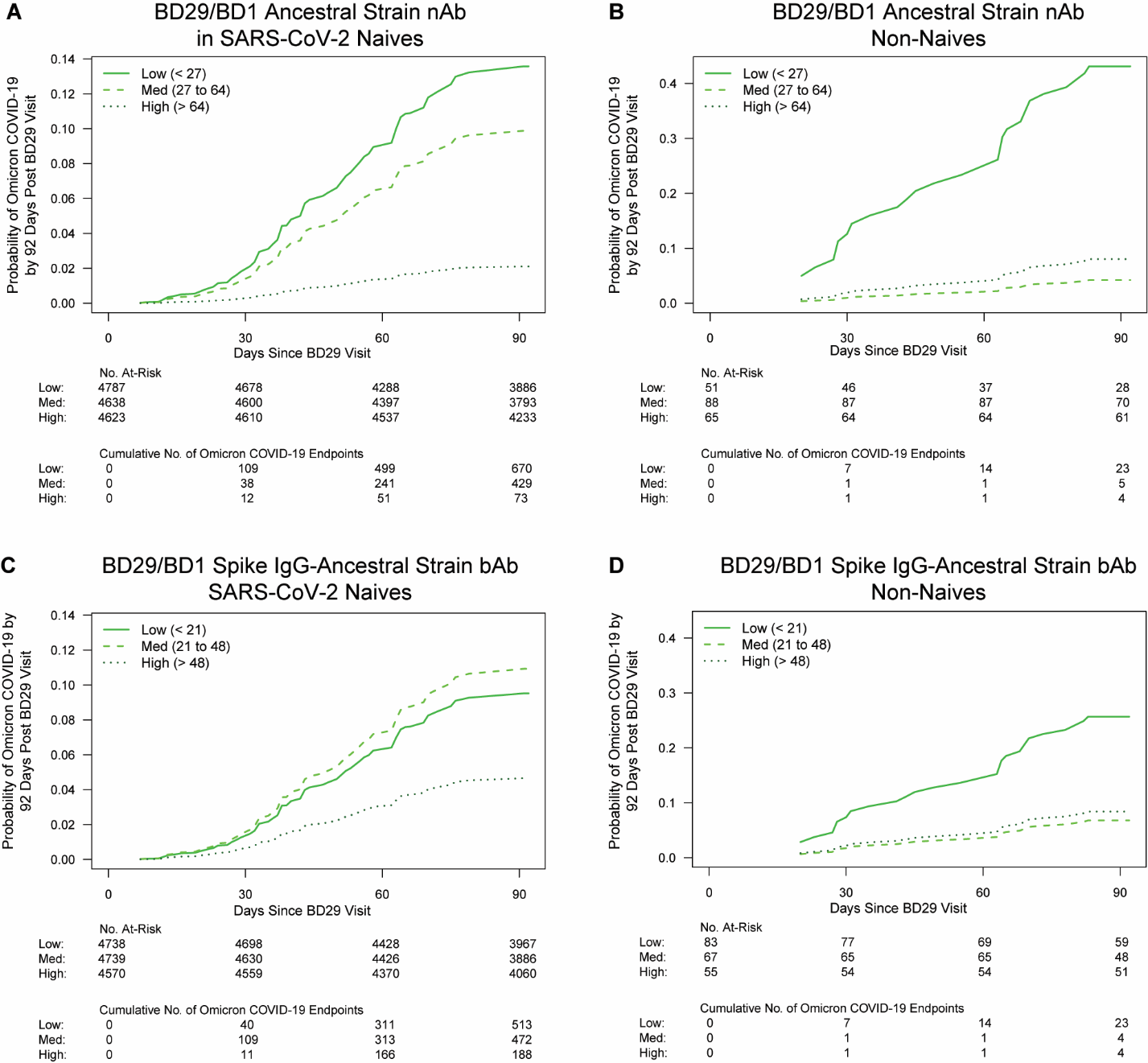

### Figure S24. Cox-model-based marginalized Omicron COVID-19 cumulative incidence curves for subgroups of per-protocol boosted (A, C) SARS-CoV-2 naive or (B, D) non-naive participants defined by fold-rise (BD29/BD1) Ancestral strain antibody tertile. A, B: Fold-rise Ancestral strain neutralizing antibody (nAb); C, D: Fold-rise Spike IgG-Ancestral strain binding antibody (bAb). No. at risk = estimated number in the population for analysis, i.e. per-protocol (A, C) SARS-CoV-2 naive or (B, D) non-naive boosted participants not experiencing the Omicron COVID-19 endpoint or SARS-CoV-2 infected through 6 days post BD29 visit. Analyses were adjusted for baseline risk score, at-risk status, and community of color status.

### Table S7. Estimated hazard ratios of Omicron COVID-19 for the medium versus low and for the high versus low tertiles of the designated BD1 BA.1 strain and Ancestral strain markers, in SARS-CoV-2 naives and in non-naives. Comparisons were made in per-protocol boosted participants. N/A, not applicable.

| **BD1 Marker** | **Tertile*** | **No. cases/**  **No. at-risk**** | **Attack rate** | **Hazard Ratio Pt. Est.** | **Hazard Ratio 95% CI** | ***P* value (2-sided)** | **Overall *P* value^¶^** | **FDR-adjusted *P* value^†^** | **FWER-adjusted *P* value^†^** |
| --- | --- | --- | --- | --- | --- | --- | --- | --- | --- |
| **A) SARS-CoV-2 Naive** |  |  |  |  |  |  |  |  |  |
| Spike IgG-BA.1 strain bAbs | Low | 342/4,734 | 0.0722 | 1 | N/A | N/A | 0.760 | 0.988 | 0.998 |
|  | Med | 515/4,681 | 0.1100 | 1.44 | (0.46, 4.47) | 0.529 |  |  |  |
|  | High | 419/4,632 | 0.0905 | 1.11 | (0.39, 3.18) | 0.850 |  |  |  |
| BA.1 Strain nAbs | Low | 667/6,196 | 0.1077 | 1 | N/A | N/A | 0.584 | 0.967 | 0.979 |
|  | Med | 361/4,267 | 0.0846 | 0.72 | (0.30, 1.77) | 0.479 |  |  |  |
|  | High | 249/3,584 | 0.0695 | 0.60 | (0.21, 1.68) | 0.328 |  |  |  |
| Spike IgG-Ancestral Strain bAbs | Low | 359/4,819 | 0.0745 | 1 | N/A | N/A | 0.775 | 0.988 | 0.998 |
|  | Med | 497/4,690 | 0.1060 | 1.42 | (0.50, 3.99) | 0.507 |  |  |  |
|  | High | 420/4,538 | 0.0926 | 1.14 | (0.43, 3.04) | 0.791 |  |  |  |
| Ancestral Strain nAbs | Low | 395/4,740 | 0.0833 | 1 | N/A | N/A | 0.954 | 0.988 | 1.000 |
|  | Med | 413/4,681 | 0.0882 | 0.97 | (0.38, 2.52) | 0.953 |  |  |  |
|  | High | 469/4,626 | 0.1014 | 1.10 | (0.40, 3.02) | 0.848 |  |  |  |
| **B) Non-Naive** |  |  |  |  |  |  |  |  |  |
| Spike IgG-BA.1 Strain bAbs | Low | 4/39 | 0.1026 | 1 | N/A | N/A | 0.720 | 0.755 | 0.886 |
|  | Med | 12/77 | 0.1558 | 1.57 | (0.27, 9.01) | 0.614 |  |  |  |
|  | High | 19/89 | 0.2135 | 1.95 | (0.37, 10.24) | 0.428 |  |  |  |
| BA.1 Strain nAbs | Low | 12/76 | 0.1579 | 1 | N/A | N/A | 0.462 | 0.662 | 0.847 |
|  | Med | 3/48 | 0.0625 | 0.48 | (0.05, 4.35) | 0.515 |  |  |  |
|  | High | 20/80 | 0.2500 | 1.70 | (0.51, 5.72) | 0.390 |  |  |  |
| Spike IgG-Ancestral Strain bAbs | Low | 3/27 | 0.1111 | 1 | N/A | N/A | 0.059 | 0.408 | 0.399 |
|  | Med | 10/115 | 0.0870 | 0.55 | (0.09, 3.23) | 0.507 |  |  |  |
|  | High | 22/62 | 0.3548 | 2.78 | (0.46, 16.71) | 0.263 |  |  |  |
| Ancestral Strain nAbs | Low | 4/41 | 0.0976 | 1 | N/A | N/A | 0.198 | 0.499 | 0.664 |
|  | Med | 9/87 | 0.1034 | 0.63 | (0.11, 3.71) | 0.613 |  |  |  |
|  | High | 22/76 | 0.2895 | 2.34 | (0.46, 1.73) | 0.303 |  |  |  |

Baseline covariates were adjusted for baseline risk score, at risk status, and community of color status, all defined identically as in Gilbert et al. (*4*).

*Antibody values defining the three tertiles were:

Spike IgG-BA.1 strain bAb: Low < 2000 AU/ml; Med 2000 to 5000 AU/ml; High > 5000 AU/ml.

BA.1 strain nAb: Low < 11 AU/ml; Med 11 to 15 AU/ml; High > 15 AU/ml.

Spike IgG-Ancestral strain bAb: Low < 12,000 AU/ml; Med 12,000 to 29,000 AU/ml; High > 29,000 AU/ml.

Ancestral strain nAb: Low < 73 AU/ml; Med 73 to 200 AU/ml; High > 200 AU/ml.

**No. at risk = estimated number in the population for analysis, i.e. per-protocol (A) SARS-CoV-2 naive and (B) non-naive boosted participants not experiencing the Omicron COVID-19 endpoint or SARS-CoV-2 infected through 6 days post BD29 visit; no. cases = number of this cohort with an observed Omicron COVID-19 endpoint.

^¶^The overall *P* value is from a generalized Wald test of whether the hazard rate of Omicron COVID-19 differed across the Low, Medium and High subgroups.

^†^*q*-value (false discovery rate, FDR) and family-wise error rate (FWER) were computed over the set of *P* values both for quantitative markers and categorical markers using the Westfall and Young permutation method (10,000 replicates).

### Table S8. Estimated hazard ratios of Omicron COVID-19 for the medium versus low and for the high versus low tertiles of the designated BD29 BA.1 strain and Ancestral strain markers, in SARS-CoV-2 naives and in non-naives. Comparisons were made in per-protocol boosted participants. N/A, not applicable.

| **BD29 Marker** | **Tertile*** | **No. cases/**  **No. at-risk**** | **Attack rate** | **Hazard Ratio Pt. Est.** | **Hazard Ratio 95% CI** | ***P* value (2-sided)** | **Overall *P* value^¶^** | **FDR-adjusted *P* value^†^** | **FWER-adjusted *P* value^†^** |
| --- | --- | --- | --- | --- | --- | --- | --- | --- | --- |
| **A) SARS-CoV-2 Naive** |  |  |  |  |  |  |  |  |  |
| Spike IgG-BA.1 Strain bAbs | Low | 693/4,801 | 0.1443 | 1 | N/A | N/A | 0.129 | 0.160 | 0.195 |
|  | Med | 255/4,692 | 0.0543 | 0.37 | (0.13, 1.02) | 0.056 |  |  |  |
|  | High | 330/4,554 | 0.0725 | 0.46 | (0.17, 1.26) | 0.131 |  |  |  |
| BA.1 Strain nAbs | Low | 699/4,677 | 0.1495 | 1 | N/A | N/A | 0.053 | 0.114 | 0.181 |
|  | Med | 411/4,859 | 0.0846 | 0.57 | (0.24, 1.38) | 0.217 |  |  |  |
|  | High | 167/4,511 | 0.0370 | 0.27 | (0.09, 0.78) | 0.016 |  |  |  |
| Spike IgG-Ancestral Strain bAbs | Low | 607/4,672 | 0.1299 | 1 | N/A | N/A | 0.403 | 0.420 | 0.418 |
|  | Med | 378/4,877 | 0.0775 | 0.58 | (0.21, 1.57) | 0.282 |  |  |  |
|  | High | 292/4,498 | 0.0649 | 0.49 | (0.17, 1.46) | 0.203 |  |  |  |
| Ancestral Strain nAbs | Low | 552/4,855 | 0.1137 | 1 | N/A | N/A | 0.018 | 0.114 | 0.101 |
|  | Med | 601/4,667 | 0.1288 | 1.14 | (0.47, 2.78) | 0.769 |  |  |  |
|  | High | 123/4,526 | 0.0272 | 0.24 | (0.07, 0.77) | 0.017 |  |  |  |
| **B) Non-Naive** |  |  |  |  |  |  |  |  |  |
| Spike IgG-BA.1 Strain bAbs | Low | 23/107 | 0.2150 | 1 | N/A | N/A | 0.145 | 0.380 | 0.539 |
|  | Med | 7/24 | 0.2917 | 0.94 | (0.28, 3.13) | 0.920 |  |  |  |
|  | High | 5/74 | 0.0676 | 0.24 | (0.06, 1.01) | 0.051 |  |  |  |
| BA.1 Strain nAbs | Low | 20/86 | 0.2326 | 1 | N/A | N/A | 0.260 | 0.434 | 0.638 |
|  | Med | 8/50 | 0.1600 | 0.49 | (0.11, 2.23) | 0.359 |  |  |  |
|  | High | 7/68 | 0.1029 | 0.32 | (0.07, 1.49) | 0.146 |  |  |  |
| Spike IgG-Ancestral Strain bAbs | Low | 19/96 | 0.1979 | 1 | N/A | N/A | 0.263 | 0.434 | 0.638 |
|  | Med | 9/42 | 0.2143 | 0.75 | (0.24, 2.39) | 0.629 |  |  |  |
|  | High | 7/65 | 0.1077 | 0.33 | (0.09, 1.24) | 0.102 |  |  |  |
| Ancestral Strain nAbs | Low | 14/69 | 0.2029 | 1 | N/A | N/A | 0.135 | 0.380 | 0.539 |
|  | Med | 13/65 | 0.2000 | 1.05 | (0.35, 3.15) | 0.933 |  |  |  |
|  | High | 8/69 | 0.1159 | 0.25 | (0.05, 1.29) | 0.098 |  |  |  |

Baseline covariates were adjusted for baseline risk score, at risk status, and community of color status, all defined identically as in Gilbert et al. (*4*).

*Antibody values defining the three tertiles are shown in Figures 2 and S22.

**No. at risk = estimated number in the population for analysis, i.e. per-protocol (A) SARS-CoV-2 naive and (B) non-naive boosted participants not experiencing the Omicron COVID-19 endpoint or SARS-CoV-2 infected through 6 days post BD29 visit; no. cases = number of this cohort with an observed Omicron COVID-19 endpoint.

^¶^The overall *P* value is from a generalized Wald test of whether the hazard rate of Omicron COVID-19 differed across the Low, Medium and High subgroups.

^†^*q*-value (false discovery rate, FDR) and family-wise error rate (FWER) were computed over the set of *P* values both for quantitative markers and categorical markers using the Westfall and Young permutation method (10,000 replicates).

### Table S9. Estimated hazard ratios of Omicron COVID-19 for the medium versus low and for the high versus low tertiles of the designated fold-rise (BD29/BD1) BA.1 strain and Ancestral strain markers, in SARS-CoV-2 naives and in non-naives. Comparisons were made in per-protocol boosted participants. N/A, not applicable.

| **BD29/BD1 Marker** | **Tertile*** | **No. cases/**  **No. at-risk**** | **Attack rate** | **Hazard Ratio Pt. Est.** | **Hazard Ratio 95% CI** | ***P* value (2-sided)** | **Overall *P* value^¶^** | **FDR-adjusted *P* value^†^** | **FWER-adjusted *P* value^†^** |
| --- | --- | --- | --- | --- | --- | --- | --- | --- | --- |
| **A) SARS-CoV-2 Naive** |  |  |  |  |  |  |  |  |  |
| Spike IgG-BA.1 Strain bAbs | Low | 653/4,664 | 0.1400 | 1 | N/A | N/A | 0.092 | 0.181 | 0.280 |
|  | Med | 417/4,841 | 0.0861 | 0.62 | (0.25, 1.55) | 0.310 |  |  |  |
|  | High | 207/4,542 | 0.0456 | 0.35 | (0.13, 0.90) | 0.030 |  |  |  |
| BA.1 Strain nAbs | Low | 581/4,688 | 0.1239 | 1 | N/A | N/A | 0.294 | 0.340 | 0.412 |
|  | Med | 456/4,668 | 0.0977 | 0.72 | (0.26, 1.98) | 0.529 |  |  |  |
|  | High | 240/4,691 | 0.0512 | 0.45 | (0.16, 1.23) | 0.119 |  |  |  |
| Spike IgG-Ancestral Strain bAbs | Low | 513/4,738 | 0.1083 | 1 | N/A | N/A | 0.178 | 0.251 | 0.412 |
|  | Med | 562/4,739 | 0.1186 | 1.16 | (0.47, 2.84) | 0.750 |  |  |  |
|  | High | 203/4,570 | 0.0444 | 0.48 | (0.18, 1.23) | 0.125 |  |  |  |
| Ancestral Strain nAbs | Low | 685/4,787 | 0.1431 | 1 | N/A | N/A | 0.002 | 0.039 | 0.021 |
|  | Med | 489/4,638 | 0.1054 | 0.71 | (0.32, 1.58) | 0.406 |  |  |  |
|  | High | 103/4,623 | 0.0223 | 0.15 | (0.05, 0.42) | <0.001 |  |  |  |
| **B) Non-Naive** |  |  |  |  |  |  |  |  |  |
| Spike IgG-BA.1 Strain bAbs | Low | 28/95 | 0.2947 | 1 | N/A | N/A | 0.018 | 0.123 | 0.194 |
|  | Med | 2/54 | 0.0370 | 0.09 | (0.01, 1.25) | 0.074 |  |  |  |
|  | High | 5/55 | 0.0909 | 0.23 | (0.07, 0.75) | 0.015 |  |  |  |
| BA.1 Strain nAbs | Low | 23/70 | 0.3286 | 1 | N/A | N/A | 0.037 | 0.123 | 0.236 |
|  | Med | 7/78 | 0.0897 | 0.24 | (0.07, 0.83) | 0.025 |  |  |  |
|  | High | 5/56 | 0.0893 | 0.29 | (0.07, 1.22) | 0.091 |  |  |  |
| Spike IgG-Ancestral Strain bAbs | Low | 26/83 | 0.3133 | 1 | N/A | N/A | 0.078 | 0.151 | 0.261 |
|  | Med | 4/67 | 0.0597 | 0.23 | (0.04, 1.21) | 0.082 |  |  |  |
|  | High | 5/55 | 0.0909 | 0.28 | (0.07, 1.08) | 0.064 |  |  |  |
| Ancestral Strain nAbs | Low | 25/51 | 0.4902 | 1 | N/A | N/A | <0.001 | 0.040 | 0.020 |
|  | Med | 5/88 | 0.0568 | 0.07 | (0.02, 0.30) | <0.001 |  |  |  |
|  | High | 5/65 | 0.0769 | 0.14 | (0.04, 0.45) | <0.001 |  |  |  |

Baseline covariates were adjusted for baseline risk score, at risk status, and community of color status, all defined identically as in Gilbert et al. (*4*).

*Antibody values defining the three tertiles are shown in Figures S23 and S24.

**No. at risk = estimated number in the population for analysis, i.e. per-protocol (A) SARS-CoV-2 naive and (B) non-naive boosted participants not experiencing the Omicron COVID-19 endpoint or SARS-CoV-2 infected through 6 days post BD29 visit; no. cases = number of this cohort with an observed Omicron COVID-19 endpoint.

^¶^The overall *P* value is from a generalized Wald test of whether the hazard rate of Omicron COVID-19 differed across the Low, Medium and High subgroups.

^†^*q*-value (false discovery rate, FDR) and family-wise error rate (FWER) were computed over the set of *P* values both for quantitative markers and categorical markers using the Westfall and Young permutation method (10,000 replicates).

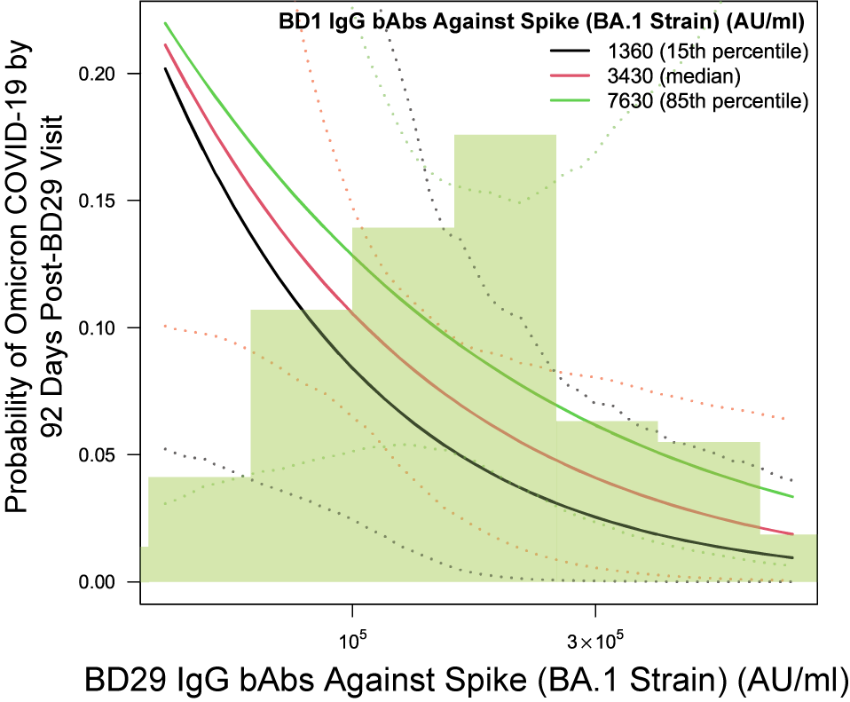

### Figure S25. Marginalized cumulative incidence curves of Omicron COVID-19 risk across a range of BD29 Spike IgG-BA.1 strain binding antibody (bAb) levels and within each tertile of BD1 Spike IgG-BA.1 strain bAb among SARS-CoV-2 naives. The Cox regression model adjusted for the minority indicator, heightened risk for severe COVID-19, predicted risk score, BD1 Spike IgG-BA.1 strain bAb and interaction between BD1 and BD29 Spike IgG-BA.1 strain bAb levels.

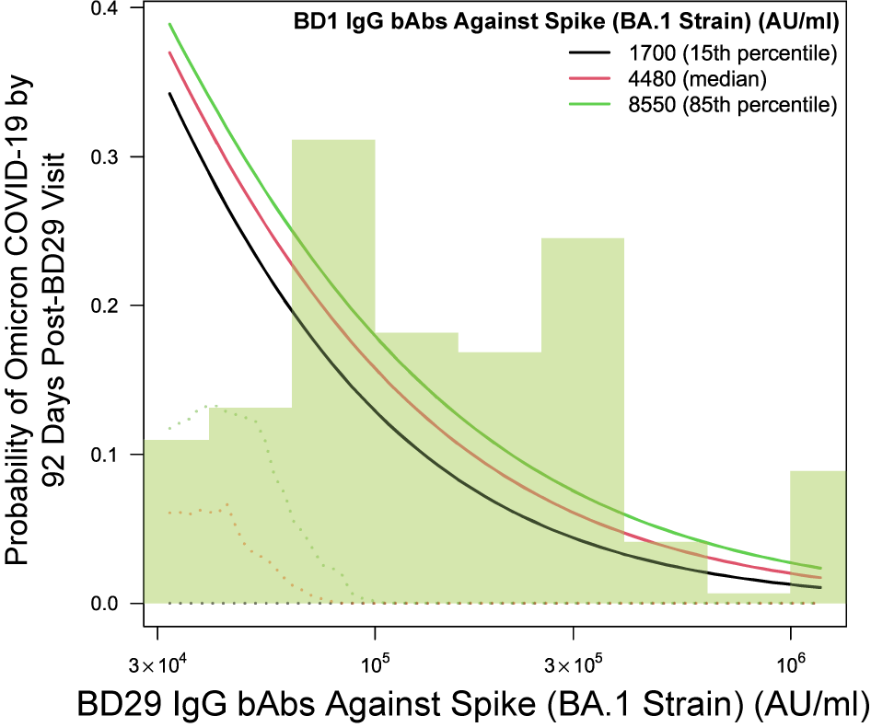

### Figure S26. Marginalized cumulative incidence curves of Omicron COVID-19 risk across a range of BD29 Spike IgG-BA.1 strain binding antibody (bAb) levels and within each tertile of BD1 Spike IgG-BA.1 strain bAb among non-naives. The Cox regression model adjusted for the minority indicator, heightened risk for severe COVID-19, predicted risk score, BD1 Spike IgG-BA.1 strain bAb and interaction between BD1 and BD29 Spike IgG-BA.1 strain bAb levels.

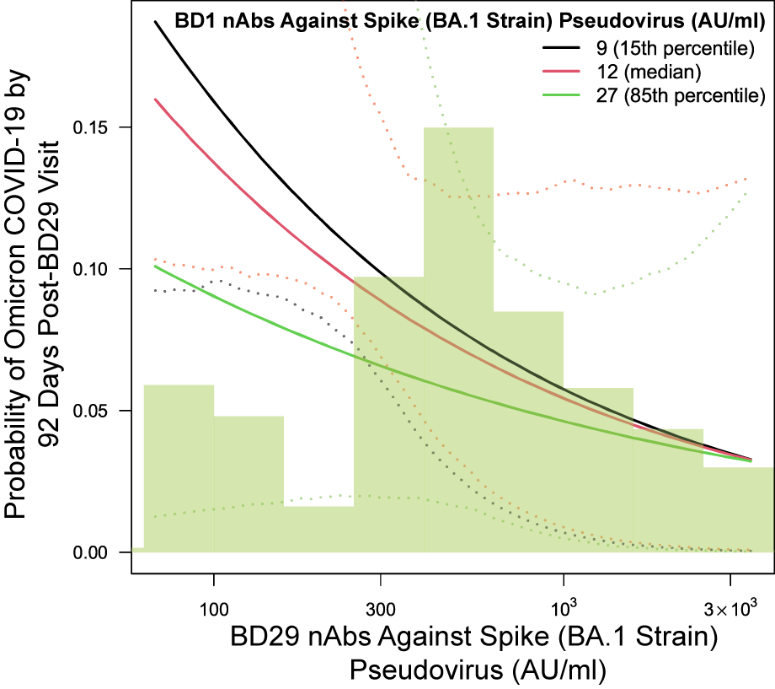

### Figure S27. Marginalized cumulative incidence curves of Omicron COVID-19 risk across a range of BD29 nAb BA.1 levels and within each tertile of BD1 BA.1 strain neutralizing antibody (nAb) level among SARS-CoV-2 naives. The Cox regression model adjusted for the minority indicator, heightened risk for severe COVID-19, predicted risk score, BD1 BA.1 strain nAb and interaction between BD1 and BD29 BA.1 strain nAb levels.

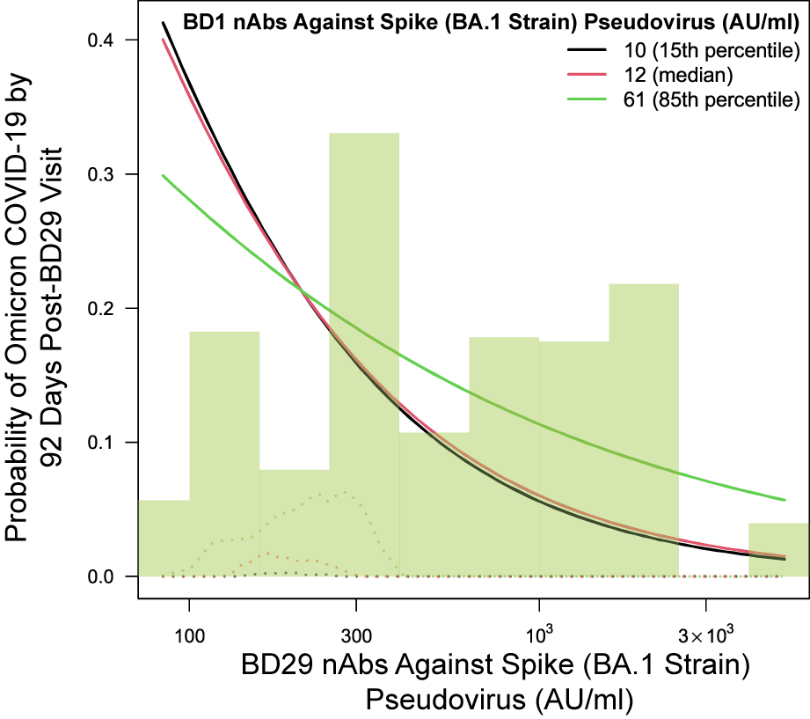

### Figure S28. Marginalized cumulative incidence curves of Omicron COVID-19 risk across a range of BD29 BA.1 strain neutralizing antibody (nAb) levels and within each tertile of BD1 BA.1 strain nAb among non-naives. The Cox regression model adjusted for the minority indicator, heightened risk for severe COVID-19, predicted risk score, BD1 BA.1 strain nAb and interaction between BD1 and BD29 BA.1 strain nAb levels.

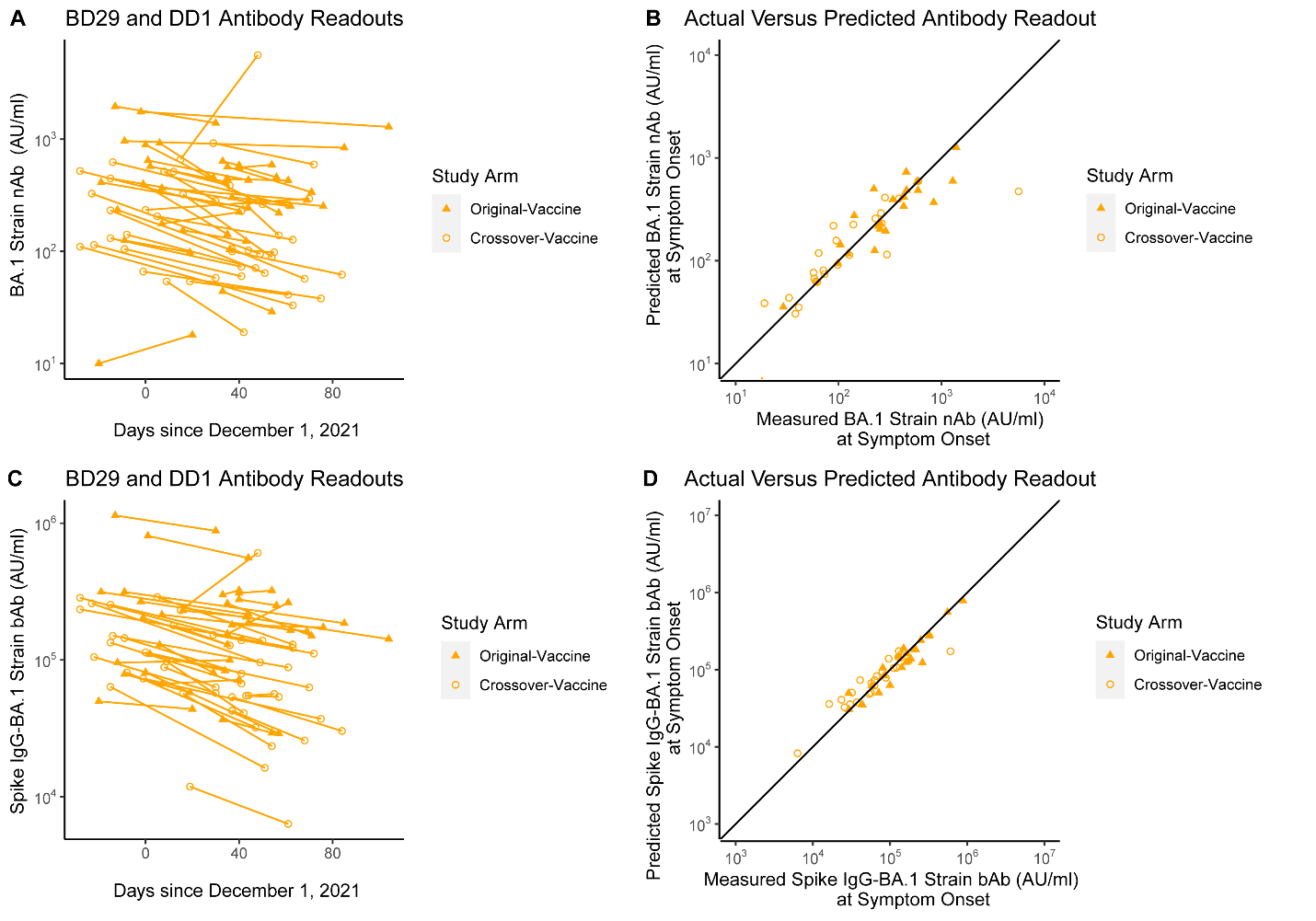
Figure S29. (A, C) BD29 and DD1 antibody levels and (B, D) predicted versus actual antibody levels at DD1 for (A, B) BA.1 strain neutralizing antibody (nAb) and (C, D) Spike IgG-BA.1 strain binding antibody (bAb) levels among SARS-CoV-2 naives. Filled orange triangles designate the original-vaccine arm; open orange circles designate the crossover-vaccine arm. The median slope for BA.1 strain nAbs was -0.0044 per day (half-life 68 days). The median slope for Spike IgG-BA.1 strain bAbs was -0.0034 per day (half-life 80 days).

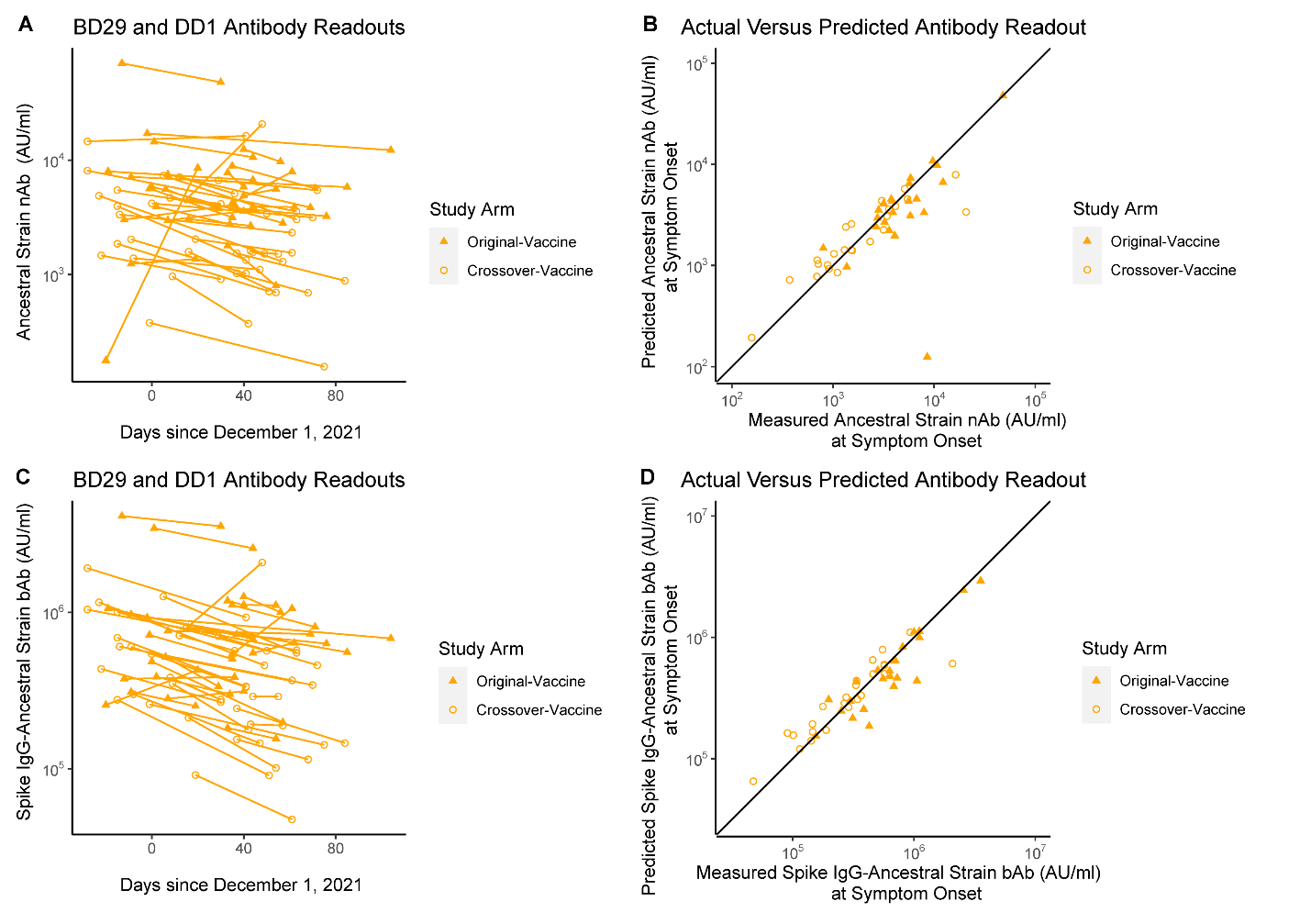

Figure S30. (A, C) BD29 and DD1 antibody levels and (B, D) predicted versus actual antibody levels at DD1 for (A, B) Ancestral strain neutralizing antibody (nAb) and (C, D) Spike IgG-Ancestral strain binding antibody (bAb) level among SARS-CoV-2 naives. Filled orange triangles designate the original-vaccine arm; open orange circles designate the crossover-vaccine arm. -0.0025. The median slope for Ancestral strain nAbs was -0.0039 per day (half-life 78 days). The median slope for Spike IgG-BA.1 strain bAbs was -0.0035 per day (half-life 86 days).

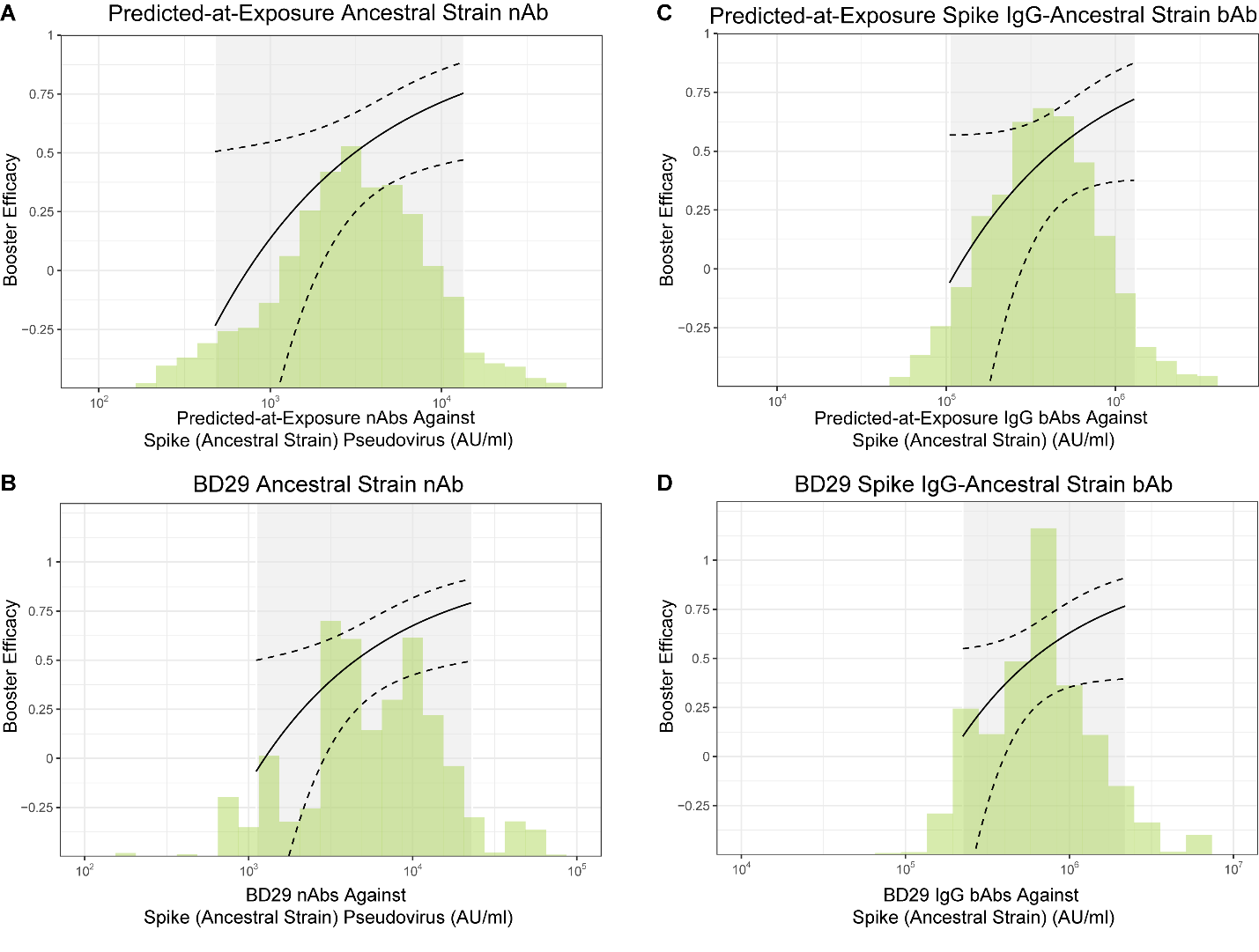

### Figure S31. Booster relative efficacy against Omicron COVID-19 among SARS-CoV-2 naives as a function of the predicted antibody level [A: Ancestral strain neutralizing antibody (nAb), B: Spike IgG-Ancestral strain binding antibody (bAb)] at the time of exposure to SARS-CoV-2 and the measured BD29 antibody level (C: Ancestral strain nAb, D: Spike IgG-Ancestral strain bAb).

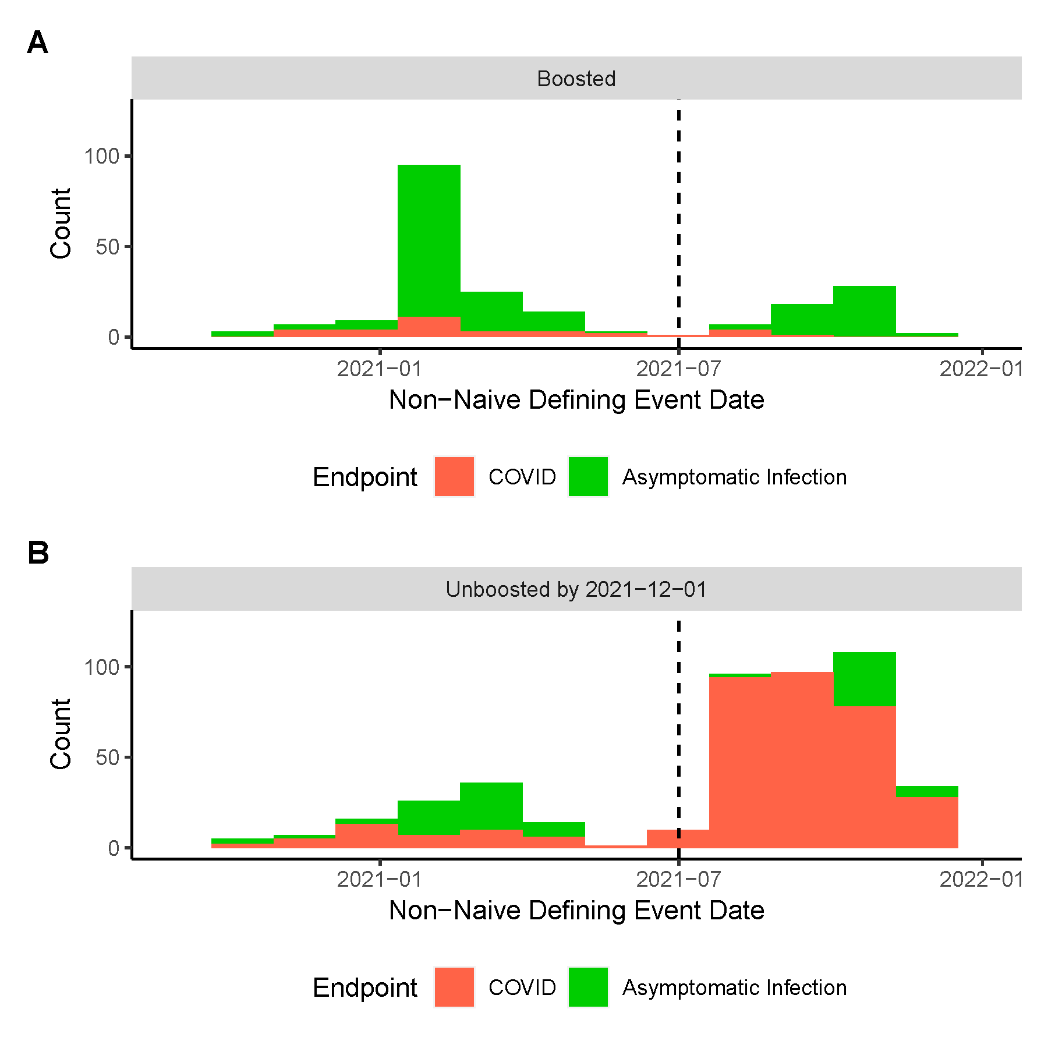

### Figure S32. Distribution of the day of the non-naive defining event for (A) boosted and (B) unboosted participants.

### Table S10. Discrete Super Learner performance across all 92 variable sets sorted by cross-validated area under the ROC (CV-AUC) performance for predicting occurrence of Omicron COVID-19 in SARS-CoV-2 naive per-protocol boosted participants. bAb = binding antibody; BRF = baseline risk factors; fold-rise = BD29/BD1; nAb = neutralizing antibody.

| **Variable Set** | **CV-AUC (95% CI)** |
| --- | --- |
| BRF + BD29-bAb-Spike-BA.1 + BD29-bAb-Spike-Ancestral + BD29-bAb-RBD-Ancestral + BD29-nAb-BA.1 + BD29-nAb-Ancestral + Fold-rise-bAb-Spike-BA.1 + Fold-rise-bAb-Spike-Ancestral + Fold-rise-bAb-RBD-Ancestral + Fold-rise-nAb-BA.1 + Fold-rise-nAb-Ancestral | 0.686 [0.600, 0.761] |
| BRF + BD1-bAb-RBD-Ancestral + BD1-nAb-BA.1 + BD1-nAb-Ancestral + Fold-rise-bAb-RBD-Ancestral + Fold-rise-nAb-BA.1 + Fold-rise-nAb-Ancestral | 0.686 [0.600, 0.761] |
| BRF + BD1-bAb-Spike-BA.1 + BD1-bAb-Spike-Ancestral + BD1-nAb-BA.1 + BD1-nAb-Ancestral + BD29-bAb-Spike-BA.1 + BD29-bAb-Spike-Ancestral + BD29-nAb-BA.1 + BD29-nAb-Ancestral | 0.686 [0.600, 0.761] |
| BRF + BD29-bAb-Spike-BA.1 + BD29-bAb-Spike-Ancestral + BD29-nAb-BA.1 + BD29-nAb-Ancestral | 0.685 [0.599, 0.760] |
| BRF + BD1-bAb-Spike-BA.1 + BD1-bAb-Spike-Ancestral + BD1-bAb-RBD-Ancestral + BD1-nAb-BA.1 + BD1-nAb-Ancestral + BD29-bAb-Spike-BA.1 + BD29-bAb-Spike-Ancestral + BD29-bAb-RBD-Ancestral + BD29-nAb-BA.1 + BD29-nAb-Ancestral | 0.685 [0.599, 0.760] |
| BRF + BD1-bAb-Spike-BA.1 + BD1-bAb-Spike-Ancestral + BD1-bAb-RBD-Ancestral + BD1-nAb-BA.1 + BD1-nAb-Ancestral + BD29-bAb-Spike-BA.1 + BD29-bAb-Spike-Ancestral + BD29-bAb-RBD-Ancestral + BD29-nAb-BA.1 + BD29-nAb-Ancestral + Fold-rise-bAb-Spike-BA.1 + Fold-rise-bAb-Spike-Ancestral + Fold-rise-bAb-RBD-Ancestral + Fold-rise-nAb-BA.1 + Fold-rise-nAb-Ancestral | 0.684 [0.598, 0.760] |
| BRF + BD29-nAb-BA.1 + Fold-rise-nAb-BA.1 | 0.684 [0.598, 0.759] |
| BRF + BD1-bAb-RBD-Ancestral + BD1-nAb-BA.1 + BD1-nAb-Ancestral + BD29-bAb-RBD-Ancestral + BD29-nAb-BA.1 + BD29-nAb-Ancestral | 0.682 [0.596, 0.758] |
| BRF + BD29-nAb-BA.1 + BD29-nAb-Ancestral | 0.682 [0.597, 0.757] |
| BRF + BD29-nAb-BA.1 + BD29-nAb-Ancestral + Fold-rise-nAb-BA.1 + Fold-rise-nAb-Ancestral | 0.681 [0.595, 0.756] |
| BRF + BD1-nAb-BA.1 + BD1-nAb-Ancestral + BD29-nAb-BA.1 + BD29-nAb-Ancestral | 0.681 [0.594, 0.756] |
| BRF + BD29-bAb-Spike-BA.1 + BD29-bAb-Spike-Ancestral + BD29-nAb-BA.1 + BD29-nAb-Ancestral + Fold-rise-bAb-Spike-BA.1 + Fold-rise-bAb-Spike-Ancestral + Fold-rise-nAb-BA.1 + Fold-rise-nAb-Ancestral | 0.681 [0.595, 0.756] |
| BRF + BD1-bAb-Spike-BA.1 + BD1-nAb-BA.1 + BD29-bAb-Spike-BA.1 + BD29-nAb-BA.1 | 0.680 [0.593, 0.755] |
| BRF + BD29-bAb-Spike-BA.1 + BD29-nAb-BA.1 + Fold-rise-bAb-Spike-BA.1 + Fold-rise-nAb-BA.1 | 0.679 [0.593, 0.755] |
| BRF + BD1-bAb-Spike-BA.1 + BD1-bAb-Spike-Ancestral + BD1-nAb-BA.1 + BD1-nAb-Ancestral + Fold-rise-bAb-Spike-BA.1 + Fold-rise-bAb-Spike-Ancestral + Fold-rise-nAb-BA.1 + Fold-rise-nAb-Ancestral | 0.679 [0.592, 0.755] |
| BRF + BD1-nAb-BA.1 + BD29-nAb-BA.1 | 0.678 [0.592, 0.754] |
| BRF + BD1-bAb-Spike-BA.1 + BD1-bAb-Spike-Ancestral + BD1-bAb-RBD-Ancestral + BD1-nAb-BA.1 + BD1-nAb-Ancestral + Fold-rise-bAb-Spike-BA.1 + Fold-rise-bAb-Spike-Ancestral + Fold-rise-bAb-RBD-Ancestral + Fold-rise-nAb-BA.1 + Fold-rise-nAb-Ancestral | 0.678 [0.592, 0.754] |
| BRF + BD1-bAb-Spike-BA.1 + BD1-nAb-BA.1 + Fold-rise-bAb-Spike-BA.1 + Fold-rise-nAb-BA.1 | 0.678 [0.591, 0.754] |
| BRF + BD29-bAb-RBD-Ancestral + BD29-nAb-BA.1 + BD29-nAb-Ancestral + Fold-rise-bAb-RBD-Ancestral + Fold-rise-nAb-BA.1 + Fold-rise-nAb-Ancestral | 0.678 [0.592, 0.753] |
| BRF + BD29-bAb-RBD-Ancestral + BD29-nAb-BA.1 + BD29-nAb-Ancestral | 0.677 [0.591, 0.753] |
| BRF + BD29-bAb-Spike-BA.1 + BD29-nAb-BA.1 | 0.677 [0.591, 0.753] |
| BRF + BD29-nAb-BA.1 | 0.677 [0.591, 0.753] |
| BRF + BD1-nAb-BA.1 + BD1-nAb-Ancestral + Fold-rise-nAb-BA.1 + Fold-rise-nAb-Ancestral | 0.677 [0.590, 0.753] |
| BRF + BD1-nAb-BA.1 + Fold-rise-nAb-BA.1 | 0.673 [0.586, 0.750] |
| BRF + BD29-bAb-Spike-Ancestral + BD29-nAb-Ancestral | 0.662 [0.575, 0.740] |
| BRF + BD29-nAb-Ancestral | 0.659 [0.572, 0.737] |
| BRF + BD29-bAb-Spike-BA.1 + BD29-bAb-Spike-Ancestral + BD29-bAb-RBD-Ancestral | 0.659 [0.571, 0.738] |
| BRF + BD1-bAb-Spike-Ancestral + BD1-nAb-Ancestral + BD29-bAb-Spike-Ancestral + BD29-nAb-Ancestral | 0.659 [0.571, 0.737] |
| BRF + BD29-bAb-RBD-Ancestral + BD29-nAb-Ancestral | 0.657 [0.569, 0.735] |
| BRF + BD1-bAb-Spike-Ancestral + BD1-bAb-RBD-Ancestral + BD1-nAb-Ancestral + BD29-bAb-Spike-Ancestral + BD29-bAb-RBD-Ancestral + BD29-nAb-Ancestral | 0.657 [0.568, 0.736] |
| BRF + BD29-bAb-Spike-Ancestral + BD29-bAb-RBD-Ancestral + BD29-nAb-Ancestral + Fold-rise-bAb-Spike-Ancestral + Fold-rise-bAb-RBD-Ancestral + Fold-rise-nAb-Ancestral | 0.656 [0.568, 0.735] |
| BRF + BD1-bAb-Spike-BA.1 + BD1-bAb-Spike-Ancestral + BD1-bAb-RBD-Ancestral + BD29-bAb-Spike-BA.1 + BD29-bAb-Spike-Ancestral + BD29-bAb-RBD-Ancestral | 0.656 [0.568, 0.734] |
| BRF + BD1-nAb-Ancestral + BD29-nAb-Ancestral | 0.655 [0.567, 0.734] |
| BRF + BD29-nAb-Ancestral + Fold-rise-nAb-Ancestral | 0.654 [0.566, 0.732] |
| BRF + BD29-bAb-Spike-Ancestral + BD29-nAb-Ancestral + Fold-rise-bAb-Spike-Ancestral + Fold-rise-nAb-Ancestral | 0.653 [0.565, 0.732] |
| BRF + BD29-bAb-RBD-Ancestral + BD29-nAb-Ancestral + Fold-rise-bAb-RBD-Ancestral + Fold-rise-nAb-Ancestral | 0.651 [0.563, 0.730] |
| BRF + BD1-bAb-RBD-Ancestral + BD1-nAb-Ancestral + BD29-bAb-RBD-Ancestral + BD29-nAb-Ancestral | 0.650 [0.562, 0.729] |
| BRF + BD1-bAb-Spike-Ancestral + BD1-bAb-RBD-Ancestral + BD29-bAb-Spike-Ancestral + BD29-bAb-RBD-Ancestral | 0.647 [0.557, 0.726] |
| BRF + BD29-bAb-Spike-Ancestral + BD29-bAb-RBD-Ancestral | 0.646 [0.557, 0.726] |
| BRF + BD29-bAb-Spike-Ancestral + BD29-bAb-RBD-Ancestral + Fold-rise-bAb-Spike-Ancestral + Fold-rise-bAb-RBD-Ancestral | 0.645 [0.555, 0.725] |
| BRF + BD1-bAb-Spike-BA.1 + BD1-nAb-BA.1 | 0.641 [0.552, 0.721] |
| BRF + BD1-bAb-Spike-BA.1 + BD1-bAb-Spike-Ancestral + BD1-nAb-BA.1 + BD1-nAb-Ancestral | 0.640 [0.551, 0.720] |
| BRF + BD1-bAb-RBD-Ancestral + BD1-nAb-BA.1 + BD1-nAb-Ancestral | 0.637 [0.548, 0.718] |
| BRF + BD1-nAb-Ancestral + Fold-rise-nAb-Ancestral | 0.637 [0.548, 0.717] |
| BRF + BD29-bAb-Spike-BA.1 + BD29-bAb-Spike-Ancestral + BD29-bAb-RBD-Ancestral + Fold-rise-bAb-Spike-BA.1 + Fold-rise-bAb-Spike-Ancestral + Fold-rise-bAb-RBD-Ancestral | 0.635 [0.546, 0.716] |
| BRF + BD1-nAb-BA.1 + BD1-nAb-Ancestral | 0.633 [0.544, 0.714] |
| BRF + BD1-nAb-BA.1 | 0.632 [0.543, 0.713] |
| BRF + BD1-bAb-Spike-Ancestral + BD1-nAb-Ancestral + Fold-rise-bAb-Spike-Ancestral + Fold-rise-nAb-Ancestral | 0.629 [0.540, 0.710] |
| BRF + BD29-bAb-RBD-Ancestral + Fold-rise-bAb-RBD-Ancestral | 0.625 [0.537, 0.706] |
| BRF + BD1-bAb-Spike-Ancestral + BD1-bAb-RBD-Ancestral + BD1-nAb-Ancestral + Fold-rise-bAb-Spike-Ancestral + Fold-rise-bAb-RBD-Ancestral + Fold-rise-nAb-Ancestral | 0.623 [0.534, 0.704] |
| BRF + BD29-bAb-Spike-BA.1 + BD29-bAb-Spike-Ancestral | 0.622 [0.533, 0.703] |
| BRF + BD1-bAb-RBD-Ancestral + BD29-bAb-RBD-Ancestral | 0.621 [0.533, 0.702] |
| BRF + BD1-bAb-Spike-Ancestral + BD1-bAb-RBD-Ancestral + Fold-rise-bAb-Spike-Ancestral + Fold-rise-bAb-RBD-Ancestral | 0.620 [0.530, 0.702] |
| BRF + BD29-bAb-RBD-Ancestral | 0.620 [0.531, 0.701] |
| BRF + BD29-bAb-Spike-BA.1 + BD29-bAb-Spike-Ancestral + Fold-rise-bAb-Spike-BA.1 + Fold-rise-bAb-Spike-Ancestral | 0.618 [0.529, 0.699] |
| BRF + BD1-bAb-RBD-Ancestral + BD1-nAb-Ancestral + Fold-rise-bAb-RBD-Ancestral + Fold-rise-nAb-Ancestral | 0.616 [0.528, 0.698] |
| BRF + BD1-bAb-Spike-BA.1 + BD1-bAb-Spike-Ancestral + BD29-bAb-Spike-BA.1 + BD29-bAb-Spike-Ancestral | 0.612 [0.523, 0.694] |
| BRF + BD1-bAb-RBD-Ancestral + Fold-rise-bAb-RBD-Ancestral | 0.611 [0.522, 0.693] |
| BRF + BD29-bAb-Spike-BA.1 + Fold-rise-bAb-Spike-BA.1 | 0.605 [0.516, 0.688] |
| BRF + Fold-rise-bAb-RBD-Ancestral + Fold-rise-nAb-BA.1 + Fold-rise-nAb-Ancestral | 0.603 [0.514, 0.686] |
| BRF + Fold-rise-bAb-Spike-Ancestral + Fold-rise-bAb-RBD-Ancestral | 0.603 [0.513, 0.686] |
| BRF + BD1-bAb-Spike-BA.1 + BD29-bAb-Spike-BA.1 | 0.603 [0.514, 0.685] |
| BRF + Fold-rise-bAb-Spike-BA.1 + Fold-rise-bAb-Spike-Ancestral + Fold-rise-nAb-BA.1 + Fold-rise-nAb-Ancestral | 0.601 [0.512, 0.685] |
| BRF + Fold-rise-nAb-BA.1 + Fold-rise-nAb-Ancestral | 0.597 [0.508, 0.681] |
| BRF + BD1-bAb-Spike-BA.1 + BD1-bAb-Spike-Ancestral + BD1-bAb-RBD-Ancestral + Fold-rise-bAb-Spike-BA.1 + Fold-rise-bAb-Spike-Ancestral + Fold-rise-bAb-RBD-Ancestral | 0.597 [0.507, 0.680] |
| BRF + Fold-rise-bAb-Spike-BA.1 + Fold-rise-bAb-Spike-Ancestral + Fold-rise-bAb-RBD-Ancestral | 0.596 [0.506, 0.680] |
| BRF + BD29-bAb-Spike-BA.1 | 0.594 [0.505, 0.677] |
| BRF + Fold-rise-bAb-Spike-BA.1 + Fold-rise-nAb-BA.1 | 0.593 [0.504, 0.676] |
| BRF + BD29-bAb-Spike-Ancestral + Fold-rise-bAb-Spike-Ancestral | 0.590 [0.501, 0.674] |
| BRF + BD1-bAb-Spike-BA.1 + Fold-rise-bAb-Spike-BA.1 | 0.587 [0.498, 0.670] |
| BRF + Fold-rise-bAb-RBD-Ancestral + Fold-rise-nAb-Ancestral | 0.586 [0.496, 0.670] |
| BRF + Fold-rise-nAb-BA.1 | 0.582 [0.493, 0.666] |
| BRF + BD1-bAb-Spike-Ancestral + BD29-bAb-Spike-Ancestral | 0.579 [0.490, 0.664] |
| BRF + BD29-bAb-Spike-Ancestral | 0.579 [0.489, 0.664] |
| BRF + BD1-bAb-Spike-BA.1 + BD1-bAb-Spike-Ancestral + Fold-rise-bAb-Spike-BA.1 + Fold-rise-bAb-Spike-Ancestral | 0.579 [0.490, 0.664] |
| BRF + Fold-rise-bAb-Spike-BA.1 + Fold-rise-bAb-Spike-Ancestral | 0.578 [0.488, 0.663] |
| BRF + Fold-rise-bAb-Spike-Ancestral + Fold-rise-nAb-Ancestral | 0.575 [0.484, 0.661] |
| BRF + Fold-rise-nAb-Ancestral | 0.573 [0.482, 0.660] |
| BRF + BD1-bAb-Spike-Ancestral + Fold-rise-bAb-Spike-Ancestral | 0.573 [0.483, 0.657] |
| BRF + Fold-rise-bAb-RBD-Ancestral | 0.570 [0.481, 0.655] |
| BRF + Fold-rise-bAb-Spike-BA.1 | 0.565 [0.475, 0.650] |
| BRF + Fold-rise-bAb-Spike-Ancestral | 0.540 [0.451, 0.627] |
| BRF + BD1-bAb-Spike-Ancestral + BD1-nAb-Ancestral | 0.534 [0.438, 0.627] |
| BRF + BD1-nAb-Ancestral | 0.527 [0.432, 0.620] |
| BRF + BD1-bAb-Spike-BA.1 + BD1-bAb-Spike-Ancestral + BD1-bAb-RBD-Ancestral | 0.524 [0.417, 0.629] |
| BRF + BD1-bAb-Spike-Ancestral + BD1-bAb-RBD-Ancestral | 0.523 [0.416, 0.629] |
| BRF + BD1-bAb-Spike-BA.1 | 0.522 [0.415, 0.628] |
| BRF + BD1-bAb-RBD-Ancestral + BD1-nAb-Ancestral | 0.522 [0.424, 0.618] |
| BRF + BD1-bAb-Spike-BA.1 + BD1-bAb-Spike-Ancestral | 0.518 [0.406, 0.630] |
| BRF + BD1-bAb-Spike-Ancestral | 0.518 [0.405, 0.629] |
| BRF + BD1-bAb-RBD-Ancestral | 0.517 [0.404, 0.629] |
| BRF (baseline risk factors) | 0.517 [0.404, 0.628] |

### Table S11. Discrete Super Learner performance across all 92 variable sets sorted by cross-validated area under the ROC (CV-AUC) performance for predicting occurrence of Omicron COVID-19 in non-naive per-protocol boosted participants. bAb = binding antibody; BRF = baseline risk factors; fold-rise = BD29/BD1; nAb = neutralizing antibody.

| **Variable Set** | **CV-AUC (95% CI)** |
| --- | --- |
| BRF + BD29-bAb-RBD-Ancestral + BD29-nAb-Ancestral | 0.712 [0.558, 0.829] |
| BRF + Fold-rise-bAb-RBD-Ancestral + Fold-rise-nAb-Ancestral | 0.683 [0.530, 0.804] |
| BRF + BD1-bAb-RBD-Ancestral + BD1-nAb-Ancestral + Fold-rise-bAb-RBD-Ancestral + Fold-rise-nAb-Ancestral | 0.681 [0.526, 0.804] |
| BRF + Fold-rise-bAb-RBD-Ancestral + Fold-rise-nAb-BA.1 + Fold-rise-nAb-Ancestral | 0.680 [0.527, 0.802] |
| BRF + BD1-bAb-RBD-Ancestral + BD1-nAb-BA.1 + BD1-nAb-Ancestral + Fold-rise-bAb-RBD-Ancestral + Fold-rise-nAb-BA.1 + Fold-rise-nAb-Ancestral | 0.673 [0.516, 0.799] |
| BRF + BD1-bAb-Spike-Ancestral + BD1-bAb-RBD-Ancestral + BD1-nAb-Ancestral + Fold-rise-bAb-Spike-Ancestral + Fold-rise-bAb-RBD-Ancestral + Fold-rise-nAb-Ancestral | 0.666 [0.510, 0.792] |
| BRF + BD1-bAb-Spike-Ancestral + BD1-bAb-RBD-Ancestral + Fold-rise-bAb-Spike-Ancestral + Fold-rise-bAb-RBD-Ancestral | 0.657 [0.499, 0.787] |
| BRF + BD29-bAb-Spike-Ancestral + BD29-nAb-Ancestral | 0.656 [0.500, 0.784] |
| BRF + Fold-rise-bAb-Spike-Ancestral + Fold-rise-bAb-RBD-Ancestral | 0.654 [0.497, 0.784] |
| BRF + Fold-rise-bAb-RBD-Ancestral | 0.651 [0.496, 0.780] |
| BRF + BD1-bAb-RBD-Ancestral + Fold-rise-bAb-RBD-Ancestral | 0.650 [0.493, 0.780] |
| BRF + BD1-bAb-Spike-BA.1 + BD1-bAb-Spike-Ancestral + BD1-bAb-RBD-Ancestral + BD1-nAb-BA.1 + BD1-nAb-Ancestral + Fold-rise-bAb-Spike-BA.1 + Fold-rise-bAb-Spike-Ancestral + Fold-rise-bAb-RBD-Ancestral + Fold-rise-nAb-BA.1 + Fold-rise-nAb-Ancestral | 0.646 [0.491, 0.775] |
| BRF + BD1-bAb-Spike-BA.1 + BD1-bAb-Spike-Ancestral + BD1-bAb-RBD-Ancestral + Fold-rise-bAb-Spike-BA.1 + Fold-rise-bAb-Spike-Ancestral + Fold-rise-bAb-RBD-Ancestral | 0.643 [0.478, 0.782] |
| BRF + Fold-rise-bAb-Spike-BA.1 + Fold-rise-bAb-Spike-Ancestral + Fold-rise-bAb-RBD-Ancestral | 0.640 [0.475, 0.780] |
| BRF + Fold-rise-bAb-Spike-Ancestral + Fold-rise-nAb-Ancestral | 0.639 [0.481, 0.772] |
| BRF + BD1-bAb-RBD-Ancestral + BD1-nAb-Ancestral + BD29-bAb-RBD-Ancestral + BD29-nAb-Ancestral | 0.636 [0.478, 0.770] |
| BRF + BD29-bAb-RBD-Ancestral + Fold-rise-bAb-RBD-Ancestral | 0.633 [0.475, 0.767] |
| BRF + BD1-bAb-Spike-Ancestral + BD1-nAb-Ancestral + Fold-rise-bAb-Spike-Ancestral + Fold-rise-nAb-Ancestral | 0.625 [0.468, 0.760] |
| BRF + Fold-rise-bAb-Spike-BA.1 + Fold-rise-bAb-Spike-Ancestral | 0.624 [0.464, 0.761] |
| BRF + BD1-bAb-Spike-BA.1 + BD1-bAb-Spike-Ancestral + BD1-nAb-BA.1 + BD1-nAb-Ancestral + Fold-rise-bAb-Spike-BA.1 + Fold-rise-bAb-Spike-Ancestral + Fold-rise-nAb-BA.1 + Fold-rise-nAb-Ancestral | 0.622 [0.463, 0.759] |
| BRF + Fold-rise-bAb-Spike-BA.1 + Fold-rise-bAb-Spike-Ancestral + Fold-rise-nAb-BA.1 + Fold-rise-nAb-Ancestral | 0.622 [0.465, 0.756] |
| BRF + BD1-bAb-RBD-Ancestral + BD1-nAb-BA.1 + BD1-nAb-Ancestral | 0.621 [0.465, 0.756] |
| BRF + BD1-nAb-Ancestral + Fold-rise-nAb-Ancestral | 0.621 [0.466, 0.755] |
| BRF + BD1-bAb-Spike-BA.1 + BD1-bAb-Spike-Ancestral + Fold-rise-bAb-Spike-BA.1 + Fold-rise-bAb-Spike-Ancestral | 0.619 [0.459, 0.757] |
| BRF + BD29-bAb-RBD-Ancestral + BD29-nAb-Ancestral + Fold-rise-bAb-RBD-Ancestral + Fold-rise-nAb-Ancestral | 0.619 [0.460, 0.756] |
| BRF + BD1-nAb-BA.1 + BD1-nAb-Ancestral | 0.617 [0.460, 0.753] |
| BRF + BD29-bAb-RBD-Ancestral + BD29-nAb-BA.1 + BD29-nAb-Ancestral | 0.616 [0.460, 0.752] |
| BRF + BD29-bAb-Spike-Ancestral + BD29-bAb-RBD-Ancestral + Fold-rise-bAb-Spike-Ancestral + Fold-rise-bAb-RBD-Ancestral | 0.615 [0.455, 0.753] |
| BRF + Fold-rise-bAb-Spike-Ancestral | 0.614 [0.457, 0.751] |
| BRF + BD29-bAb-Spike-Ancestral + BD29-bAb-RBD-Ancestral + BD29-nAb-Ancestral + Fold-rise-bAb-Spike-Ancestral + Fold-rise-bAb-RBD-Ancestral + Fold-rise-nAb-Ancestral | 0.614 [0.456, 0.751] |
| BRF + Fold-rise-bAb-Spike-BA.1 | 0.611 [0.454, 0.748] |
| BRF + BD1-bAb-Spike-Ancestral + Fold-rise-bAb-Spike-Ancestral | 0.609 [0.452, 0.747] |
| BRF + BD29-nAb-BA.1 + BD29-nAb-Ancestral | 0.607 [0.443, 0.752] |
| BRF + BD1-bAb-Spike-BA.1 + Fold-rise-bAb-Spike-BA.1 | 0.606 [0.452, 0.741] |
| BRF + BD1-bAb-RBD-Ancestral + BD1-nAb-Ancestral | 0.603 [0.443, 0.744] |
| BRF + BD1-bAb-RBD-Ancestral + BD29-bAb-RBD-Ancestral | 0.603 [0.444, 0.742] |
| BRF + BD1-nAb-BA.1 + BD1-nAb-Ancestral + Fold-rise-nAb-BA.1 + Fold-rise-nAb-Ancestral | 0.602 [0.443, 0.742] |
| BRF + BD1-bAb-Spike-Ancestral + BD1-nAb-Ancestral + BD29-bAb-Spike-Ancestral + BD29-nAb-Ancestral | 0.601 [0.444, 0.738] |
| BRF + BD1-bAb-Spike-BA.1 + BD1-bAb-Spike-Ancestral + BD1-nAb-BA.1 + BD1-nAb-Ancestral | 0.599 [0.440, 0.739] |
| BRF + Fold-rise-bAb-Spike-BA.1 + Fold-rise-nAb-BA.1 | 0.598 [0.441, 0.736] |
| BRF + Fold-rise-nAb-BA.1 + Fold-rise-nAb-Ancestral | 0.597 [0.423, 0.752] |
| BRF + BD29-bAb-Spike-BA.1 + BD29-bAb-Spike-Ancestral + BD29-bAb-RBD-Ancestral | 0.596 [0.441, 0.734] |
| BRF + BD29-bAb-Spike-Ancestral + Fold-rise-bAb-Spike-Ancestral | 0.595 [0.427, 0.745] |
| BRF + BD29-bAb-Spike-Ancestral + BD29-nAb-Ancestral + Fold-rise-bAb-Spike-Ancestral + Fold-rise-nAb-Ancestral | 0.595 [0.415, 0.756] |
| BRF + BD1-bAb-Spike-Ancestral + BD1-bAb-RBD-Ancestral + BD1-nAb-Ancestral + BD29-bAb-Spike-Ancestral + BD29-bAb-RBD-Ancestral + BD29-nAb-Ancestral | 0.594 [0.434, 0.737] |
| BRF + BD1-bAb-Spike-BA.1 + BD1-nAb-BA.1 + Fold-rise-bAb-Spike-BA.1 + Fold-rise-nAb-BA.1 | 0.593 [0.438, 0.731] |
| BRF + BD1-bAb-Spike-Ancestral + BD1-nAb-Ancestral | 0.591 [0.432, 0.732] |
| BRF + BD1-bAb-RBD-Ancestral + BD1-nAb-BA.1 + BD1-nAb-Ancestral + BD29-bAb-RBD-Ancestral + BD29-nAb-BA.1 + BD29-nAb-Ancestral | 0.590 [0.432, 0.732] |
| BRF + BD29-bAb-RBD-Ancestral + BD29-nAb-BA.1 + BD29-nAb-Ancestral + Fold-rise-bAb-RBD-Ancestral + Fold-rise-nAb-BA.1 + Fold-rise-nAb-Ancestral | 0.590 [0.434, 0.730] |
| BRF + BD29-bAb-Spike-BA.1 + BD29-bAb-Spike-Ancestral + BD29-bAb-RBD-Ancestral + Fold-rise-bAb-Spike-BA.1 + Fold-rise-bAb-Spike-Ancestral + Fold-rise-bAb-RBD-Ancestral | 0.590 [0.432, 0.731] |
| BRF + BD1-nAb-BA.1 + BD1-nAb-Ancestral + BD29-nAb-BA.1 + BD29-nAb-Ancestral | 0.590 [0.425, 0.737] |
| BRF + BD1-nAb-Ancestral + BD29-nAb-Ancestral | 0.586 [0.423, 0.733] |
| BRF + Fold-rise-nAb-Ancestral | 0.586 [0.428, 0.728] |
| BRF + BD29-bAb-Spike-BA.1 + BD29-bAb-Spike-Ancestral + Fold-rise-bAb-Spike-BA.1 + Fold-rise-bAb-Spike-Ancestral | 0.585 [0.409, 0.745] |
| BRF + BD1-bAb-Spike-Ancestral + BD29-bAb-Spike-Ancestral | 0.584 [0.419, 0.733] |
| BRF + BD29-bAb-Spike-BA.1 + Fold-rise-bAb-Spike-BA.1 | 0.584 [0.415, 0.736] |
| BRF + BD1-bAb-Spike-BA.1 + BD1-bAb-Spike-Ancestral + BD1-bAb-RBD-Ancestral + BD29-bAb-Spike-BA.1 + BD29-bAb-Spike-Ancestral + BD29-bAb-RBD-Ancestral | 0.583 [0.418, 0.733] |
| BRF + BD1-nAb-Ancestral | 0.582 [0.426, 0.724] |
| BRF + BD29-bAb-Spike-Ancestral + BD29-bAb-RBD-Ancestral | 0.580 [0.417, 0.729] |
| BRF + BD1-bAb-Spike-BA.1 + BD29-bAb-Spike-BA.1 | 0.580 [0.405, 0.739] |
| BRF + BD29-bAb-Spike-BA.1 + BD29-bAb-Spike-Ancestral + BD29-nAb-BA.1 + BD29-nAb-Ancestral | 0.578 [0.418, 0.725] |
| BRF + BD29-bAb-RBD-Ancestral | 0.577 [0.411, 0.728] |
| BRF + BD29-nAb-Ancestral + Fold-rise-nAb-Ancestral | 0.576 [0.413, 0.725] |
| BRF + Fold-rise-nAb-BA.1 | 0.575 [0.408, 0.727] |
| BRF + BD1-bAb-Spike-Ancestral + BD1-bAb-RBD-Ancestral + BD29-bAb-Spike-Ancestral + BD29-bAb-RBD-Ancestral | 0.574 [0.404, 0.731] |
| BRF + BD1-bAb-Spike-BA.1 + BD1-bAb-Spike-Ancestral | 0.573 [0.407, 0.726] |
| BRF + BD1-bAb-Spike-BA.1 + BD1-bAb-Spike-Ancestral + BD1-bAb-RBD-Ancestral + BD1-nAb-BA.1 + BD1-nAb-Ancestral + BD29-bAb-Spike-BA.1 + BD29-bAb-Spike-Ancestral + BD29-bAb-RBD-Ancestral + BD29-nAb-BA.1 + BD29-nAb-Ancestral | 0.573 [0.403, 0.728] |
| BRF + BD29-bAb-Spike-Ancestral | 0.573 [0.410, 0.722] |
| BRF + BD1-bAb-Spike-Ancestral + BD1-bAb-RBD-Ancestral | 0.570 [0.406, 0.721] |
| BRF + BD29-bAb-Spike-BA.1 + BD29-bAb-Spike-Ancestral + BD29-bAb-RBD-Ancestral + BD29-nAb-BA.1 + BD29-nAb-Ancestral + Fold-rise-bAb-Spike-BA.1 + Fold-rise-bAb-Spike-Ancestral + Fold-rise-bAb-RBD-Ancestral + Fold-rise-nAb-BA.1 + Fold-rise-nAb-Ancestral | 0.569 [0.412, 0.713] |
| BRF + BD1-bAb-Spike-BA.1 + BD1-bAb-Spike-Ancestral + BD1-bAb-RBD-Ancestral + BD1-nAb-BA.1 + BD1-nAb-Ancestral + BD29-bAb-Spike-BA.1 + BD29-bAb-Spike-Ancestral + BD29-bAb-RBD-Ancestral + BD29-nAb-BA.1 + BD29-nAb-Ancestral + Fold-rise-bAb-Spike-BA.1 + Fold-rise-bAb-Spike-Ancestral + Fold-rise-bAb-RBD-Ancestral + Fold-rise-nAb-BA.1 + Fold-rise-nAb-Ancestral | 0.569 [0.412, 0.713] |
| BRF + BD29-nAb-BA.1 + BD29-nAb-Ancestral + Fold-rise-nAb-BA.1 + Fold-rise-nAb-Ancestral | 0.568 [0.380, 0.742] |
| BRF + BD1-nAb-BA.1 + Fold-rise-nAb-BA.1 | 0.567 [0.388, 0.733] |
| BRF + BD1-bAb-Spike-BA.1 + BD1-bAb-Spike-Ancestral + BD1-bAb-RBD-Ancestral | 0.566 [0.402, 0.718] |
| BRF + BD29-bAb-Spike-BA.1 + BD29-nAb-BA.1 + Fold-rise-bAb-Spike-BA.1 + Fold-rise-nAb-BA.1 | 0.565 [0.390, 0.728] |
| BRF + BD1-bAb-Spike-BA.1 + BD1-bAb-Spike-Ancestral + BD29-bAb-Spike-BA.1 + BD29-bAb-Spike-Ancestral | 0.565 [0.403, 0.715] |
| BRF + BD29-bAb-Spike-BA.1 + BD29-bAb-Spike-Ancestral | 0.564 [0.404, 0.713] |
| BRF + BD1-bAb-Spike-BA.1 | 0.563 [0.391, 0.724] |
| BRF + BD1-bAb-Spike-Ancestral | 0.563 [0.400, 0.715] |
| BRF + BD29-nAb-BA.1 + Fold-rise-nAb-BA.1 | 0.563 [0.393, 0.720] |
| BRF + BD1-bAb-Spike-BA.1 + BD1-bAb-Spike-Ancestral + BD1-nAb-BA.1 + BD1-nAb-Ancestral + BD29-bAb-Spike-BA.1 + BD29-bAb-Spike-Ancestral + BD29-nAb-BA.1 + BD29-nAb-Ancestral | 0.562 [0.397, 0.716] |
| BRF + BD1-bAb-Spike-BA.1 + BD1-nAb-BA.1 | 0.562 [0.394, 0.718] |
| BRF + BD1-bAb-RBD-Ancestral | 0.561 [0.398, 0.713] |
| BRF (baseline risk factors) | 0.561 [0.389, 0.720] |
| BRF + BD29-bAb-Spike-BA.1 | 0.560 [0.397, 0.712] |
| BRF + BD29-nAb-BA.1 | 0.559 [0.389, 0.719] |
| BRF + BD1-bAb-Spike-BA.1 + BD1-nAb-BA.1 + BD29-bAb-Spike-BA.1 + BD29-nAb-BA.1 | 0.559 [0.386, 0.721] |
| BRF + BD29-nAb-Ancestral | 0.558 [0.377, 0.727] |
| BRF + BD29-bAb-Spike-BA.1 + BD29-nAb-BA.1 | 0.557 [0.394, 0.710] |
| BRF + BD1-nAb-BA.1 + BD29-nAb-BA.1 | 0.557 [0.385, 0.718] |
| BRF + BD29-bAb-Spike-BA.1 + BD29-bAb-Spike-Ancestral + BD29-nAb-BA.1 + BD29-nAb-Ancestral + Fold-rise-bAb-Spike-BA.1 + Fold-rise-bAb-Spike-Ancestral + Fold-rise-nAb-BA.1 + Fold-rise-nAb-Ancestral | 0.556 [0.370, 0.731] |
| BRF + BD1-nAb-BA.1 | 0.555 [0.384, 0.715] |

### Supplementary Text

#### Comparison of Duke and PPD calibration experiments to WHO IS

Both Duke and PPD obtained the WHO International Standard (IS) lot 20/136 (*15*) and performed multiple pseudovirus-based neutralizing antibody assays (nAb) (*4, 11*) on the standard, which was assigned by the WHO to have 50% inhibitory dilution (ID50) nAb activity of 1000 International Units (IU)/ml. Duke reconstituted the WHO IS twice and obtained 10 nAb assay readouts of reconstitution 1 and 28 of reconstitution 2 (*16*). PPD obtained 20 readouts of a single reconstitution (*17*). The results are shown below in **Figure S33**. Below is a plot of the readouts and the arithmetic average (black line).

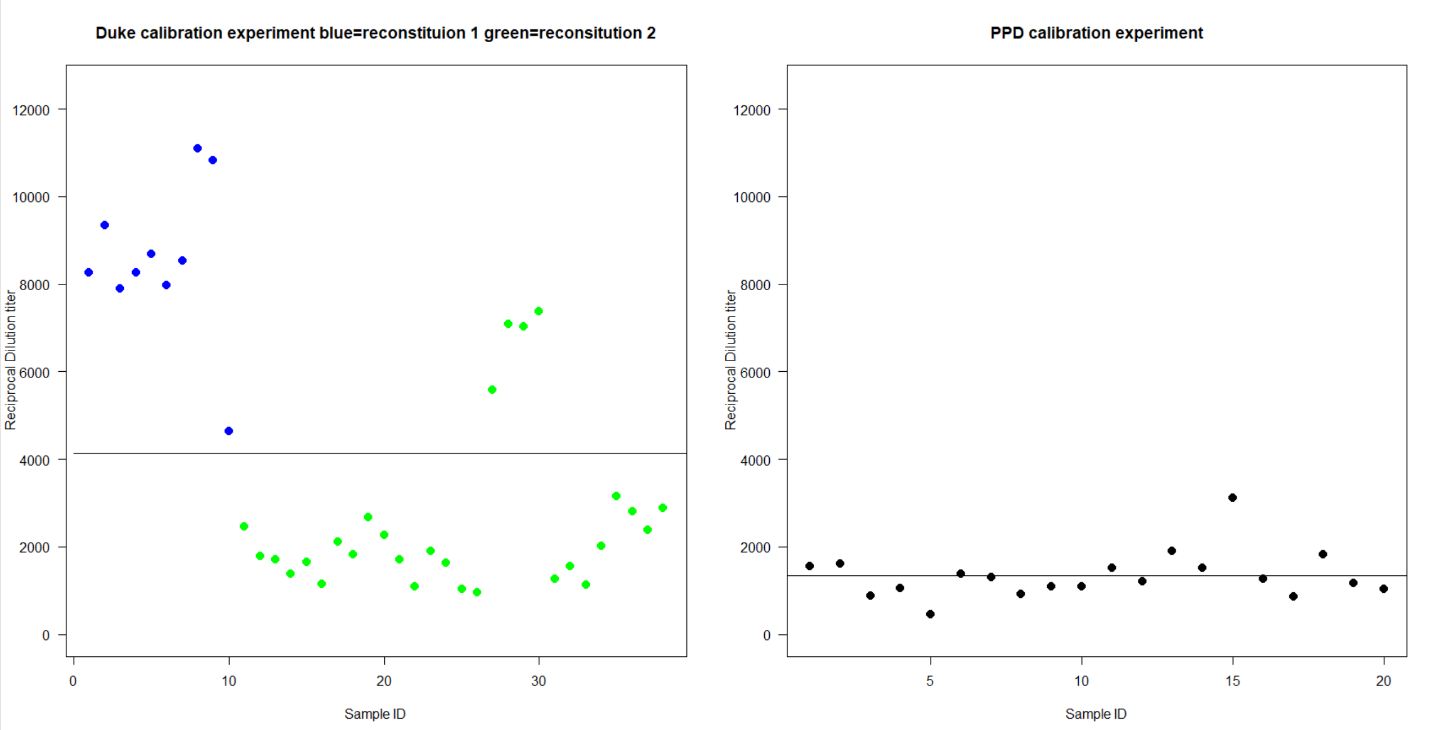

Figure S33. Reciprocal 50% inhibitory dilution pseudovirus neutralizing antibody (nAb ID50) titers from the Duke calibration experiment (left panel) and the PPD calibration experiment (right panel). In the left panel, the blue dots represent samples from reconstitution 1 of the WHO IS 20/136 and the green dots represent samples from reconstitution 2 of the WHO IS 20/136. The horizontal black line in each panel is the arithmetic average across all samples. Each dot represents a single assay run.

ID50 and 80% inhibitory dilution (ID80) titers are converted to IU50s and IU80s, respectively, by dividing 1000 IU/mL by either the mean, median or geometric mean ID50 and ID80 titer as a dilution factor [further explained in the Supplementary Material of (*4*)]. The arithmetic mean for Duke was 4135, thus the conversion factor for mean ID50 is 1000 ÷ 4135 = 0.242. The geometric mean for PPD was 1248, which was close to the conversion factor used by PPD (a more sophisticated analysis yielded 1275).

The ID50 titers from reconstitution 2 of Duke and those of the PPD reconstitution look similar, except for 4 green ‘outliers’ from four sample IDs. Using reconstitution 2 only and excluding these four sample IDs, Duke’s geometric mean is 1753, which is closer to the PPD geometric mean of 1248. Reconstitution 1 and the four outliers have a geometric mean of 7855, more than 4-fold higher.

PPD’s comparison of PPD with Historical Duke(*10*) showed that the two readouts were very similar (**Figure S34**). This led to the conclusion that the analysis “supports the equivalency of PPDs VSDVAC62 and Duke ID0 with a slope of 0.98 and a bias of 1.04” [page 12 in (*10*)]. It was recommended to use a conversion factor VAC62 to Duke ID50 of VAC62/1.04 [page 130 in (*10*)].

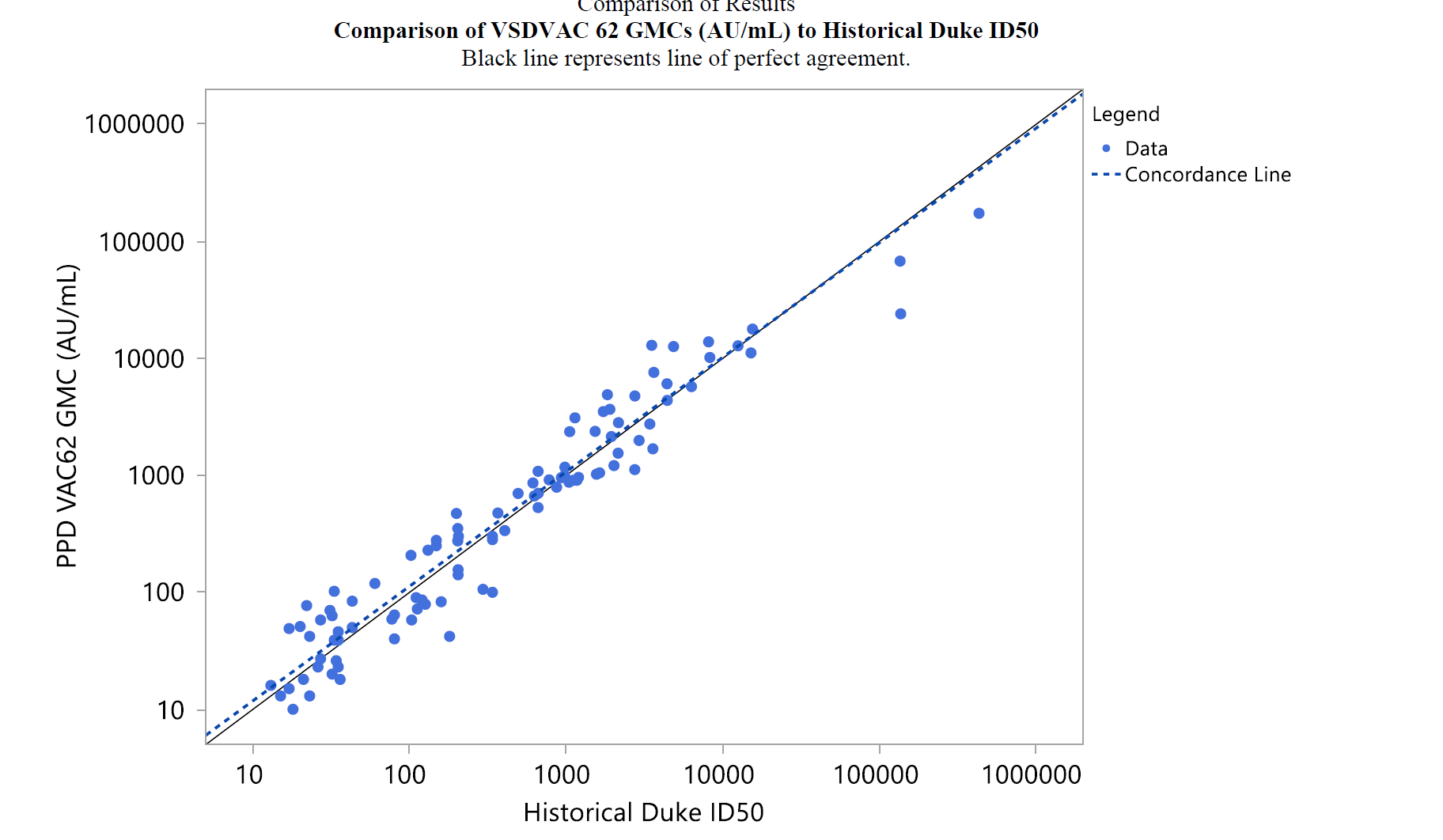

### Figure S34. Comparison of 100 Duke nAb ID50 readouts (i.e. not converted to IU50/ml) versus an average of 3 PPD readouts in arbitrary units (AU)/ml. The black line represents perfect agreement.

**Figure S34** shows no outliers where Duke is pulled to the right, as might be expected from the left panel in **Figure S33**. Attachment I on page 24 in (*10*) shows that the VAC62/Duke ID50 ratio is generally around 1 and always with [0.18, 3.49], never 4-fold.

Cumulatively, these findings provide strong evidence that typically Duke ID50 and PPD are approximately equivalent. A likely explanation for why the two conversion factors (1.275 PPD to WHO IU50/ml, 4.13 Duke to WHO IU) are so different is that Duke’s experiment had some aberrations and then used a mean not median conversion factor.

Based on the above considerations, we analyze ID50 readouts of the form VAC62/1.04 to make it like Duke ID50 and as recommended in (*10*), and then multiply by 0.242 as the conversion factor Duke uses to convert to IU50/ml. Both ID50 readouts (against the Ancestral strain and against the BA.1 strain) are multiplied by (0.242/1.04), where only the Ancestral strain readouts can be interpreted to be on the IU50/ml scale.
